# Integrative multi-omics QTL colocalization maps regulatory architecture in aging human brain

**DOI:** 10.1101/2025.04.17.25326042

**Authors:** Xuewei Cao, Haochen Sun, Ru Feng, Rahul Mazumder, Carlos F Buen Abad Najar, Yang I. Li, Philip L. de Jager, David Bennett, The Alzheimer’s Disease Functional Genomics Consortium, Kushal K. Dey, Gao Wang

## Abstract

Multi-trait QTL (xQTL) colocalization has shown great promises in identifying causal variants with shared genetic etiology across multiple molecular modalities, contexts, and complex diseases. However, the lack of scalable and efficient methods to integrate large-scale multi-omics data limits deeper insights into xQTL regulation. Here, we propose *ColocBoost*, a multi-task learning colocalization method that can scale to hundreds of traits, while accounting for multiple causal variants within a genomic region of interest. *ColocBoost* employs a specialized gradient boosting framework that can adaptively couple colocalized traits while performing causal variant selection, thereby enhancing the detection of weaker shared signals compared to existing pairwise and multi-trait colocalization methods. We applied *ColocBoost* genome-wide to 17 gene-level single-nucleus and bulk xQTL data from the aging brain cortex of ROSMAP individuals (average *N* = 595), encompassing 6 cell types, 3 brain regions and 3 molecular modalities (expression, splicing, and protein abundance). Across molecular xQTLs, *ColocBoost* identified 16,503 distinct colocalization events, exhibiting 10.7(±0.74)-fold enrichment for heritability across 57 complex diseases/traits and showing strong concordance with element-gene pairs validated by CRISPR screening assays. When colocalized against Alzheimer’s disease (AD) GWAS, *ColocBoost* identified up to 2.5-fold more distinct colocalized loci, explaining twice the AD disease heritability compared to fine-mapping without xQTL integration. This improvement is largely attributable to *ColocBoost*’s enhanced sensitivity in detecting gene-distal colocalizations, as supported by strong concordance with known enhancer-gene links, highlighting its ability to identify biologically plausible AD susceptibility loci with underlying regulatory mechanisms. Notably, several genes including *BLNK* and *CTSH* showed sub-threshold associations in GWAS, but were identified through multi-omics colocalizations which provide new functional support for their involvement in AD pathogenesis.

## Introduction

Quantitative trait loci mapping of molecular phenotypes (xQTL) has been highly informative in dissecting the regulatory mechanisms underlying complex disease susceptibility loci^1–9^. Statistical colocalization methods have been instrumental in identifying causal variants shared across molecular phenotypes and complex diseases, further facilitating the prioritization of candidate causal genes and relevant cell types or tissues. However, as xQTL studies expand to encompass diverse molecular modalities, cell types, and tissues, existing colocalization methods have struggled to adequately address the resulting analytical challenges^10–12^. Existing colocalization methods are limited in their ability to handle multiple causal variants and scale to large numbers of traits. For example, the widely used COLOC^13,14^ method allows for multiple causal variants but is restricted to pairwise trait analysis, while HyPrColoc^15^, despite scaling to many traits, is limited to single causal variant per trait per gene. Other methods like eCAVIAR^16^, MOLOC^17^, Sherlock^18^, Primo^19^ are constrained in both computational scalability and the complexity of causal configurations they can accommodate. Critically, in our empirical analyses, these approaches have consistently struggled to capture gene-distal GWAS-xQTL colocalizations. These limitations underscore the need for a more versatile framework that scales to many traits, accommodates multiple causal variants, and robustly detects subtle gene-distal effects in order to fully leverage growing resources of heterogeneous, high-dimensional multi-omics data.

Here we propose *ColocBoost*, a new multi-task learning colocalization method that efficiently scales to tens or hundreds of traits while accommodating multiple causal variants within genomic regions of interest. *ColocBoost* employs an iterative gradient boosting framework in which weak learners are applied in a coupled manner across traits at each boosting round. Putative causal variants are identified through linkage disequilibrium (LD) proximity smoothing to account for correlation among nearby variants. We extensively benchmark *ColocBoost* against existing colocalization methods (COLOC^13,14^, MOLOC^17^, HyPrColoc^15^, OPERA^20^) using real-world data informed simulations, ensuring robust performance across a wide range of practical and theoretical conditions. We applied *ColocBoost* to the newly developed FunGen-xQTL resource from the Alzheimer’s Disease Sequencing Project (ADSP) Functional Genomics Consortium (FunGen-AD)^21,22^, analyzing 17 gene-level molecular traits measured in range from 226 to 784 (average *N*=595) post-mortem donors^23,24^, encompassing 3 brain tissues, 6 brain cell types, and 3 gene-level molecular modalities (expression, splicing, and protein abundance). *ColocBoost* was deployed in both disease-agnostic (xQTL-only) and disease-prioritized settings for Alzheimer’s disease (AD) GWAS (GWAS-xQTL). We demonstrate improved performance of *ColocBoost* over existing methods on (i) disease heritability enrichment of identified variants (xQTL-only) across 57 relatively independent (genetic correlation *r*_*g*_< 0.7) diseases and traits (average *N*=305K)^25^ using Stratified LD Score Regression (S-LDSC)^26,27^, and (ii) enrichment of regulatory element-gene links validated by CRISPRi screening assays^28–31^ and promoter-capture Hi-C^32,33^. In the AD GWAS-prioritized analysis, *ColocBoost* provides functional insights into numerous GWAS loci by leveraging rich multi-omics data across molecular layers. In addition to refining loci with genome-wide significance, it further improves the identification of putative causal variants at sub-threshold GWAS loci, revealing regulatory mechanisms with support by orthogonal evidence from the ENCODE-rE2G enhancer-gene catalogs^34^.

## Results

### Overview of *ColocBoost*

*ColocBoost* is a multi-trait colocalization method that accommodates multiple causal variants at a locus, scales efficiently to hundreds of traits, and detects colocalization events for variants with relatively weak effects (**Table 1**). Each colocalization event {*s*} is characterized by a triplet {*CoS*_*α*_(*s*), *T*(*s*), *g*(*s*)}, where *CoS* is a *Colocalization Confidence Set* of candidate causal variants designed to capture the shared causal variant alongside its high-LD proxies, *T* is the *Causal Configuration* specifying which traits share the effect, and *g* is the *Genomic Region. ColocBoost* formulates colocalization as a multi-task regression for *L* traits, each with phenotype (centered at zero) and standardized genotype data 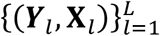, and effect-size vectors ***β***_*l*_ = (*β*_1*l*_, ⋯, *β*_*Pl*_) for *P* variants, assuming 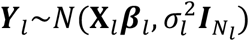 where *N*_*l*_ is the sample size for trait *l*. A variant *j* is *colocalized* if it has non-zero effect size (*β*_*jl*_ ≠ 0) for two or more traits. Two complementary modes are provided, a *disease-agnostic* mode for xQTL-only integration and a *disease-prioritized* mode for GWAS-xQTL colocalization where the latter focuses on a focal disease trait (**Figure 1**; **Methods** and **Supplementary Note**).

**Table 1.**
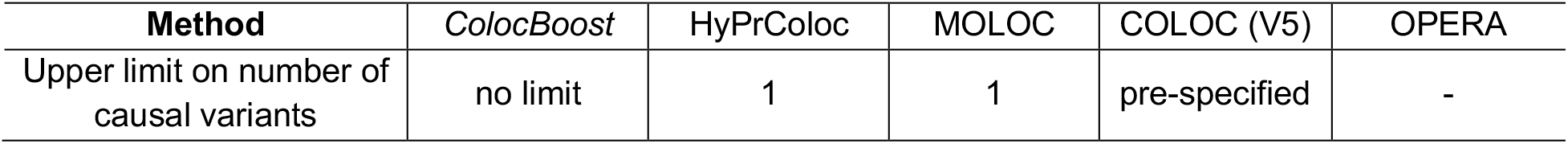
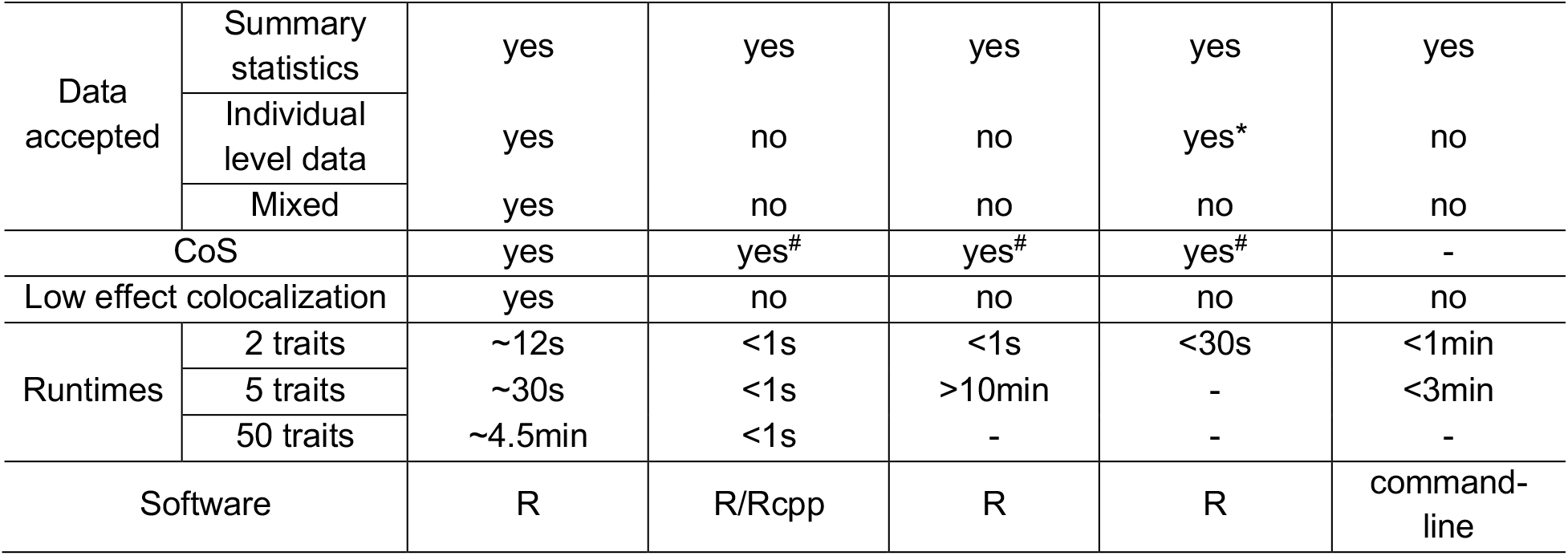
Methodological advantages of *ColocBoost* over existing colocalization approaches. A comparison of *ColocBoost* with four other colocalization methods — COLOC (V5), MOLOC, HyPrColoc and OPERA — focusing on (i) their capacity for multiple causal variants, (ii) support to diverse data-type models, (iii) ability to capture weak, distal xQTL effects, (iv) runtimes and (v) software implementation. Runtimes were assessed for a single simulated locus using a sample size of *N* =1,160 and *P* ≈ 10K variants. For methods based on the summary statistics, the time required to generate them from individual-level simulated data were not included. ^#^indicates that colocalization confidence/credible sets (CoS) were not provided in the original methods but can be computed using available outputs (**Methods**); * COLOC(V5) only supports summary statistics by default although as a two-stage approach it can use SuSiE to process individual-level data, followed by colocalization analysis; - indicates that the method was not applicable to the specified scenario due to its intrinsic limitations.

| Method | *ColocBoost* | HyPrColoc | MOLOC | COLOC (V5) | OPERA |
| --- | --- | --- | --- | --- | --- |
| Upper limit on number of causal variants | no limit | 1 | 1 | pre-specified | - |

|  |  |  |  |  |  |  |
| --- | --- | --- | --- | --- | --- | --- |
| Data accepted | Summary statistics | yes | yes | yes | yes | yes |
|  | Individual level data | yes | no | no | yes* | no |
|  | Mixed | yes | no | no | no | no |
| CoS |  | yes | yes<sup>#</sup> | yes<sup>#</sup> | yes<sup>#</sup> | - |
| Low effect colocalization |  | yes | no | no | no | no |
| Runtimes | 2 traits | ~12s | <1s | <1s | <30s | <1min |
|  | 5 traits | ~30s | <1s | >10min | - | <3min |
|  | 50 traits | ~4.5min | <1s | - | - | - |
| Software |  | R | R/Rcpp | R | R | command-line |

**Figure 1.**
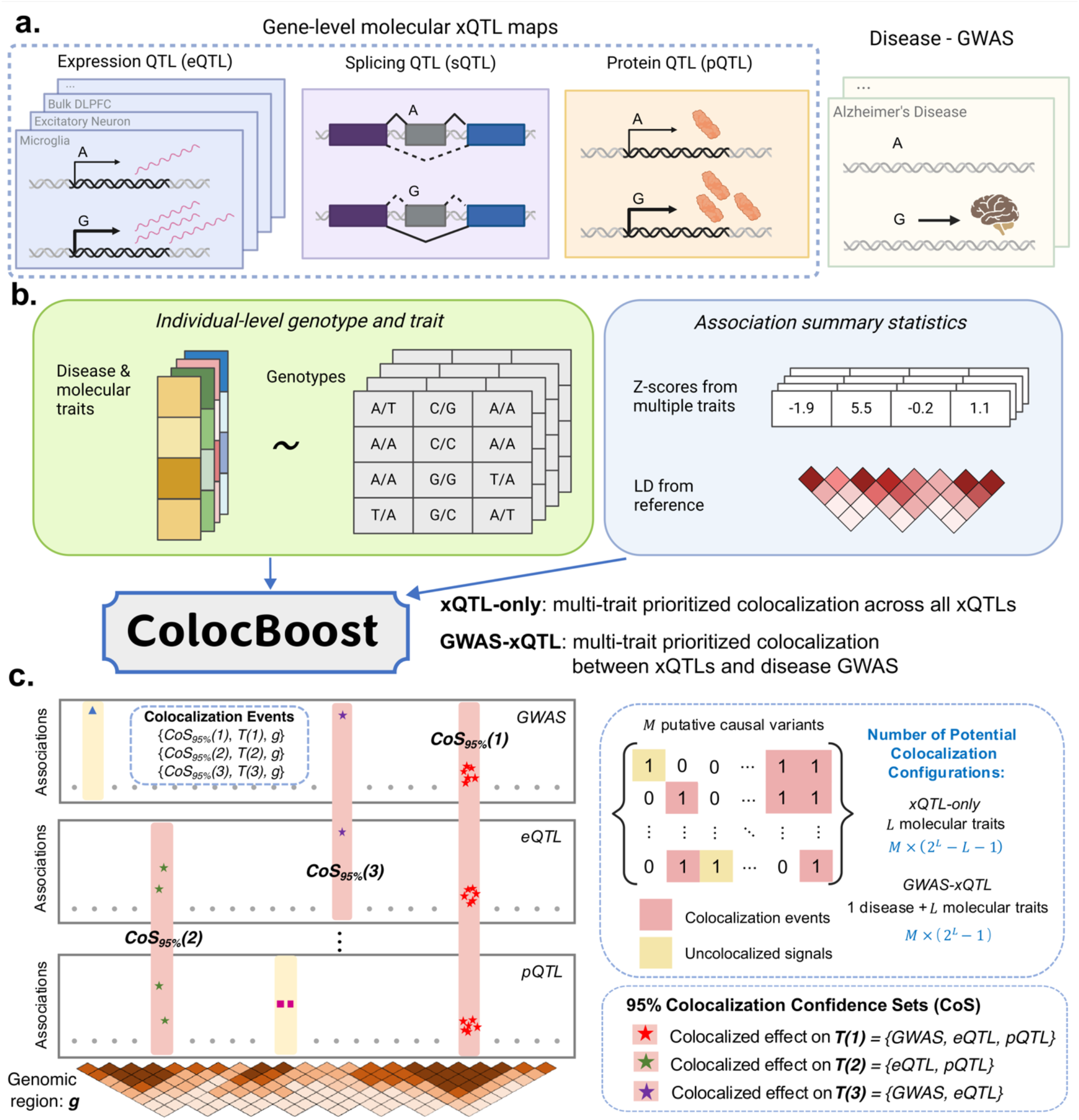
Overview of *ColocBoost* for multi-trait colocalization. **a.** *ColocBoost* uses a multi-task gradient boosting model with proximity smoothing to identify colocalized variants across multiple molecular traits (demonstrated here with gene expression, splicing and protein abundance traits) and when applicable, further integrating GWAS (demonstrated here with Alzheimer’s GWAS). **b**. *ColocBoost* takes as input either (i) individual-level genotype and trait data, or (ii) summary-level association statistics for different traits with a well-matched LD reference panel, and performs analysis in either of the xQTL-only and GWAS-xQTL modes. **c**. By allowing for multiple causal variants (*M*) and molecular traits (*L*), xQTL-only and GWAS-xQTL *ColocBoost* can accommodate up to *M* × (2^*L*^ − *L* − 1) and *M* × (2^*L*^ − 1) potential variant-trait causal configurations. In the schematic, distinct colocalization signals (red) are represented by 95% colocalization confidence sets (CoS) associated with traits sharing this signal, each capturing a putative causal variant with 95% probability. Uncolocalized signals unique to specific traits (yellow) are distinguished by *ColocBoost* from the colocalized events. More details on *ColocBoost* model and algorithm can be found in **Methods** and **Supplementary Figure S1**.

*ColocBoost* employs an iterative gradient-boosting algorithm^35^ with specialized coupled weak learners termed *Single-Effect Couplers* (SEC) that incrementally refine effect estimates at each boosting round (**Figure S1**). For each trait, *ColocBoost* introduces *Proximity Smoothing Gradient Boosting* (PSGB), which replaces hard variant selection with a soft probabilistic assignment across LD-correlated variants (**Methods**). At iteration *k*, a SEC performs this proximity smoothing around a *best update variant* 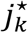 for a set of traits 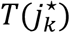 with evidence of *coupled* effects. The effect size update is

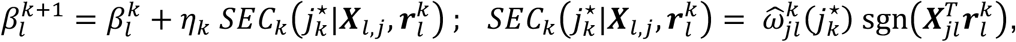

where 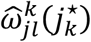 is the iteration-level single-effect conditional inclusion probability; 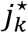 and *l* denote the selected variant and trait indices for iteration *k*. Summing the SEC updates for trait *l* yields 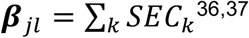. Besides standard SEC, we introduce 1) a *delayed SEC* (D-SEC) to first fit stronger trait-specific signals and then re-evaluate whether the remaining evidence supports a shared effect; and 2) an *expedited SEC* (E-SEC) prioritizes variants associated with the disease trait of interest and improves computational efficiency in the *disease-prioritized* mode (**Methods** and **Supplementary Note**).

At convergence, *ColocBoost* summarizes colocalization evidence at the variant and event levels (**Methods, Figure 1, Figure S1**). For each single-effect component, iteration-level variant probabilities are averaged using their profile log-likelihood improvements and combined across colocalizing traits by logarithmic pooling. *ColocBoost* then integrates evidence across components under a factorized working distribution to obtain a *Variant Colocalization Probability* (VCP) for each variant (**Methods**). Each colocalization event is represented by a causal configuration and a colocalization confidence set (CoS) with default 95% coverage. The CoS retains the candidate causal variant and its high-LD proxies, with construction based on the single-effect colocalization probability shown below.

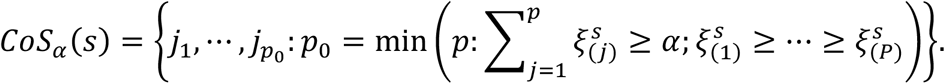

*ColocBoost* filters events with low CoS purity^36^, defined as minimum absolute correlation (|*r*|_*min*_) between all pairs of variants within CoS, or small conditional contributions to the trait-specific profile log-likelihood (**Methods**). CoS and VCP play roles analogous to credible sets and PIP in single-trait fine-mapping^3,38,39^. When *ColocBoost* is applied separately to multiple genes, MaxVCP assigns each variant its highest VCP across genes for downstream annotation analyses^40^.

*ColocBoost* differs from existing methods in its ability to analyze many traits while allowing multiple causal variants at a locus (**Table 1**). Its coupled updates can borrow information across traits when a shared effect is weak in one trait, while CoS retains uncertainty among high-LD variants. The method does not require the maximum number of causal variants to be prespecified. An open-source R package is available under **Code Availability**.

### *ColocBoost* outperforms existing colocalization methods in simulation studies

We compared *ColocBoost* against three established colocalization methods, COLOC (V5) ^13,14^, HyPrColoc^15^, and MOLOC^17^, in a realistic simulation design encompassing multiple scenarios varying in the number of causal variants, variant-trait causal configurations, and effect-size heterogeneity (**Figure S2a-i**). We considered whole-genome sequencing data of 1,162 donors from The Religious Orders Study and Memory and Aging Project (ROSMAP), focusing on *cis* loci (TSS±1.5Mb) of 1,287 unique genes, where each gene was sampled from a distinct topologically associating domain (TAD) out of 1,381 human brain TADs genome-wide^42^. We simulated 2, 5, 10, 20, and 50 molecular traits per gene, ascertaining the effect size of each variant such that it explains a fixed proportion of the trait variance at 5%, and allowing up to five causal variants per trait per locus (**Methods**). For each causal variant, we simulated its colocalization trait configuration by randomly selecting a subset of traits, with the number of colocalized traits determined by the distribution of overlapping genome-wide fine-mapping results from 62 FunGen-xQTL contexts^22^. Statistical power was defined as the proportion of correctly identified 95% CoS that contain the true causal variant, weighted by the number of true colocalizing traits. False discovery rate (FDR) was computed as the fraction of detected CoS that either failed to capture a true causal variant or incorrectly incorporated at least one non-colocalizing trait (**Methods**). Our definitions of power and FDR are intentionally stringent, penalizing both the incorrect assignment of colocalized sets and any misattribution of traits within those sets. Since CoS is not a native metric to competing methods, we extended each method to define the best analogous CoS (**Methods**). Additionally, we conducted variant-level benchmarking by treating variant-level posterior probabilities (from COLOC, HyPrColoc, and MOLOC) or VCP (from *ColocBoost*) as comparable measures of colocalization evidence (**Methods**).

Across different simulation scenarios, *ColocBoost* consistently achieved high statistical power (averaging 0.934) while controlling the FDR below the expected 5% for CoS-level benchmarks (**Figure 2a**). In pairwise colocalization settings with multiple causal variants, *ColocBoost* exhibited 2.9 to 5.2-fold higher power over HyPrColoc and MOLOC respectively, while maintaining significantly lower FDR. *ColocBoost* and COLOC (V5) showed comparable performance under this setting. COLOC’s power varied considerably with the prior colocalization probability *p*_12_, ranging from 0.95 at *p*_12_ = 10^−4^ to 0.40 at *p*_12_ = 10^−7^ (**Figure S2b**); *ColocBoost* achieved power comparable to the highest COLOC value in this grid without a user-specified colocalization prior. For 5, 10, and 20 traits, we compared *ColocBoost* only with HyPrColoc due to scalability constraints of COLOC (V5) and MOLOC (**Table 1**). Even in a one-causal-variant regime which aligns with the HyPrColoc model assumptions, *ColocBoost* attained higher power and lower FDR (**Figure 2a**). Under multiple causal variants, *ColocBoost* further outperformed HyPrColoc, demonstrating up to 73% higher power and 60% lower FDR, with performance gains increasing proportionally to the number of causal variants (**Figure 2a, Figure S2c**). Two representative multiple-causal-variant simulations illustrate settings in which HyPrColoc loses power or error control: (i) heterogeneous effects across traits where HyPrColoc fails to detect either causal variant despite typically being expected to recover at least one under a single-causal assumption; and (ii) a non-causal variant in shared LD with two causal variants and having the strongest marginal association, HyPrColoc admits false positives driven by marginal signal strength (**Figure 2b-i, 2b-ii**). *ColocBoost* can resolve the locus and identify the two causal variants in both cases. We also compared *sizes* and *purity* of 95% CoS among methods as benchmark for how precisely true causal variants are colocalized (**Methods**), and found that all methods produced CoS with average variant counts under 10 and maintained purity above 0.95 (**Figure S2d**). When the number of traits increased to 50, *ColocBoost* continued to show improved power and significantly better controlled FDR compared to HyPrColoc in both single and multiple causal variant scenarios (**Figure S2e**). Additionally, we benchmarked *ColocBoost* against HyPrColoc across four multi-trait simulation designs adopted in HyPrColoc manuscript^15^ (**Figure S2a-ii, Supplementary Note S.6**), along with a fully colocalized scenario where multiple causal variants affect all traits. In all cases, *ColocBoost* outperformed HyPrColoc in power while HyPrColoc showed inflated FDR under multiple causal variants setting (**Figure S2f,g**), particularly in the fully colocalized scenario. As a sanity check, we also evaluated a null scenario with no causal variants across any trait; both methods maintained well-controlled Type I error rates (T1E < 0.005; **Figure S2h**).

**Figure 2.**
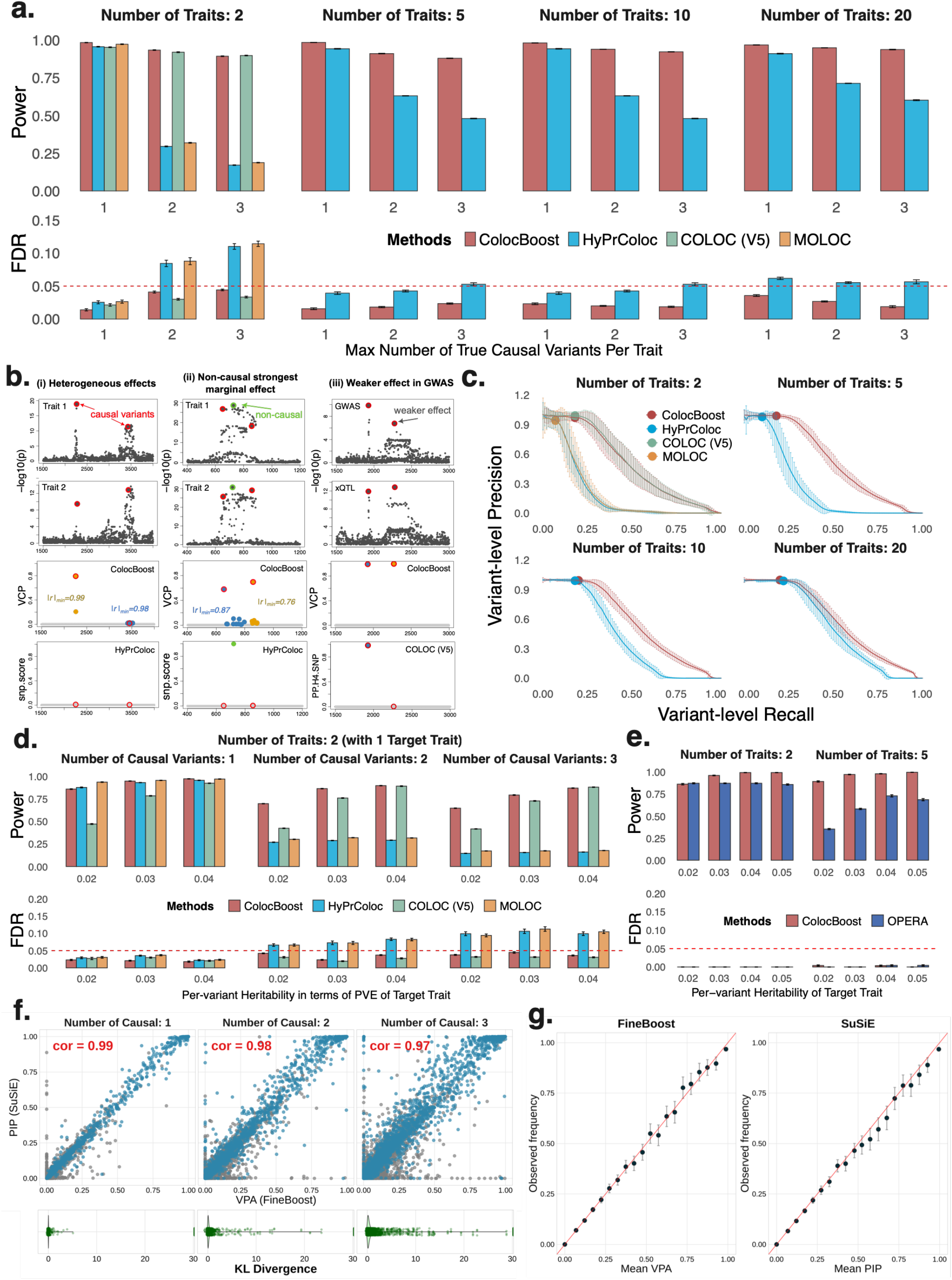
Performance comparison of *ColocBoost* with other multi-trait colocalization methods in simulation benchmarks. **a.** Statistical power and False Discovery Rate (FDR) comparisons of *ColocBoost* against COLOC (V5), MOLOC and HyPrColoc, in simulation settings involving 2, 5, 10, and 20 traits, up to five causal variants per trait per locus, with genotype data and induced colocalization configurations designed to mimic real-world xQTL datasets. Details of power and FDR calculations are provided in **Methods**. The X-axis represents the maximum number of causal variants present within each colocalization configuration across all evaluated traits. Causal variant configurations are calibrated to the observed overlap of fine-mapping credible sets in the FunGen-xQTL atlas, where most causal variants are shared across only a subset of traits and configurations with 4-5 shared variants are rare. Results are displayed only for configurations with >10% of total replicates to avoid reporting power and FDR estimates from sparsely represented configurations. **b**. Representative simulation examples illustrating limitations of competing methods: HyPrColoc with “one-causal-variant-per-trait” assumption fails to detect causal signals under (i) heterogeneous effects distribution across traits and (ii) where another non-causal variant with shared LD with causal variants has strongest marginal effect; (iii) COLOC (V5) shows reduced sensitivity to weak causal effects in the “disease-like” trait. **c**. Variant-level precision-recall curves by varying the colocalization score threshold (*ColocBoost* uses VCP; COLOC/HyPrColoc/MOLOC use variant-level scores from their respective methods; **Methods and Supplementary Note S.6**). **d**. Statistical power and FDR comparisons for disease-prioritized mode of *ColocBoost* (GWAS-xQTL) where there is a simulated “disease trait”, for which we lower the per-SNP heritability in the locus relative to other molecular traits to reflect expectations from real world applications. **e**. Statistical power and FDR comparison of *ColocBoost* with OPERA for GWAS colocalization, evaluated at the gene level (**Methods**). **f**. Evaluation of variant-level probability of association (VPA) for *FineBoost* (special case of *ColocBoost* with *L* = 1 for single-trait fine-mapping analysis). We compare VPA against PIP computed by SuSiE, where each point depicts a single variable in one of the simulations with true effect variable (blue) and no effect variable (grey). **g**. Calibration of VPA in *FineBoost* and PIP in SuSiE. Variants across all simulations were grouped into bins according to their reported value. The plots show the average VPA or PIP for each bin against the proportion of effect variants in that bin. For each bin, the x-axis shows the mean reported probability and y-axis shows the fraction of variants that are truly causal in the simulations. A well-calibrated method should produce points near the *x* = *y* line. The red dashed line in panels **a, d**, and **e** denote the FDR threshold 0.05 corresponding to 95% CoS. Numerical results are reported in **Supplementary Data**.

Next, under the primary simulation design informed by FunGen-xQTL data, we evaluated variant-level performance of the above methods by comparing the precision-recall curves generated by varying the threshold of variant-level colocalization probability from 0 to 1 and computing area under the precision-recall curve (AUPRC) (see **Methods** for variant-level probability scores of each method). *ColocBoost* outperformed HyPrColoc and MOLOC, achieving 2.56 ± 0.22 and 2.48 ± 0.22-fold higher AUPRC respectively across all simulation scenarios (**Figure 2c**). In the two-trait scenario, *ColocBoost* showed similar precision-recall as COLOC (*p* = 0.41, two-sided *t*-test). While variant-level comparisons are informative for benchmarking, we caution against focusing on a single top-ranked variant for any colocalization method: in regions of strong LD, such a variant can be merely a proxy of the true causal variant, thus constituting false discoveries at variant level (**Figure S3a**). Consequently, we recommend evaluations based on 95% CoS as they offer more robust inference by accounting for uncertainties due to LD.

In practice, it is often of interest to colocalize a disease GWAS with multiple molecular QTL traits to elucidate the functional basis of disease associations. An important technical aspect of this colocalization is that most GWAS traits tend to have significantly lower per-variant contribution to heritability compared to xQTL on molecular traits. To simulate this scenario, we create a “disease-like trait” with per-variant heritability varying between 2% and 4% (while such per-variant heritability is relatively large for a disease GWAS, we note that real-world GWAS typically benefits from much larger sample sizes to detect weaker effects with comparable power). We then applied *ColocBoost* in the GWAS-xQTL mode using E-SEC on the disease trait. In the two-trait scenario with high per-variant heritability, *ColocBoost* showed similar performance with COLOC (V5). However, when the disease trait had lower per-variant heritability, *ColocBoost* demonstrated greater advantage in both power and FDR, demonstrating its capacity to detect subtler signals (**Figure 2d**; with an illustrative example in **Figure 2b-iii**). We next compared *ColocBoost* to the recently proposed OPERA^20^ method designed for GWAS-centric multi-trait colocalization. Since OPERA only offers gene-level outputs, we defined a parallel gene-level metric for *ColocBoost* to enable a fair comparison (**Methods**). While both methods controlled FDR well below 0.05 in this setting, GWAS-xQTL *ColocBoost* achieved up to 2.53-fold higher power compared to OPERA, with performance gains increasing proportionally to the number of traits and the number of causal variants involved (**Figure 2e, Methods**). Moreover, *ColocBoost* demonstrated improved power (at well controlled FDR) over OPERA under an additional simulation setting where OPERA has been reported to show excellent performance^20^ (**Figure S2a-ii, Figure S3b**).

To additionally assess calibration of the single-trait variant probability when *ColocBoost* reduces to *FineBoost* (**Supplementary Note S.1.4; Figure S4**), we compared *FineBoost* variant-level probability of association (VPA) with SuSiE’s posterior inclusion probability (PIP) across simulations with one to five causal variants. VPA and PIP were highly concordance (correlation>0.97), and for both methods, the fraction of truly causal variants closely matched the reported probabilities across probability bins, indicating good empirical calibration (**Figures 2f–g**). These results support calibration of VPA in the single-trait setting, consistent with the case of VCP evaluated through previous multi-trait simulations.

Overlapping samples are common in xQTL analyses spanning multiple modalities from donors of the same cohort. Since this can induce dependencies (correlated residuals independent of genetic effects) across traits, in principle, accounting for such correlations may improve method performance. However, Foley *et al*.^15^ showed that treating studies as independent typically yielded controlled FDR and robust power in capturing colocalizing traits while conferring an advantage of reduced computational complexity. To validate this, we considered extensions of the multi-trait simulation design in **Figure 2a**, incorporating realistic cross-trait correlations estimated from FunGen-xQTL data (**Figure S2a-iii, Supplementary Note S.6**). Despite not modeling these correlations by default, *ColocBoost* maintained comparable power and FDR across all simulation scenarios (**Figure S3c,d**). While we recommend the current *ColocBoost* implementation for its balance of performance and scalability to many traits, we provide technical details to incorporate sample overlap adjustments within *ColocBoost* (**Supplementary Note S.2.5**).

Finally, we performed three secondary analyses to assess *ColocBoost* under specialized or less typical circumstances. First, we considered a scenario where each trait has available association summary statistics but not the matching LD reference. In this scenario, like fine-mapping, a single-causal-variant assumption without LD-proximity smoothing was used for colocalization, under which *ColocBoost* continued to exhibit substantially better FDR control with slight loss of power compared to HyPrColoc (**Supplementary Note S.2.5**; **Figure S3e**). Second, we explored a version of *ColocBoost* with only one trait, referred to as *FineBoost*, to compare its fine-mapping performance with SuSiE^38^. Even though *FineBoost* is primarily a special case of *ColocBoost* rather than a dedicated fine-mapping tool, it matched SuSiE in power while more effectively controlling FDR in scenarios featuring multiple (up to five) causal variants, thereby underscoring its robustness as a statistically rigorous model for both colocalization and single-trait genetic association studies (**Figure S3f**). Third, we tested different choices of the probabilistic simplex ***ω***_*l*_ by varying the way in which it integrates marginal association evidence with proximal LD. We observed an overall robust performance of *ColocBoost* to these choices, with slightly better performance under the default choice (**Methods; Figure S5**).

We conclude that *ColocBoost* demonstrates robust performance over existing colocalization methods across diverse simulation designs, including multiple causal variants, large sets of traits, complicated colocalization patterns, lower causal effect sizes, overlapping samples, and absence of LD data.

### Colocalization performance in split-sample analyses

We evaluated colocalization robustness and reproducibility through independent random splits in two real-data settings (**Supplementary Note S.10**). First, we divided the ROSMAP DLPFC eQTL samples (*n* = 784) into two and three non-overlapping subsets and colocalized each subset with the same AD GWAS. In the two-way split, both COLOC and *ColocBoost* identified fewer loci than in the complete dataset as expected, and their full-data recovery rates were nearly identical (87%). However, *ColocBoost* recovered more full-data loci (41 vs. 20; **Figures S6a-b**). Relative to 95% CS from SuSiE fine-mapping on the complete eQTL dataset, *ColocBoost* recovered 50.2% of all CS and 59.6% of single-CS gene regions, compared with 36.0% and 40.9% for COLOC (**Figure S6c**). Between the splits, *ColocBoost* identified nearly twice as many reproducible loci as COLOC (26 vs. 14 shared loci), while maintaining high average CoS purity (>0.94). Under the three-way split, *ColocBoost* recovered 55.6% of all complete-data CS and 48.3% of single-CS gene regions, compared with approximately 40% and 30% for SuSiE fine-mapping applied to the individual subsets. Second, we divided the UK Biobank LDL cholesterol GWAS sample into two subsets and colocalized each subset with the same GTEx liver eQTL data. *ColocBoost* recovered 65.7% of full-data loci compared with 51.5% for COLOC and identified more cross-split shared loci (30 vs. 19) (**Figure S6d**).

### xQTL-only *ColocBoost* decodes molecular regulatory architecture in aging brain

We conducted a genome-wide xQTL-only *ColocBoost* analysis across 17 cis-xQTL traits from the aging brain cortex of ROSMAP subjects (average *N*=595; **Table 2**)^22,24,43–45^, spanning 16,928 genes and three gene-level molecular trait modalities including gene expression, splicing and protein abundance. Our data consist of 6 pseudo-bulk expression QTL (eQTL) datasets derived from different cell types in brain dorsolateral pre-frontal cortex (DLPFC), 4 bulk eQTL datasets spanning three brain cortical regions and CD14^+^/CD16^-^ monocytes from peripheral blood mononuclear cells (PBMCs), 6 bulk splicing QTL (sQTL) datasets comprising both productive and unproductive splicing in the same three cortical regions^46^, and one protein abundance QTL (pQTL) dataset from DLPFC^22^ (**Methods, Table 2**). Overall, *ColocBoost* identified 18,654 colocalization events involving 11,257 genes and 16,504 distinct 95% CoS (**Table S1**). At least one colocalization event was detected for 66.5% genes; of these, 27.6% represented cross-modality colocalization bridging at least two of the three modalities, and 2.3% spanned all three modalities (**Figure 3a**). A large fraction (61.8%) of colocalization events were shared across more than two traits (**Figure 3b, Figure S7a**). eQTLs and sQTLs contributed to more colocalization events compared to pQTLs, partly due to protein abundance data being available for only 7,641 of the 16,928 genes (**Table 2**). Restricting the analysis to 3,655 genes profiled across all three modalities yielded more balanced colocalization patterns, though eQTLs and sQTLs still demonstrated more colocalizations than pQTL (**Figure S7b**).

**Table 2.**
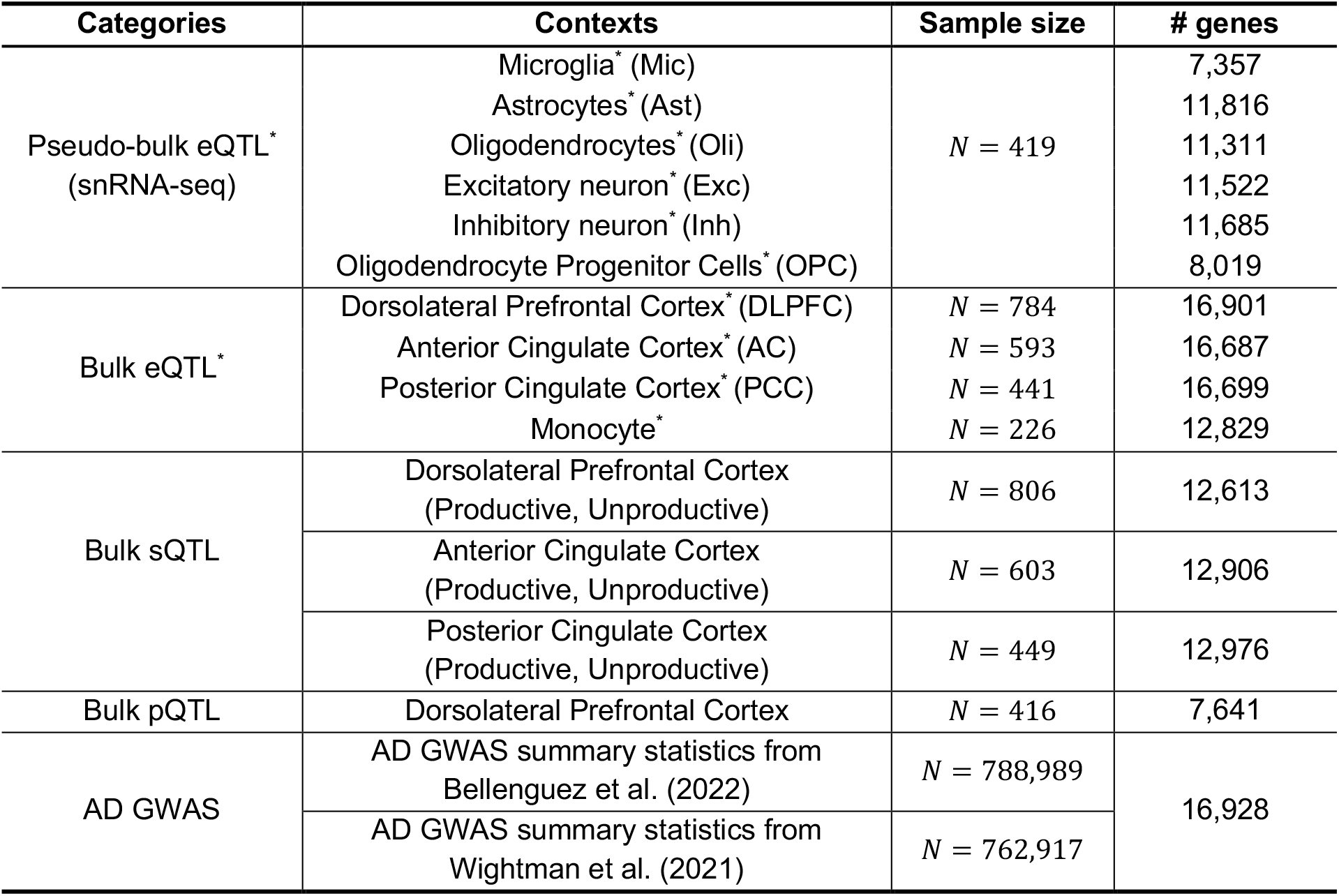
Overview of 17 *cis*-xQTL datasets from ROSMAP brain cortex. Each row lists the molecular trait modalities and GWAS, abbreviated dataset name, cell types or brain regions corresponding to the xQTL data, and sample sizes analyzed for each dataset. Asterisks (*) mark modalities included in eQTL specific colocalization analysis as an ablation study.

| Categories | Contexts | Sample size | # genes |
| --- | --- | --- | --- |
| Pseudo-bulk eQTL*<br>(snRNA-seq) | Microglia* (Mic) | N = 419 | 7,357 |
|  | Astrocytes* (Ast) |  | 11,816 |
|  | Oligodendrocytes* (Oli) |  | 11,311 |
|  | Excitatory neuron* (Exc) |  | 11,522 |
|  | Inhibitory neuron* (Inh) |  | 11,685 |
|  | Oligodendrocyte Progenitor Cells* (OPC) |  | 8,019 |
| Bulk eQTL* | Dorsolateral Prefrontal Cortex* (DLPFC) | N = 784 | 16,901 |
|  | Anterior Cingulate Cortex* (AC) | N = 593 | 16,687 |
|  | Posterior Cingulate Cortex* (PCC) | N = 441 | 16,699 |
|  | Monocyte* | N = 226 | 12,829 |
| Bulk sQTL | Dorsolateral Prefrontal Cortex<br>(Productive, Unproductive) | N = 806 | 12,613 |
|  | Anterior Cingulate Cortex<br>(Productive, Unproductive) | N = 603 | 12,906 |
|  | Posterior Cingulate Cortex<br>(Productive, Unproductive) | N = 449 | 12,976 |
| Bulk pQTL | Dorsolateral Prefrontal Cortex | N = 416 | 7,641 |
| AD GWAS | AD GWAS summary statistics from<br>Bellenguez et al. (2022) | N = 788,989 | 16,928 |
|  | AD GWAS summary statistics from<br>Wightman et al. (2021) | N = 762,917 |  |

**Figure 3.**
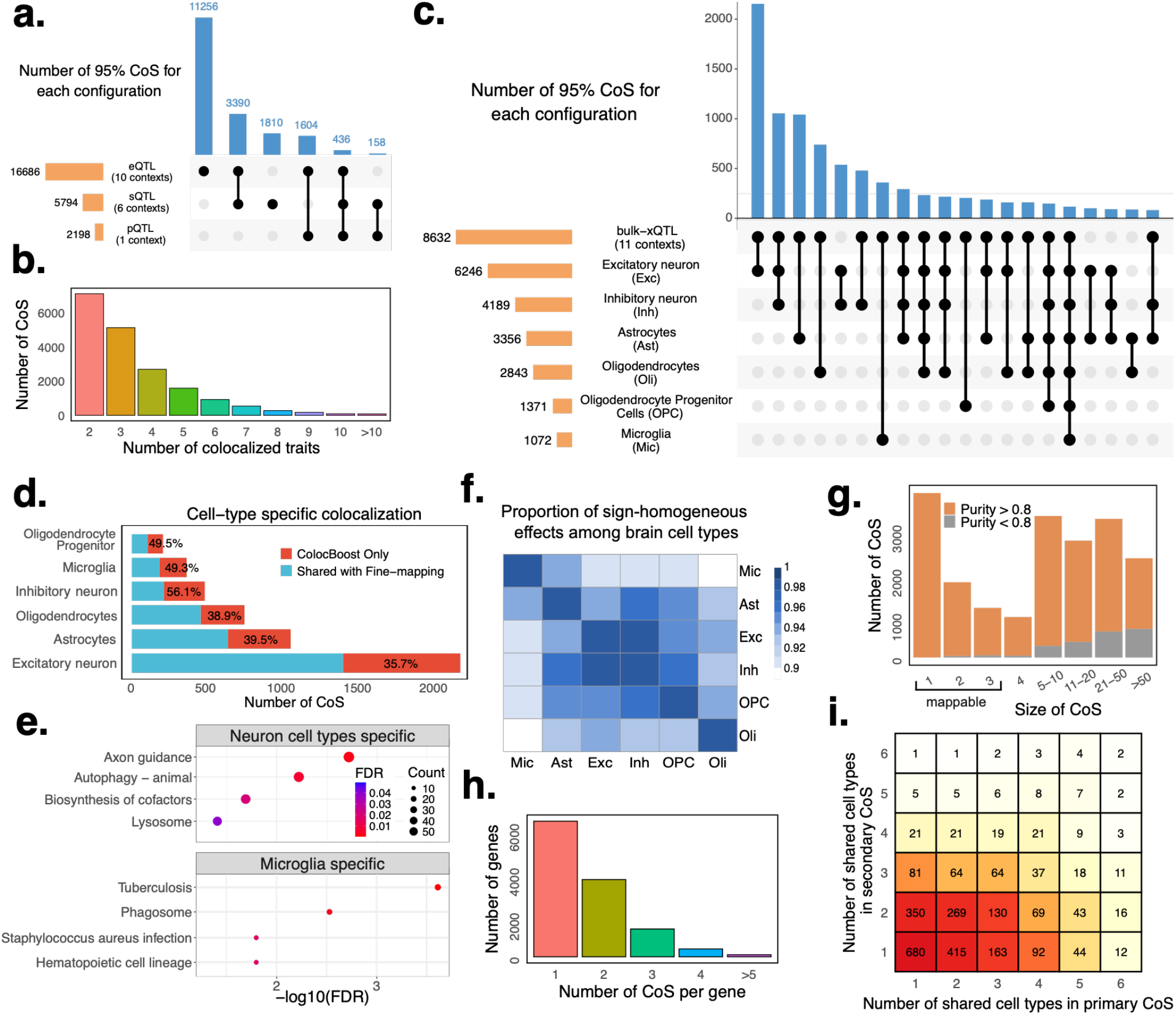
*ColocBoost* xQTL analysis across cell types and traits modalities. *ColocBoost* was applied to 17 gene-level cis-xQTL datasets from the aging brain cortex of ROSMAP subjects (average *N* = 595) spanning 16,928 genes. **a.** UpSet plot summarizing the colocalization patterns across three different molecular trait modalities (expression, splicing, protein abundance). See **Table 2** for details of each modality. **b**. Distribution of the number of 95% CoS corresponding to the different numbers of colocalized traits per locus; we omit from consideration 0.55% loci with more than 10 colocalized traits. **c**. UpSet plot summarizing the colocalization patterns across 6 pseudo-bulk brain cell-type eQTL data, along with bulk-xQTL data. We highlight putative causal variants with (i) shared effect across multiple brain cell types, and (ii) cell-type specific effect obtained through colocalization between bulk xQTL and pseudo-bulk eQTL for that cell type. **d**. The fraction of cell-type specific colocalizations (measured at the level of CoS) for different brain cell types recovered by fine-mapping credible set within the cell type. **e**. Top 4 significantly enriched (FDR<0.05) pathways (using enrichKEGG^58^) for eGenes linked to cell-type specific colocalizations in excitatory neurons and microglia. Size of the circle denotes the number of eGenes matched to the pathway and the color denotes the level of FDR-adjusted significance. **f**. For each pair of brain cell types, we assess the homogeneity in estimated causal effects directions by measuring the proportion of colocalized signals exhibiting concordant effect size signs. **g**. Empirical distribution of the number of 95% CoS in terms of the number of variants each CoS contained. CoS are color-coded by *purity*, with high-purity sets (*purity* > 0.8) distinguished from moderate-purity sets (0.5 < *purity* ≤ 0.8). **h**. Distribution of the number of CoS per gene; only 0.8% genes harbor more than four colocalized sets. **i**. For each gene with multiple CoS, we consider a pair of primary CoS and secondary CoS, and evaluate the number of brain cell type pseudo-bulk eQTLs showing shared effects for each CoS in the pair. We present a heatmap of the number of pairs of primary and secondary CoS for the same gene, showing different patterns of sharing across brain cell type eQTLs. Numerical results are reported in **Supplementary Data**.

Next, we focused on xQTL-only colocalization events where the 95% CoS included at least one eQTL from a specific brain cell type. Of the 18,654 total events identified, 10,237 (55%) involved at least one cell-type eQTL (**Figure 3c**). Within this subset, 4,973 events (48.6%) featured colocalization between a single-cell-type eQTL and one or more bulk xQTL. Overall, 27.9% of these cell-type-specific events mapped to microglia or astrocytes, whereas 54.1% involved neuronal subtypes (**Figure 3c**). Notably, 35.7–56.1% of these cell-type specific colocalization signals were not captured by the matched cell-type eQTL fine-mapping using SuSiE (**Figure 3d, Figure S7c**), while on the other hand, 58.7–76.6% of fine-mapped cell-type eQTLs were identified to colocalize with other xQTL (**Figure S7c**). This underscores the utility of integrating bulk xQTL data with pseudo-bulk cell-type level data in boosting detection power for weaker cell-type level signals, as previously demonstrated for eQTL mapping^47^. To evaluate the impact of modality beyond eQTL, we conducted an ablation *ColocBoost* analysis that removed all sQTL and pQTL datasets, retaining only the 10 eQTL contexts (6 cell-type eQTL and 4 bulk eQTL). Although a large fraction (82.1%) of these cell-type involved colocalizations can be detected by including only bulk eQTL without sQTL or pQTL (**Figure S7d**), the remaining 17.9% represent regulatory signals that could only be identified through integrated multi-omics analysis. eGenes corresponding to neuron-specific colocalization showed significant enrichment in axon guidance pathways, while microglia-specific signals showed enrichment in infectious disease pathways^48,49^ (**Figure 3e; Table S2**); microglia are known to be equipped with receptors to respond to detect and respond to a wide array of pathogens^50^. We also observed a 1.36 to 10.32-fold excess-of-overlap (EOO) of the cell-type specific colocalization eGenes with cell-type gene programs from external brain single-cell RNA-seq data^25,51,52^ (**Methods**; **Figure S7e**). Among the 5,264 colocalization events spanning multiple cell types, excitatory and inhibitory neurons jointly appeared in 61.8% of them, with 99.5% of these neuronal-shared events maintaining consistent causal effect-size directions (**Figure 3f**). 132 colocalization events showed coupling across all six brain cell types. 69.5% of multiple cell type colocalizations involved a bulk QTL; ablating these bulk xQTLs resulted in a mere 3.6% reduction in multiple cell-type colocalizations (**Table S1**).

Among all *ColocBoost* 95% CoS, 3,712 (19.9%) consists of a single variant and 6,532 (35%) contain three or less variants, which we refer to as *mappable* CoS (**Figure 3g**). Variants in these low-complexity CoS exhibit strong functional enrichment: 10.6-fold in cV2F regulatory score^53^ and 3.31-fold in RegulomeDB annotations^54,55^ relative to the remaining CoS (**Figure S8a**). 84.5% of baseline-LD v2.2 annotations^56^ are significantly enriched in *mappable* CoS^57^ (**Figure S8b** where p-values of enrichment are estimated by Jackknife method through a leave-one-chromosome-out strategy). Notably, 45.9% of genes contain two or more colocalized events, highlighting the importance of a colocalization method capable of accommodating multiple causal variants (**Figure 3h**). For each gene, we designated the 95% CoS associated with the highest increase in profile log-likelihood as the *primary* colocalization and others as *secondary* (**Methods, Figure S9a-b**). We observed that secondary CoS were on average 2.24 times more distal from gene TSS and 44.5% more cell-type-specific compared to primary CoS (**Figure 3i**). Moreover, primary CoS showed significantly higher coding and promoter enrichment, especially among variants with MaxVCP > 0.5 (**Figure S9c**). As an illustrative example, we present *ARSB* featuring two distinct colocalization loci: the primary CoS comprises a single variant with shared effect across neuron and glial cells, whereas the secondary CoS comprises of eight variants and shared only among glial cells (**Figure S9d**). These discrete regulatory events suggest that *ARSB* expression may be modulated by multiple, partially independent mechanisms across different cell types.

As a secondary analysis, we applied *ColocBoost* to eQTL data from 13 GTEx brain tissues with moderate sample size (*N* =168, **Table S3**). Of the colocalizations identified in three cortical GTEx tissues—brain cortex, anterior cingulate cortex (BA24), and frontal cortex (BA9)—a substantial fraction (46.9%) overlapped with ROSMAP, whereas only 10.4% of ROSMAP colocalized events appeared in GTEx (**Figure S10)**. This asymmetry likely arises from three factors: (i) larger sample size in ROSMAP, which increases power to detect weaker QTL, and (ii) the cell-type-level resolution available in ROSMAP, which can reveal additional layers of regulatory variation masked by bulk tissue data, and (iii) differences in donor age, with ROSMAP consisting of an aging population (average age: 88 years) versus the younger GTEx cohort (average age: 53.5 years), potentially capturing additional age-related genetic effects.

### Colocalized xQTL are concordant with CRISPR and enhancer-gene maps

We systematically evaluated the concordance of CoS-gene maps implicated by *ColocBoost* based on the enrichment for regulatory element-gene links derived from orthogonal functional assays from cell lines. These benchmarking datasets include (i) 569 element-gene links from aggregated CRISPRi data (KRAB-dCas9) of 3 studies^29–31^ as used in Gschwind et al.^34^, (ii) 8,192 element-gene links from the recent STING-seq assay using an updated CRISPRi protocol (KRAB-dCas9-MeCP2)^29^ covering 500 GWAS loci across 44 blood-related traits, and (iii) 118,389 regulatory element-gene maps from Promoter Capture Hi-C assay in the dorsolateral prefrontal cortex brain tissue^32,33^, which profiles targeted 3D contacts linking gene promoters to proximal or distal regulatory elements. Specifically, we constructed 95% CoS-gene links and compared them with (i) all SNP-Gene links derived from conventional xQTL association analysis and (ii) SuSiE fine-mapped 95% credible set (CS)-gene links, merged across all 17 ROSMAP xQTLs (**Figure 4a, Methods**). CoS-gene pairs from xQTL-only *ColocBoost* exhibited a high 15.42-fold (*p* = 8 × 10^−4^) and 6.38-fold (*p* = 4.8 × 10^−5^) variant set-level EOO with aggregated CRISPRi and STING-seq links in K562 respectively, significantly exceeding the EOO observed for standard xQTL SNP-Gene and fine-mapped CS-gene links (one-sided *t*-test of difference, *p* = 0.0024 and 0.027 respectively) (**Methods**; **Figure 4a**). Consistent with these findings, *ColocBoost* CoS outperformed xQTL CS by 1.73-fold EOO in brain cortical PCHi-C (**Figure S11a**).

**Figure 4.**
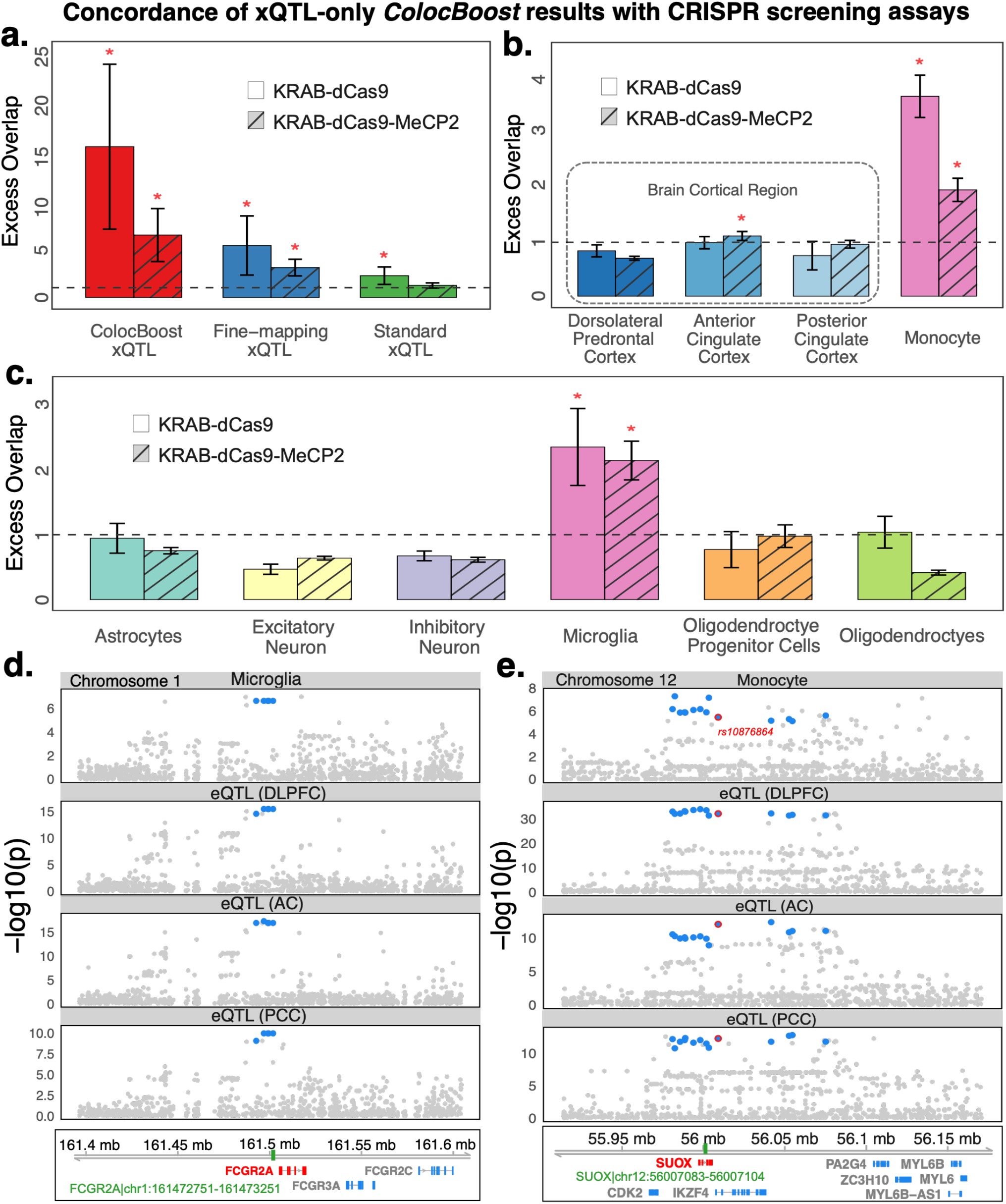
Validation of ColocBoost colocalization signals using CRISPR data. **a.** Variant set-level excess-of-overlap (EOO) analysis of (i) 95% CoS-gene links from xQTL-only ColocBoost, (ii) standard marginal xQTL-gene associations (FDR<0.05), merged across all 17 xQTL datasets and (iii) 95% credible set (CS)-gene links from SuSiE fine-mapping, merged across all 17 xQTL datasets (**Methods**), against 569 silver-standard element-gene links from 3 aggregated CRISPR interference (CRISPRi) datasets in K562 cell line using KRAB-dCas9 protocol, and 8,192 silver-standard element-gene links from the recent STING-seq data in K562 using a newer KRAB-dCas9-MeCP2 protocol. **b, c**. Relative EOO analysis of 95% CoS-gene links across **b**. 3 different brain cortical regions and CD14+CD16-bulk monocytes, and **c**. 6 different brain cell types, against element-gene links from aggregated CRISPR (dCas9) and STING-seq datasets. Red asterisks denote significant enrichment based on EOO test adjusted for multiple testing (Bonferroni adjusted p-value<0.05). **d**. As an illustration, a CoS-gene link from microglia-specific eQTL colocalization in *FCGR2A* is validated by CRISPRi in K562 cells. **e**. A second CoS-gene link, reflecting colocalization between monocyte eQTLs and bulk brain region data at the *SUOX* gene, is similarly validated by CRISPRi in K562 cells. The green box denotes the CRISPRi enhancer linked to the target gene. Numerical results are reported in **Supplementary Data**.

Overall, 53 CoS-gene links were validated by at least one of the two CRISPR datasets, of which 15 (28.3%) involved immune cell type eQTLs in microglia or monocyte among the coupled traits (**Figure 4b,c, Table S4**). CoS-gene links in 3 brain cortical regions did not show significant EOO relative to all CoS-gene links; however, CoS-gene links in CD14+/CD16- monocytes showed high enrichment (EOO=3.71, *p* = 1 × 10^−41^ in CRISPRi and EOO=1.97, *p* = 8 × 10^−19^ in STING-seq; **Figure 4b**). Concordant with this observation, out of the 6 brain cortical cell types, only CoS-gene links involving microglia were uniquely enriched (EOO=2.35, *p* = 8 × 10^−6^ in CRISPRi and EOO=2.14, *p* = 6 × 10^−14^ in STING-seq; **Figure 4c)**. This confirms the expected regulatory concordance between K562 and immune cell types in the brain, while also highlighting its discordance with broader brain cell types. Although K562 is an erythroleukemia cell line that does not closely match most brain cell types, it remains the most extensively studied line for CRISPR screening assays containing hundreds of element-gene links, providing reasonable statistical power for benchmarking purpose in our work. We assessed concordance between CoS-gene links involving different brain cell types and bulk brain regions against the brain PCHi-C links, and found significant relative EOO in oligodendrocyte progenitor cells and oligodendrocytes, but not in any bulk brain region (**Figure S11b,c**).

An illustrative example of the concordance between *ColocBoost* and CRISPR data is the 95% CoS *chr1:161493252-161502998*, comprising of 5 variants mapped to *FCGR2A*^59,60^. It overlaps the regulatory element identified by CRISPRi (*chr1:161502961-161503461*) mapped to the same gene (**Figure 4d**). The CoS is microglia specific among single cell contexts—consistent with the role of *FCGR2A* encoding a cell-surface receptor involved in clearing immune complexes^61^—and propagates across all 3 bulk brain region eQTLs,. Another example of such concordance is a relatively broader 95% CoS mapped to *SUOX*, with 14 variants spanning *chr12:55977282-56078047* which contains the element *chr12:56007083-56007104* mapped to the same gene by CRISPRi. The CoS spans 9 molecular traits, comprising of bulk eQTL data from monocytes and 3 brain cortical regions, alternative splicing QTLs, as well as with other brain cell type eQTLs (**Figure 4e, Figure S11d**). One variant *rs10876864* (*chr12:56007301:G:A*) in the CoS has been previously associated with autoimmune^62^ and allergic conditions^63^.

### Colocalized xQTL variants are enriched in complex disease heritability

Genome-wide MaxVCP scores derived from our multi-omics colocalization map can serve as a new variant-level functional annotation score. To evaluating its informativeness across a broad range of complex diseases, we annotated 9,991,229 variants with minor allele counts≥5 in the 1000 Genomes Project Europeans using 5 different MaxVCP derived features: (i) *MaxVCP*-*xQTL*, based on xQTL-only *ColocBoost* analysis of all 17 ROSMAP xQTL datasets; (ii) *MaxVCP*-*eQTL*, restricting the analysis to 10 eQTL traits; (iii) *MaxVCP*-*pseudobulk*, restricting the analysis to 6 cell-type pseudo-bulk eQTLs; (iv) *MaxVCP*-*bulk*, restricting the analysis to bulk eQTLs in three brain regions; and (v) *MaxVCP*-*GTEx*, obtained by applying xQTL-only *ColocBoost* to 13 GTEx brain tissues (**Methods, Table S5**). These five annotations showed moderate inter-correlation (average *r*=0.39; **Figure S12a**). To assess their relevance to disease heritability, we applied Stratified LD Score Regression^27^ (S-LDSC) across 57 relatively independent (*r*_*g*_<0.7) complex diseases and traits (average GWAS sample size *N*=305K), including 22 blood-related and 18 brain-related traits (**Table S6**). We compared the heritability enrichment and the standardized effect sizes (*τ*^∗^) of these annotations conditional on 97 baseline-LD v2.2 annotations, comprising coding, conserved, regulatory, and LD-linked features^56,64^ (**Methods**).

All five MaxVCP annotations attained high heritability enrichments in meta-analysis across all 57 traits combined (average 10.6±0.74-fold), as well as across brain-related traits (**Figure 5a)**. 4 out of 5 MaxVCP annotations—except MaxVCP-pseudobulk—showed significant heritability enrichment for blood traits meta-analysis. When conditioned on the baseline-LD annotations, MaxVCP-xQTL attained the highest *τ*^∗^ (0.51 and 0.50 respectively) compared to other MaxVCP annotations (annotations with *τ*^∗^ ≥ 0.5 are generally considered to be highly impactful for disease^40,65^). In a subsequent joint heritability enrichment analysis including five MaxVCP annotations, MaxVCP-xQTL remained significant with moderate to high values of joint *τ*^∗^—0.33, 0.51 and 0.46 for all traits, brain traits, and blood traits meta-analysis respectively. The consistently stronger heritability signal for MaxVCP-xQTL over other MaxVCP scores highlights the additional advantage conferred by integrating multiple QTL modalities and contexts. We next compared the heritability information of MaxVCP-xQTL against an analogous score from similarly applying HyPrColoc to all modalities of xQTL data (MaxPPH-xQTL). When considered individually, both annotations were enriched for disease heritability and exhibited high marginal *τ*^∗^ (Bonferroni corrected *p* < 0.05) conditioned on the baseline-LD annotations (**Figure S12b**). However, in the joint analysis of the two annotations, only MaxVCP-xQTL remained significant in all traits and brain traits meta-analyses (joint *τ*^∗^ = 0.45 and 0.52 respectively; **Figure 5b**), effectively subsuming the information MaxPPH-xQTL captured. In comparison, for blood traits meta-analyses, both the annotations showed comparable joint *τ*^∗^ signal.

**Figure 5.**
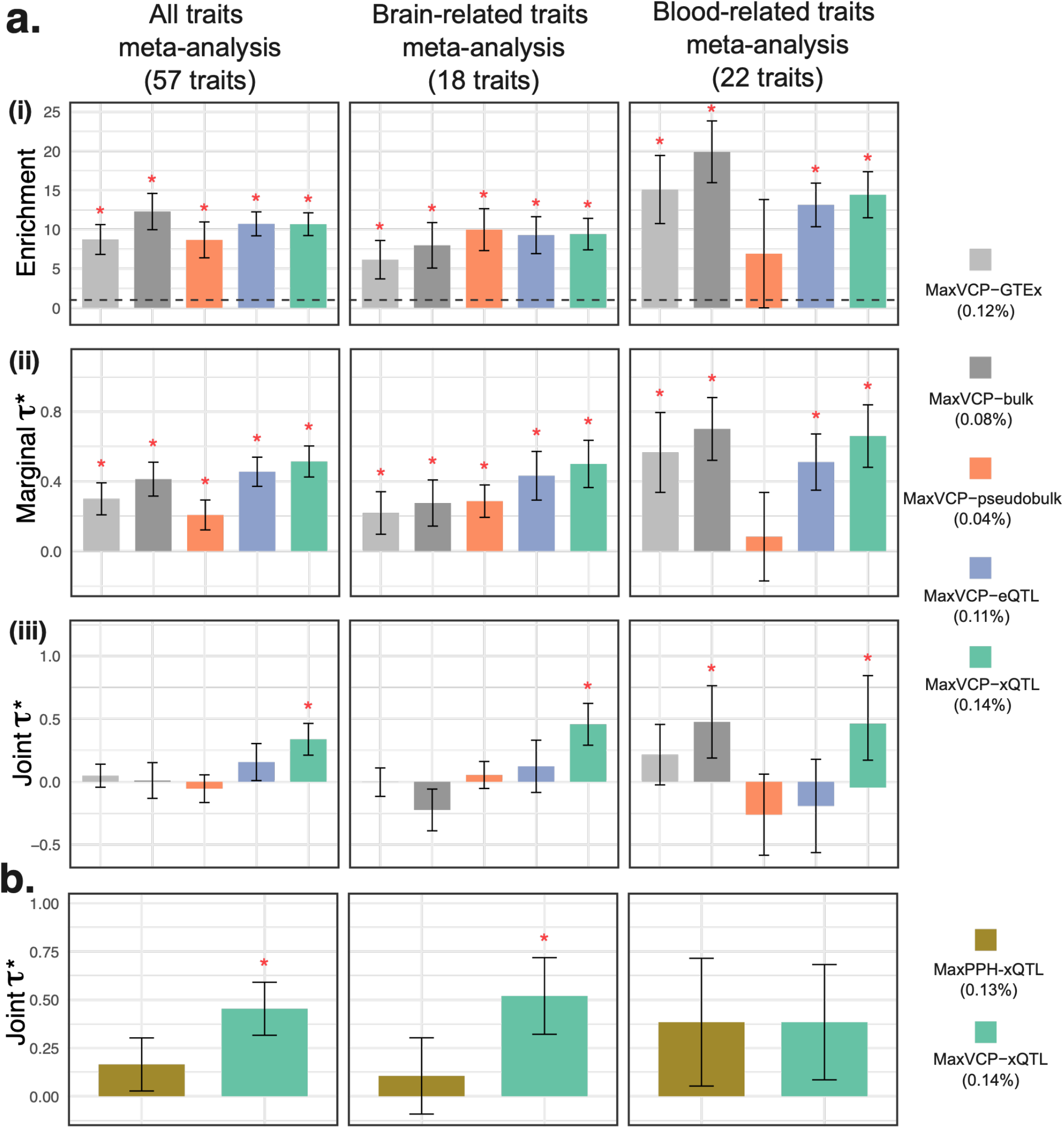
Disease heritability analyses of variant-level functional annotations derived from *ColocBoost*. **a.** We generated 5 MaxVCP variant-level functional annotation scores by performing xQTL-only *ColocBoost* (**Results**) and performed S-LDSC heritability analysis of the resulting annotations. **(i)** Heritability enrichment conditional on 97 baseline-LD v2.2 annotations. **(ii)** Standardized effect sizes of the 5 MaxVCP scores, each conditional on 97 baseline-LD v2.2 annotations (marginal *τ*^∗^). **(iii)** Standardized effect sizes, jointly analyzing 97 baseline-LD v2.2 annotations and all 5 MaxVCP scores (joint *τ*^∗^). **b**. Standardized effect sizes of the MaxVCP-xQTL and an analogous score based on the HyPrColoc method in a joint model involving other 97 baseline-LD v2.2 annotations. All results are meta-analyzed across 57 complex traits as well as subsets of 18 brain-related and 22 blood-related traits following from S-LDSC recommendations^56^. The asterisks indicate statistical significance (Bonferroni adjusted *p* < 0.05). Error bars indicate 95% confidence intervals. Numerical results are reported in **Supplementary Data**.

We conducted five secondary analyses to evaluate robustness of MaxVCP-xQTL. First, we compared MaxVCP-xQTL with a *CoS*-*xQTL* annotation that labels each variant as 1 if it appears in a 95% CoS (**Methods**). CoS-xQTL showed lower heritability enrichment than MaxVCP-xQTL and was not conditionally informative when jointly analyzed with MaxVCP-xQTL (**Figure S12c**). Second, we tested if binarizing MaxVCP-xQTL at various thresholds affected heritability signals, finding that these discrete annotations performed comparably or worse than the continuous MaxVCP-xQTL (**Figure S12d**). Third, we contrasted MaxVCP-xQTL with a *MaxPIP* annotation built from maximum SuSiE fine-mapping PIP across all 17 xQTL traits. MaxVCP-xQTL showed higher standardized effect sizes both in marginal and in joint analysis, although SuSiE still contributed substantially to the joint analysis for brain-related traits. This likely reflects additional contribution from trait-specific xQTL not colocalizing with other modalities or contexts (**Figure S12e**). We next investigated the enrichment of MaxVCP-xQTL in variants confidently fine-mapped (PIP>0.95) from 94 UK Biobank traits^34^ and 930 Million Veteran Program (MVP) GWAS traits^66^ using an EOO approach (**Methods**). We observed a high 17 ± 0.70-fold and 11.8 ± 0.62-fold enrichment respectively (**Figure S12f**), and prioritized 129 such variants with MaxVCP-xQTL ≥ 0.5 (**Table S7**). Finally, we stratified MaxVCP-xQTL into cell-type-specific (colocalization of a single cell type with bulk QTL) and cell-type-shared (colocalization among multiple cell types) annotations to assess whether cell-type specificity dominates disease heritability signal. Both annotations remained jointly informative in the meta-analyses for all traits and brain traits (**Figure S12g**), suggesting that while cell-type specificity in colocalization is important, shared regulatory architecture also substantially contributes to disease risk.

### GWAS–xQTL *ColocBoost* identifies regulatory variants for Alzheimer’s disease

We applied disease-prioritized *ColocBoost* to integrate 17 xQTL data with an Alzheimer’s Disease (AD) GWAS from a recent meta-analysis study^67^ (*N*_*cases*_ = 111,326, *N*_*controls*_ = 677,663 including 46,828 proxy cases from UK Biobank), identifying 188 colocalization events spanning 120 distinct loci where a 95% CoS linked AD to at least one gene-level xQTL trait (**Figure 6a**). 61.8% of these loci were not discovered by GWAS-only fine-mapping using SuSiE at 95% credible-set (CS) coverage, suggesting that integration with molecular QTLs can uncover disease associations that are not marginally significant in GWAS. To assess their credibility, we applied *ColocBoost* to a second AD GWAS which has a smaller effective sample size especially for non-proxy AD cases^68^ (*N*_*cases*_ =86,531, *N*_*controls*_ =676,386 including 46,613 proxy cases from UK Biobank); this study differs from our primary AD meta-analysis GWAS in cohort composition, proxy case definitions, linkage disequilibrium, and phenotype ascertainment (details in **Supplementary Note S.8**). In this study, *ColocBoost* identified 55 CoS with xQTLs, 20 of which overlapped with the fine-mapped CS from the same GWAS. Among the remaining 35, 17 overlapped with fine-mapped CS in the larger study^67^. While this 48.5% overlap offer support for new GWAS loci *ColocBoost* can identify, it represents an underestimate of replicability given that even the larger study may still be underpowered for some loci, and that this comparison is not a strict downsample-based replication given the aforementioned differences between the two GWAS studies.

**Figure 6.**
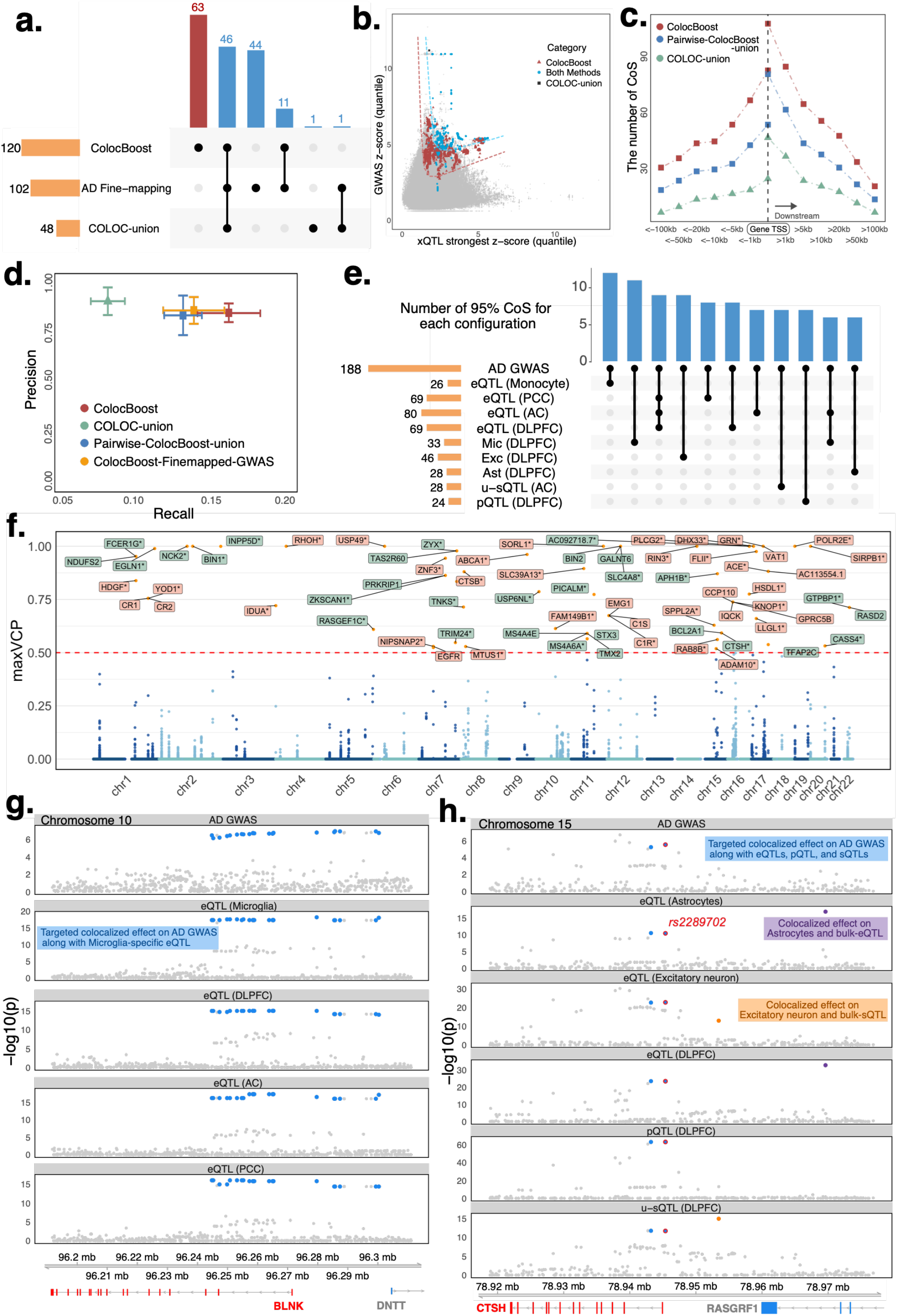
AD–xQTL *ColocBoost* identifies colocalized variants between xQTLs and AD GWAS. **a.** UpSet plot of distinct (i) 95% CoS identified by GWAS–xQTL *ColocBoost*, (ii) union of 95% CoS identified by applying COLOC on each xQTL with AD GWAS (COLOC-union), and (iii) 95% SuSiE credible sets (CS) from AD GWAS fine-mapping. **b**. Scatter plot across variants comparing the marginal association z-scores in AD GWAS against the quantile-matched z-scores corresponding to the strongest marginal association signals (in terms of the magnitude of z-scores) across all xQTL traits for each CoS. We color-code variants within CoS corresponding *ColocBoost*, COLOC-union and both methods. **c**. Distribution of distances from the gene TSS for CoS from AD–xQTL *ColocBoost*, COLOC-union, and the union of CoS identified by *ColocBoost* applied pairwise to one AD GWAS and each of xQTL traits (pairwise-ColocBoost-union). **d**. Precision-recall analysis comparing CoS-gene links from AD-xQTL *ColocBoost*, COLOC-union, pairwise-ColocBoost-union, and a version limiting AD-xQTL *ColocBoost* to AD fine-mapped variants (ColocBoost-Finemapped-GWAS) against enhancer-gene links predicted by ENCODE-rE2G across 354 biosamples. Error bars along both axis indicate 95% confidence intervals. **e**. An UpSet plot focusing showing the distinct colocalization patterns across xQTLs exhibited by 95% CoS from AD-xQTL *ColocBoost*. **f**. Manhattan plot of variant level MaxVCP from *ColocBoost*, with labeled genes containing variants with MaxVCP>0.5 and highlighting microglia contributions (green). **g**. An example case of AD-xQTL colocalization demonstrating a single CoS with cell-type specific colocalization in microglia for *BLNK* gene. **h**. A second example case of AD-xQTL colocalization demonstrating three distinct CoS showing different colocalization patterns across multiple brain cell types for the *CTSH* gene. Numerical results are reported in **Supplementary Data**.

To compare with existing methods, we compiled additional sets of CoS via two pairwise approaches: by taking the union of CoS identified via applying COLOC on each xQTL with AD GWAS (hereafter referred as *COLOC*-*union*) and by performing the same pairwise analysis using *ColocBoost* (hereafter referred as *pairwise*-*ColocBoost*-*union*). *ColocBoost* recovered 96% of the events identified by COLOC-union, whereas COLOC-union reproduced only 38% of *ColocBoost* CoS (**Figure 6b**). Based on S-LDSC analysis, variants implicated by *ColocBoost* explained nearly twice as much AD heritability as COLOC-union, and the combination of *ColocBoost* CoS and AD fine-mapping CS explained 1.6 times more AD heritability than fine-mapping alone (**Table S8**). Although the overall outcome indicates that integrating xQTL data via AD-xQTL *ColocBoost* meaningfully strengthens causal AD variant and target gene discovery, we caution that LDSC heritability estimates for a single disease, like AD, can be noisy^69^. We further examined gene-proximal versus gene-distal (outside of gene TSS±10kb) colocalizations, finding that *ColocBoost* and pairwise-*ColocBoost*-union each identified more gene-distal regulatory loci (2.9-fold and 1.9-fold, respectively) than COLOC-union (**Figure 6c**). We further validated these CoS via ENCODE-rE2G model^34^, a predictive E2G framework integrating 1D chromatin activity and 3D chromatin contact features based on their concordance with CRISPR data across 354 biosamples from different cell types, tissues, and contexts^36,37^. A precision-recall analysis of CoS-gene maps with enhancer–gene maps showed 1.94 times higher recall for *ColocBoost* compared to COLOC-union, with minimal drop in precision (**Figure 6d**). A similar precision was observed when restricting the analysis to 63 unique *Colocboost* CoS not identified by AD fine-mapping. This recall enhancement is largely driven by (i) gene-distal CoS (**Figure S13a**) and (ii) cell-type or brain region-specific CoS (**Figure S13b**). While pairwise-*ColocBoost*-union also exhibited improved recall over COLOC-union, its performance remained slightly poorer than *ColocBoost* incorporating all xQTL data. We excluded HyPrColoc from this application because it identified only one colocalization event—between *BIN1* microglia eQTL and AD—when applied to all 18 traits (17 xQTLs and 1 AD GWAS). As previously illustrated (**Figure 2b-i**), HyPrColoc may struggle with heterogeneous signals across traits, which likely explains its limited yield in this application integrating diverse xQTL modalities and GWAS.

Out of 188 colocalization events detected by AD-xQTL *ColocBoos*t, 34% (63 events) involved only a single brain cell type, with 19 microglia specific events despite being underpowered as a relatively less common cell type in DLPFC (cell proportion < 5%) (**Figure 6e**). This finding aligns with the recognized importance of microglia in AD pathogenesis^70–72^. Overall, 68 genes were linked by *ColocBoost* to a variant with high colocalization probability (MaxVCP>0.5; **Figure 6f**); 44% of them involved immune cells microglia or bulk monocyte. In comparison, 66 genes were associated with variants having MaxVCP>0.5 for pairwise-*ColocBoost*-union, whereas only 23 genes met this threshold for COLOC-union (**Figure S13c,d**, and **Figure S14a,b** for a relaxed score of MaxVCP>0.1). 38.1% of microglia or monocyte colocalizations also showed shared effects with other brain cell types (**Figure 6e**). We further evaluated microglia-colocalized genes against three curated microglia gene sets from Yang *et al*.^73^ (**Figure S14c**), and observed significant enrichment with two of them: microglia-specific IFNβ-responsive genes (EOO=2.49, *p* = 0.001) and differentially expressed microglia genes under IFNβ treatment (EOO=2.29, *p* = 2 × 10^−36^). We also identified strong enrichment (EOO=4.96, *p* < 0.01) with AD disease-progression genes in microglia^25^. A list of AD colocalization and their trait configurations is provided in supplementary material (**Table S1, Figure S15a**). We also identified several variants spanning multiple genes: for example, *rs679515* (*chr1:207577223:T:C*) colocalized with *CR2* in DLPFC bulk eQTLs, *YOD1* in oligodendrocytes, and *CR1* across oligodendrocytes and three additional bulk cortical eQTLs^74,75^ (MaxVCP=0.755; **Figure S15b**).

We highlight two examples of AD colocalization at the cis-regulatory loci of *BLNK* and *CTSH*, which were uniquely identified by *ColocBoost* as neither gene reached genome-wide significance of 5 × 10^−8^ in AD GWAS (p-values of 1.1 × 10^−7^ and 2.18 × 10^−7^ respectively, indication suggestive associations), which partly explains why they were missed by COLOC-union and HyPrColoc. In the first example, a single 95% CoS links *BLNK* to AD specifically in microglia (**Figure 6g**). Although *BLNK* is considered an AD risk gene (significantly upregulated in Aβ-treated cells^76^), a microglia-specific eQTL underlying suggestive AD association has not previously been reported^77,78^. This CoS comprises 22 variants in strong LD (purity 0.945), resulting in highest VCP of only 0.095. Therefore, a variant-level colocalization score (e.g., even at relaxed MaxVCP>0.1) would overlook this locus, whereas 95% CoS, a set-level metric including LD proxies, uncovered the microglial eQTL colocalization. This observation highlights the advantage of CoS compared to per-variant colocalization probabilities. *CTSH* offers a more complicated example where three CoS were identified. One of them colocalizes with AD GWAS in astrocytes and excitatory neurons (**Figure 6h, Figure S16a**) and contains two tightly correlated variants (*r*=0.975). One variant, *rs2289702* (VCP=0.415), is located 29kb downstream of *CTSH* gene TSS, and was recently shown to reduce *CTSH* expression in luciferase assays by significantly disrupting a transcription-factor binding site (ZFP69B)^79–81^. The same CoS also encompasses an unproductive splicing QTL (u-sQTL, associated with splicing junction between Exon 3 and 4) and a pQTL in DLPFC, with effect directions suggesting a coherent cascade of decreased expression and protein abundance (**Figure S16b**). The other two CoS at *CTSH* did not colocalize with AD GWAS: one showed cell-type specific colocalization in excitatory neurons with links to unproductive splicing, while the other showed cell-type colocalization in astrocytes (**Figure 6h, Figure S16**). The low-to-moderate correlation across all three CoS (average *r* =0.53) suggest potentially independent regulatory paths, illustrating how multi-modal colocalization can unravel intricate regulatory architectures despite challenges in completely disentangling multiple correlated causal alleles as is the case of the first CoS.

## Discussion

*ColocBoost* fills a critical methodological gap in large-scale xQTL colocalization analysis by providing a calibrated statistical approach that handles many traits and multiple causal variants while representing each putative causal variant with its LD proxies. As a multi-task variable selection framework with specialized gradient boosting updates, *ColocBoost* effectively identifies regulatory variants with shared causal effects across traits, improving sensitivity to map weaker effects in both simulations and real-world applications. As demonstrated by our large-scale brain xQTL analysis and AD GWAS integration, *ColocBoost* generates variant-level multi-omics colocalization maps that connect genetic loci to shared molecular effects relevant to complex diseases. Our multi-omics colocalization map is strongly supported by extensive orthogonal functional genomic data, underscoring its potential for guiding CRISPR screening and other functional follow-up studies.

Although *ColocBoost* draws inspiration from the single-effect regression (SER) concept in SuSiE^38^, including its extensions for multi-trait fine-mapping^38,82–84^, it differs from these methods in both its statistical target and computational construction. Multi-trait fine-mapping like mvSuSiE^84^ identifies variants that contribute globally to *at least one* trait, whereas *ColocBoost*, a multi-trait colocalization method, elucidates local colocalization events and the specific configuration of traits sharing each effect. Compared to mvSuSiE which fits a multivariate regression with a genome-wide estimated prior on effect-sharing patterns, *ColocBoost*, as a multi-task learning model, learns local sharing patterns through weak proximity-smoothed updates, SEC decisions across traits, and adaptive scheduling, providing a more flexible framework to identify heterogeneous effects across traits not captured by global Bayesian priors. In contrast to Bayesian colocalization which can be sensitive to prior specification^8,14,15,17,85^, *ColocBoost* does not need a user-specified colocalization prior. Its *ℓ*_(∞,1)_-norm objective favors variants with cross-trait gradient contributions^86,87^, and the SEC determines which traits to couple at each locus using profile log-likelihood comparisons. Each learner contributes a small update, so an early coupling decision does not determine the final call. The weak-learner construction and adaptive scheduling introduce several hyperparameters, and simulations across the evaluated settings support the default choices (**Methods, Supplementary Note S.5.2, Figure S5**). *ColocBoost* also accommodates partially overlapping samples and variant sets and allows trait-specific LD references. **Table S9** summarizes the assumptions and limitations of related methods. Of these methods we note that CAFEH^88^, another SuSiE extension intended to perform fine-mapping and colocalization, suffers from poor calibration (high FDR)^82^, leading us to exclude it from direct comparisons in this work.

Our work has several downstream applications. First, the gradient boosting framework can potentially be repurposed for cross-ancestry GWAS fine-mapping by assembling SEC whose variant-trait configurations support cross-ancestry signals. Second, *ColocBoost* can likewise serve as a pleiotropy identification method by extending the application to multiple GWAS traits. Third, MaxVCP and CoS generated from *ColocBoost* can be used to improve functionally informed polygenic risk prediction within and across ancestries, as well as to inform functional characterization experiments, such as CRISPR^28,89^ and MPRA^90,91^. Finally, while we here showcased only 17 xQTL contexts, we have already applied *ColocBoost* in over 80 distinct molecular QTL datasets beyond eQTL, sQTL, and pQTL, and leveraged the resulting MaxVCP to general novel resources for disease-informed variant-to-function links, as reported in the FunGen-xQTL flagship manuscript^22^.

Our work has several limitations. First, certain omics data—such as thousands of highly correlated chromatin accessibility peaks or methylation sites—can pose scalability challenges, although simple dimensionality-reduction steps^92^ or wavelet-based transformations can help^93^ (but we are yet to test these scenarios using *ColocBoost*). Second, *ColocBoost* may less well calibrated in one setting: strongly correlated residuals across traits (*ρ* ≥ 0.7) can inflate FDR under some causal configurations, although residual correlations in our xQTL analyses were only moderately correlated 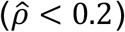 (**Figure S17**). Third, the multi-task regression currently comprises linear models, though in principle one can accommodate generalized likelihoods for count-based, binary, or mixed outcomes by developing corresponding SEC updates. Fourth, like most fine-mapping methods, *ColocBoost* assumes consistent LD reference data with summary statistics data; violation of this assumption may yield false discoveries, even though in such scenarios *ColocBoost* controls FDR reasonably well compared to SuSiE-RSS in single-trait fine-mapping analysis (**Figure S18**). An LD-free implementation, mirroring HyPrColoc with single-causal assumption, is available for applications without a reliable LD panel (**Figure S3g**). Fifth, *ColocBoost* does not currently integrate functional annotations into SEC updates^97^. Given that functional annotations have been shown to significantly enhance fine-mapping, incorporating them into *ColocBoost* could potentially provide similar benefits. Finally, the application of *ColocBoost* in this work has been focused on eQTLs, sQTLs and pQTLs; as part of the FunGen-xQTL effort, we are currently expanding the application to additional non-coding RNA QTLs, APA QTLs, m6A QTLs, as well as exploring multi-ancestry QTL data. We envisage *ColocBoost* as a core framework for our next wave of integrative xQTL and GWAS studies.

## Methods

### Multi-trait colocalization problem and colocalization events

The multi-trait colocalization problem concerns identifying shared causal variants across *L* traits within a genomic region of interest. The result of a *ColocBoost* analysis at a locus is a set of *Colocalization Events*, where each event {*s*} is summarized by a triplet {*g*(*s*), *T*(*s*), *CoS*_*α*_(*s*)} where:

i. *g*(*s*) denotes a *Genomic Region* or locus underlying the *ColocBoost* analysis,
ii. *T*(*s*) is the *Causal Configuration* specifying which subset of the analyzed traits share the same causal variant,
iii. *CoS*_*α*_(*s*) is a *Colocalization Confidence Set* representing the smallest set of variants containing the causal variant with probability at least α (default 95%).

The *CoS*_*α*_ plays the same role in multi-trait analysis that the credible set plays in single-trait fine-mapping: under realistic linkage disequilibrium (LD), the data cannot uniquely identify a single causal variant from its tightly linked proxies, but it can identify the set of variants among which the causal variant must lie. We therefore adopt *CoS*_*α*_ as the primary unit of colocalization inference, in contrast to existing approaches that report a regional colocalization probability paired with a single nominated variant (for example, COLOC’s PP.H4 with the top SNP, or HyPrColoc’s regional posterior probability with a candidate variant).

The challenge in multi-trait colocalization is that the configuration space grows combinatorially with the number of traits. At each locus with *M* putative causal variants that may influence two or more of *L* traits, enumerating all possible colocalization events requires evaluating *M* × (2^*L*^ − 1) configurations. This combinatorial explosion becomes computationally prohibitive as the number of traits increases. Previous Bayesian colocalization methods require explicitly defining this massive hypothesis space a priori, which becomes intractable for even moderate numbers of traits^14–16^. *ColocBoost* avoids enumeration by discovering supported configurations through a sequential boosting algorithm rather than evaluating each configuration in advance.

### Rationale of multi-trait gradient boosting algorithm for colocalization detection

*ColocBoost* characterizes colocalization events through a *Proximity Smoothing Gradient Boosting* (PSGB) algorithm, extending a recent iterative gradient-boosting framework^35^ to jointly fit multi-trait regression models while accounting for high correlation among genetic variants. At *L* = 1, this reduces to single-trait fine-mapping (referred to as *FineBoost*; **Supplementary Note S.1**); at *L* > 1, it generalizes to multi-trait colocalization through coupled updates across traits. The multi-trait extension combines two ideas: (i) a joint objective that inherently favors variants associated across multiple traits and decides which traits share the variant effect, and (ii) a smoothing strategy that handles LD-induced uncertainty in selecting causal variants from their LD proxies within each trait.

For *L* traits 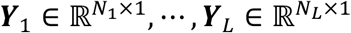 (standardized) and corresponding standardized (centered and scaled) genotype matrices 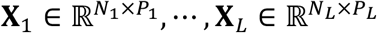 (with *N*_*l*_ denoting the number of individuals and *P*_*l*_ denoting the number of genetic variants for trait *l*), we consider the joint loss function 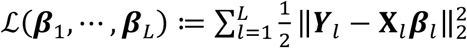. This can be interpreted as the log-likelihood under a multiple linear regression model 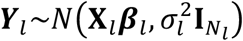 with fixed effect size ***β***_*l*_. The trait-indexed dimensions accommodate settings where samples and variant sets differ across traits; for better clarity of presentation, we assume *P*_*l*_ = *P* and *N*_*l*_ = *N*. The *ℓ*_(∞,1)_-norm of the gradient of ℒ is given by

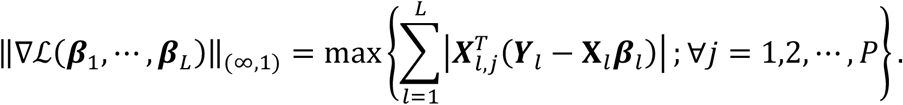

This aggregates gradient contributions across traits at each variant via an *ℓ*_1_-type sum and then maximizes across variants via an *ℓ* _∞_-type maximum. This norm structure encourages selection of variants with large cross-trait gradient contributions, structurally favoring colocalized signals without requiring an explicit prior over which traits should share variants. *ColocBoost* minimizes this objective sequentially through gradient boosting, with each iteration’s update determined by the PSGB and *Single-Effect Coupler* (SEC) mechanisms described in the next two subsections.

### Generalization of trait-specific proximity smoothing gradient boosting algorithm

For a single trait (*L* = 1), standard gradient boosting selects the single variant by maximizing the trait-specific gradient contribution 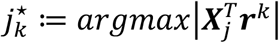 at iteration *k*, where 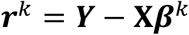 is the residualized phenotype. The algorithm then updates only the effect size of this single variant through a weak learner with a dynamic learning rate^35^. This hard-selection rule makes the update sensitive to small differences among high LD variants. For example, when two variants *j* and *j*^′^ are in high LD, 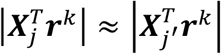 and select one candidate variant to update over other high-LD neighbors can lead to (i) selecting wrong proxy tagging variants and missing out on the true causal variant, and (ii) high sensitivity of the update variants under small fluctuations of the data.

To address this issue, *ColocBoost* employs a PSGB replacing the hard selection in each standard gradient update with a soft assignment, distributing the update across variants in proportion to their evidence and LD with the leading variant. The soft assignment arises as the closed-form solution of a Nesterov-smoothed optimization problem. The hard maximum across variants in the *ℓ*_∞_-norm objective 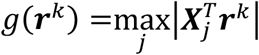 is replaced by an entropy-regularized approximation,

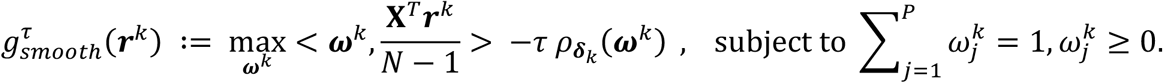

where ⟨·,·⟩ is the Euclidean inner product, ***δ***^*k*^ ∈ ℝ^*P*^ is a positive LD-informed reference measure, and 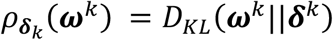 is the Kullback-Leibler (KL) divergence between ***ω***^*k*^ and ***δ***^*k*^ defined below. 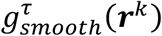 is strictly concave, thus admits a *unique* maximizer, 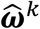, with the *closed-form* softmax representation:

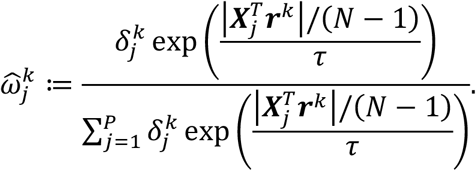

Here, 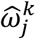 represents a single-effect conditional inclusion probability which is a Gibbs distribution over variant identity, with ***δ***^*k*^ as its reference measure and the exponentiated gradient as its data-dependent term. It is structurally analogous to SuSiE’s single effect posterior ***α***_*l*_, but it is induced by the Nesterov-smoothed variational problem rather than a Bayesian model. At boosting round 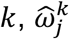 is the probability mass assigned to variant *j* for the current signle-effect update. It replaces the hard selection of 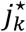 by distributing the update across candidate variants (**Proposition S1, Supplementary Note S.3.2**). The temperature *τ* > 0 controls its concentration and the approximation error (default at *τ* = 0.01). As 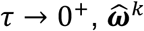 concentrates on 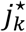 with maximal gradient evidence^35^, and as 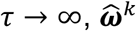 coverages to ***δ***^*k*^. Its calibration against simulated causal variants and comparison with SuSiE^38^ are shown in **Figure S4d**. We also evaluated the sensitivity of 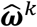 is to both *τ* and the regularization term (**Figures S4c-e**; **Supplementary Note S.5.2**).

The resulting smoothed objective is convex and differentiable,

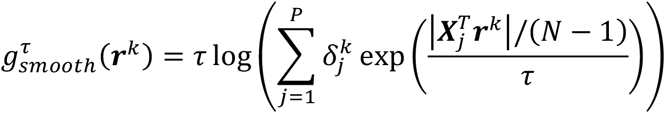

with gradient with respect to 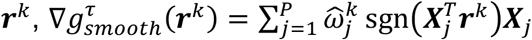, which distributes the update across all variables weighted by 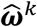, in contrast to the standard sub-gradient supported on a single variable.

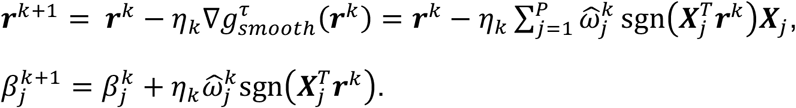

The dynamic learning rate *η*_*k*_ is adaptively tuned at each boosting round at a sufficiently small value to ensure each update behaves as a *weak learner*, making controlled, incremental updates to prevent overfitting while capturing weak effects (**Supplementary Note S.1**).

The regularization term 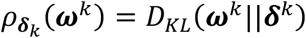 has two components requiring justification, the KL functional form and the LD-informed reference measure ***δ***^*k*^. (1) The KL form yields the analytical solution for 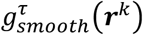 (**Supplementary Note S.3.3**). For fixed ***δ***^*k*^, the approximation satisfies 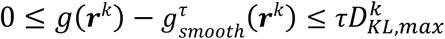, where 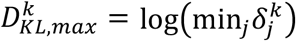. This reduces to *τ*log(*P*) for a uniform reference measure. (2) Sensitivity to the reference measure ***δ***^*k*^ (**Figure S4e**). We specify 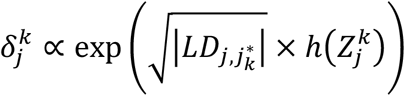 with 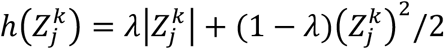. Here, *λ* is a mixing parameter, 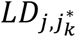 denotes LD between variants *j* and 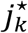, and 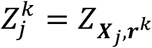 is the z-score testing association between ***X***_*j*_ and ***r***^*k*^ at *k* th iteration (**Figure S1a**). The function *h*(·) combines the |*Z*| component, which arises from the *ℓ*_∞_-norm *Correlation Minimization* framework underlying boosting^35^, with the *Z*^2^/2 component, which resembles the evidence used in normal-theory Bayes factors. The LD weighting concentrates mass on variants in high LD with the leading variant. We use *λ* = 0.5 as the midpoint of this mixture and evaluated sensitivity across the full range *λ* ∈ [0,1]. Compared with the default, ***ω***^*k*^ remained stable, with correlations exceeding 0.96 and lower KL divergence (**Figure S4, Figure S5**, and **Supplementary Note S.5.2**).

### Multi-trait coupling updates via the Single-Effect Coupler

*ColocBoost* extends single-trait PSGB to multiple traits through *Single-Effect Couplers* (SEC). At each boosting iteration, a SEC evaluates whether traits share a common causal effect and, if so, performs coupled proximity-smoothed updates (**Figure S1**). The coupling decision is made locally by comparing joint versus separate trait updates using profile log-likelihood. Over successive iterations, these decisions progressively refine which traits are coupled, and the resulting trait groupings define the causal configuration *T*(*s*).

We first introduce a simplified version of *ColocBoost* with *L* = 2 traits via SEC as a special case to help readers more easily understand the strategy in a pairwise colocalization setting. At boosting round *k*, we define the best single-effect variant 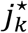 for *joint updates* across two traits as

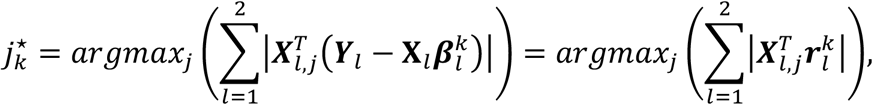

and the *trait-specific* best single-effect variant 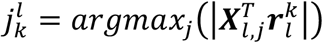 for *l* = 1,2. We define *equivalence* of 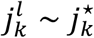 between 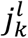 and 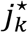 if 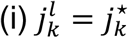 or they are in strong LD with each other (|*r*| > 0.8), and (ii) have comparable effects, i.e., Δ*loglik*(·; ***Y***_*l*_) for trait *l* at the two variants is smaller than *ε* (default *ε* = 0.1), where Δ*loglik*(·5 ***Y***_*l*_) measures the drop in trait *l*’s profile log-likelihood when effect at a given variant is set to zero. Therefore, determining which traits to update jointly reduces to evaluating whether each trait’ 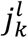 is equivalent to 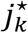. We propose a *dynamic coupling strategy* as follows (**Figure S1b**):

#### Logic 1 (SEC) – two traits shared the same single-effect

If 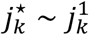 and 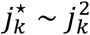, we *couple* the updates across both traits at the same single-effect variant 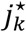 with trait-specific proximity smoothing (**Figure S1c**).

#### Logic 2 – two traits had stronger distinct trait-specific single-effects

If 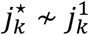 and 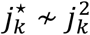, we update each of trait 1 and trait 2 separately at 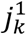 and 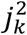, respectively.

#### Logic 3 (delayed SEC) – one trait has a stronger trait-specific single effect

If 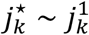 but 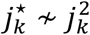 (or vice versa) because trait 2 has a stronger trait-specific signal at 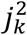. This can occur when trait 2 has both a shared and a trait-specific effect, or when distinct causal variants in LD mimic sharing. *ColocBoost* addresses this through delayed SEC (D-SEC); it first updates trait 2 at 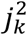 and then re-evaluates 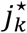. A shared signal can remain in the residuals and be coupled later, whereas an association induced by LD should weaken after the trait-specific update (**Figure S1d**; **Supplementary Note S.2.3**).

The pairwise coupling strategy generalizes naturally to *L* > 2 traits. At each boosting round, traits are partitioned into equivalence groups based on their trait-specific best single-effect variants, and the same three-logic framework determines whether to couple (logic 1), separate (logic 2), or defer (logic 3) updates across groups. The complete multi-trait specification is provided in **Supplementary Note S.2**.

### Aggregate PSGB iterations to colocalization events

*ColocBoost* provides variant-level probability measures that propagate uncertainty from individual boosting iterations through multi-trait aggregation to a final per-variant quantity. The probability hierarchy traces four levels: (1) per-iteration variant probability 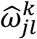, arising from the Nesterov-smoothed objective (**Proposition S1**; **Supplementary Note S.3.2**); (2) trait-specific single-effect probability 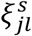, averaging 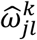 across iterations within one traits and a single-effect component; (3) single-effect colocalization probability 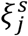, combining the trait-specific 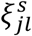 for the colocalizing traits in that component; and (4) variant colocalization probability (VCP), combining evidence across single-effect components under the same conditional independence assumption used by SuSiE for PIP^38^. The full derivation of this probability hierarchy is summarized below.

Upon convergence, *ColocBoost* identifies a total of *S* colocalization events within a genomic region *g*. As previously defined, each colocalization event *s* (for *s* = 1, ⋯, *S*) corresponds to a triplet {*CoS*_*α*_(*s*), *T*(*s*), *g*}, where *T*(*s*) specifies the causal configuration of traits and *CoS*_*α*_(*s*) represents the *α*-level colocalization confidence set.

#### Determination of T(s)

The trait configuration *T*(*s*) is represented a binary vector 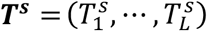, where 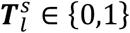 indicates whether the variant has a putative effect on trait *l*. For any given variant, the total number of possible colocalization configurations across *L* traits is 2^*L*^ − *L* −1, excluding single-trait and non-causal cases. The configuration ***T***^***s***^ is determined by examining which traits share evidence for genetic effects across boosting iterations, as captured by SEC. Therefore, let *K*_*s*_ denote the subset of boosting iterations corresponding to configuration ***T***^*s*^.

#### Determination of CoS_α_(s)

For each trait configuration ***T***^*s*^ and corresponding boosting iterations *K*_*s*_, the *α*-level CoS is constructed through the following sequential steps.

1. Clustering of single-effect components. For each trait *l* with 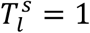, we define 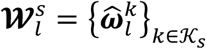 as the collection of single-effect probability vectors 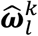 for *k* ∈ *K*_*s*_, where 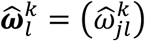 denotes the single-effect probability for all variants at iteration *k* as previously introduced in the section describing proximal gradient boosting algorithm. By virtue of being weak learners, different SEC may update the same or nearby (LD-proximal) variants over multiple rounds for the same causal effect. This often yields similar weight vectors 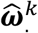 and 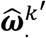, which, by design, further enhances accommodation of inherent signal uncertainly in high-LD regions. To assemble these SECs, we aggregate 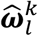 for *k* ∈ *K*_*s*_ using a modularity-based hierarchical clustering approach (**Supplementary Note S.4**).
2. Defining of single-effect colocalization probability. Within each cluster of component *s*, we aggregate 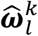 for trait *l* ∈ *T*(*s*) using the joint profile log-likelihood improvements, 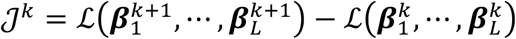.

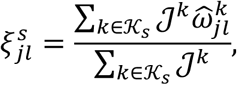

where the weighting iterations by their improvement in fit follows the gradient boosting ensemble construction^100^. The normalized weights are non-negative and sum to one, so 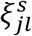 remains a probability distribution on {1, ⋯, *P*} inheriting the properties established for 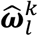 in **Proposition S1** (**Supplementary Note S.3.4**). Finally, 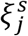 are aggregated across colocalizing traits (*l* ∈ *T*(*s*)) using logarithmic pooling^101^, 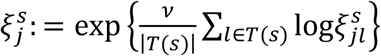. After normalization, 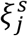 is a *single-effect colocalization probability* for variant *j* supporting *T*(*s*) for every *ν* > 0. The parameter *ν* controls concentration without changing the variant ranking. We use *ν* = 1.5 by default. Across simulations with 2, 5, and 10 traits, power changed modestly over *ν* ∈ {1,1.5,2} and FDR remained below 0.05 (**Figure S4; Supplementary Note S.1.4**).
3. Construction of *CoS*_*α*_(*s*). As previously described (in the **Overview of Methods**), 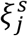 can be used to construct the *α*-level CoS under *T*(*s*),

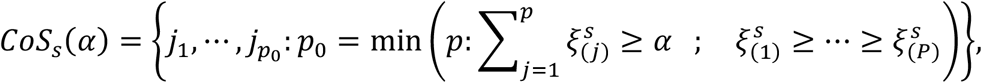

where 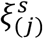 denotes the *j*-th largest probability 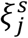. Similar to SuSiE^38^, we discard the CoS with low *purity* (minimum absolute correlation between all pairs of variants within CoS, default to purity < 0.5) and with small Δℒ_l_ and 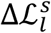, the relative contribution to the trait-specific profile log-likelihood and CoS-trait-specific log-likelihood respectively (Δℒ_*l*_ <0.025 and 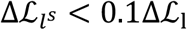 by default). We also introduce strategies to further handle highly correlated CoS, to diagnose quality of CoS due to incomplete LD and/or mismatch with summary statistics, and to filter potentially spurious signals (**Supplementary Note S.4**).

#### Determination of variant colocalization probability (VCP)

We define VCP by combining evidence across single-effect colocalization probabilities, ***ξ***^*s*^, under the same conditional independence assumption used by SuSiE for posterior inclusion probability (PIP). Specifically, for variant *j*,

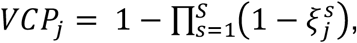

where in both frameworks, the independence assumption arises because each component is fitted *conditionally* on residuals from all other components. VCP serves as an analog of PIP in single-trait statistical fine-mapping^3,38,39^. This parallels the iterative Bayesian stepwise selection (IBSS) procedure in SuSiE^38^, where each single-effect regression is fitted to residuals from all other effects, ensuring conditional independence across components.

### Expediated boosting learners for disease-prioritized *ColocBoost*

We introduce an *expedited SEC* (E-SEC) to prioritize identifying colocalization with a focal trait, e.g., a GWAS, implementing the disease-prioritized mode of *ColocBoost*. We start with the best single-effect variant for GWAS of interest 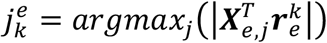, and construct its equivalence group 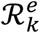 with respect to 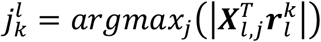 for each trait *l* ≠ *e* by checking if 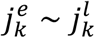. If 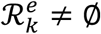, we re-compute 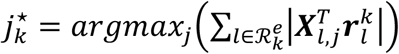 across traits in 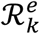 as the candidate variant to update in current boosting round, where by design, 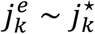. We then employ the dynamic coupling strategy previously described to guide subsequent boosting rounds. This procedure is iterated until 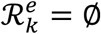. In doing so, traits in 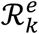 are prioritized for coupling at variant 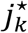 during earlier rounds, thereby improving detection of colocalizations with the focal trait.

### Summary statistics extension and related practical features

We provide implementation details in the **Supplementary Note S.2.5** describing how *ColocBoost* addresses common challenges in real-world genetic data integration. First, *ColocBoost* supports both individual-level and summary-statistics-based analyses, with optional use of multiple external LD reference panels when individual-level data are unavailable. Second, it accommodates mixed data types across traits, as in our GWAS– xQTL integration. Third, it handles non-overlapping variant sets without requiring imputation or variant exclusion while retaining variants with partially missing LD data. Finally, it provides an LD-free mode under a single causal variant assumption, enabling analysis when LD is unavailable or poorly matched.

### Simulation settings for multi-trait colocalization methods performance benchmark

To evaluate multi-trait colocalization methods, we used genotype data from the ROSMAP cohort (*N* =1,162) on 1,287 gene regions (±1.5Mb from gene TSS) randomly drawn from topologically associating domains (TAD) across the genome^42^. Each region contains variants filtered by missing rate less than 0.1 and minor allele frequency (MAF) greater than 0.05, resulting in between 2,069 to 40,000 variants per region (average 9,216). Let **X** be the standardized genotype matrix in each region and ***β***_*l*_ be the effect sizes for variants on trait *l*, where *P*_*c*_ approximate independent variants were sampled to have causal (non-zero) effects and all the remaining were set to non-causal (zero effect). We simulate the normalized effect sizes such that the percentage of phenotypic variance explained (PVE) by each variant 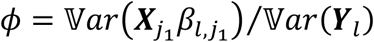 for 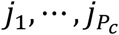 is kept fixed. Then, we generated simulated traits based on ***Y***_*l*_ = **X*β***_*l*_ + ***ϵ*** with 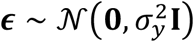, where 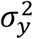 is chosen such that a modest *ϕ* = 0.05 was used for the xQTL-only colocalization setting, and a lower *ϕ* = 0.02 use for disease-prioritized setting. While this *ϕ* may appear relatively large for some GWAS traits, real GWAS typically benefit from much larger sample sizes to detect weaker effects with comparable power. Our choice of *ϕ* for simulation purpose is merely a practical trade-off given limited availability to large-scale genome sequence data. For under the null, we set ***β***_*l*_ = 0 and vary 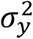 from 0.1 to 1. To simulate dependency between traits due to non-genetic factors such as sample overlap, we model residual as 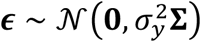, where **Σ** was estimated from FunGen-xQTL atlas used in our real data analysis (**Supplementary Note S.6**).

Our primary numerical study considered three broad scenarios based on different number of traits simulated: (i) For simulations with only two traits, we assumed both traits shared the same *P*_*c*_ causal variants, where *P*_*c*_ varied from 1 to 3. (ii) For simulations with 5,10, and 20 traits, we adopted a more complex simulation design to mimic the number of causal variants and variant-trait configurations in real world data analyses. Specifically, we varied the number of causal variants, ranging from 1 to 5, and randomly assigned each variant to subset of traits to determine the colocalization patterns at each gene locus based on an empirical probability distribution derived from the observed genome-wide overlap of fine-mapping credible sets in FunGen-xQTL fine-mapped atlas^22^ (**Table S10**). For example, when simulating 3 causal variants per gene across 10 traits, we randomly selected 2-10 colocalized traits for each causal variant with probabilities 38.0%, 28.5%, 16.3%, 7.9%, 5.1%, 2.4%, 1.2%, 0.4%, 0.2%. (iii) For simulations with 50 traits, we also varied the number of causal variants, ranging from 1 to 5, and randomly selected 10-25 colocalized traits for each causal variant. Notably, as the number of traits increases, the colocalization patterns tend to become sparser. In some cases, distinct subsets of traits may share different causal variants (i.e., traits {1,2,3} shared one causal variant, while traits {4,5,6} shared another one). In addition, we conducted four other simulation experiments each featuring simpler colocalization patterns drawn from published work describing competing methods, whose design are briefly described in **Results** and detailed in **Supplementary Note S.6**.

### Performance metrics for benchmarking multi-trait colocalization methods

We used two principal metrics to compare *ColocBoost* with other methods: (i) statistical power and (ii) false discovery rate (FDR), evaluated at the level of a colocalization configuration, for both the 95% CoS and the set of traits involved in this configuration. We define *power* as the proportion of correctly identified CoS-trait pairs over the total number of true variant-trait pairs in simulated causal configurations. Specifically, let *T*_*j,true*_ be the set of true colocalized traits for the true causal variant *j* for *j ∈* ℳ_*causal*_, where ℳ_*causal*_ is the set of true causal variants. Let *CoS*_*s*_ denote the estimated 95% CoS for colocalization event *s* and 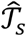 be the set of colocalizing traits associated with this event. For each true causal variant *j, power* is calculated as

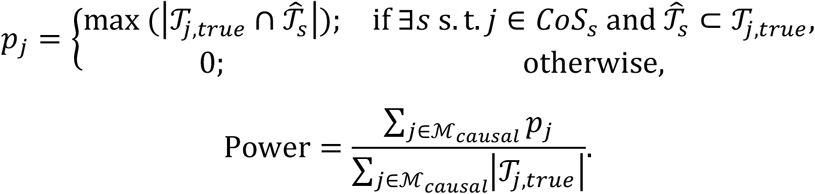

where the notation |*T*| denotes the number of elements in the set *T*. This definition provides a stringent measure of detection accuracy: it ensures that the detected colocalized traits must be a subset of the true set for that variant, and among such cases, power is proportional to the number of correctly identified traits.

We defined FDR as the proportion of incorrectly detected CoS over all detected CoS. Specifically, for each detected *CoS*_*s*_ and colocalized traits 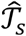, we considered two stringent conditions to classify false discoveries: (i) *CoS*_*s*_ does not include the true causal variant and (ii) *CoS*_*s*_ includes a causal variant *j*, but also includes incorrect traits 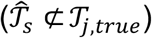. We assigned *F*_*s*_ = 1 if either of these two conditions was met and *F*_*s*_ = 0 otherwise. The FDR is calculated as

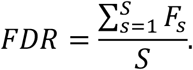

We further evaluate *variant-level* detection accuracy using *precision-recall curves (PRC)*, a standard practice adopted in a similar context (fine-mapping) when variant-level scores are determined by both signal strength and its LD proximity and is difficult to separate apart these two factors^38,83,102–105^. In brief, we applied a range of thresholds [0,1] to each variant’s colocalization score to compute precision and recall rates. For *ColocBoost*, we used VCP as the colocalization score; see **Supplementary Note S.6** for details of colocalization score of other methods.

### Comparison with OPERA using region-level benchmarks

We next compared *ColocBoost* with the recently proposed multi-omics colocalization method OPERA by simulating data at the region level for 2, 5, and 10 traits. In each locus, the first trait served as the *target* trait with per-variant PVE, *ϕ*, ranging from 0.02 to 0.05, while the remaining traits each had *ϕ* = 0.05. The number of causal variants and colocalizing traits per variant were randomly assigned based on distribution drawn from genome-wide overlaps of fine-mapped credible sets previously described. Because OPERA requires genome-wide GWAS summary statistics for estimating its hyperparameters, we computed summary statistics for 500 of the 1,287 independent, non-overlapping gene regions previously described, to serve as 500 replicates for power and FDR calculations (**Supplementary Note S.6**).

Unlike *ColocBoost* and other methods which identifies variant-level (VCP) and locus-level (CoS) colocalizations, OPERA only provides gene-level test for whether a colocalized signal exists within an entire gene, without pinpointing the specific variant(s). OPERA does not perform xQTL-only colocalization because it requires an explicit target trait (e.g. GWAS). To enable a fair comparison between OPERA and *ColocBoost*, we consider the GWAS-prioritized mode of *ColocBoost*, and defined power and FDR at gene-level evaluated on the target trait. Specifically, we defined power as the proportion of correctly identified gene regions colocalized with the target trait over the total true colocalized gene regions. FDR is defined as the proportion of incorrectly identified gene regions colocalized with the target trait over all identified regions. In practice, OPERA may report multiple overlapping subsets ambiguous as to which true colocalization they correspond to due to a lack of variant level information. For instance, for cases with two distinct colocalizations {1,2} and {1,2,3} where trait 1 is the target trait, OPERA often reports three subsets {1,2,3}, {1,2} and {1,3}, the last of which is ambiguous as to whether it is a false positive driven by a incorrect causal variant, or merely subset of {1,2,3} with the correct underlying causal variant. We adopted a lenient evaluation strategy for OPERA: for both the ground truth and reported colocalization, we merged any subsets that are identical or strictly nested into their largest compatible set (here, {1,2,3}) to enable a consistent gene-level assessment. To compare on unified gene-level basis, we likewise apply the same merging strategy to *ColocBoost* CoS, despite it often identifies multiple distinct and correct colocalization events per gene without merging. This procedure ensures a fair comparison for OPERA at gene-level despite underestimating the power of *ColocBoost* at event-level and overlooking finer colocalization patterns it can capture.

### Unified variant-level benchmarking for other competing colocalization methods

Because many existing colocalization methods report only a single top-ranked variant, we observed inflated FDRs, particularly under multiple causal variants scenarios, due to failure to account for LD-driven uncertainty in variant selection (**Figure S2e**). In contrast, our proposed colocalization confidence set (CoS) framework offers a more informative and robust summary (**Figure 2c**). However, since CoS is unique to *ColocBoost* and not natively supported by competing methods, we developed a unified evaluation approach by extending each method to construct 95% credible sets analogous to our 95% CoS. These were built using reported variant-level probabilities of each method (detailed below) and filtered with the same purity threshold (requiring purity>0.5) to ensure fair and interpretable comparisons. The construction of 95% CoS analogs for each method is as follows:

- COLOC (V5): for colocalization configuration where PP.H4 is greater than other PPs (PP.H0, PP.H1, PP.H2, PP.H3), we use ‘SNP.PP.H4’ as variant-level scores to define 95% colocalization credible set similar to how we defined 95% CoS. Unlike many analyses that apply a high PP.H4 cutoff (e.g., 0.7–0.8), we do not impose one here for methods comparison purposes, as the COLOC paper does not rigorously recommend a particular threshold, and our simulations show that omitting it preserves well-controlled FDR while maintaining power comparable to *ColocBoost*. Nonetheless, users requiring more stringent evidence can adopt a higher PP.H4 cutoff in practice.
- HyPrColoc: for identified colocalizations with posterior probability higher than uncolocalized configurations, we used ‘snp.scores’ as the variant-level scores to define 95% colocalization credible set. As with COLOC, we omit an explicit configuration probability cutoff here for fair comparison.
- MOLOC: for colocalization configuration where PPA.ab is greater than other PPAs (PPA.a, PPA.b, PPA.a,b, and zero), we used ‘SNP.PP.ab’ as the variant-level scores to define 95% colocalization credible set.

### Multi-omics and GWAS data sources and preprocessing

In our primary analyses, we used 17 *cis*-xQTL contexts spanning 16,928 genes and 3 modalities (gene expression, splicing, protein abundance) from the aging brain cortex of ROSMAP donors (average *N*=595; **Table 2**)^22,24,43–45^. Bulk RNA-seq were generated from postmortem tissues across the dorsolateral prefrontal cortex (DLPFC), posterior cingulate cortex (PCC), and head of caudate nucleus (AC). Bulk monocytes were isolated from peripheral blood mononuclear cells through negative selection, followed by fluorescent antibody labeling and sorting of live CD14+CD16-cells for downstream analysis^106^. Single-nucleus RNA-seq was performed on 479 DLPFC samples and we focused on six major cell types with abundant cell proportions to robustly perform eQTL calling: excitatory neurons (Exc), inhibitory neurons (Inh), microglia (Mic), oligodendrocyte precursor cells (OPC), oligodendrocytes (Oli), and astrocytes (Ast)^43^. For bulk RNA-seq, standard QC and adapter trimming was performed on raw reads, followed by alignment via STAR and gene expression quantification via RNASeqQC^22^. Genes were removed if over 20% samples have TPM expression level of 10% or less. Quantile normalization was applied to gene-level expression. Additionally, LeafCutter2 was applied to generate productive and unproductive splicing events for sQTL analysis, as described in another FunGen-xQTL companion project^46^. For snRNA-seq data, gene expressions were summed into pseudo bulk counts for each donor-cell pair, quantified as counts per million (CPM) and then quantile normalized. Selected Reaction Monitoring (SRM) proteomics was performed on DLPFC with manual inspection to ensure correct peak assignment and peak boundaries^24^. After quality control and filtering for proteins quantified in at least 50% of samples, the peptide relative abundances were log2 transformed and centered at the median, followed by missing protein values imputation using the grouped Empirical Bayes Matrix Factorization (gEBMF) method^107^. For all normalized molecular phenotypes, we adjusted the technical factors (batch, RNA integrity number, post-mortem interval), biological covariates (sex, age at death), the top 15 genotype PC, and additional hidden confounders using PCA on each normalized phenotype matrix with the number of PCs determined by the Marchenko-Pastur limit.

In GTEx eQTLs colocalization analysis, normalized gene expression for 17,834 genes from all samples across 13 GTEx brain tissues and genotype data were downloaded from the open access dataset from public release GTEx Analysis V8^6^ (average *N*=168; **Table S3**). Following from recommendations of GTEx consortium we adjusted 68 covariates on both normalized gene expression and genotype, including top 5 genotyping PCs, 60 PEER factors, sequencing platform and protocol, and sex.

For AD-xQTL colocalization, we constructed a custom LD reference panel from the Alzheimer’s Disease Sequencing Project (ADSP), consisting of approximately 17,000 whole-genome sequenced (WGS) individuals of European ancestry. We primarily used a recent AD GWAS meta-analysis summary statistics^67^ (*N*_*cases*_ =111,326, *N*_*controls*_ =677,663) after performing quality control against this reference panel, including allele harmonization and LD mismatch detection using SLALOM^108^, with flagged suspicious variants removed. As a secondary AD GWAS, we considered meta-analysis summary statistics with a smaller effective sample size especially for non-proxy AD cases^68^ (*N*_*cases*_ = 86,531, *N*_*controls*_ = 676,386 including 46,613 proxy cases from UK Biobank) (**Supplementary Note S.8**).

### Excess of overlap (EOO) analysis for variant-gene links

We quantify the concordance between two sets of variant-gene (VG) links using an *excess of overlap (EOO) statistic*^51^, computed by comparing the fraction of shared VG pairs to their expected overlap by chance. Specifically, for sets *VG*_1_ and *VG*_2_ of sizes |*VG*_1_| and |*VG*_2_| out of a total tested *T*_1_ and *T*_2_ VG pairs, with *T*_12_ pairs commonly tested by both methods, we define:

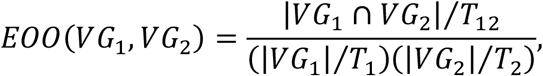

so that values above 1 indicate more overlap than expected by chance. Standard error of EOO can be estimated by leave-one-chromosome-out strategy. Analogously, we define discordance as the fraction of variants linked to different genes between two sets, divided by the amount of discordance expected by chance.

In evaluating element-gene maps from CRISPR or promoter-capture datasets, we adopt different definitions of *T* and *VG* tailored to specific tasks. We defined *VG* for element-gene maps as all pairs of variant-gene links with the element of interest. For variant-gene pairs across the genome (total 360,092,003), *VG* is defined by the 95% CoS-gene links from 95% CoS-gene links for *ColocBoost*, SNP-Gene links of marginal QTL associations, and fine-mapped eQTL 95% credible set-gene links. For the cell-type-specific analyses, we focus on 95% CoS-gene links (433,927 identified), and *VG* is defined to by the variant-gene pair for the corresponding cell-type.

### MaxVCP-based variant annotation and heritability analysis

To evaluate whether variants implicated by multi-trait colocalization inform disease risk, we constructed a new variant annotation based on the *maximum variant colocalization probability* (MaxVCP). Specifically, for variants in 95% CoS obtained by *ColocBoost*, let *G*_*j*_ denote the set of genes whose 95% CoS contains variant *j*, we then define

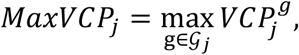

where 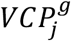 is the variant colocalization probability for variant *j* and gene *g*. Using ROSMAP and GTEx data we created five distinct MaxVCP based annotations as previously described in **Results**, for 9,991,229 variants with minor allele count ≥ 5 from 1000 Genomes Project Europeans (**Table S1**). We then applied S-LDSC to quantify how well these annotations capture disease heritability across 57 GWAS summary statistics data-sets (**Table S6**), conditioning on 97 functional annotations from the baseline-LD v2.2 model^56^. We consider two metrics, heritability enrichment and standardized effect size *τ*^∗^. We meta-analyze these metrics across all 57 traits, as well as separately across the blood and brain-related traits.

Enrichment is defined as the relative contribution of heritability explained by colocalized annotation (MaxVCP),

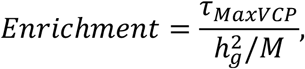

where 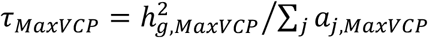 quantifies heritability explained by per-variant within colocalized annotation. Denote 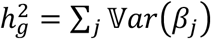 as the estimated variant heritability across all annotations, then 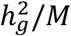 represents the proportion of heritability explained by per-variant within all annotations, where *M* is the total number of variants and V*ar*(*β*_*j*_) is the per-variant heritability of variant *j*. Standardized effect size (*τ*^∗^) is defined as the proportionate change in per-variant heritability associated with a one standard deviation increase in the value of the colocalized annotation, conditional on other annotations included in the model. Both enrichment and 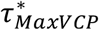 are estimated via a genomic block-jackknife procedure with 200 blocks that also provides standard errors. We considered two versions of 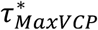: a marginal version conditional on baseline-LD annotations only, and a joint version where the conditioning set includes other colocalization-derived annotations in addition to baseline-LD. More detailed description of metrics can be found in **Supplementary Note S.7**.

### Benchmarking AD-colocalized CoS-gene links against ENCODE-rE2G regulatory annotations

We benchmarked AD-colocalized 95% CoS-gene links from *ColocBoost* and COLOC (V5) against element-gene links in the ENCODE-rE2G model, using precision and recall to assess concordance. *Recall* was defined as the fraction of high-confidence element-gene links—*i*.*e*., distinct element-gene pairs across all biosamples harboring AD GWAS loci— captured by each colocalization method. To define distinct regulatory elements, overlapping links across biosamples were merged using *bedtools*, and we retained those elements containing at least one AD GWAS variant with p-value < 10^−5^ (known as the “suggestive” GWAS significance threshold). *Precision* was defined as the fraction of 95% CoS-gene links from each method that overlapped with ENCODE-rE2G element-gene links.

## Data Availability

All data produced in the present study are available upon reasonable request to the authors

https://github.com/StatFunGen/colocboost-paper

## Data availability

The complete xQTL summary statistics for the 17 brain xQTL molecular contexts, including expression, splicing and protein abundance xQTL summary statistics are available to the reviewers through the following Synapse link (https://www.synapse.org/Synapse:syn66511452) with username “statfungen_guest” and password “cao_et_al_april_2025” and LD reference from the Alzheimer’s Disease Sequencing Project (ADSP) are available at the following Synapse link (https://www.synapse.org/Synapse:syn53171227), for reviewers to assess our work. These data will formally be available at the time of publication via The National Institute on Aging Genetics of Alzheimer’s Disease Data Storage Site (NIAGADS) at https://adsp.niagads.org/, as part of the FunGen-xQTL data release.

The download information of full colocalization results from the xQTL-only and GWAS-xQTL *ColocBoost* can be found in **Supplementary Table 1** and **Supplementary Data**, including the CoS and VCP scores.

**Supplementary Data** for numerical results with reproducible codes is available at https://doi.org/10.5281/zenodo.15215225. The Supplementary Data is categorized into Figures and Supplementary Figures, with a ‘data’ sub-folder and ‘codes’ sub-folder under each category comprising of the numerical results (in .*rds* format, to be loaded into R using *readRDS*), and the codes to generate the figure panels from the data respectively.

MaxVCP-based colocalization annotations in the S-LDSC analysis are available at https://doi.org/10.5281/zenodo.15215225 (under ‘Supplementary_Tables/annotations_maxVCP’ sub-folder) and 97 baseline-LD v2.2 annotations in the S-LDSC analysis are available at https://console.cloud.google.com/storage/browser/broad-alkesgroup-public-requester-pays/ UKBiobank fine-mapping data: https://www.finucanelab.org/data MVP fine-mapping data: Verma et al^66^. https://drive.google.com/drive/folders/1SKm5caoFziLBu5VST2zt0TevbMLNRHqm

ENCODE-rE2G predictions are available on the ENCODE portal https://www.encodeproject.org/search/?type=Annotation&searchTerm=encoderE2G

Element-gene maps from Promoter Capture Hi-C assay in the dorsolateral prefrontal cortex brain tissue are available at Table S3 in ref^34^ https://www.sciencedirect.com/science/article/pii/S0092867416313228 - app2

Combined CRISPRi data (KRAB-dCas9) is available as part of the CRISPR benchmarking pipeline at https://github.com/EngreitzLab/CRISPR_comparison/tree/main/resources/crispr_data

Element-gene links from the STING-seq assay using an updated CRISPRi protocol (KRAB-dCas9-MeCP2) is available at Table S3F in Morris, J.A. et al^29^. https://www.science.org/doi/10.1126/science.adh7699#supplementary-materials

cV2F scores are available at Table S3 in Fabiha, T. et al^53^. https://mskcc.box.com/shared/static/hsrogtr3fddtmd53hyy5ph7dlp20eq72.txt

## Code availability

ColocBoost (v1.0) is implemented in the R package *colocboost*, freely available on CRAN (https://cran.r-project.org/web/packages/colocboost/index.html) and at https://github.com/StatFunGen/colocboost. The code and supplementary data to reproduce main and supplementary figures are available at https://statfungen.github.io/colocboost-paper.

## Acknowledgements

We thank members of the Wang and Dey labs for discussions and support. We thank the members of the Alzheimer’s Disease Sequencing Project Functional Genomics Consortium (FunGen-AD) for providing the FunGen-xQTL resource. We thank Dr. Daniel Nachun (Stanford) and Pei Tian (Columbia) for assistance in the development of colocboost R package. This work was supported by NIH/NIA R01AG076901 (G.W., X.C. H.S), a FunGen-AD Consortium Grant U01AF072572 (G.W., R.F., P.L.), a grant from The Urbut Family Foundation (G.W.), NIH/NHGRI grant R00HG012203 (K.K.D., H.S), NIH/NHGRI grant R01HG014008 (K.K.D.), NIH/NCI Cancer Center Support Grant P30CA008748 (K.K.D, H.S), Josie Robertson Investigator Program (K.K.D.), NIH/NIGMS R35GM153249 (Y.I.L.) and a GREGoR Consortium Grant (Y.I.L. and C.F.B.A.N.).

## Author contributions

G.W., K.K.D. jointly supervised research. X.C., H.S., R.M., K.K.D., G.W. developed the method and algorithm. X.C., H.S., K.K.D., G.W. conceived and designed the experiments, performed analyses, interpreted the results and implemented to *colocboost* R package with documentation and example vignettes. R.F. contributed to testing *colocboost* R package and developing bioinformatics pipelines implementing *ColocBoost* on real-world data. R.F., C.F.B.A.N., Y.I.L. contributed splicing QTL data production. P.L.D., D.B., G.W. contributed and supervised other molecular QTL data production. X.C., H.S., R.F., K.K.D., G.W. wrote and revised the manuscript.

All authors critically reviewed the manuscript, suggested revisions as needed and approved the final version.

## Supplementary Figures Legend

**Figure S1. Computational framework of *ColocBoost* for multi-trait colocalization analysis. a**. Comparison between standard gradient boosting algorithm and proximity smoothing gradient boosting algorithm in *ColocBoost*. Standard gradient boosting updates one variant in each iteration based on the strongest association of the genotypic profile of variants with the residual phenotype. Proximity smoothing gradient boosting updates this candidate best variant as well as some of its strong LD neighbors through an adaptive smoothing, determined by the proximal LD and the strength of the signal. **b**. Schematic representations of multi-trait weak learner, Single-Effect Couplers (SEC), across two traits that models a single causal variant by first evaluating the coupling across traits and then performing proximity smoothed updates at the variant for each of the coupled traits. **c**. Delayed-SEC (D-SEC) updates across two traits are designed to remove or substantially weaken uncoupled signals before performing coupled updates. **d**. Comprehensive workflow diagram depicting the integration of SEC and D-SEC within an iterative gradient boosting informed algorithm in *ColocBoost* for multi-trait colocalization analysis.

**Figure S2. Extended performance comparison of *ColocBoost* against other multi-trait colocalization methods. a**. Schematics of a broad range of simulation designs used for comprehensive benchmarking, including realistic simulation analyses, simulation designs adopted in original publications of competing methods, and simulation with correlated traits. (See details in **Supplementary Note**). **b**. Statistical power and FDR comparison of *ColocBoost* and COLOC (V5) across prior colocalization probabilities *p*_12_ *∈* {10^−4^, 10^−5^, 10^−6^, 10^−7^}, with default *p*_12_ = 5 × 10^−6^ also shown, in a two-trait simulation with 1 to 3 causal variants. **c**. Scatter plot of the negative logarithm of False Discovery Rate (-log(FDR)) and power of *ColocBoost* and HyPrColoc across varying numbers of true causal variants (see **Methods** for details on the computation of power and FDR). The vertical red line represents the FDR cut-off of 0.5. **d**. Size of CoS and purity comparisons in simulation designs involving 2, 5, 10, and 20 traits. **e**. Statistical power and FDR of *ColocBoost* and HyPrColoc in simulation design involving 50 traits, up to five causal variant per trait per locus. **f**. Statistical power and FDR of *ColocBoost* and HyPrColoc in simulation designs adopted in HyPrColoc with 10 traits and four different causal configurations of 10 traits (codenamed “3+3+2+2”, “5+5”, “5+5+1rand”, “5+5+1rand+1rand” with definitions provided in **Supplementary Note S.6**). **g**. Statistical power and FDR of *ColocBoost* and HyPrColoc in a fully colocalized scenario where multiple causal variants affects all traits, involving 5,10, and 20 phenotypes with up to five true causal variants. **h**. Comprehensive comparison of type I error rate of *ColocBoost* and HyPrColoc under a null scenario with no causal variants across any trait. The error bars in panels **c**-**g** represent 95% confidence intervals. Numerical results for these analyses are reported in **Supplementary Data**.

**Figure S3. Additional metrics to benchmark performance of *ColocBoost* with other multi-trait colocalization methods. a**. Statistical power and FDR of all methods based on the single top-ranked variant, involving 2, 5,10, and 20 phenotypes with up to five true causal variants, with genotype data and induced colocalization configurations designed to mimic real xQTL datasets (**Methods**). **b**. Statistical power and FDR comparison of *ColocBoost* and OPERA for GWAS targeted colocalization, evaluated at the gene level, under the same simulation design used in OPERA. **c**. Statistical power and FDR comparison and **d**. variant-level precision-recall curves by varying the colocalization score threshold in a simulation design incorporating trait-trait correlations across 2, 5,10, and 20 phenotypes with up to five true causal variants. **e**. Statistical power and FDR of *ColocBoost* and HyPrColoc assuming diagonal LD matrix and imposing a single-causal-variant assumption without LD-proximity smoothing in *ColocBoost*. **f**. Statistical power and FDR for single trait fine-mapping analysis (*FineBoost*) with up to five causal variants using SuSiE and *FineBoost* (the single-trait version of *ColocBoost*) in simulation scenarios comprising up to five causal variants. Error bars in panels **a**-**f** represent 95% confidence intervals. Numerical results are reported in Supplementary Data.

**Figure S4. Calibration of single-effect conditional inclusion probability, omega, and variant probability of association, VPA, in fine-mapping analysis. a**. Single-effect conditional inclusion probabilities (omega) when the true causal variant lies in a perfect-LD block (top) or among independent variants (bottom). **b**. Distribution of omega for true causal variants across 1,000 simulations, showing equal weights within the perfect-LD block (left) and concentration near 1 in the independent-variant setting (right). **c**. Assessment of omega calibration. Variants across all simulations were grouped into 20 equally spaced bins according to their reported omega. The plots show the average omega for each bin against the proportion of effect variants in that bin. For each bin, the x-axis shows the mean reported probability and y-axis shows the fraction of variants that are truly causal in the simulations. A well-calibrated method should produce points near the x = y line. Gray error bars show 95% confidence intervals. The comparison panel evaluates omega from *FineBoost* against alpha from SuSiE. **d**. Sensitivity analysis of omega to temperature parameter *τ*, showing KL divergence (left) and correlation with default *τ* = 0.01 (right) across values ranging from 0.001 to 100. **e**. Sensitivity analysis of omega to regularization parameter *λ*, showing KL divergence (left) and correlation with default *λ* = 0.5 (right) across values ranging from 0 to 1. **f–g**. Assessment of VPA calibration. Variants across all simulations were grouped into bins according to their reported VPA. The plots show the average VPA for each bin against the proportion of effect variants in that bin. A well-calibrated method should produce points near the x = y line. Gray error bars show 95% confidence intervals. The comparison panel evaluates VPA from *FineBoost* against PIP from SuSiE. See detailed definitions of each parameter in **Supplementary Note S.5.2**. Numerical results are reported in **Supplementary Data**.

**Figure S5. Hyperparameter sensitivity and robustness analysis for *ColocBoost***. Performances of *ColocBoost* with different hyperparameters are evaluated under the simulation designs involving 2, 5, and 10 traits, up to five causal variants per trait per locus, with genotype data and induced colocalization configurations designed to mimic real xQTL datasets. **a**. Simulation studies to benchmark different choice of probabilistic simplex *δ*^*k*^ used in proximity smoothing in *ColocBoost*. **b**. Sensitivity analysis of *λ ∈* [0,1] that controls the strength of proximity smoothing in *ColocBoost*. **c**. Sensitivity analysis of threshold to determine the equivalence across best updates. (**d, e**). Sensitivity of two parameters related to D-SEC, code-named *coloc_thresh* and *func_compare*. **f**. Evaluation of performance for different weight fudge factor *ν*. (**g, h**). Robustness against weak spurious signals removing criterions. **i**. Robustness of correlation between confidence sets. See detailed definitions of each parameters in **Supplementary Note S.5.2**. Numerical results are reported in **Supplementary Data**.

**Figure S6. Colocalization performance in split-sample analyses. a**. Concordance between *ColocBoost* VCP and COLOC SNP.PP.H4 in pairwise simulation studies, shown separately for loci where both methods identify the same CoS variants (left) and loci with partially overlapping variants (right). **b–c**. Split-sample reproducibility analysis of AD GWAS colocalization with DLPFC bulk eQTL (784 samples) using e. two and f. three independent splits. UpSet plots show the overlap of colocalized loci identified by COLOC (left) and *ColocBoost* (right) across the full dataset (d) and independent subsets (d1, d2, d3). Barplots show the proportion of SuSiE 95% CS recovered by each method, evaluated across all 95% CS and single-CS gene regions. **d**. Split-sample reproducibility analysis of low-density lipoprotein (LDL) cholesterol GWAS from UK Biobank with GTEx liver eQTL. UpSet plots show the overlap of colocalized loci identified by COLOC (left) and ColocBoost (right) using the full UK Biobank sample (d) and two random sample splits (d1 and d2).

**Figure S7. Extended *ColocBoost* xQTL analysis in aging human brain**. *ColocBoost* was applied to 17 gene-level cis-xQTL datasets from the aging brain cortex of ROSMAP subjects. **a**. UpSet plot summarizing the top 80 colocalized events in xQTL-only *ColocBoost* analysis across 17 molecular traits. **b**. UpSet plot displaying colocalization patterns across 3 molecular modalities (expression, splicing, and protein abundance) restricted to 3,655 genes with data available for all modalities. **c**. The fraction of cell-type specific fine-mapping (measuring as 95% credible sets) for different brain cell types, that are recovered by cell-type specific colocalizations separately. **d**. Summary of cell-type specific and cell-type shared colocalization events from (left penal) xQTL colocalization analysis spanning 17 molecular traits, and (right panel) eQTL colocalization analysis spanning eQTL data from 6 brain cell types and 4 bulk tissues. The diagonal entries indicate the number of cell-type specific colocalizations and off-diagonal entries indicate the number of colocalizations shared between each pairs of cell types. **e**. Excess-of-overlap (EOO) analysis of the cell-type specific colocalization of eGenes with cell-type gene programs from external brain single-cell RNA-seq data. Numerical results are reported in **Supplementary Data**.

**Figure S8. Functional enrichment of colocalized variants. a**. Excess-of-overlap (EOO) analysis of “mappable” CoS (number of colocalized variants ≤ 3) and other 95% CoS with cV2F score and RegulomeDB annotations. **b**. Enrichment analysis of variants comprising “mappable” CoS are significantly enriched in regulatory functional annotations from baseline-LD v2.2, including enhancer, promoter, histone modifications, genomic features, conservation, and other annotations. Error bars denote 95% confidence intervals. Numerical results are reported in **Supplementary Data**.

**Figure S9. Properties of genes with multiple causal variants. a**. Scatter plot of the change in log-likelihood between primary CoS and a secondary CoS, where each point is a primary-secondary CoS pair for the gene. **b**. A boxplot summarizing the relationship between the number of colocalized traits and log likelihood changes. **c**. Functional annotation enrichment comparison between high MaxVCP scored variants (MaxVCP>0.5) in primary CoS versus secondary CoS, including annotations from enhancer, promoter, coding, and repressed categories. **d**. An example case of xQTL-only colocalization in gene *ARSB* showing two CoS, one colocalized among all brain cell types and the other colocalized across microglia, astrocytes and oligodendrocytes only. Error bars denote 95% confidence intervals. Numerical results are reported in **Supplementary Data**.

**Figure S10. xQTL-only *ColocBoost* for eQTLs across 13 brain tissues in GTEx. a**. UpSet plot summarizing the top 50 colocalized events in xQTL-only *ColocBoost* analysis across 13 brain tissues in GTEx. **b**. CoS-level overlap across ROSMAP xQTL colocalization events, ROSMAP eQTL colocalization events, and GTEx colocalization events involving brain cortex (3 tissues). **c**. Properties of genes with multiple causal variants, including log likelihood changes between primary and secondary CoS, the relationship between the number of colocalized traits and log likelihood changes, and the number of pairs of primary and secondary CoS across brain tissue eQTLs. Error bars denote 95% confidence intervals. Numerical results are reported in **Supplementary Data**.

**Figure S11. Validation of *ColocBoost* colocalization. a**. Variant set-level excess-of-overlap (EOO) analysis of (i) 95% CoS-gene links from xQTL-only *ColocBoost*, (ii) standard marginal xQTL-gene associations, merged across all 17 xQTL datasets and (iii) 95% credible set (CS)-gene links from SuSiE fine-mapping, merged across all 17 xQTL datasets, against 118,389 regulatory element-gene maps from Promoter Capture Hi-C assay in the dorsolateral prefrontal cortex brain tissue, which profiles targeted 3D contacts linking gene promoters to proximal or distal regulatory elements. (**b, c**) EOO analysis of 95% CoS-gene links across (b) 3 different brain cortical regions and CD14+/CD16-bulk monocytes, and (c) 6 different brain cell types, against regulatory element-gene maps from PCHi-C data. **e**. Colocalization for *SUOX* in detail, also validated by CRIPRi in K562 cells. Error bars denote 95% confidence intervals. Numerical results are reported in **Supplementary Data**.

**Figure S12. Extended disease heritability analyses of variant-level functional annotations derived from *ColocBoost*. a**. Correlation matrix of five functional annotations based on the MaxVCP scores from *ColocBoost*. **b**. Heritability enrichment and standardized effect sizes, meta-analyzed across all, brain-related and blood-related traits, conditional on 97 baseline-LD v2.2 annotations of the 5 versions of xQTL MaxPPH derived from HyPrColoc. **c**. Standardized effect sizes, conditional on the 97 baseline-LD v2.2 annotations + MaxVCP-xQTL and CoS-xQTL annotations (joint τ^∗^). **d**. Standardized effect sizes, conditional on the 97 baseline-LD v2.2 annotations + MaxVCP-xQTL and binarized MaxVCP-xQTL scores at different binarization thresholds (joint τ^∗^). **e**. Heritability enrichment and standardized effect sizes (marginal τ^∗^ and joint τ^∗^) of MaxVCP-xQTL against MaxPIP annotation from SuSiE fine-mapping analysis. **f**. EOO analysis of MaxVCP-xQTL in the disease fine-mapped variants (PIP>0.95) from 94 UK Biobank traits and 930 Million Veteran Program (MVP) traits. **g**. Heritability enrichment and standardized effect sizes (marginal τ^∗^ and joint τ^∗^) of MaxVCP-xQTL annotations for cell-type-specific variants and cell-type-shared variants. Error bars denote 95% confidence intervals. Numerical results are reported in **Supplementary Data**. Asterisk denotes FDR<5% is computed by Bonferroni correction of the p-values for the number of annotations tested in the S-LDSC analysis.

**Figure S13. Extended evaluations of GWAS-xQTL colocalization from *ColocBoost* and competing methods. a**. Precision-recall analysis comparing CoS-gene links from AD-xQTL *ColocBoost*, COLOC-Union, pairwise-*ColocBoost*-union, and a version of AD-xQTL *ColocBoost* limited to AD fine-mapped variants (*ColocBoost*-finemapped-GWAS) against enhancer-gene links predicted by ENCODE-rE2G across 354 biosamples. Error bars along both axis indicate 95% confidence intervals. **b**. Precision-Recall for the comparison with enhancer-gene links for each eQTL. COLOC-union only identified colocalization events for a few xQTL, and is excluded from the figure due to a lack of reliable standard error estimation. Manhattan plot of the MaxVCP functional annotation scores of variants from **c**. Pairwise-*ColocBoost*-union and **d**. COLOC-union, with labeled genes containing variants with MaxVCP>0.5. Microglia contributions are highlighted in green. Error bars denote 95% confidence intervals. Numerical results are reported in **Supplementary Data**.

**Figure S14. Additional summary of genome-wide GWAS-xQTL colocalization results. a**. UpSet plot of colocalization events and identified by AD-xQTL *ColocBoost*. **b**. Manhattan plot of MaxVCP scores, with labeled genes containing variants with MaxVCP>0.1, and microglia contributions are highlighted in green. **c**. Excess overlap of genes showing colocalization in microglia with three microglia specific gene-sets as benchmarks. Error bars denote 95% confidence intervals. Numerical results are reported in **Supplementary Data**.

**Figure S15. Example cases of xQTL colocalizations in known AD risk genes. a**. Example of established AD gene *BIN1* across multiple molecular contexts and **b**. example of colocalized variants spanning multiple genes *CR1, CR2, YOD1*. One variant, rs669515 (MaxVCP=0.755), colocalized with *CR2* in DLPFC bulk eQTLs, *YOD1* in oligodendrocytes, and *CR1* across oligodendrocytes and all three additional bulk cortical eQTLs. Numerical results are reported in **Supplementary Data**.

**Figure S16. Additional details of colocalizations in gene *CTSH*. a**. Marginal association p-values (at −log10 scale) and **b**. estimated multi-trait coefficients by *ColocBoost*, including all molecular events colocalized. Numerical results are reported in **Supplementary Data**.

**Figure S17. Calibration of residual correlation across phenotypes for colocalization analysis. a**. Performance of *ColocBoost* under residual correlations *ρ ∈* {0.3,0.5,0.7,0.9} based on an AR(1) model, reporting power, FDR, median CoS size, and mean set purity across simulations with 2, 5, and 10 traits and varying numbers of causal variants per trait. The corresponding independent-sample baseline (*ρ* = 0) is reported in Figure 2 and Figures S2. **b**. Estimated residual correlation matrix across ten eQTL datasets from the ROSMAP cohort, representing cell-type-specific and bulk-tissue contexts with 100% sample overlap.

**Figure S18. Impact of LD reference mismatch on summary statistics based inference**. Data were simulated using GTEx genotypes (*N* = 838), whereas LD was estimated from independent European-ancestry reference panels from either the Alzheimer’s Disease Sequencing Project (ADSP; *N* = 19,292) or the 1000 Genomes Project (*N* = 489). We randomly selected 500 TAD regions from a total of 1,361 TADs spanning the entire genome. SuSiE exhibited a higher false discovery rate (FDR) than *FineBoost* (the single-trait version of *ColocBoost*) when the smaller 1000 Genomes reference panel was used. The larger ADSP reference panel improved FDR control, consistent with reduced LD reference mismatch, as widely accepted in the literature.

## Supplementary Tables Legend

**Table S1**. xQTL-only and AD-xQTL *ColocBoost* colocalization loci and variants.

**Table S2**. Significantly enriched pathways for genes underlying neuron- and microglia-specific colocalizations.

**Table S3**. GTEx eQTL from 13 bulk brain tissues.

**Table S4**. *ColocBoost* 95% CoS-gene links validated by two CRISPRi data.

**Table S5**. List of annotations used in S-LDSC.

**Table S6**. List of 57 complex diseases used in disease heritability enrichment analysis.

**Table S7**. List of 129 variants with MaxVCP>0.5 enriched in variants confidently fine-mapped (PIP>0.95) in 94 UK Biobank traits and 930 Million Veteran Program (MVP) GWAS traits.

**Table S8**. Heritability enrichment results for AD GWAS in Bellenguez 2022 conditional on 97 baseline-LD annotations.

**Table S9**. Feature comparisons of multi-trait fine-mapping and colocalization methods (green: feature supported or trivial to enable).

**Table S10**. Realistic cross-trait sharing patterns from genome-wide fine-mapping results from 62 FunGen-xQTL contexts by calculating how often signals overlapped among those contexts and then sampled from these observed overlap frequencies to define what subset of traits each simulated causal variant share.

