## Supplementary Figures for "Integrative multi-omics QTL colocalization maps regulatory architecture in aging human brain"

**Figure S1. Computational framework of *ColocBoost* for multi-trait colocalization analysis. a**. Comparison between standard gradient boosting algorithm and proximity smoothing gradient boosting algorithm in *ColocBoost*. Standard gradient boosting updates one variant in each iteration based on the strongest association of the genotypic profile of variants with the residual phenotype. Proximity smoothing gradient boosting updates this candidate best variant as well as some of its strong LD neighbors through an adaptive smoothing, determined by the proximal LD and the strength of the signal. **b**. Schematic representations of multi-trait weak learner, Single-Effect Couplers (SEC), across two traits that models a single causal variant by first evaluating the coupling across traits and then performing proximity smoothed updates at the variant for each of the coupled traits. **c**. Delayed-SEC (D-SEC) updates across two traits are designed to remove or substantially weaken uncoupled signals before performing coupled updates. **d**. Comprehensive workflow diagram depicting the integration of SEC and D-SEC within an iterative gradient boosting informed algorithm in *ColocBoost* for multi-trait colocalization analysis.

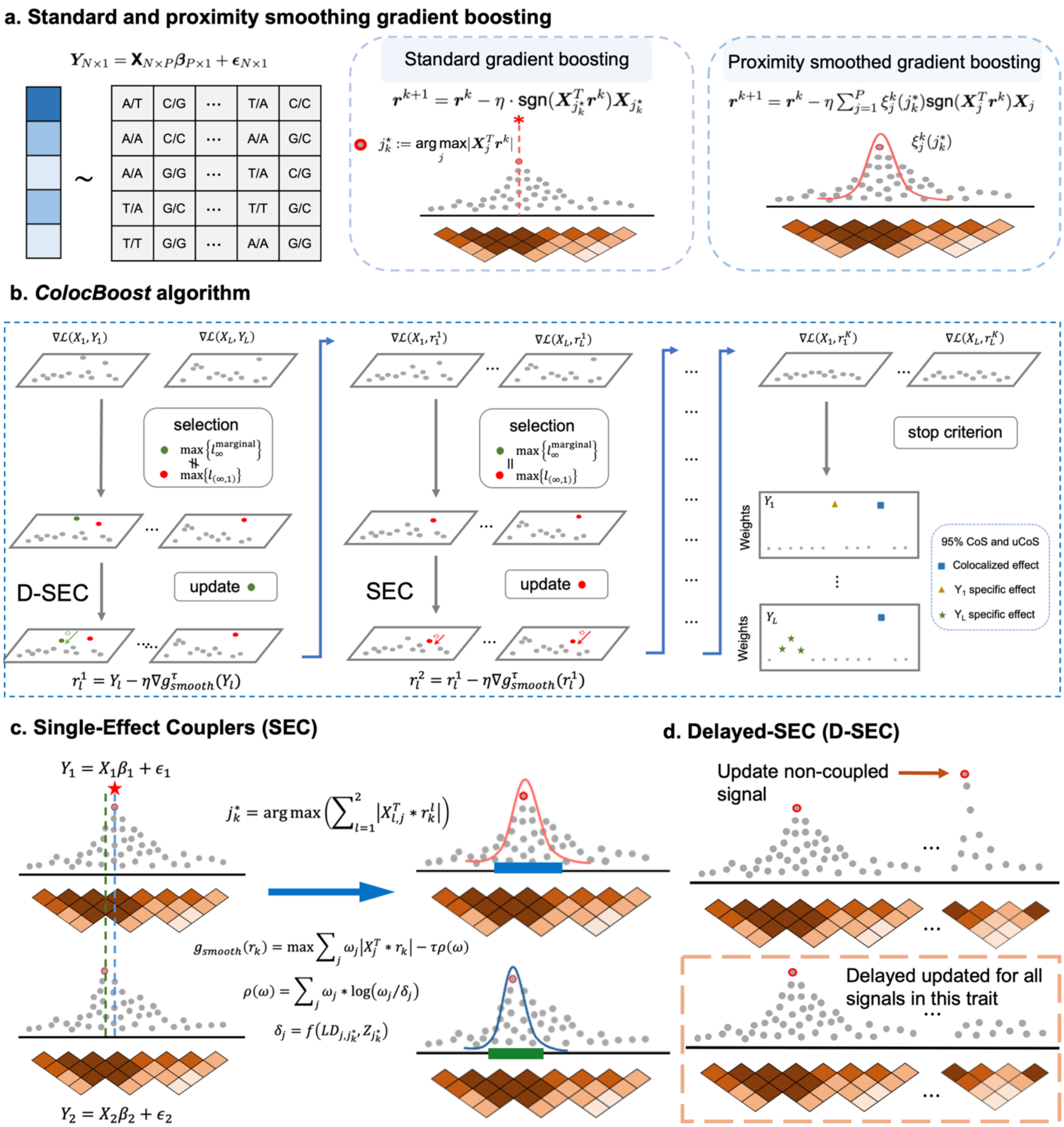

a. Standard and proximity smoothing gradient boosting
$\mathbf{Y}_{N\times 1} = \mathbf{X}_{N\times P}\boldsymbol{\beta}_{P\times 1} + \boldsymbol{\epsilon}_{N\times 1}$
Standard gradient boosting
$\boldsymbol{r}^{k+1} = \boldsymbol{r}^{k} - \eta \cdot \text{sgn}(\boldsymbol{X}_{j_k^*}^T \boldsymbol{r}^k)\boldsymbol{X}_{j_k^*}$
$j_k^* := \arg\max_j |\boldsymbol{X}_j^T \boldsymbol{r}^k|$
Proximity smoothed gradient boosting
$\boldsymbol{r}^{k+1} = \boldsymbol{r}^{k} - \eta \sum_{j=1}^{P} \xi_j^k(j_k^*)\text{sgn}(\boldsymbol{X}_j^T \boldsymbol{r}^k)\boldsymbol{X}_j$
$\xi_j^k(j_k^*)$
b. ColocBoost algorithm
$\nabla\mathcal{L}(X_1, Y_1)$
$\nabla\mathcal{L}(X_L, Y_L)$
selection
$\max\{l_\infty^{\text{marginal}}\}$
$\max\{l_{(\infty,1)}\}$
D-SEC
update
$r_l^1 = Y_l - \eta\nabla g_{smooth}^{\tau}(Y_l)$
$\nabla\mathcal{L}(X_1, r_1^1)$
$\nabla\mathcal{L}(X_L, r_L^1)$
SEC
$r_l^2 = r_l^1 - \eta\nabla g_{smooth}^{\tau}(r_l^1)$
$\nabla\mathcal{L}(X_1, r_1^K)$
$\nabla\mathcal{L}(X_L, r_L^K)$
stop criterion
Weights
95% CoS and uCoS
Colocalized effect
$Y_1$ specific effect
$Y_L$ specific effect
c. Single-Effect Couplers (SEC)
$Y_1 = X_1\beta_1 + \epsilon_1$
$j_k^* = \arg\max\left(\sum_{l=1}^{2}\left|X_{l,j}^T * r_k^l\right|\right)$
$g_{smooth}(r_k) = \max\sum_j \omega_j|X_j^T * r_k| - \tau\rho(\omega)$
$\rho(\omega) = \sum_j \omega_j * \log(\omega_j/\delta_j)$
$\delta_j = f(LD_{j,j_k^*}, Z_{j_k^*})$
$Y_2 = X_2\beta_2 + \epsilon_2$
d. Delayed-SEC (D-SEC)
Update non-coupled signal
Delayed updated for all signals in this trait

**Figure S2. Extended performance comparison of *ColocBoost* against other multi-trait colocalization methods. a.** Schematics of a broad range of simulation designs used for comprehensive benchmarking, including realistic simulation analyses, simulation designs adopted in original publications of competing methods, and simulation with correlated traits. (See details in **Supplementary Note**). **b.** Statistical power and FDR comparison of *ColocBoost* and COLOC (V5) across prior colocalization probabilities $p_{12} \in \{10^{-4}, 10^{-5}, 10^{-6}, 10^{-7}\}$, with default $p_{12} = 5 \times 10^{-6}$ also shown, in a two-trait simulation with 1 to 3 causal variants. **c.** Scatter plot of the negative logarithm of False Discovery Rate (-log(FDR)) and power of *ColocBoost* and HyPrColoc across varying numbers of true causal variants (see **Methods** for details on the computation of power and FDR). The vertical red line represents the FDR cut-off of 0.5. **d.** Size of CoS and purity comparisons in simulation designs involving 2, 5, 10, and 20 traits. **e.** Statistical power and FDR of *ColocBoost* and HyPrColoc in simulation design involving 50 traits, up to five causal variant per trait per locus. **f.** Statistical power and FDR of *ColocBoost* and HyPrColoc in simulation designs adopted in HyPrColoc with 10 traits and four different causal configurations of 10 traits (codenamed "3+3+2+2", "5+5", "5+5+1rand", "5+5+1rand+1rand" with definitions provided in **Supplementary Note S.6**). **g.** Statistical power and FDR of *ColocBoost* and HyPrColoc in a fully colocalized scenario where multiple causal variants affects all traits, involving 5,10, and 20 phenotypes with up to five true causal variants. **h.** Comprehensive comparison of type I error rate of *ColocBoost* and HyPrColoc under a null scenario with no causal variants across any trait. The error bars in panels **c**-**g** represent 95% confidence intervals. Numerical results for these analyses are reported in **Supplementary Data**.

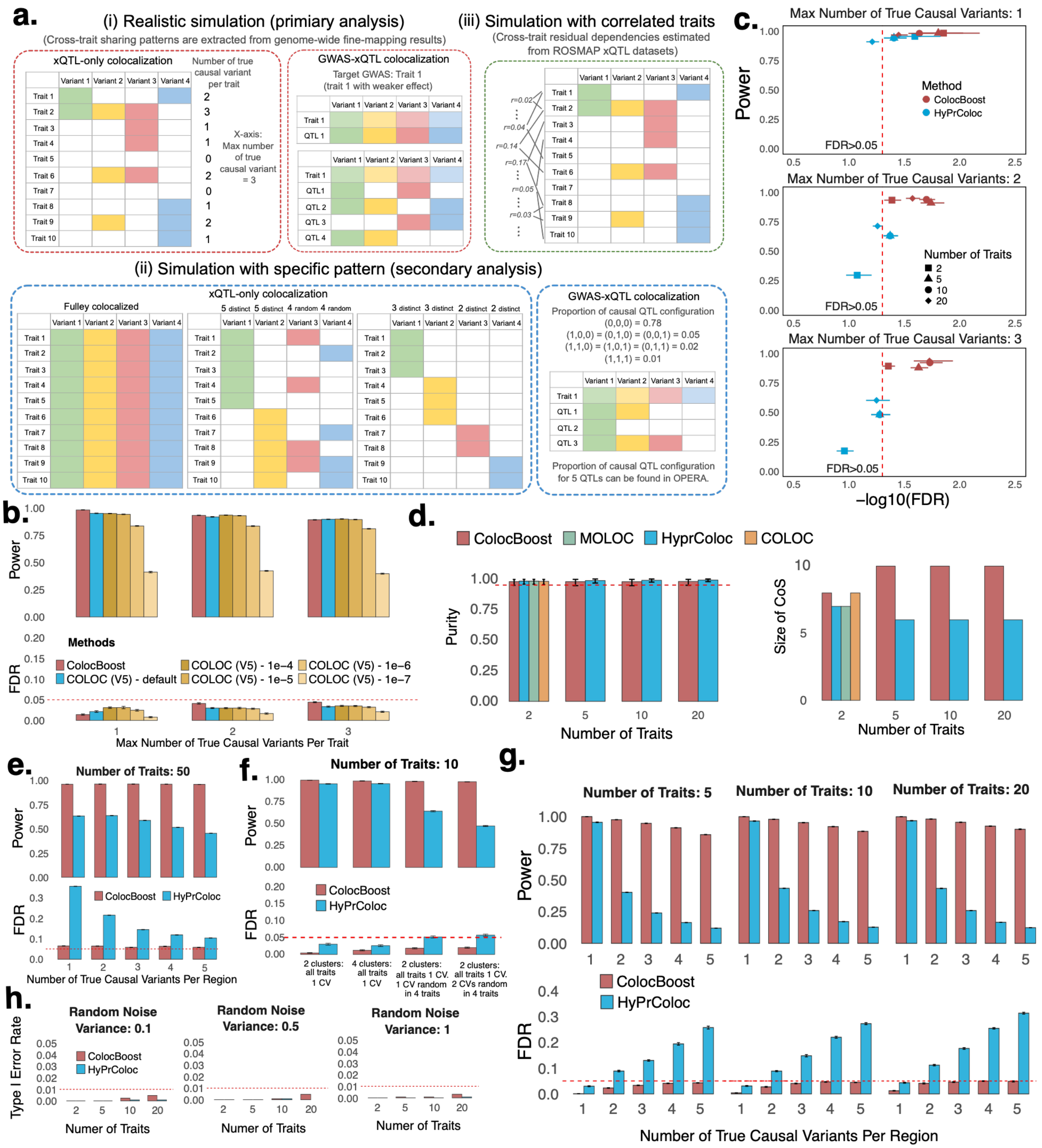

a.
(i) Realistic simulation (primiary analysis)
(Cross-trait sharing patterns are extracted from genome-wide fine-mapping results)
xQTL-only colocalization
Number of true causal variant per trait
X-axis: Max number of true causal variant = 3
GWAS-xQTL colocalization
Target GWAS: Trait 1 (trait 1 with weaker effect)
(iii) Simulation with correlated traits
(Cross-trait residual dependencies estimated from ROSMAP xQTL datasets)
(ii) Simulation with specific pattern (secondary analysis)
xQTL-only colocalization
Fulley colocalized
5 distinct 5 distinct 4 random 4 random
3 distinct 3 distinct 2 distinct 2 distinct
GWAS-xQTL colocalization
Proportion of causal QTL configuration
(0,0,0) = 0.78
(1,0,0) = (0,1,0) = (0,0,1) = 0.05
(1,1,0) = (1,0,1) = (0,1,1) = 0.02
(1,1,1) = 0.01
Proportion of causal QTL configuration for 5 QTLs can be found in OPERA.
b.
Power
FDR
Methods
ColocBoost
COLOC (V5) - default
COLOC (V5) - 1e−4
COLOC (V5) - 1e−5
COLOC (V5) - 1e−6
COLOC (V5) - 1e−7
Max Number of True Causal Variants Per Trait
c.
Max Number of True Causal Variants: 1
Max Number of True Causal Variants: 2
Max Number of True Causal Variants: 3
Power
Method
ColocBoost
HyPrColoc
Number of Traits
FDR>0.05
−log10(FDR)
d.
ColocBoost
MOLOC
HyprColoc
COLOC
Purity
Size of CoS
Number of Traits
e.
Number of Traits: 50
Power
FDR
ColocBoost
HyPrColoc
Number of True Causal Variants Per Region
f.
Number of Traits: 10
Power
FDR
ColocBoost
HyPrColoc
2 clusters: all traits 1 CV
4 clusters: all traits 1 CV
2 clusters: all traits 1 CV. 1 CV random in 4 traits
2 clusters: all traits 1 CV. 2 CVs random in 4 traits
g.
Number of Traits: 5
Number of Traits: 10
Number of Traits: 20
Power
FDR
ColocBoost
HyPrColoc
Number of True Causal Variants Per Region
h.
Random Noise Variance: 0.1
Random Noise Variance: 0.5
Random Noise Variance: 1
Type I Error Rate
ColocBoost
HyPrColoc
Numer of Traits

**Figure S3. Additional metrics to benchmark performance of *ColocBoost* with other multi-trait colocalization methods. a**. Statistical power and FDR of all methods based on the single top-ranked variant, involving 2, 5,10, and 20 phenotypes with up to five true causal variants, with genotype data and induced colocalization configurations designed to mimic real xQTL datasets (**Methods**). **b.** Statistical power and FDR comparison of *ColocBoost* and OPERA for GWAS targeted colocalization, evaluated at the gene level, under the same simulation design used in OPERA. **c.** Statistical power and FDR comparison and **d.** variant-level precision-recall curves by varying the colocalization score threshold in a simulation design incorporating trait-trait correlations across 2, 5,10, and 20 phenotypes with up to five true causal variants. **e.** Statistical power and FDR of *ColocBoost* and HyPrColoc assuming diagonal LD matrix and imposing a single-causal-variant assumption without LD-proximity smoothing in *ColocBoost*. **f.** Statistical power and FDR for single trait fine-mapping analysis (*FineBoost*) with up to five causal variants using SuSiE and *FineBoost* (the single-trait version of *ColocBoost*) in simulation scenarios comprising up to five causal variants. Error bars in panels **a**-**f** represent 95% confidence intervals. Numerical results are reported in Supplementary Data.

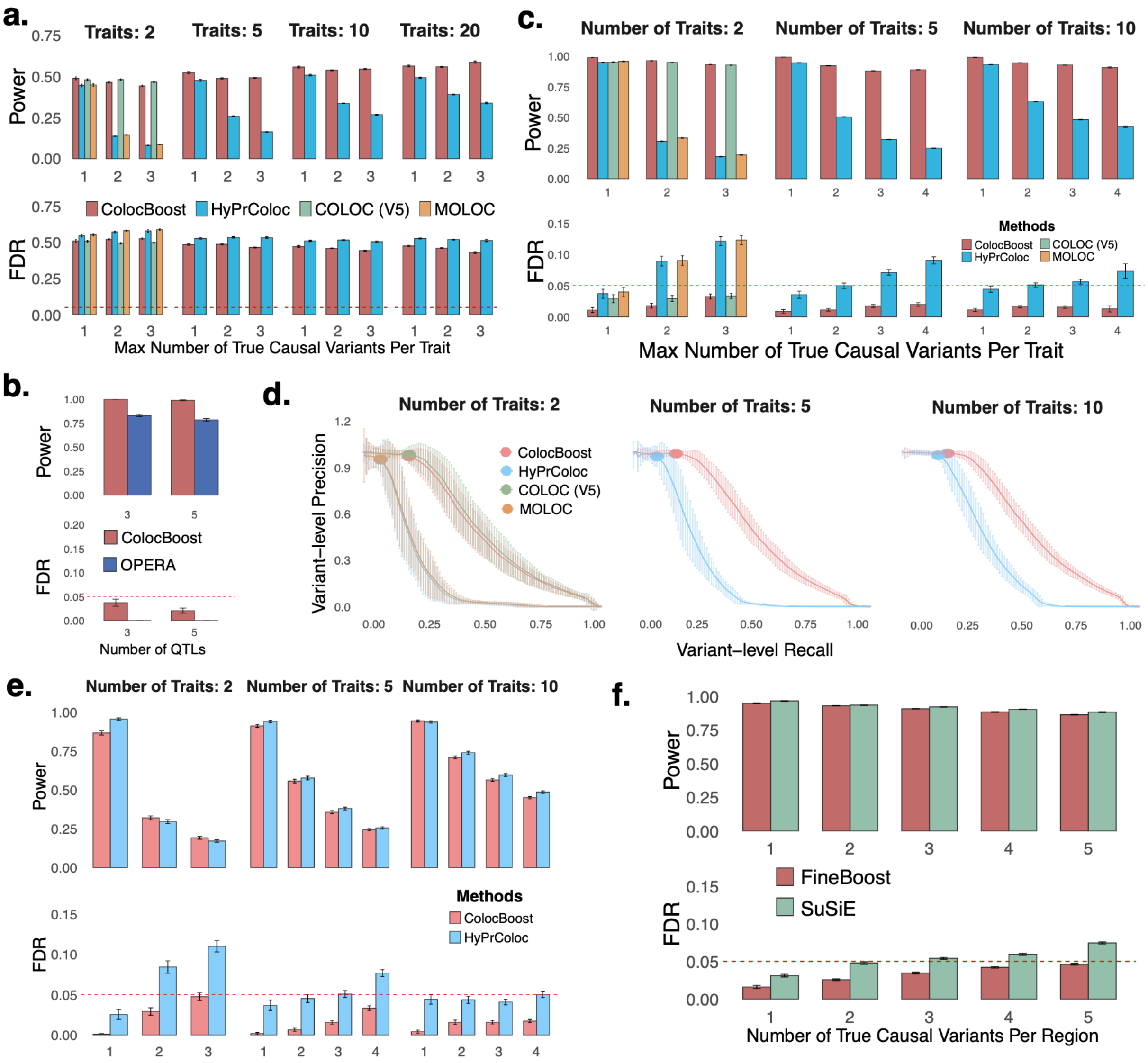

a.
Traits: 2
Traits: 5
Traits: 10
Traits: 20
Power
FDR
ColocBoost
HyPrColoc
COLOC (V5)
MOLOC
Max Number of True Causal Variants Per Trait
b.
Power
FDR
ColocBoost
OPERA
Number of QTLs
c.
Number of Traits: 2
Number of Traits: 5
Number of Traits: 10
Power
FDR
Methods
ColocBoost
HyPrColoc
COLOC (V5)
MOLOC
Max Number of True Causal Variants Per Trait
d.
Number of Traits: 2
Number of Traits: 5
Number of Traits: 10
Variant−level Precision
Variant−level Recall
ColocBoost
HyPrColoc
COLOC (V5)
MOLOC
e.
Number of Traits: 2
Number of Traits: 5
Number of Traits: 10
Power
FDR
Methods
ColocBoost
HyPrColoc
f.
Power
FDR
FineBoost
SuSiE
Number of True Causal Variants Per Region

**Figure S4. Calibration of single-effect conditional inclusion probability, omega, and variant probability of association, VPA, in fine-mapping analysis. a.** Single-effect conditional inclusion probabilities (omega) when the true causal variant lies in a perfect-LD block (top) or among independent variants (bottom). **b.** Distribution of omega for true causal variants across 1,000 simulations, showing equal weights within the perfect-LD block (left) and concentration near 1 in the independent-variant setting (right). **c.** Assessment of omega calibration. Variants across all simulations were grouped into 20 equally spaced bins according to their reported omega. The plots show the average omega for each bin against the proportion of effect variants in that bin. For each bin, the x-axis shows the mean reported probability and y-axis shows the fraction of variants that are truly causal in the simulations. A well-calibrated method should produce points near the x = y line. Gray error bars show 95% confidence intervals. The comparison panel evaluates omega from *FineBoost* against alpha from SuSiE. **d.** Sensitivity analysis of omega to temperature parameter $\tau$, showing KL divergence (left) and correlation with default $\tau = 0.01$ (right) across values ranging from 0.001 to 100. **e.** Sensitivity analysis of omega to regularization parameter $\lambda$, showing KL divergence (left) and correlation with default $\lambda = 0.5$ (right) across values ranging from 0 to 1. **f–g.** Assessment of VPA calibration. Variants across all simulations were grouped into bins according to their reported VPA. The plots show the average VPA for each bin against the proportion of effect variants in that bin. A well-calibrated method should produce points near the x = y line. Gray error bars show 95% confidence intervals. The comparison panel evaluates VPA from *FineBoost* against PIP from SuSiE. See detailed definitions of each parameter in **Supplementary Note S.5.2**. Numerical results are reported in **Supplementary Data**.

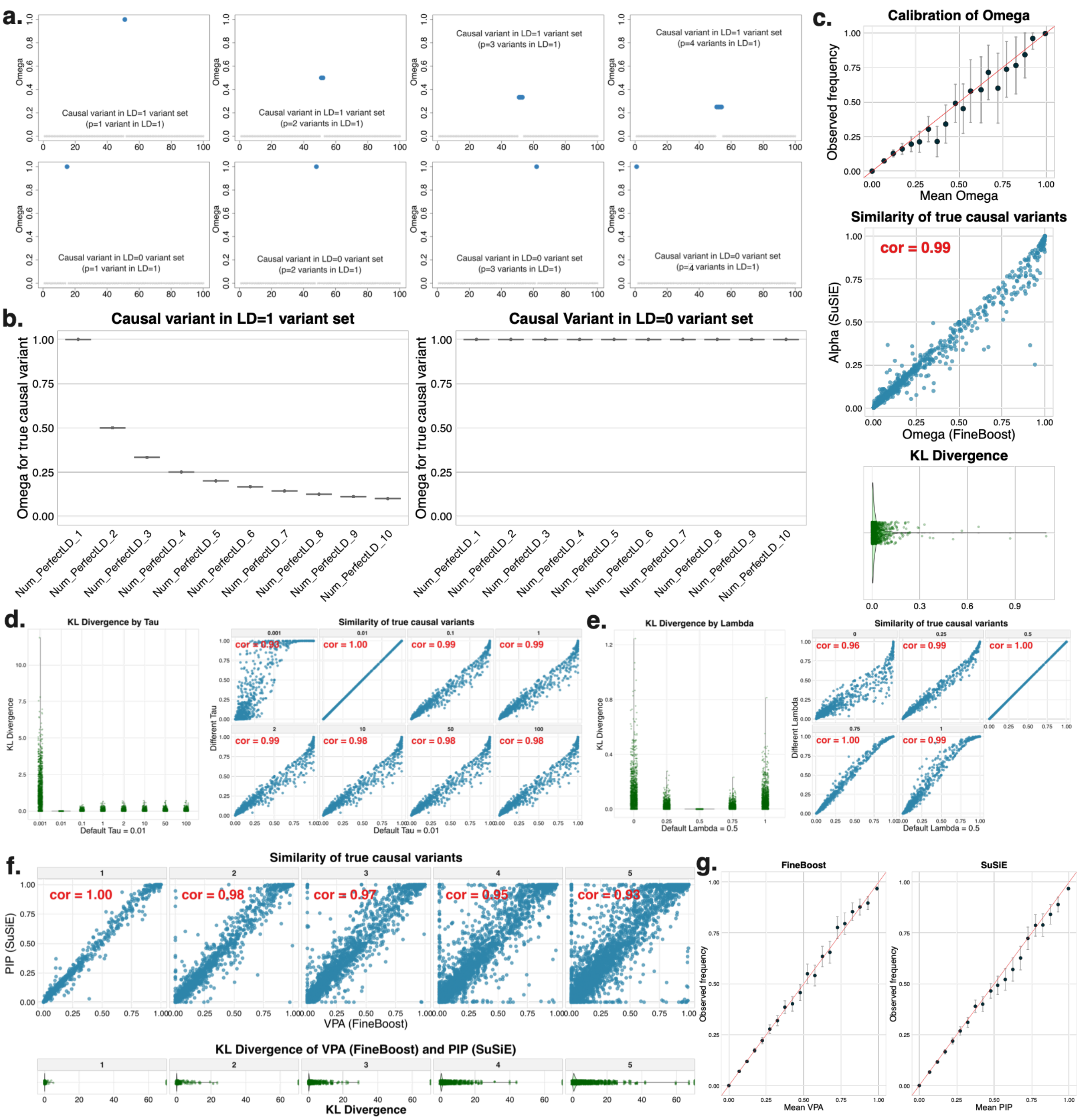
a.
Causal variant in LD=1 variant set
(p=1 variant in LD=1)
Causal variant in LD=1 variant set
(p=2 variants in LD=1)
Causal variant in LD=1 variant set
(p=3 variants in LD=1)
Causal variant in LD=1 variant set
(p=4 variants in LD=1)
Causal variant in LD=0 variant set
(p=1 variant in LD=1)
Causal variant in LD=0 variant set
(p=2 variants in LD=1)
Causal variant in LD=0 variant set
(p=3 variants in LD=1)
Causal variant in LD=0 variant set
(p=4 variants in LD=1)
Omega
b.
Causal variant in LD=1 variant set
Causal Variant in LD=0 variant set
Omega for true causal variant
Num_PerfectLD_1
Num_PerfectLD_2
Num_PerfectLD_3
Num_PerfectLD_4
Num_PerfectLD_5
Num_PerfectLD_6
Num_PerfectLD_7
Num_PerfectLD_8
Num_PerfectLD_9
Num_PerfectLD_10
c.
Calibration of Omega
Observed frequency
Mean Omega
Similarity of true causal variants
cor = 0.99
Alpha (SuSiE)
Omega (FineBoost)
KL Divergence
d.
KL Divergence by Tau
KL Divergence
Default Tau = 0.01
Similarity of true causal variants
Different Tau
cor = 0.93
cor = 1.00
cor = 0.99
cor = 0.99
cor = 0.99
cor = 0.98
cor = 0.98
cor = 0.98
Default Tau = 0.01
e.
KL Divergence by Lambda
KL Divergence
Default Lambda = 0.5
Similarity of true causal variants
Different Lambda
cor = 0.96
cor = 0.99
cor = 1.00
cor = 1.00
cor = 0.99
Default Lambda = 0.5
f.
Similarity of true causal variants
PIP (SuSiE)
cor = 1.00
cor = 0.98
cor = 0.97
cor = 0.95
cor = 0.93
VPA (FineBoost)
KL Divergence of VPA (FineBoost) and PIP (SuSiE)
KL Divergence
g.
FineBoost
SuSiE
Observed frequency
Mean VPA
Mean PIP

**Figure S5. Hyperparameter sensitivity and robustness analysis for *ColocBoost*.** Performances of *ColocBoost* with different hyperparameters are evaluated under the simulation designs involving 2, 5, and 10 traits, up to five causal variants per trait per locus, with genotype data and induced colocalization configurations designed to mimic real xQTL datasets. **a**. Simulation studies to benchmark different choice of probabilistic simplex $\delta^k$ used in proximity smoothing in *ColocBoost*. **b**. Sensitivity analysis of $\lambda \in [0,1]$ that controls the strength of proximity smoothing in *ColocBoost*. **c.** Sensitivity analysis of threshold to determine the equivalence across best updates. (**d, e**). Sensitivity of two parameters related to D-SEC, code-named *coloc_thresh* and *func_compare*. **f**. Evaluation of performance for different weight fudge factor $\nu$. (**g, h**). Robustness against weak spurious signals removing criterions. **i**. Robustness of correlation between confidence sets. See detailed definitions of each parameter in **Supplementary Note S.5.2**. Numerical results are reported in **Supplementary Data**.

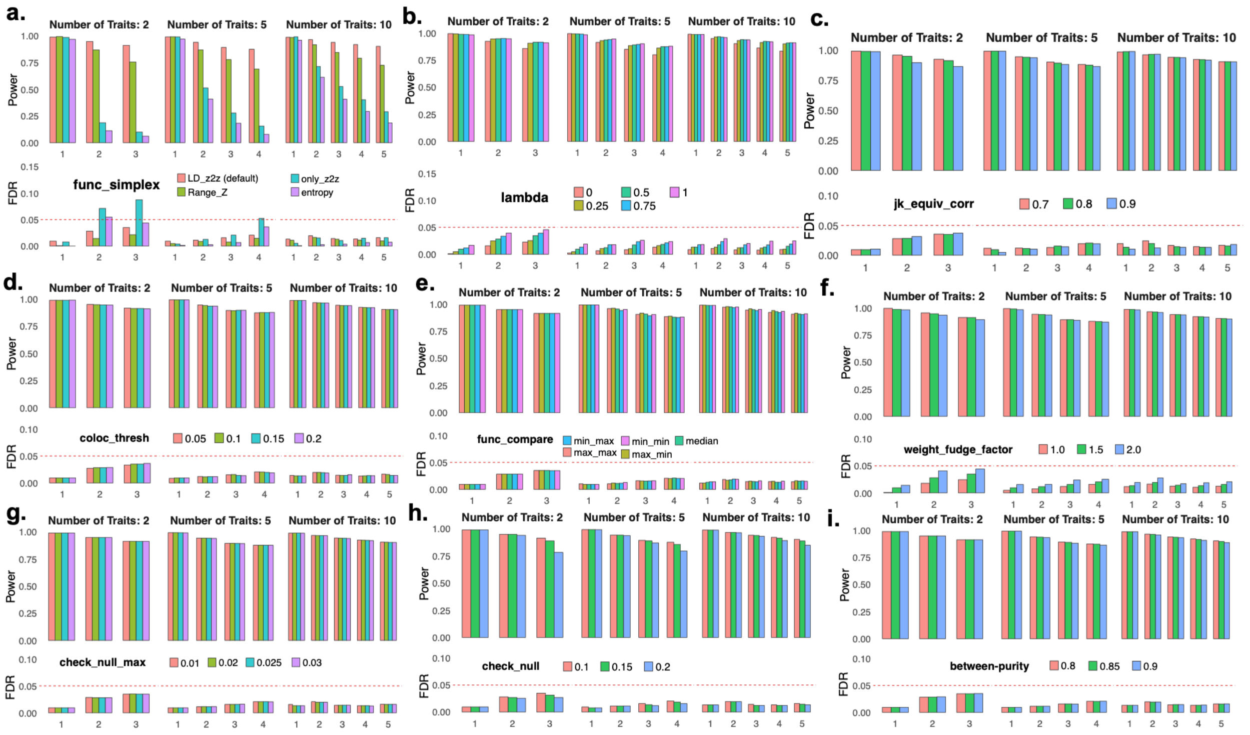

**Figure S6. Colocalization performance in split-sample analyses. a.** Concordance between *ColocBoost* VCP and COLOC SNP.PP.H4 in pairwise simulation studies, shown separately for loci where both methods identify the same CoS variants (left) and loci with partially overlapping variants (right). **b–c**. Split-sample reproducibility analysis of AD GWAS colocalization with DLPFC bulk eQTL (784 samples) using e. two and f. three independent splits. UpSet plots show the overlap of colocalized loci identified by COLOC (left) and *ColocBoost* (right) across the full dataset (d) and independent subsets (d1, d2, d3). Barplots show the proportion of SuSiE 95% CS recovered by each method, evaluated across all 95% CS and single-CS gene regions. **d.** Split-sample reproducibility analysis of low-density lipoprotein (LDL) cholesterol GWAS from UK Biobank with GTEx liver eQTL. UpSet plots show the overlap of colocalized loci identified by COLOC (left) and ColocBoost (right) using the full UK Biobank sample (d) and two random sample splits (d1 and d2).

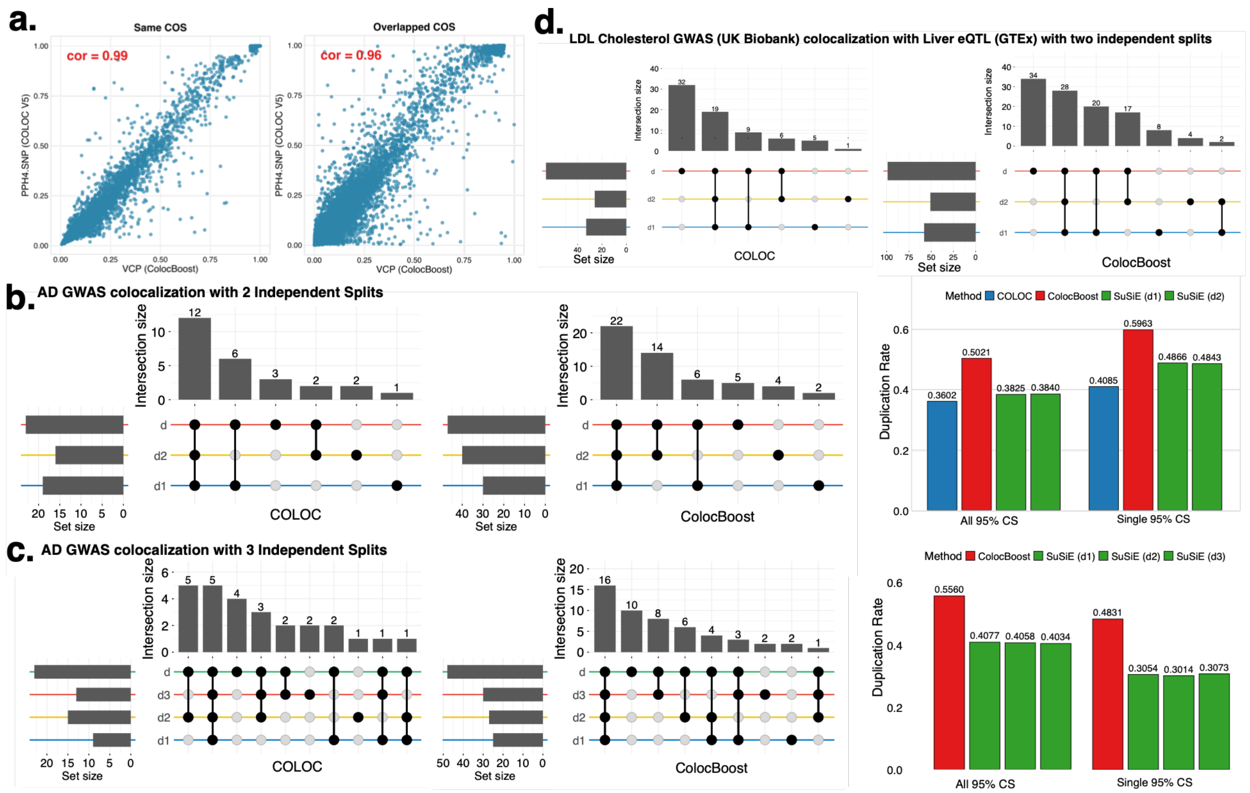

**Figure S7. Extended *ColocBoost* xQTL analysis in aging human brain.** *ColocBoost* was applied to 17 gene-level cis-xQTL datasets from the aging brain cortex of ROSMAP subjects. **a**. UpSet plot summarizing the top 80 colocalized events in xQTL-only *ColocBoost* analysis across 17 molecular traits. **b.** UpSet plot displaying colocalization patterns across 3 molecular modalities (expression, splicing, and protein abundance) restricted to 3,655 genes with data available for all modalities. **c.** The fraction of cell-type specific fine-mapping (measuring as 95% credible sets) for different brain cell types, that are recovered by cell-type specific colocalizations separately. **d.** Summary of cell-type specific and cell-type shared colocalization events from (left penal) xQTL colocalization analysis spanning 17 molecular traits, and (right panel) eQTL colocalization analysis spanning eQTL data from 6 brain cell types and 4 bulk tissues. The diagonal entries indicate the number of cell-type specific colocalizations and off-diagonal entries indicate the number of colocalizations shared between each pairs of cell types. **e.** Excess-of-overlap (EOO) analysis of the cell-type specific colocalization of eGenes with cell-type gene programs from external brain single-cell RNA-seq data. Numerical results are reported in **Supplementary Data**.

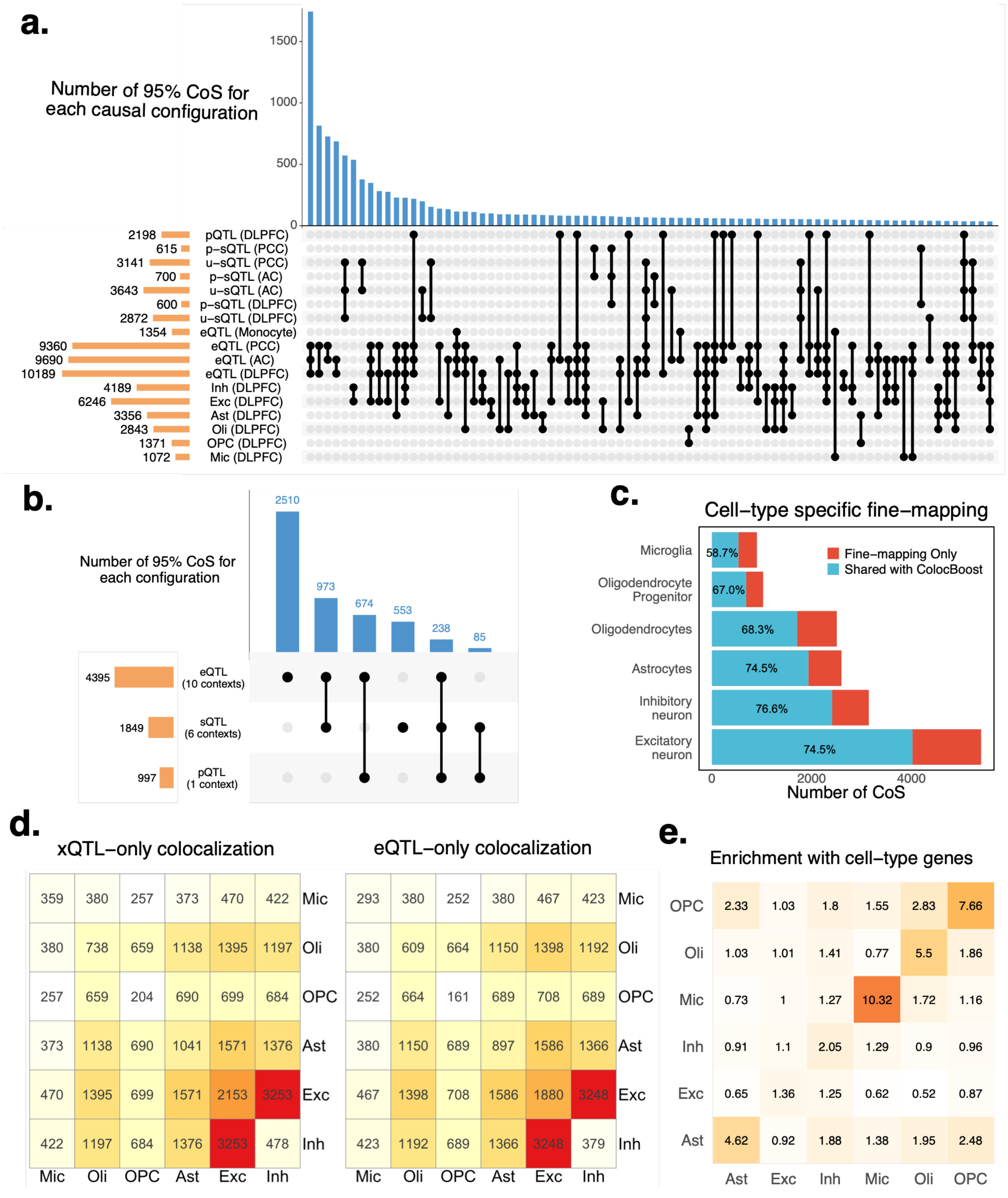
a.
Number of 95% CoS for each causal configuration
1500
1000
500
0
2198 pQTL (DLPFC)
615 p–sQTL (PCC)
3141 u–sQTL (PCC)
700 p–sQTL (AC)
3643 u–sQTL (AC)
600 p–sQTL (DLPFC)
2872 u–sQTL (DLPFC)
1354 eQTL (Monocyte)
9360 eQTL (PCC)
9690 eQTL (AC)
10189 eQTL (DLPFC)
4189 Inh (DLPFC)
6246 Exc (DLPFC)
3356 Ast (DLPFC)
2843 Oli (DLPFC)
1371 OPC (DLPFC)
1072 Mic (DLPFC)
b.
Number of 95% CoS for each configuration
2510
973
674
553
238
85
4395 eQTL (10 contexts)
1849 sQTL (6 contexts)
997 pQTL (1 context)
c.
Cell–type specific fine–mapping
Microglia 58.7%
Oligodendrocyte Progenitor 67.0%
Oligodendrocytes 68.3%
Astrocytes 74.5%
Inhibitory neuron 76.6%
Excitatory neuron 74.5%
Fine–mapping Only
Shared with ColocBoost
0
2000
4000
Number of CoS
d.
xQTL–only colocalization
359 380 257 373 470 422 Mic
380 738 659 1138 1395 1197 Oli
257 659 204 690 699 684 OPC
373 1138 690 1041 1571 1376 Ast
470 1395 699 1571 2153 3253 Exc
422 1197 684 1376 3253 478 Inh
Mic Oli OPC Ast Exc Inh
eQTL–only colocalization
293 380 252 380 467 423 Mic
380 609 664 1150 1398 1192 Oli
252 664 161 689 708 689 OPC
380 1150 689 897 1586 1366 Ast
467 1398 708 1586 1880 3248 Exc
423 1192 689 1366 3248 379 Inh
Mic Oli OPC Ast Exc Inh
e.
Enrichment with cell–type genes
OPC 2.33 1.03 1.8 1.55 2.83 7.66
Oli 1.03 1.01 1.41 0.77 5.5 1.86
Mic 0.73 1 1.27 10.32 1.72 1.16
Inh 0.91 1.1 2.05 1.29 0.9 0.96
Exc 0.65 1.36 1.25 0.62 0.52 0.87
Ast 4.62 0.92 1.88 1.38 1.95 2.48
Ast Exc Inh Mic Oli OPC

**Figure S8. Functional enrichment of colocalized variants a.** Excess-of-overlap (EOO) analysis of "mappable" CoS (number of colocalized variants $\leq 3$) and other 95% CoS with cV2F score and RegulomeDB annotations. **b.** Enrichment analysis of variants comprising "mappable" CoS are significantly enriched in regulatory functional annotations from baseline-LD v2.2, including enhancer, promoter, histone modifications, genomic features, conservation, and other annotations. Error bars denote 95% confidence intervals. Numerical results are reported in **Supplementary Data**.

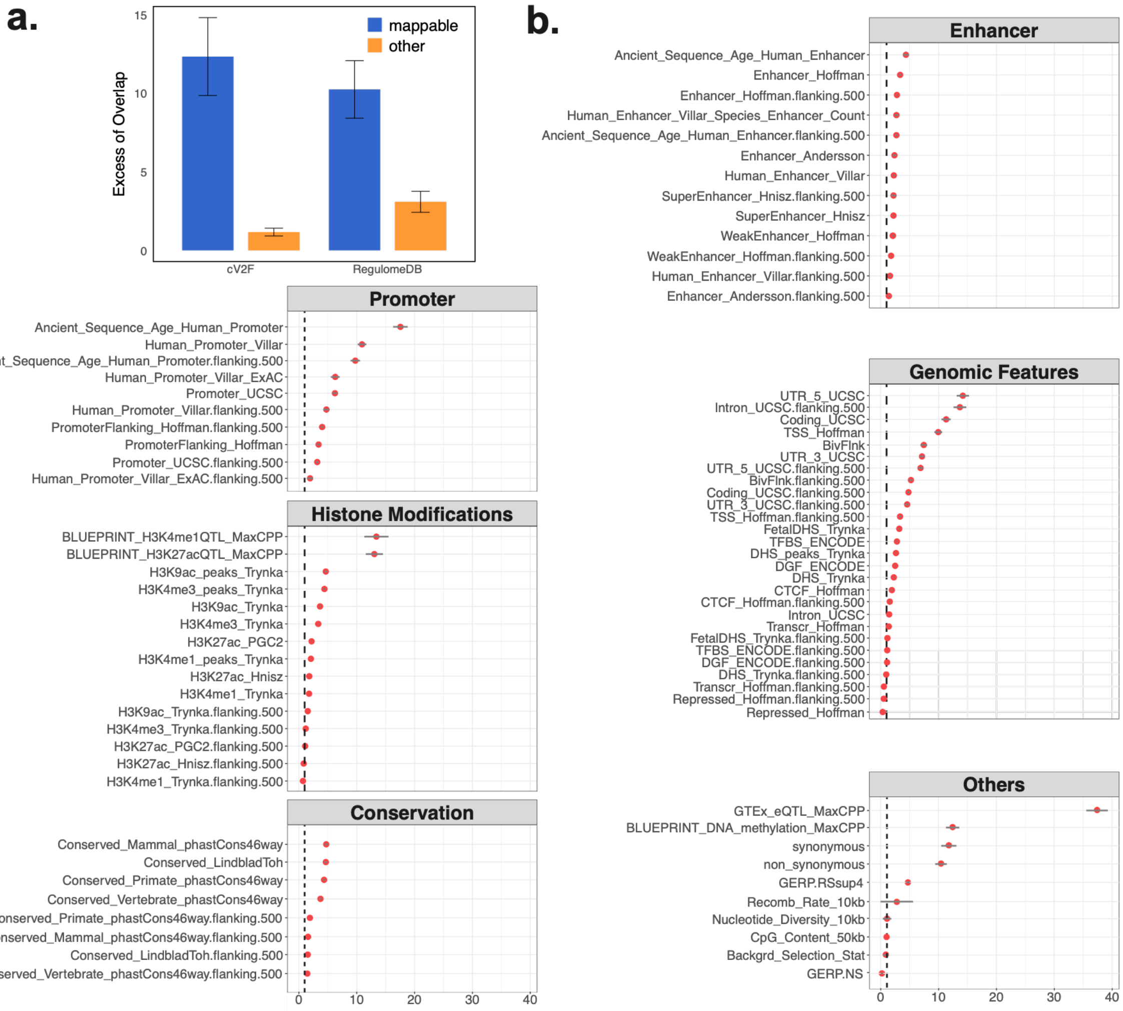

**Figure S9. Properties of genes with multiple causal variants. a.** Scatter plot of the change in log-likelihood between primary CoS and a secondary CoS, where each point is a primary-secondary CoS pair for the gene. **b.** A boxplot summarizing the relationship between the number of colocalized traits and log likelihood changes. **c.** Functional annotation enrichment comparison between high MaxVCP scored variants (MaxVCP>0.5) in primary CoS versus secondary CoS, including annotations from enhancer, promoter, coding, and repressed categories. **d**. An example case of xQTL-only colocalization in gene *ARSB* showing two CoS, one colocalized among all brain cell types and the other colocalized across microglia, astrocytes and oligodendrocytes only. Error bars denote 95% confidence intervals. Numerical results are reported in **Supplementary Data**.

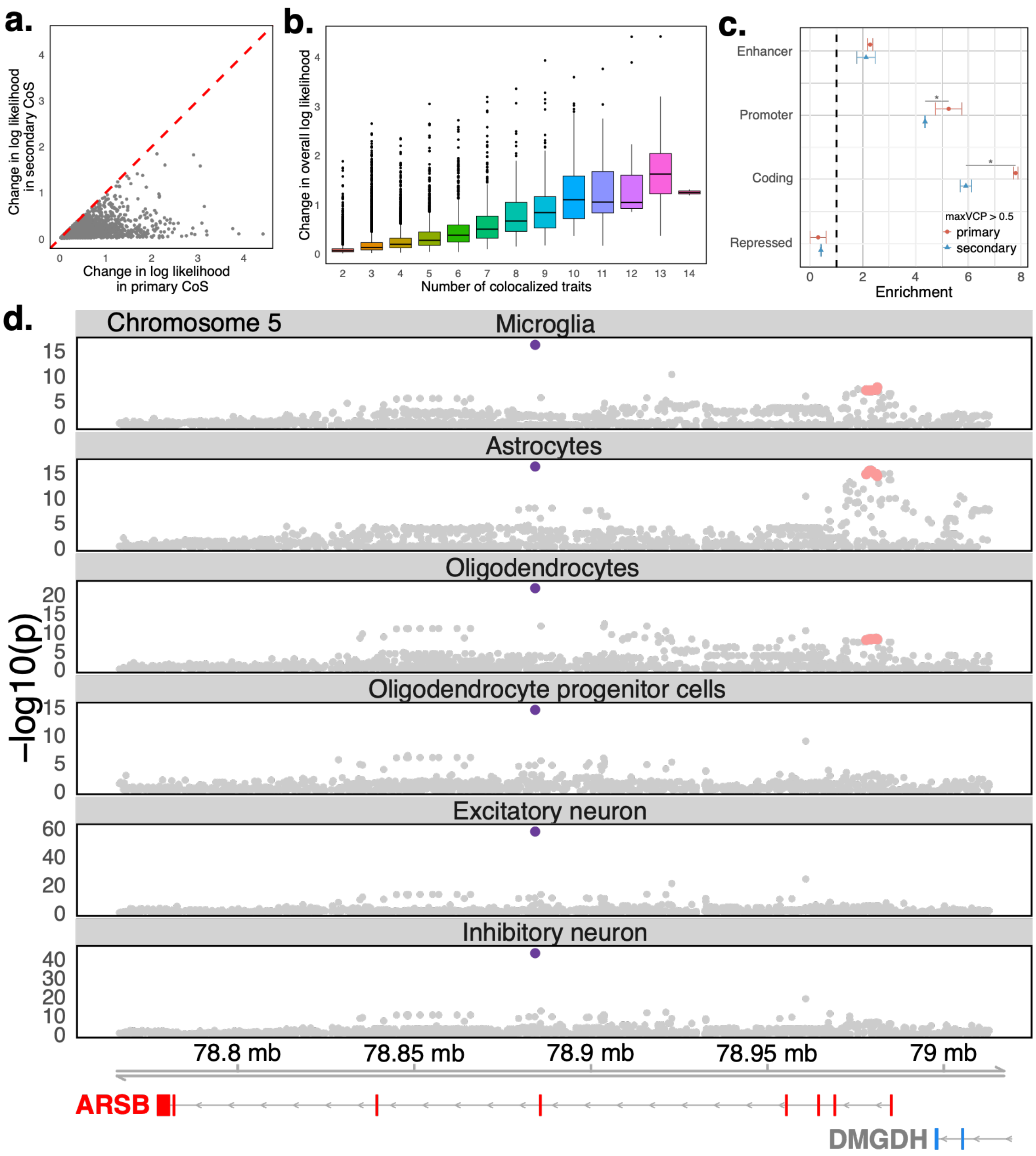

**Figure S10. xQTL-only *ColocBoost* for eQTLs across 13 brain tissues in GTEx. a.** UpSet plot summarizing the top 50 colocalized events in xQTL-only *ColocBoost* analysis across 13 brain tissues in GTEx. **b.** CoS-level overlap across ROSMAP xQTL colocalization events, ROSMAP eQTL colocalization events, and GTEx colocalization events involving brain cortex (3 tissues). **c.** Properties of genes with multiple causal variants, including log likelihood changes between primary and secondary CoS, the relationship between the number of colocalized traits and log likelihood changes, and the number of pairs of primary and secondary CoS across brain tissue eQTLs. Error bars denote 95% confidence intervals. Numerical results are reported in **Supplementary Data**.

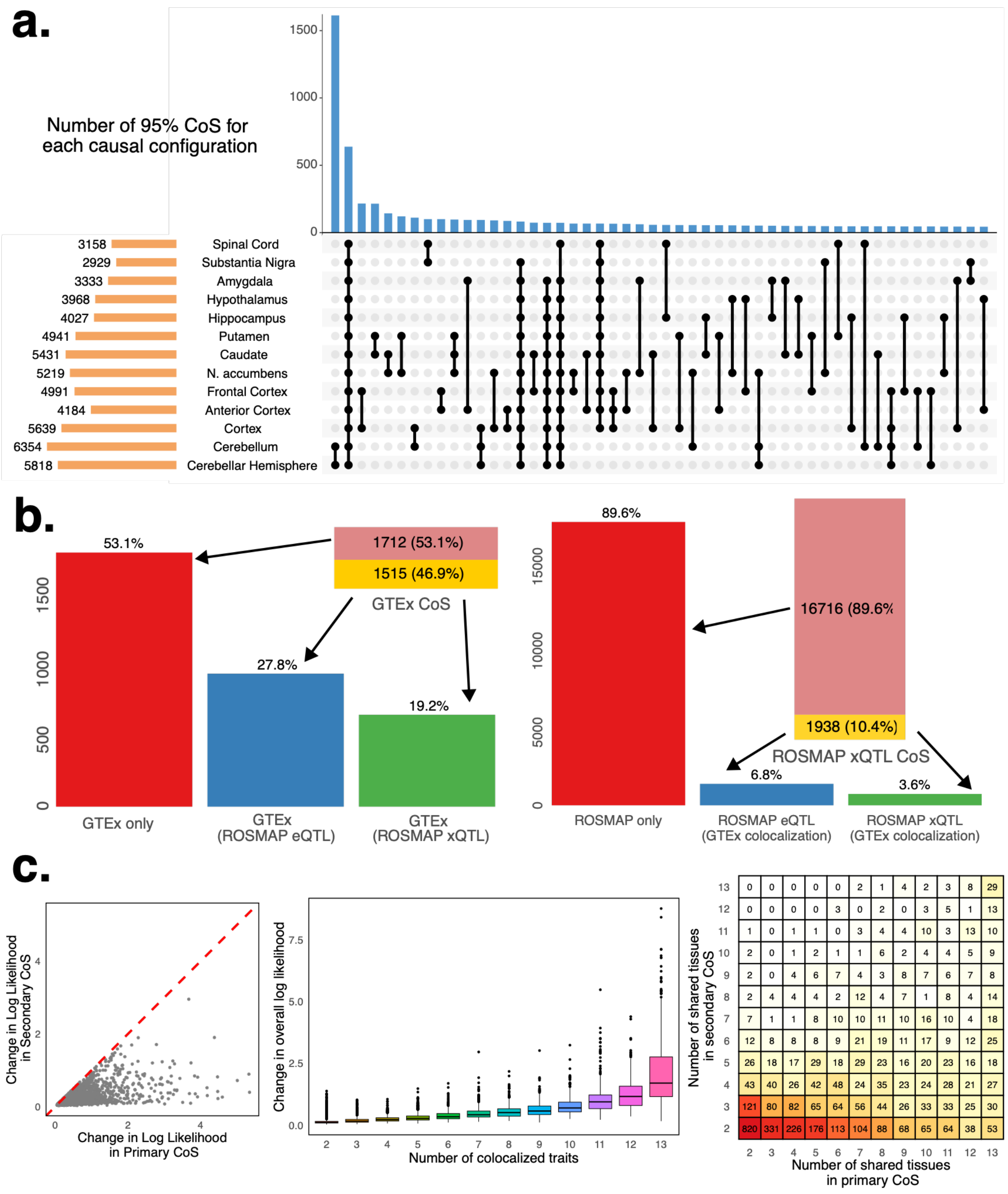

**Figure S11. Validation of *ColocBoost* colocalization. a.** Variant set-level excess-of-overlap (EOO) analysis of (i) 95% CoS-gene links from xQTL-only *ColocBoost*, (ii) standard marginal xQTL-gene associations, merged across all 17 xQTL datasets and (iii) 95% credible set (CS)-gene links from SuSiE fine-mapping, merged across all 17 xQTL datasets, against 118,389 regulatory element-gene maps from Promoter Capture Hi-C assay in the dorsolateral prefrontal cortex brain tissue, which profiles targeted 3D contacts linking gene promoters to proximal or distal regulatory elements. (**b, c**) EOO analysis of 95% CoS-gene links across (b) 3 different brain cortical regions and CD14+/CD16- bulk monocytes, and (c) 6 different brain cell types, against regulatory element-gene maps from PCHi-C data. **e**. Colocalization for *SUOX* in detail, also validated by CRIPRi in K562 cells. Error bars denote 95% confidence intervals. Numerical results are reported in **Supplementary Data**.

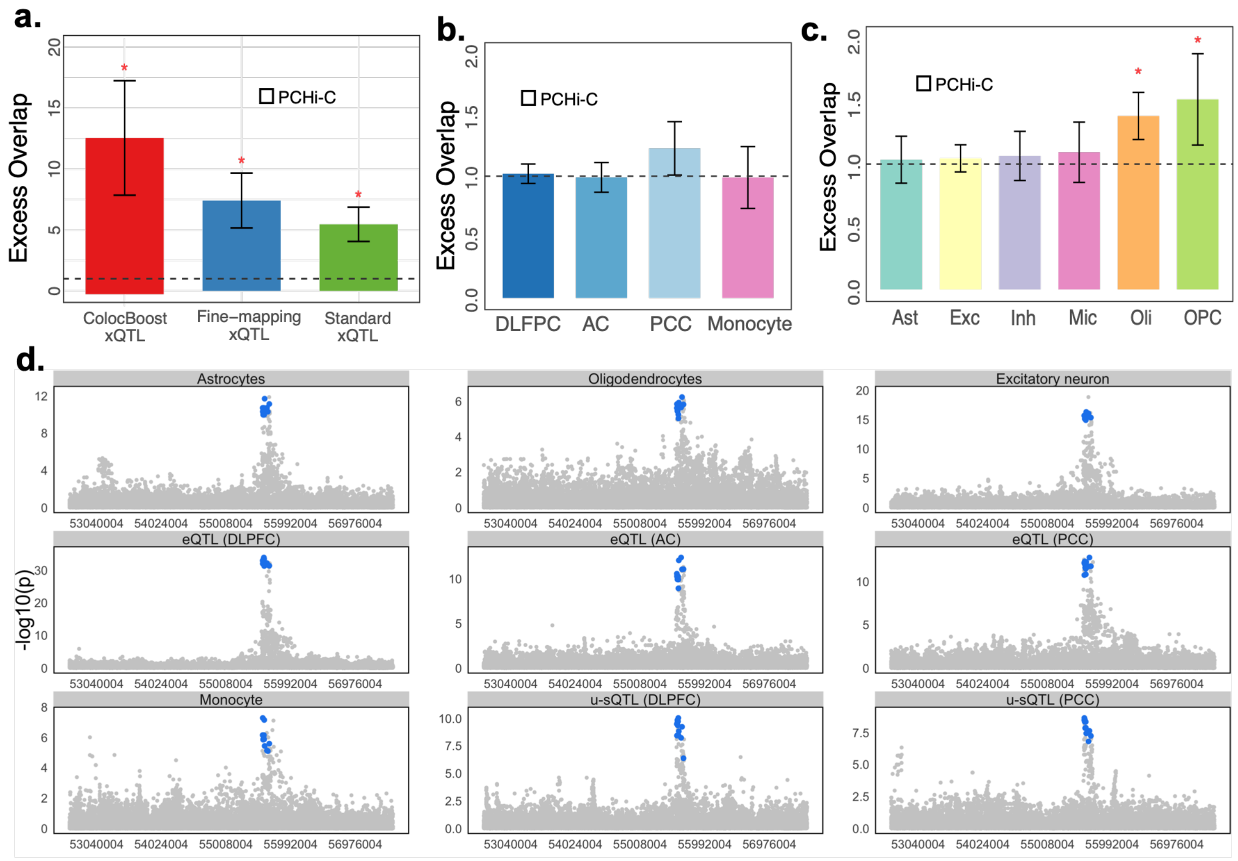

**Figure S12. Extended disease heritability analyses of variant-level functional annotations derived from *ColocBoost*. a.** Correlation matrix of five functional annotations based on the MaxVCP scores from *ColocBoost*. **b.** Heritability enrichment and standardized effect sizes, meta-analyzed across all, brain-related and blood-related traits, conditional on 97 baseline-LD v2.2 annotations of the 5 versions of xQTL MaxPPH derived from HyPrColoc. **c.** Standardized effect sizes, conditional on the 97 baseline-LD v2.2 annotations + MaxVCP-xQTL and CoS-xQTL annotations (joint $\tau^*$). **d.** Standardized effect sizes, conditional on the 97 baseline-LD v2.2 annotations + MaxVCP-xQTL and binarized MaxVCP-xQTL scores at different binarization thresholds (joint $\tau^*$). **e.** Heritability enrichment and standardized effect sizes (marginal $\tau^*$ and joint $\tau^*$) of MaxVCP-xQTL against MaxPIP annotation from SuSiE fine-mapping analysis. **f.** EOO analysis of MaxVCP-xQTL in the disease fine-mapped variants (PIP>0.95) from 94 UK Biobank traits and 930 Million Veteran Program (MVP) traits. **g.** Heritability enrichment and standardized effect sizes (marginal $\tau^*$ and joint $\tau^*$) of MaxVCP-xQTL annotations for cell-type-specific variants and cell-type-shared variants. Error bars denote 95% confidence intervals. Numerical results are reported in **Supplementary Data**. Asterisk denotes FDR<5% is computed by Bonferroni correction of the p-values for the number of annotations tested in the S-LDSC analysis.

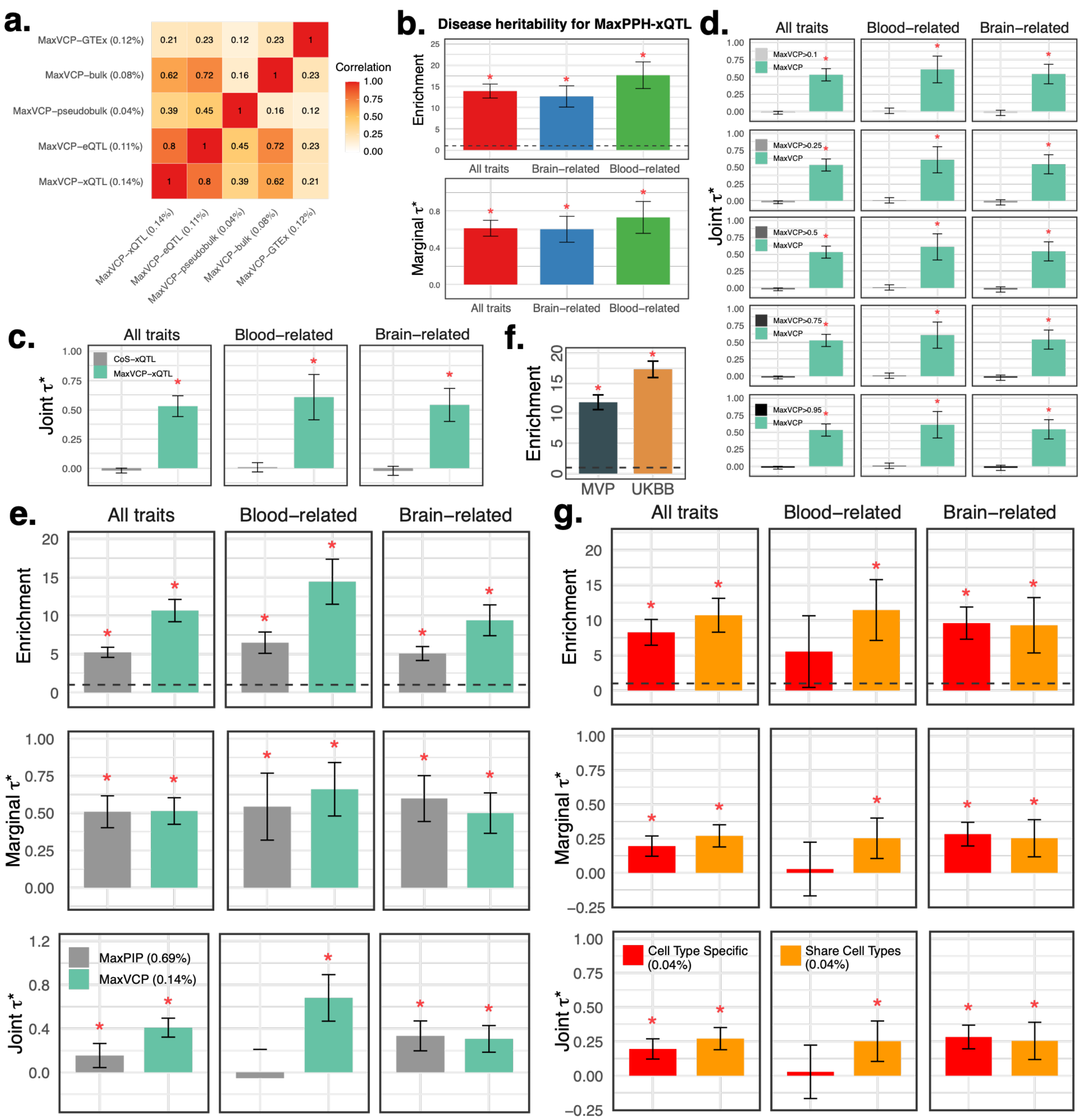

a.
MaxVCP−GTEx (0.12%)
MaxVCP−bulk (0.08%)
MaxVCP−pseudobulk (0.04%)
MaxVCP−eQTL (0.11%)
MaxVCP−xQTL (0.14%)
Correlation
b.
Disease heritability for MaxPPH-xQTL
Enrichment
Marginal τ*
All traits
Brain−related
Blood−related
c.
Joint τ*
CoS−xQTL
MaxVCP−xQTL
d.
MaxVCP>0.1
MaxVCP>0.25
MaxVCP>0.5
MaxVCP>0.75
MaxVCP>0.95
MaxVCP
e.
MaxPIP (0.69%)
MaxVCP (0.14%)
f.
MVP
UKBB
g.
Cell Type Specific (0.04%)
Share Cell Types (0.04%)

**Figure S13. Extended evaluations of GWAS-xQTL colocalization from *ColocBoost* and competing methods. a.** Precision-recall analysis comparing CoS-gene links from AD-xQTL *ColocBoost*, COLOC-Union, pairwise-*ColocBoost*-union, and a version of AD-xQTL *ColocBoost* limited to AD fine-mapped variants (*ColocBoost*-finemapped-GWAS) against enhancer-gene links predicted by ENCODE-rE2G across 354 biosamples. Error bars along both axis indicate 95% confidence intervals. **b**. Precision-Recall for the comparison with enhancer-gene links for each eQTL. COLOC-union only identified colocalization events for a few xQTL, and is excluded from the figure due to a lack of reliable standard error estimation. Manhattan plot of the MaxVCP functional annotation scores of variants from **c.** Pairwise-*ColocBoost*-union and **d.** COLOC-union, with labeled genes containing variants with MaxVCP>0.5. Microglia contributions are highlighted in green. Error bars denote 95% confidence intervals. Numerical results are reported in **Supplementary Data**.

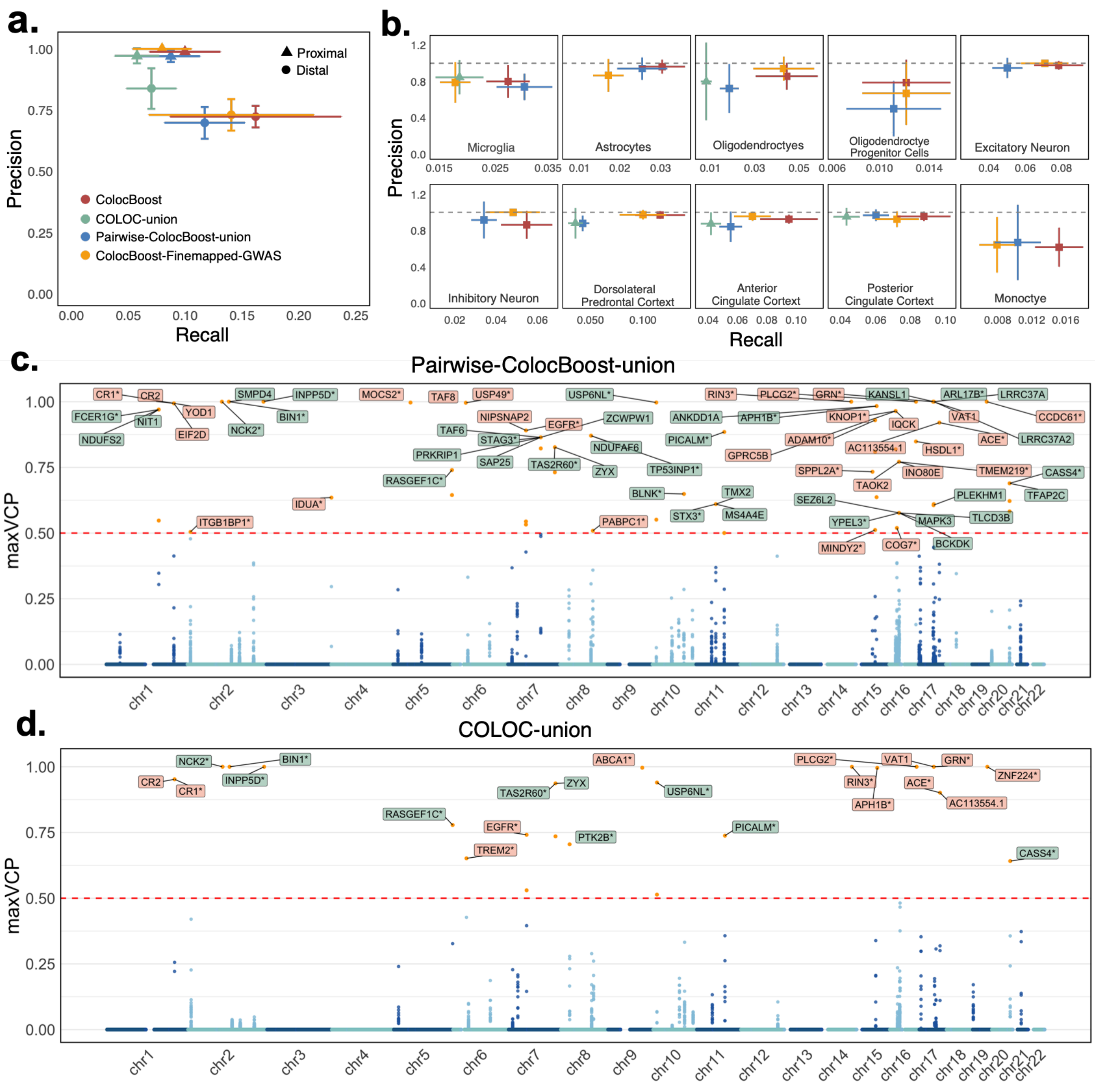

**Figure S14. Additional summary of genome-wide GWAS-xQTL colocalization results. a.** UpSet plot of colocalization events and identified by AD-xQTL *ColocBoost*. **b.** Manhattan plot of MaxVCP scores, with labeled genes containing variants with MaxVCP>0.1, and microglia contributions are highlighted in green. **c.** Excess overlap of genes showing colocalization in microglia with three microglia specific gene-sets as benchmarks. Error bars denote 95% confidence intervals. Numerical results are reported in **Supplementary Data**.

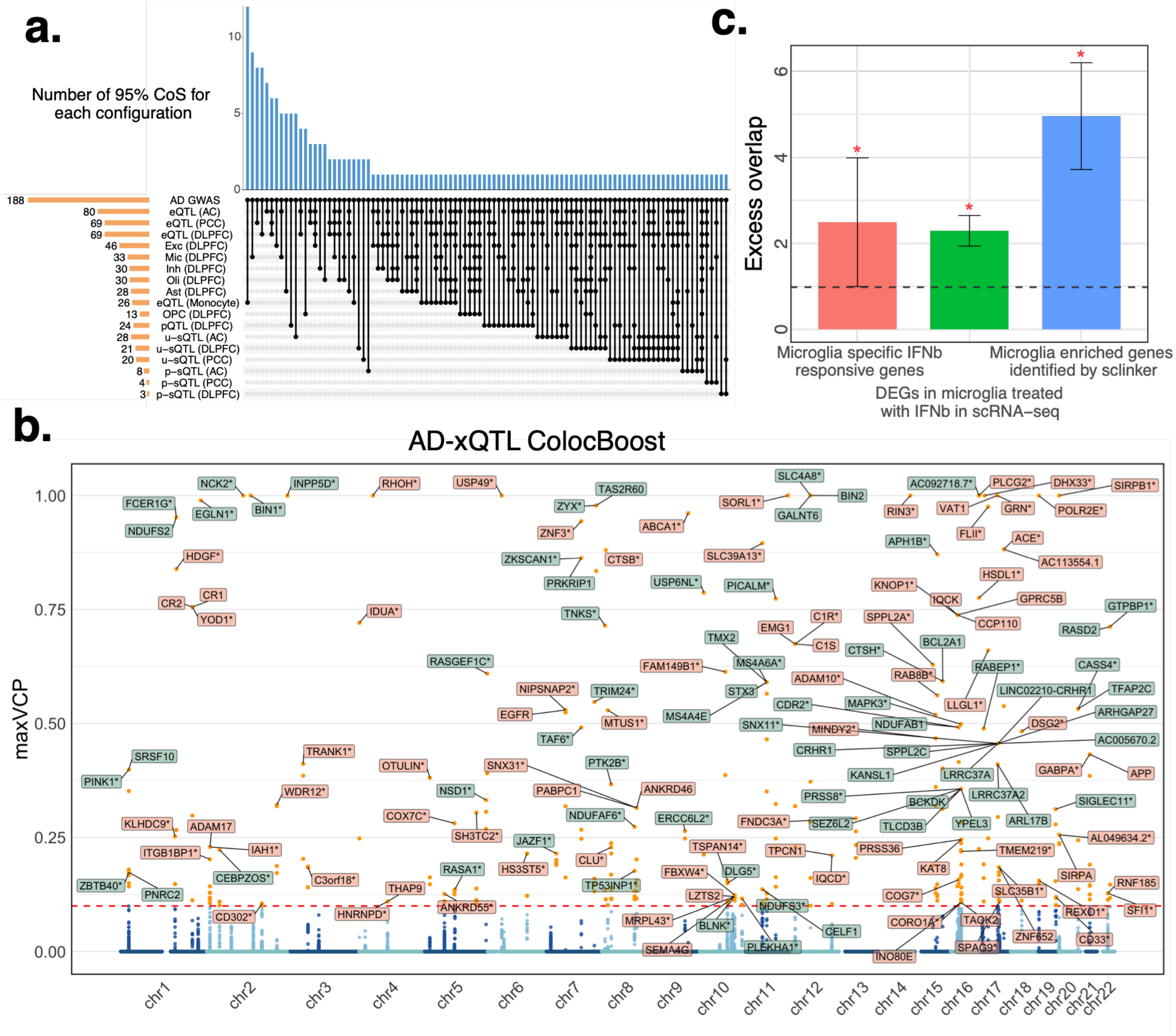

**Figure S15. Example cases of xQTL colocalizations in known AD risk genes. a.** Example of established AD gene *BIN1* across multiple molecular contexts and **b**. example of colocalized variants spanning multiple genes *CR1, CR2, YOD1*. One variant, rs669515 (MaxVCP=0.755), colocalized with *CR2* in DLPFC bulk eQTLs, *YOD1* in oligodendrocytes, and *CR1* across oligodendrocytes and all three additional bulk cortical eQTLs. Numerical results are reported in **Supplementary Data**.

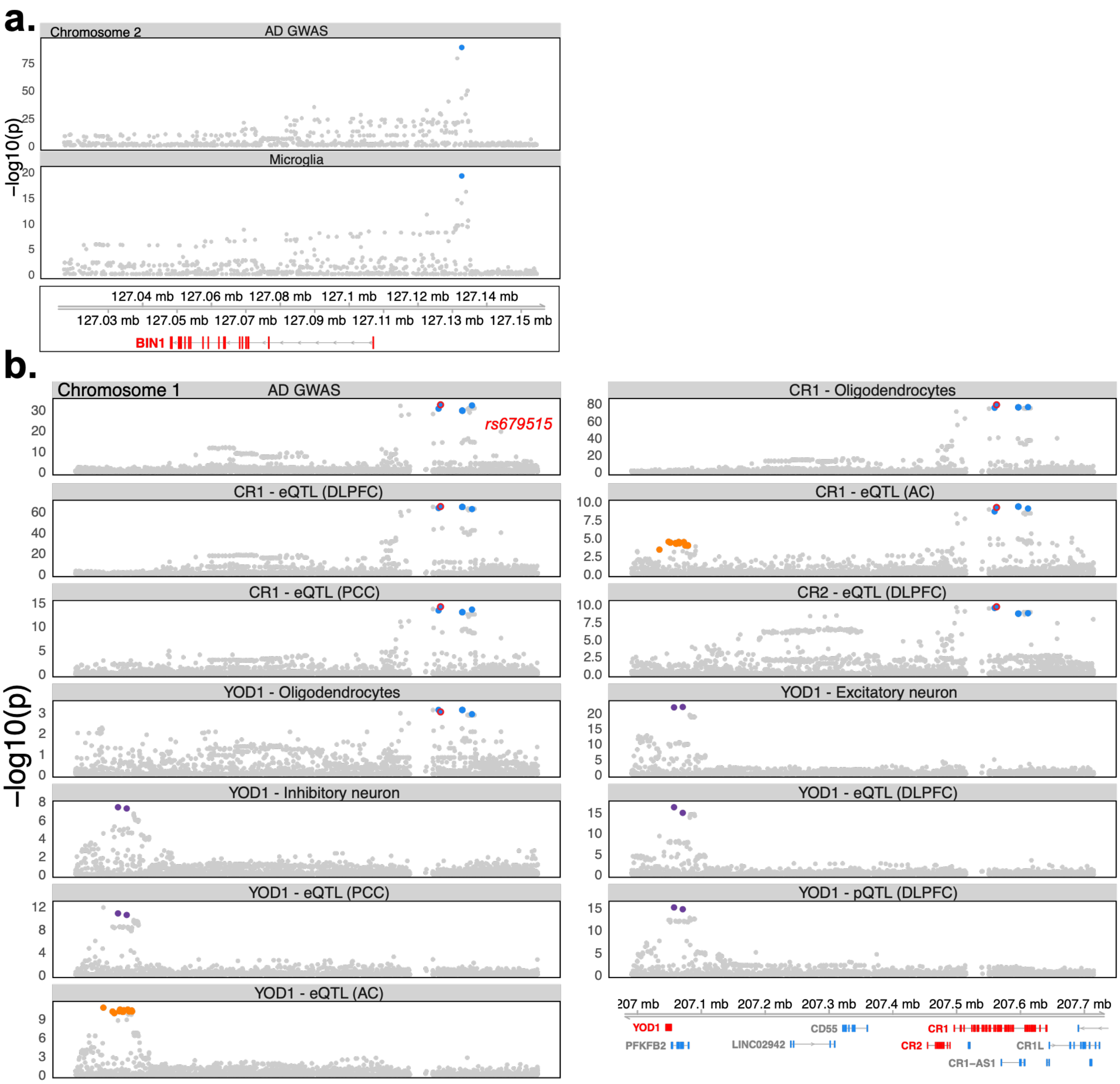

**Figure S16. Additional details of colocalizations in gene *CTSH*. a.** Marginal association p-values (at -log10 scale) and **b.** estimated multi-trait coefficients by *ColocBoost*, including all molecular events colocalized. Numerical results are reported in **Supplementary Data**.

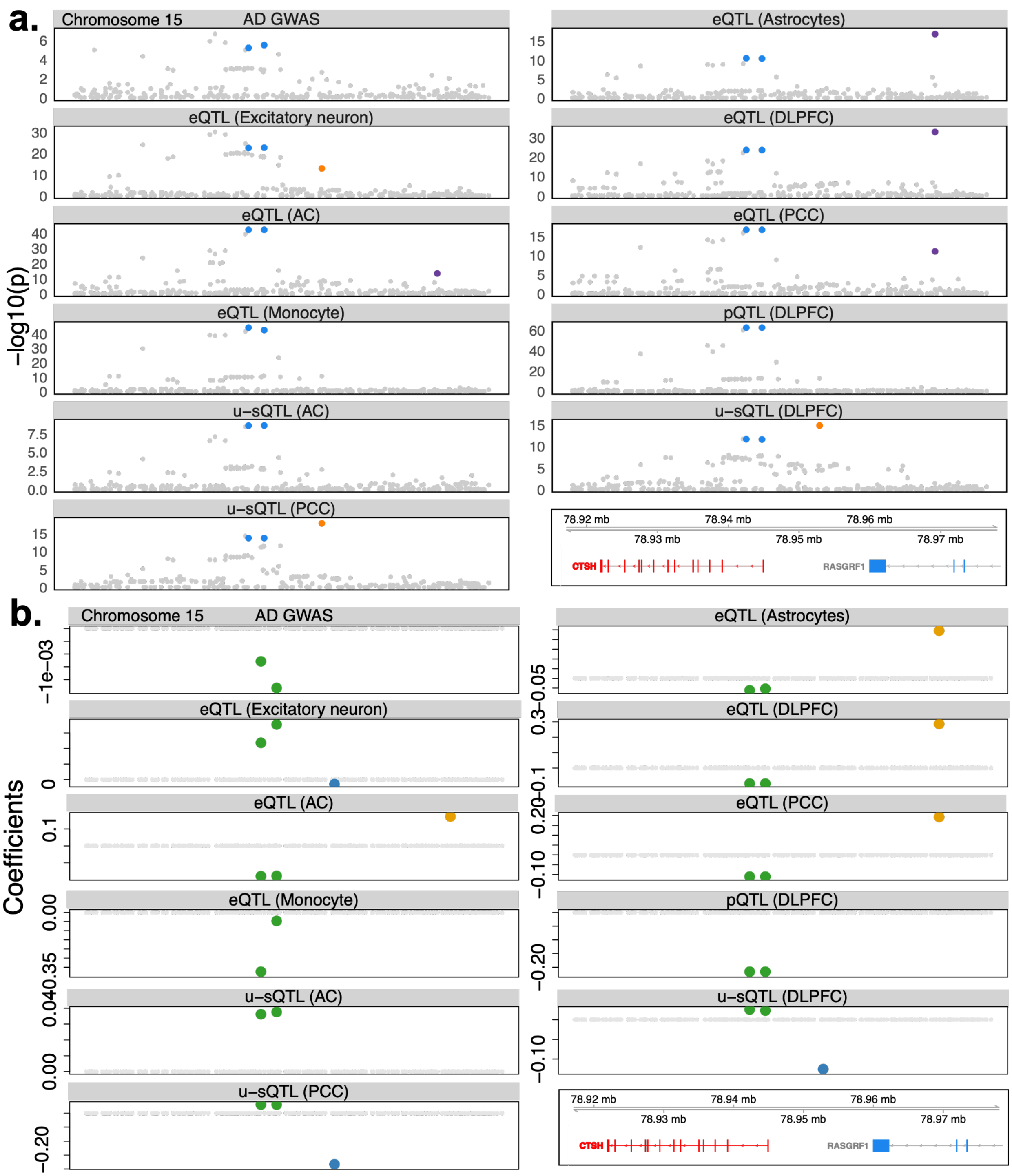

**Figure S17. Calibration of residual correlation across phenotypes for colocalization analysis. a.** Performance of *ColocBoost* under residual correlations $\rho \in \{0.3, 0.5, 0.7, 0.9\}$ based on an AR(1) model, reporting power, FDR, median CoS size, and mean set purity across simulations with 2, 5, and 10 traits and varying numbers of causal variants per trait. The corresponding independent-sample baseline ($\rho = 0$) is reported in Figure 2 and Figures S2. **b.** Estimated residual correlation matrix across ten eQTL datasets from the ROSMAP cohort, representing cell-type-specific and bulk-tissue contexts with 100% sample overlap.

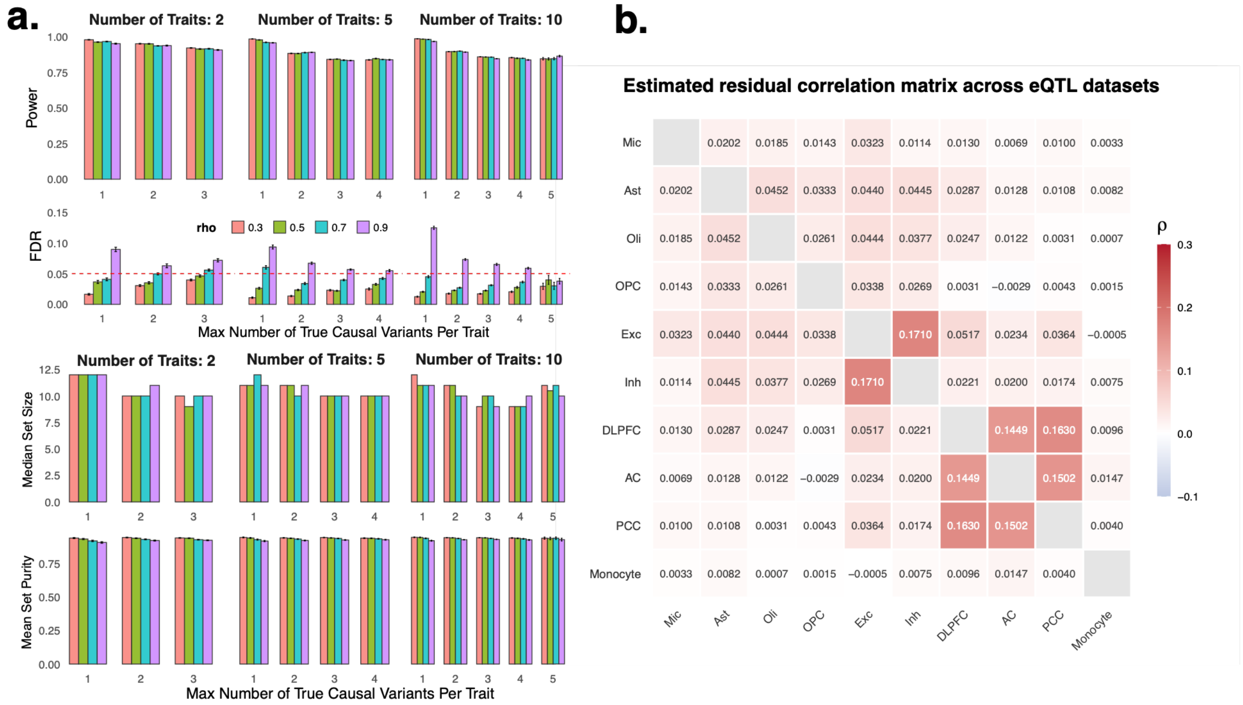

**Figure S18. Impact of LD reference mismatch on summary statistics based inference.** Data were simulated using GTEx genotypes ($N = 838$), whereas LD was estimated from independent European-ancestry reference panels from either the Alzheimer's Disease Sequencing Project (ADSP; $N = 19{,}292$) or the 1000 Genomes Project ($N = 489$). We randomly selected 500 TAD regions from a total of 1,361 TADs spanning the entire genome. SuSiE exhibited a higher false discovery rate (FDR) than *FineBoost* (the single-trait version of *ColocBoost*) when the smaller 1000 Genomes reference panel was used. The larger ADSP reference panel improved FDR control, consistent with reduced LD reference mismatch, as widely accepted in the literature.

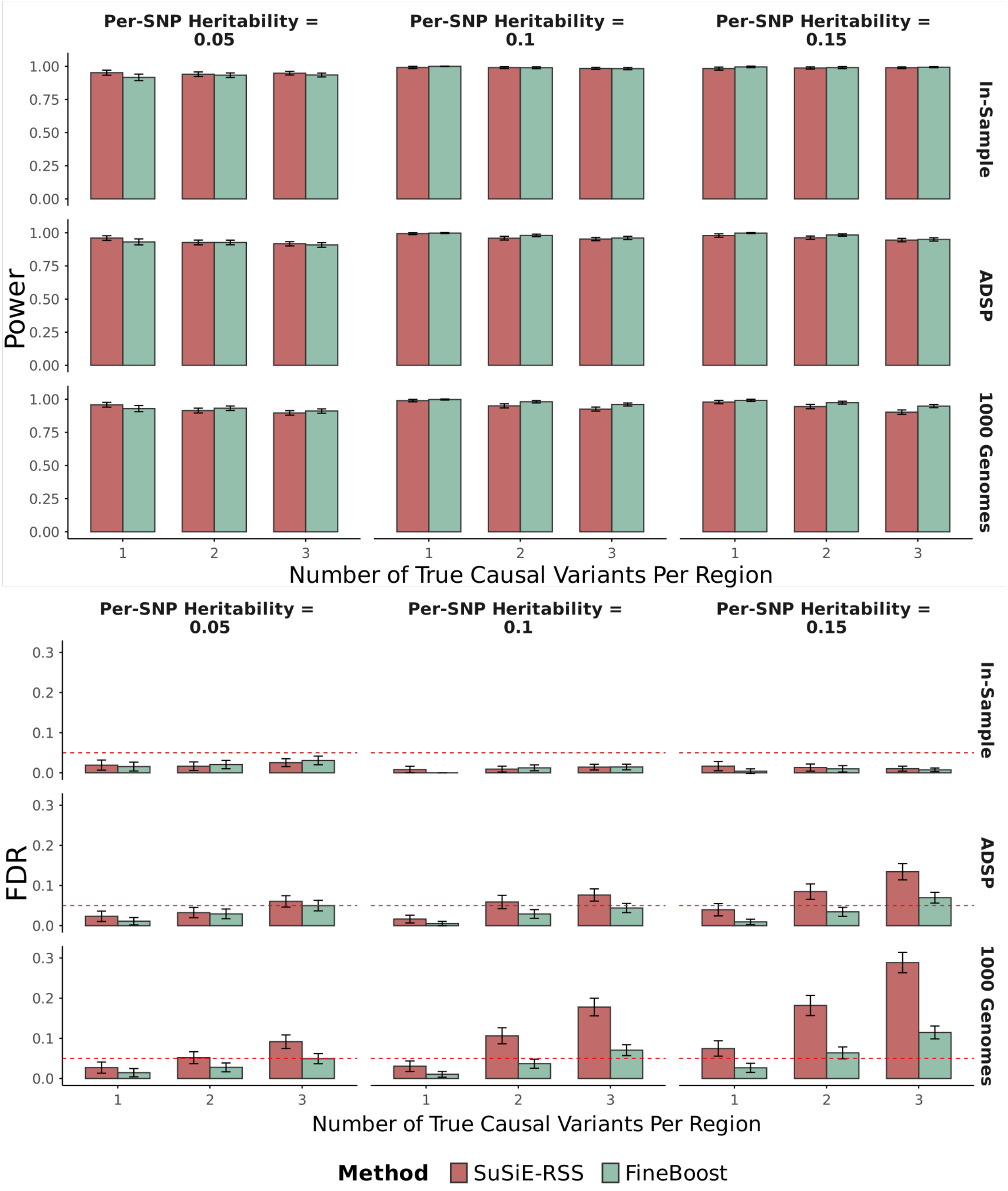
