## Supplementary Note for "Integrative multi-omics QTL colocalization maps regulatory architecture in aging human brain"

### S.1 *FineBoost*: Proximity smoothing gradient boosting algorithm in single-trait fine-mapping analysis

In statistical fine-mapping in genetic association studies, we start with a vector of phenotype, $\boldsymbol{Y}_{N\times 1}$, where $N$ is the number of samples. $Y_i$ can represent a continuous trait of individual $i$, such as height and BMI; or, represent a molecular trait, such as the gene expression of a specific gene observed in sample $i$. Fine-mapping is used to analyze the joint association of $P$ genetic variants to identify putative causal genetic variants (variants with non-zero effects) among highly correlated variants due to linkage disequilibrium (LD)[1]. Let $\mathbf{X}_{N\times P}$ be the matrix of genotypes, coded as $0, 1, 2$ for each of the $P$ genetic variants in a region of interest. Without loss of generosity we assume centered phenotype vector $\boldsymbol{Y}$, that is, $\bar{Y} = \sum_{i=1}^{N} Y_i = 0$ (in practice we can subtract $\bar{Y}$ from the original $\boldsymbol{Y}$ to center the phenotype vector). Then, for a quantitative trait $\boldsymbol{Y}$ following a normal distribution, relationship between $\boldsymbol{Y}$ and $\mathbf{X}$ can be modeled as the following multiple linear regression model:

$$\boldsymbol{Y}_{N\times 1} = \mathbf{X}_{N\times P}\boldsymbol{\beta}_{P\times 1} + \boldsymbol{\epsilon}_{N\times 1} \text{ with } \boldsymbol{\epsilon} \sim N(\mathbf{0}, \sigma^2\mathbf{I}_N), \tag{S.1.1}$$

where $\boldsymbol{\beta}$ is the fixed effect sizes of $P$ genetic variants on $\boldsymbol{Y}$, $\sigma^2$ is the variance of the phenotype with $\mathbf{I}_N$ an identity matrix with the diagonal elements of 1s. We refer to variables $j$ with non-zero effects ($\beta_j \neq 0$) as *effect variables*. The distribution of $i$-th sample phenotype can be written in terms of parameters $\boldsymbol{\beta}$ and data $(\boldsymbol{X}_i, Y_i)$,

$$Pr(Y_i|\boldsymbol{X}_i, \boldsymbol{\beta}) = \frac{1}{\sigma\sqrt{2\pi}} \text{exp}\left\{-\frac{1}{2}\left(\frac{Y_i - \boldsymbol{X}_i^T\boldsymbol{\beta}}{\sigma}\right)^2\right\}. \tag{S.1.2}$$

Therefore, the joint distribution of independent $Y_1, Y_2, \cdots, Y_N$ is

$$\begin{aligned} P(\boldsymbol{Y}|\mathbf{X}, \boldsymbol{\beta}) &= \prod_{i=1}^{N} \frac{1}{\sigma\sqrt{2\pi}} \text{exp}\left\{-\frac{1}{2}\left(\frac{Y_i - \boldsymbol{X}_i^T\boldsymbol{\beta}}{\sigma}\right)^2\right\} \\ &= (2\pi\sigma^2)^{\frac{N}{2}} \text{exp}\left\{-\frac{1}{2\sigma^2}\sum_{i=1}^{N}(Y_i - \boldsymbol{X}_i^T\boldsymbol{\beta})^2\right\}. \end{aligned} \tag{S.1.3}$$

To find the maximum likelihood estimate of $\boldsymbol{\beta}$, we use the negative log-likelihood as the loss function,

$$\begin{aligned} \mathcal{L}(\boldsymbol{\beta}) &= -\text{log}(P(\boldsymbol{Y}|\mathbf{X}, \boldsymbol{\beta})) \\ &= -\frac{N}{2}\text{log}(2\pi\sigma^2) + \frac{1}{2\sigma^2}\sum_{i=1}^{N}(Y_i - \boldsymbol{X}_i^T\boldsymbol{\beta})^2 \\ &:= \frac{1}{2}||\boldsymbol{Y} - \mathbf{X}\boldsymbol{\beta}||_2^2, \end{aligned} \tag{S.1.4}$$

where $||\cdot||_2^2$ is the squared $\ell_2$ norm of a vector, that is, $||\boldsymbol{Y} - \mathbf{X}\boldsymbol{\beta}||_2^2 = \sum_{i=1}^{N}(Y_i - \boldsymbol{X}_i^T\boldsymbol{\beta})^2$.

#### S.1.1 Overview of the standard gradient boosting algorithm

For the loss function of the multiple linear regression given in Equation (S.1.4), its gradient with respect to $\boldsymbol{\beta}$ is $\nabla\mathcal{L}(\boldsymbol{\beta}) := \nabla_{\boldsymbol{\beta}}\mathcal{L}(\boldsymbol{\beta}) = \mathbf{X}^T(\boldsymbol{Y} - \mathbf{X}\boldsymbol{\beta})$. We obtain the maximum likelihood estimate of $\boldsymbol{\beta}$ by solving $\nabla\mathcal{L}(\boldsymbol{\beta}) = 0$. Since $\mathbf{X}$ is not of full column rank, there is no unique solution. A more flexible way is to solve the following minimization problem

$$\boldsymbol{\beta}^\star := \arg\min_{\boldsymbol{\beta}} ||\nabla\mathcal{L}(\boldsymbol{\beta})||_\infty, \tag{S.1.5}$$

where $||\nabla\mathcal{L}(\boldsymbol{\beta})||_\infty = \max\left\{\left|\boldsymbol{X}_j^T(\boldsymbol{Y} - \mathbf{X}\boldsymbol{\beta})\right|; \forall j = 1, 2, \cdots, P\right\}$ is the $\ell_\infty$ norm. This renders a sub-gradient optimization problem as the optimizing function is a piece-wise linear function of $\boldsymbol{\beta}$. Typically such setup is likely non-smooth and non-differentiable at the transition points from one linear function to another[2]; yet this function is convex as it is point-wise maximum of convex (here affine) functions. To optimize, we can compute the sub-gradient and then use gradient descent on $\boldsymbol{\beta}$. Specifically, we write the above Equation (S.1.5) in the following form

$$\boldsymbol{\beta}^\star := \arg\min_{\boldsymbol{\beta}} \{g(\boldsymbol{\beta})\} \tag{S.1.6}$$

where $g(\boldsymbol{\beta})$ is defined as $g(\boldsymbol{\beta}) := \max\left\{|\boldsymbol{X}_j^T(\boldsymbol{Y} - \mathbf{X}\boldsymbol{\beta})|; \forall j = 1, 2, \cdots, P\right\}$. This can also be re-written as a function of the residuals $\boldsymbol{r} = \boldsymbol{Y} - \mathbf{X}\boldsymbol{\beta}$,

$$g(\boldsymbol{r}) := \max\left\{|\mathbf{X}_j^T\boldsymbol{r}|; \forall j = 1, 2, \cdots, P\right\}. \tag{S.1.7}$$

Therefore, maximizing $g(\boldsymbol{\beta})$ with respect to effects $\boldsymbol{\beta}$ is equivalent to maximize $g(\boldsymbol{r})$ with respect to residuals $\boldsymbol{r}$. A standard gradient descent algorithm on $\boldsymbol{r}$ can be used to fit the model by maximizing $g(\boldsymbol{r})$. Suppose at boosting round $k$, we have an update of $\boldsymbol{r}$ denoted as $\boldsymbol{r}^k$. Then, the subgradient direction is determined by the *best single effect* variant,

$$j_k^\star := \arg\max_j |\boldsymbol{X}_j^T\boldsymbol{r}^k|. \tag{S.1.8}$$

The gradient descent updates at boosting round $k$ through a weak boosting learner is given by

$$\boldsymbol{r}^{k+1} = \boldsymbol{r}^k - \eta_k \cdot \nabla g(\boldsymbol{r}^k) = \boldsymbol{r}^k - \eta_k \cdot \text{sgn}(\boldsymbol{X}_{j_k^\star}^T\boldsymbol{r}^k)\boldsymbol{X}_{j_k^\star}, \tag{S.1.9}$$

where $\nabla g(\boldsymbol{r}^k)$ is the gradient of $g(\boldsymbol{r})$ with respect to $\boldsymbol{r}^k$. Note that $\boldsymbol{r}^{k+1} - \boldsymbol{r}^k = \boldsymbol{Y} - \mathbf{X}\boldsymbol{\beta}^{k+1} - \boldsymbol{Y} + \mathbf{X}\boldsymbol{\beta}^k = -\mathbf{X}(\boldsymbol{\beta}^{k+1} - \boldsymbol{\beta}^k)$, so we can update effect size $\boldsymbol{\beta}$ by

$$\beta_{j_k^\star}^{k+1} = \beta_{j_k^\star}^k + \eta_k \cdot \text{sgn}(\boldsymbol{X}_{j_k^\star}^T\boldsymbol{r}^k) \text{ and } \beta_j^{k+1} = \beta_j^k, \forall j \neq j_k^\star. \tag{S.1.10}$$

Here, we employed a dynamic learning rate $\eta_k > 0$, which adaptively adjusts the optimization trajectory throughout the iterative gradient boosting process[3]. Specifically, we define $\eta_k = \eta_0/(1 + kd)$ as a decaying learning rate, where $\eta_0$ (default to 0.5) represents the initial learning rate and $d$ (default to 1.0) controls the decay rate of $\eta_k$ over iterations, effectively determining how far the algorithm moves along the negative gradient direction during each boosting round. Without a proper learning rate, gradient descent may fail to converge or require excessive computational resources. To balance the trade-off between convergence speed and stability at the further boosting rounds, we also implement a lower bound of 0.01, such that $\eta_k = \max(\eta_0/(1+kd), 0.01)$ (see details in Supplementary Note S.4.5).

Equation (S.1.10) gives the effect size $\boldsymbol{\beta}$ update at each boosting round. But in practice, updating the effect only at the best single-effect variant $j_k^\star$ identified by optimizing the objective may be suboptimal in the presence of multiple highly correlated variants—i.e., variants in high LD—as the selected variant might not correspond to the one with non-zero effect (hereafter this is used interchangeably with the word "causal"). A similar limitation arises in the widely accepted forward stepwise (FS) procedures for conditional analysis for association studies, and is typically mitigated by Bayesian inferences to account for LD in variable selection. For example SuSiE addresses this using an iterative Bayesian stepwise selection (IBSS), where per-variant contribution to a single-effect regression is assessed and averaged to compute posterior inclusion probabilities (PIPs) to reflect uncertainty due to LD[1]. In our framework, we extend the FS-style approach with proximity smoothing and gradient-weighted updates to diffuse signal across LD-linked variants, achieving a similar effect to IBSS with slightly lower computational cost since unlike SuSiE, we employ simpler (and weaker) updates without estimating hyperparameters and computing Bayes factors over all variants at each iteration, as will be elaborated in the next section.

#### S.1.2 *FineBoost* algorithm: A proximity smoothing gradient boosting algorithm for fine-mapping analysis

The standard gradient boosting selects the single variant by maximizing the trait-specific gradient contribution $j_k^\star = \arg\max_j |\boldsymbol{X}_j^T \boldsymbol{r}^k|$ at iteration $k$. The algorithm then updates only the effect size of this single variant through a weak learner with a dynamic learning rate. This hard-selection rule makes the update sensitive to small differences among high LD variants. For example, when two variant $j$ and $j'$ are in high LD, $|\boldsymbol{X}_j^T \boldsymbol{r}^k| \approx |\boldsymbol{X}_{j'}^T \boldsymbol{r}^k|$ and select one candidate variant to update over other high-LD neighbors can lead to (i) selecting wrong proxy tagging variants and missing out on the true causal variant, and (ii) high sensitivity of the update variants under small fluctuations of the data. We assess its calibration against simulated causal status and separately compare these probabilities with the corresponding single-effect probabilities from SuSiE.

To account for LD-induced uncertainty in variable selection, *FineBoost* replaces the hard update at the best variant $j_k^\star$ with a proximity-smoothed update over candidate variants through a *Proximity Smoothing Gradient Boosting* (PGSB) algorithm. The resulting *Single Effect Learner* (SEL) assigns mass using the current gradient evidence and an LD-informed positive reference measure. It does not claim to identify one variant uniquely when the data cannot distinguish variants in strong LD. Specifically, we define

$$(\hat{\omega}_j^k, \boldsymbol{r}^{k+1}, \beta_j^{k+1}) \leftarrow SEL(j_k^\star | \mathbf{X}, \boldsymbol{r}^k), \ \forall j = 1, 2, \cdots, P, \qquad \text{(S.1.11)}$$

that applies this smoothing constraint during each boosting round. The objective function shown in Equation (S.1.7) can be re-written at boosting round $k$ as:

$$g_{smooth}(\boldsymbol{r}^k) = \max_{\boldsymbol{\omega}^k} \sum_j \omega_j^k |\boldsymbol{X}_j^T \boldsymbol{r}^k|/(N-1) - \tau \cdot \rho(\boldsymbol{\omega}^k) \text{ with } \sum_{j=1}^P \omega_j^k = 1,\ \omega_j^k \geq 0, \tag{S.1.12}$$

where $\rho(\boldsymbol{\omega}^k)$ is a Kullback-Leibler (KL) divergence-based regularizer between $\boldsymbol{\omega}^k$ and $\boldsymbol{\delta}^k$,

$$\rho(\boldsymbol{\omega}^k) = \sum_{j=1}^P \omega_j^k \log(\omega_j^k) - \sum_{j=1}^P \omega_j^k \log(\delta_j^k) := KL(\boldsymbol{\omega}^k||\boldsymbol{\delta}^k), \tag{S.1.13}$$

where $\boldsymbol{\delta}^k \in \mathbb{R}^P$ is a positive LD-informed reference measure defined below.

The first term of this regularizer, $\sum_{j=1}^P \omega_j^k \log(\omega_j^k)$, denotes the entropy of $\boldsymbol{\omega}^k$ which encourages the otherwise mass update at $j_k^\star$ to be spread more uniformly onto other variants. The second term, $\sum_{j=1}^P \omega_j^k \log(\delta_j^k)$, denotes the cross-entropy with a data-driven local association simplex $\boldsymbol{\delta}^k$ which guides the update to concentrate around LD proxies of $j_k^\star$. We chose $\delta_j^k \propto \exp\left(\sqrt{|LD_{j,j_k^\star}|} \times h(Z_{\boldsymbol{X}_j,\boldsymbol{r}^k})\right)$, where $LD_{j,j_k^\star}$ denotes LD between variants $j$ and $j_k^\star$, and $h(Z_{\boldsymbol{X}_j,\boldsymbol{r}^k}) = (1-\lambda) \times |Z_{\boldsymbol{X}_j,\boldsymbol{r}^k}| + \frac{\lambda}{2} \times Z^2_{\boldsymbol{X}_j,\boldsymbol{r}^k}$ is an elastic-net style function of z-score for testing the association between $\boldsymbol{X}_j$ and $\boldsymbol{r}^k$. This formulation ensures the simplex stay close to $j_k^\star$ while taking into consideration local LD structure. We set the default value of $\lambda$ to 0.5, based on extensive numerical studies (see details in Supplementary Note S.6.2).

Solving the optimization over $\boldsymbol{\omega}^k$ and then plug back the estimate $\hat{\boldsymbol{\omega}}^k$ into Equation (S.1.12) yields a close-form, smoothed solution of the sub-gradient optimization problem as follows (see Supplementary Note S.4.1 for derivation details),

$$g^\tau_{smooth}(\boldsymbol{r}^k) = \tau \log\left(\sum_{j=1}^P \delta_j^k \exp\left(\frac{|\boldsymbol{X}_j^T \boldsymbol{r}^k|/(N-1)}{\tau}\right)\right), \tag{S.1.14}$$

where $g^\tau_{smooth}$ remains convex and differentiable with respect to $\boldsymbol{r}^k$, as it is a log-sum-exp function of a linear term of $\boldsymbol{r}^k$, but it is also smooth and has a valid gradient instead of having a sub-gradient as in case of $g$ (Equation S.1.7). The temperature (smoothing) parameter $\tau$ controls the degree of smoothing over $g$, with smaller values concentrating the update closer around $j_k^\star$ (default to $\tau = 0.01$). Therefore, the gradient of $g^\tau_{smooth}(\boldsymbol{r}^k)$ with respect to $\boldsymbol{r}^k$ can be written as

$$\nabla g^\tau_{smooth}(\boldsymbol{r}^k) = \sum_{j=1}^P \hat{\omega}_j^k \text{sgn}(\boldsymbol{X}_j^T \boldsymbol{r}^k)\boldsymbol{X}_j \quad \text{with} \tag{S.1.15}$$

$$\hat{\omega}_j^k := \frac{\frac{1}{N-1} \cdot \delta_j^k \exp\left(\frac{|\boldsymbol{X}_j^T \boldsymbol{r}^k|/(N-1)}{\tau}\right)}{\sum_{j=1}^P \delta_j^k \exp\left(\frac{|\boldsymbol{X}_j^T \boldsymbol{r}^k|/(N-1)}{\tau}\right)}, \tag{S.1.16}$$

where $\hat{\boldsymbol{\omega}}^k$ analogous a single-effect conditional inclusion probability vector of variants at iteration $k$, with $\hat{\omega}_j^k = \mathbb{P}(\beta_j^k \neq 0|\boldsymbol{X}_j, \boldsymbol{r}^k)$ (see details in Supplementary Note S.4.1). It is a Gibbs distribution over variant identity, with $\boldsymbol{\delta}^k$ as its reference masure and the exponentiated gradient as its data-dependent term. It is structurally analogous to SuSiE's single effect posterior $\boldsymbol{\alpha}_l$, but it is induced by the Nesterov-smoothed variational problem rather than a Bayesian model. At boosting round $k$, $\hat{\omega}_j^k$ is the probability mass assigned to variant $j$ for the current single-effect update. It replaces the hard selection of $j_k^\star$ by distributing the update across candidate variants (Proposition S1, Supplementary Note S.3.2). The temperature $\tau$ controls its concentration and the approximation error (default $\tau = 0.01$). As $\tau \to 0^+$, $\hat{\boldsymbol{\omega}}^k$ concentrates on $j_k^\star$, which has the maximal gradient evidence[2]; as $\tau \to \infty$, it converges to $\boldsymbol{\delta}^k$. The details of $\tau$ and the regularization term are provided in Supplementary Note S.5.2.

Subsequent updates of residual and effects using a dynamic learning rate $\eta_k$ yields

$$\boldsymbol{r}^{k+1} = \boldsymbol{r}^k - \eta_k \nabla g^{\tau}_{smooth}(\boldsymbol{r}^k) = \boldsymbol{r}^k - \eta_k \sum_{j=1}^{P} \hat{\omega}_j^k \text{sgn}(\boldsymbol{X}_j^T \boldsymbol{r}^k)\boldsymbol{X}_j, \tag{S.1.17}$$

$$\beta_j^{k+1} = \beta_j^k + \eta_k \hat{\omega}_j^k \text{sgn}(\boldsymbol{X}_j^T \boldsymbol{r}^k), \tag{S.1.18}$$

with $\eta_k$ adaptively tuned at each boosting round at a sufficiently small value to ensure each update behave as a *weak learner*, making controlled, incremental updates to prevent overfitting while capable of capturing weak effects. The difference between Equation (S.1.10) and Equation (S.1.18) is that unlike the former, the latter updates all the indices of $\boldsymbol{\beta}$ but the updates are weighted by $\hat{\omega}_j^k$ at each boosting round. For most variants, the value of $\hat{\omega}_j^k$ will be small, but in local neighborhoods of high LD with $j_k^\star$, $\hat{\omega}_j^k$ are non-zero for most variants in that neighborhood in a data-driven fashion, instead of picking one of them as in Equation (S.1.10). Regularization in *FineBoost* occurs implicitly through the dynamic learning rate $\eta_k$ and the number of boosting rounds $K$.

The complete *FineBoost* algorithm is described in **Algorithm S1**. Here, $\Delta\mathcal{L}(\boldsymbol{\beta}^{k+1}, \boldsymbol{\beta}^k)$ is the relative change of loss function, where $\Delta\mathcal{L}(\boldsymbol{\beta}^{k+1}, \boldsymbol{\beta}^k) = [\mathcal{L}(\boldsymbol{\beta}^{k+1}) - \mathcal{L}(\boldsymbol{\beta}^k)]/\mathcal{L}(\boldsymbol{\beta}^k)$ with $\mathcal{L}(\boldsymbol{\beta}^k)$ shown in Equation (S.1.4). The convergence guarantee of the standard gradient boosting algorithm has already been discussed in Freund et al[2]. For **Algorithm S1**, we analogously provide convergence guarantees (see Supplementary Note S.3.3).

---

**Algorithm S1** PGSB algorithm for fine-mapping in one trait

---

**procedure** *FineBoost*($\mathbf{X}, \boldsymbol{Y}$)
- $X_{n,j} \to X_{n,j} - \bar{X}_j$ ▷ mean centering of **X** columns
- $X_{n,j} \to X_{n,j} / \sqrt{\sum_n X_{n,j}^2}$ ▷ ($\ell_2$-norm scaling of **X** columns)
- $\boldsymbol{r}^0 \leftarrow \boldsymbol{Y} - \bar{Y}$,
- $\beta_j^0 \leftarrow 0, \quad \forall j = 1, 2, \cdots, P$ ▷ (Initialize parameters as 0)
- **while** $k < M$ **do**
  - $j_k^\star \leftarrow \arg\max_j |\boldsymbol{X}_j^T \boldsymbol{r}^k|$
  - $LD_{j,j_k^\star} \leftarrow \text{cor}(\mathbf{X}, \boldsymbol{X}_{j_k^\star})$
  - $(\hat{\omega}_j^k, \boldsymbol{r}^{k+1}, \boldsymbol{\beta}^{k+1}) \leftarrow SEL(j_k^\star | \mathbf{X}, \boldsymbol{r}^k), \ \forall j = 1, 2, \cdots, P$
  - **break if** $|\Delta\mathcal{L}(\boldsymbol{\beta}^{k+1}, \boldsymbol{\beta}^k)| < t$ ▷ Stop criterion
- **end while**
- **return** $\boldsymbol{\beta}^K$ and $\boldsymbol{\xi}^k, \ \forall k = 1, \cdots, K$

**end procedure**

---

We additionally implemented a multiple-testing correction based metric—via q-value[4] or local false sign rate[5]—as an optional termination criterion: we terminate the algorithm if all adjusted variant-residual association p-values $p_j^k \geq p_{t_1}$ before the convergence criterion $|\Delta\mathcal{L}(\boldsymbol{\beta}^{k+1}, \boldsymbol{\beta}^k)| < t$ is met. The current default implementation sets $p_{t_1} = 1$ as the most conservative threshold to ensure comprehensive assessment of all variants without biased towards candidate loci due to stronger marginal associations.

#### S.1.3 $\alpha-$level confidence sets of *FineBoost*

Suppose that *FineBoost* converge after $K$ boosting rounds. Let $\boldsymbol{\mathcal{W}} = \{\xi_j^k\}$ be the collection of weight vectors, where $\xi_j^k$ is the weight of variant $j$ updated at boosting round $k$ for $j = 1, \cdots, P$ and $k = 1, \cdots, K$. As mentioned in Supplementary Note S.1.2, *FineBoost* updates for variant $j$ is weighted by $\xi_j^k$ in Equation S.1.16. By nature, different SEL may choose to update the same or nearby (LD-proximal) variants across multiple rounds for the same causal effect; for example, when variants $j_k^\star$ and $j_{k'}^\star$ between two rounds are close LD proxies, the weights $\xi_\cdot^k$ and $\xi_\cdot^{k'}$ will be similar. On the other hand, different SEL can also update independents variants for the distinct causal effects with different weights $\xi_\cdot^k$ and $\xi_\cdot^{k'}$. To assemble SEL across boosting rounds at convergence, we clustered $\{\xi_j^k\}$ for $k = 1, \cdots, K$ to identify truly distinct causal effects using a modularity-based hierarchical clustering approach (see Supplementary Note S.5.1)[6]. The clustering step separates repeated weak learner visits to one LD-defined signal from distinct effects. Within each retained cluster, the iteration-level vectors are aggregated using their profile log-likelihood improvements to obtain one trait-specific single-effect distribution.

Let $S$ be the number of clusters obtained from the hierarchical clustering. For example, $S = 1$ indicates that all boosting rounds only focus on the same variant or the set of variants with high LDs. Let $\mathcal{K}_s$ be the set of boosting rounds within $s$-th cluster for $s = 1, \cdots, S$. We then re-normalize the per-round weights $\xi_j^k$ by the associated profile log-likelihood change $\Delta\mathcal{L}(k + 1, k) := \mathcal{L}(\boldsymbol{\beta}^{k+1}) - \mathcal{L}(\boldsymbol{\beta}^k)$ into a single, *modified* weight for $\mathcal{K}_s$,

$$W_j^s = \frac{\sum_{k \in \mathcal{K}_s} |\Delta\mathcal{L}(k+1,k)| \times \xi_j^k}{\sum_{k \in \mathcal{K}_s} |\Delta\mathcal{L}(k+1,k)|}, \quad \forall s = 1, \cdots, S; j = 1, \cdots, P. \tag{S.1.19}$$

Similar to SuSiE[1], we construct $\alpha$-level confidence sets $CS_s$, saying $\alpha = 95\%$, from the modified weights $W_1^s, \cdots, W_P^s$,

$$CS_s(\alpha) = \left\{ j_1, \cdots, j_{p_0} : p_0 = \min\left( p : \textstyle\sum_{j=1}^p W_{(j)}^s \geq \alpha; \quad W_{(1)}^s \geq \cdots \geq W_{(P)}^s \right) \right\}. \tag{S.1.20}$$

Also similar to SuSiE, we discard the CS with *purity* less than 0.5, where we define purity as the minimum absolute correlation between all pairs of variants within the same CS. This is motivated by the fact that CS containing many uncorrelated variants are usually too large and are of essentially little inferential value even if they sum to 95% coverage. As weak learners, some SEL in the proximity smoothing gradient boosting algorithm may capture noise as putative signals, potentially introducing false positives. To mitigate this risk, we implement an additional filter to discard CS with minimal contribution to the profile log-likelihood ($\Delta\mathcal{L} < 0.025$ by default). This is analogous to the test on estimated prior variance of a single effect in SuSiE for removing potentially insignificant effects (details in Supplementary Note S.5.2).

Following the removal of impure (purity < 0.5) and spurious (minimal contribution to profile log-likelihood) signals, we obtain $\tilde{S} \leq S$ confidence sets and compute the variant-level probability of association (VPA) for *FineBoost*, similar to Bayesian PIP, as:

$$\text{VPA}_j := 1 - \prod_{s=1}^{\tilde{S}}(1 - W_j^s). \tag{S.1.21}$$

#### S.1.4 Calibration of singe-effect conditonal inclusion probability and variant-level probability of association

We evaluated calibration at two distinct levels of the *FineBoost* probability hierarchy. The first analysis concerns the single-effect conditional inclusion probability $\hat{\omega}^k$ produced at one boosting round, before information is aggregated across rounds or single-effect components. The second concerns the VPA obtained after the complete single-trait *FineBoost* procedure, including clustering of boosting rounds, profile log-likelihood-weighted aggregation, and removal of impure or weakly supported components as described above. We evaluated each quantity against simulated causal status. Comparisons with the corresponding quantities from SuSiE were treated separately as assessments of concordance rather than as substitutes for calibration against the simulation truth.

*Perfect-LD diagnostic.* To isolate the behavior of $\hat{\omega}^k$ under exact symmetry, we constructed a synthetic design with $P = 100$ variants and $N = 1{,}162$ samples. For each block size $p = 1, \ldots, 10$, $p$ standardized genotype columns were placed in perfect LD, so that any two columns in the block were equal up to sign, and the remaining $100 - p$ variants were independent of one another and of the block. Phenotypes were generated using the same per-variant PVE of 5% and Gaussian residual model used in the realistic-LD simulations. We considered two causal placements: the causal variant was either a member of the perfect-LD block or one of the independent variants. Each combination of block size and causal placement was evaluated over 1,000 independently generated replicates. For every replicate, we retained the complete iteration-level probability vector, the mass assigned to the designated causal variant, and the total mass assigned to the perfect-LD block. When the causal variant lies in the block, all block members have identical absolute gradient evidence and identical reference weights; the diagnostic therefore tests whether the implementation assigns them equal shares of the block's total probability mass. Each member has probability $1/p$ only when the block accounts for all of the probability mass; otherwise, each receives one $p$th of the mass assigned to the block. The independent-causal-variant setting serves as the corresponding control without LD-induced ambiguity. This experiment is a numerical diagnostic of the symmetry property established in Proposition S1 of Supplementary Note S.3.2, rather than a general calibration analysis.

*Calibration of the iteration-level probability.* We evaluated $\hat{\omega}^k$ before component aggregation by retaining the probability vector from one *FineBoost* iteration for each ROSMAP-based simulation. Candidate variants across the simulations were assigned to 20 equally spaced bins over $[0, 1]$ according to their reported $\hat{\omega}_j^k$. Within each non-empty bin, we calculated the mean reported probability and the observed proportion of variants that were simulated as causal, together with a 95% confidence interval for the observed proportion. Plotting the observed causal frequency against the mean reported probability assesses whether variants assigned a given amount of probability mass are causal at the corresponding frequency. For a matched method comparison, we fitted SuSiE to the same simulated data and compared $\hat{\boldsymbol{\omega}}^k$ with its corresponding single-effect probability vector $\boldsymbol{\alpha}$. Agreement between the two variant-level vectors was summarized using correlation and KL divergence. This comparison evaluates concordance with an established single-effect fine-mapping quantity; calibration itself is defined by comparison with simulated causal status.

*Sensitivity of the iteration-level probability.* We varied one parameter at a time while holding the simulated data, causal configuration, and all remaining analysis settings fixed. For the temperature analysis, we evaluated $\tau \in \{0.001, 0.01, 0.1, 1, 2, 10, 50, 100\}$ and compared each resulting probability vector with that obtained at the default $\tau = 0.01$. For the reference-measure analysis, we evaluated $\lambda \in \{0, 0.25, 0.5, 0.75, 1\}$ and compared each probability vector with that obtained at the default $\lambda = 0.5$, with $\tau$ fixed at its default. For both analyses, changes in the complete variant-level probability vector were assessed by correlation and KL divergence relative to the corresponding default analysis. The $\lambda$ analysis evaluates sensitivity within the specified association–LD reference-measure family; it does not compare alternative regularization functionals.

*Calibration of the full-pipeline VPA.* We next evaluated calibration after aggregation across the single-effect components of the complete *FineBoost* analysis. We simulated one to five approximately independent causal variants per region under the same ROSMAP-based design and ran *FineBoost* through its full fitting and post-processing procedure. We applied SuSiE to the same simulated replicates and used its PIP as the corresponding full-model variant-level quantity. We assessed VPA–PIP concordance separately for each number of simulated causal variants by comparing the two reported scores for the same variants. For truth-based calibration, variants were grouped according to their reported VPA or PIP, and the mean reported value in each bin was compared with the observed proportion of simulated causal variants, with 95% confidence intervals. Thus, the scatterplot comparison evaluates agreement between methods, whereas the binned analysis evaluates calibration against known causal status. These experiments directly assess the single-trait iteration-level probability and VPA. Multi-trait VCP additionally depends on cross-trait probability aggregation, and its performance is evaluated separately in the multi-trait simulation studies described later in this Supplementary Note.

#### S.1.5 *FineBoost* with summary association statistics

To implement the proposed PGSB algorithm used in *FineBoost* on the summary data for fine-mapping analysis, we first describe the *FineBoost* model using sufficient statistics, then provide two strategies to obtain sufficient statistics from summary data.

##### S.1.5.1 *FineBoost* with sufficient statistics

Let $\mathbf{X}^T\boldsymbol{Y}$, $\mathbf{X}^T\mathbf{X}$, and $\boldsymbol{Y}^T\boldsymbol{Y}$ be the sufficient statistics. The multiple linear regression model for *FineBoost* was introduced in Supplementary Note (S.1), $\boldsymbol{Y}_{N\times 1} = \mathbf{X}_{N\times P}\boldsymbol{\beta}_{P\times 1} + \boldsymbol{\epsilon}_{N\times 1}, \boldsymbol{\epsilon} \sim N(\mathbf{0}, \sigma^2\mathbf{I}_N)$, with the optimization problem given by $\mathcal{L}(\boldsymbol{\beta}) := \frac{1}{2}||\boldsymbol{Y} - \mathbf{X}\boldsymbol{\beta}||_2^2$. We can re-write this optimization problem based on sufficient statistics as

$$\begin{aligned}\mathcal{L}(\boldsymbol{\beta}) &= \frac{1}{2}(\boldsymbol{Y}-\mathbf{X}\boldsymbol{\beta})^T(\boldsymbol{Y}-\mathbf{X}\boldsymbol{\beta}) \\ &= \frac{1}{2}(\boldsymbol{Y}^T\boldsymbol{Y} - 2\boldsymbol{\beta}^T\mathbf{X}^T\boldsymbol{Y} + \boldsymbol{\beta}^T\mathbf{X}^T\mathbf{X}\boldsymbol{\beta})\end{aligned} \tag{S.1.22}$$

and the maximization yields the best single-effect variant $j_k^\star$

$$j_k^\star := \arg\max_j |\boldsymbol{X}_j^T\boldsymbol{r}^k| = \arg\max_j |\boldsymbol{X}_j^T\boldsymbol{Y} - \boldsymbol{X}_j^T\mathbf{X}\boldsymbol{\beta}^k|, \tag{S.1.23}$$

where $\boldsymbol{X}_j^T\boldsymbol{r}^k = \boldsymbol{X}_j^T\boldsymbol{Y} - \boldsymbol{X}_j^T\mathbf{X}\boldsymbol{\beta}^k$ can also be calculated by the sufficient statistics given $\boldsymbol{\beta}^k$. SEL under the sufficient statistics can be reparameterized as $(\xi_j^k, \boldsymbol{X}_j^T\boldsymbol{r}^{k+1}, \beta_j^{k+1}) \leftarrow SEL_{SS}(j_k^\star|\mathbf{X}^T\mathbf{X}, \boldsymbol{X}_j^T\boldsymbol{r}^k)$, $\forall j = 1, 2, \cdots, P$, where

$$\beta_j^{k+1} = \beta_j^k + \eta_k\xi_j(\boldsymbol{r}^k)\text{sgn}(\boldsymbol{X}_j^T\boldsymbol{r}^k), \tag{S.1.24}$$

$$\boldsymbol{X}_j^T\boldsymbol{r}^{k+1} = \boldsymbol{X}_j^T\boldsymbol{r}^k - \eta_k\boldsymbol{X}_j^T\nabla g_{smooth}^{\tau}(\boldsymbol{r}^k) = \boldsymbol{r}^k - \eta_k\boldsymbol{X}_j^T\sum_{j=1}^{P}\xi_j(\boldsymbol{r}^k)\text{sgn}(\boldsymbol{X}_j^T\boldsymbol{r}^k)\boldsymbol{X}_j, \tag{S.1.25}$$

with $\xi_j(\boldsymbol{r}^k)$ calculated from Equation (S.1.16). This yields the algorithm of *FineBoost* under the sufficient statistics, **Algorithm S2**.

---

**Algorithm S2** PGSB algorithm for one trait using sufficient statistics

---

**procedure** *FineBoost*-SS($\mathbf{X}^T\mathbf{X}, \mathbf{X}^T\boldsymbol{Y}, \boldsymbol{Y}^T\boldsymbol{Y}$)
  $\mathbf{X}^T\boldsymbol{r}^1 \leftarrow \mathbf{X}^T\boldsymbol{Y}$
  $\beta_j^1 \leftarrow 0\ \forall j = 1, 2, \cdots, P$   ▷ (Initialize parameters as 0)
  **while** $k < M$ **do**
    $j_k^\star \leftarrow \arg\max_j |\boldsymbol{X}_j^T\boldsymbol{r}^k|$
    $(\xi_j^k, \boldsymbol{X}_j^T\boldsymbol{r}^{k+1}, \beta_j^{k+1}) \leftarrow SEL_{SS}(j_k^\star|\mathbf{X}^T\mathbf{X}, \boldsymbol{X}_j^T\boldsymbol{r}^k),\ \forall j = 1, 2, \cdots, P$
    **break if** $|\Delta\mathcal{L}(\boldsymbol{\beta}^{k+1}, \boldsymbol{\beta}^k)| < t$   ▷ Stop criterion
  **end while**
  **return** $\boldsymbol{\beta}^K$ and $\boldsymbol{\xi}^k\ \forall k = 1, \cdots, K$
**end procedure**

---

The relative change of loss function, $\Delta\mathcal{L}(\boldsymbol{\beta}^{k+1}, \boldsymbol{\beta}^k)$, can be calculated based on the sufficient statistics. Given that $(\boldsymbol{Y}-\mathbf{X}\boldsymbol{\beta}^k)^T(\boldsymbol{Y}-\mathbf{X}\boldsymbol{\beta}^k) = \boldsymbol{Y}^T\boldsymbol{Y} - 2(\boldsymbol{\beta}^k)^T\mathbf{X}^T\boldsymbol{Y} + (\boldsymbol{\beta}^k)^T\mathbf{X}^T\mathbf{X}\boldsymbol{\beta}^k$, we have

$$\begin{aligned}\Delta\mathcal{L}(\boldsymbol{\beta}^{k+1}, \boldsymbol{\beta}^k) &= \frac{\left[(\boldsymbol{Y}-\mathbf{X}\boldsymbol{\beta}^{k+1})^T(\boldsymbol{Y}-\mathbf{X}\boldsymbol{\beta}^{k+1}) - (\boldsymbol{Y}-\mathbf{X}\boldsymbol{\beta}^k)^T(\boldsymbol{Y}-\mathbf{X}\boldsymbol{\beta}^k)\right]}{(\boldsymbol{Y}-\mathbf{X}\boldsymbol{\beta}^k)^T(\boldsymbol{Y}-\mathbf{X}\boldsymbol{\beta}^k)} \\ &= \frac{(\boldsymbol{\beta}^{k+1}-\boldsymbol{\beta}^k)^T\mathbf{X}^T\boldsymbol{Y} + (\boldsymbol{\beta}^{k+1})^T\mathbf{X}^T\mathbf{X}\boldsymbol{\beta}^{k+1} - (\boldsymbol{\beta}^k)^T\mathbf{X}^T\mathbf{X}\boldsymbol{\beta}^k}{\boldsymbol{Y}^T\boldsymbol{Y} - 2(\boldsymbol{\beta}^k)^T\mathbf{X}^T\boldsymbol{Y} + (\boldsymbol{\beta}^k)^T\mathbf{X}^T\mathbf{X}\boldsymbol{\beta}^k}.\end{aligned} \tag{S.1.26}$$

##### S.1.5.2 Approximate sufficient statistics from summary association statistics

For most studies, it is difficult to access the individual-level data ($\mathbf{X}$ and $\boldsymbol{Y}$). However, the sufficient statistics can be approximated from the types of summary data that are commonly available (detailed derivations in Supplementary Note S.4.3).

*Approximate $\boldsymbol{X}_j^T\boldsymbol{Y}$ from z-scores*. Many studies provide z-scores for testing the associations between each of genetic variants and the trait of interest. Let $\boldsymbol{z} = (z_1, \cdots, z_P)$ be the z-scores for $P$ genetic variants, where

$$\boldsymbol{X}_j^T\boldsymbol{Y} = \frac{(N-1)z_j}{\sqrt{N-2+z_j^2}}\sigma_j\sigma. \tag{S.1.27}$$

Here, $\sigma_j$ represents the standard deviation of $\boldsymbol{X}_j$. If the minor allele frequency (MAF) is provided, $\sigma_j \approx 2 \times \text{MAF}_j \times (1 - \text{MAF}_j)$; if MAF is not provided, we assume the genotype is standardized in the simple linear regression, and $\sigma_j = 1$. Since $\sigma$ is the standard deviation of $\boldsymbol{Y}$, $\boldsymbol{Y}^T\boldsymbol{Y} = (N-1)\sigma^2$. If $\sigma$ is not provided in the summary data, we assume the phenotype is standardized such that $\sigma = 1$, resulting in $\boldsymbol{Y}^T\boldsymbol{Y} = N - 1$.

If z-scores are not available in the summary data, we can use the other information to recover z-score. (i) Many studies provide $\hat{\boldsymbol{\beta}} = (\hat{\beta}_1, \cdots, \hat{\beta}_P)^T$ and $\hat{\boldsymbol{se}}(\hat{\boldsymbol{\beta}}) = (\hat{se}(\hat{\beta}_1), \cdots, \hat{se}(\hat{\beta}_P))^T$ containing the estimates of marginal association for each variant $j$ on the trait and the corresponding standard errors. Therefore, we approximate z-score as $z_j = \hat{\beta}_j/\hat{se}(\hat{\beta}_j)$. (ii) Many studies provide $\hat{\beta}_j$ and the p-value $p_j$ for testing the association between variant $j$ and trait. Then, z-score can be recovered by $z_j = \text{sgn}(\hat{\beta}_j)\mathcal{F}^{-1}_{\chi_1^2}(1 - p_i)$, where $\mathcal{F}^{-1}_{\chi_1^2}(\cdot)$ is the inverse $\chi^2$ distribution with 1 degree of freedom.

*Approximate $\boldsymbol{X}_j^T\boldsymbol{X}_j$ from LD matrix*. Given the in-sample LD matrix **LD**, we can estimate $\boldsymbol{X}_j^T\boldsymbol{X}_j$ as

$$\boldsymbol{X}_j^T\boldsymbol{X}_j = (N-1)\boldsymbol{\Sigma}_X \cdot \textbf{LD} \cdot \boldsymbol{\Sigma}_X, \tag{S.1.28}$$

where $\boldsymbol{\Sigma}_X = \text{diag}(\sigma_1, \cdots, \sigma_P)$ is a diagonal matrix of the standard errors of genotype. Often, the estimate $\boldsymbol{X}_j^T\boldsymbol{X}_j$ is approximated by an "out-of-sample" LD matrix, that is, the sample covariance matrix of the same $P$ genetic variants in a suitable reference panel, chosen to be genetically similar to the study population.

### S.2 *ColocBoost*: A multi-task regression for scalable multi-trait colocalization analysis

The PGSB algorithm of *FineBoost* using SEL for one trait can be extended to multiple-trait colocalization analysis. We propose *ColocBoost*, a multi-trait colocalization method, that (i) accommodates multiple variants in LD with shared causal genetic effects at a locus, (ii) detects colocalization events with relatively weak effects shared across traits, (iii) identifies as many as colocalized loci within a region of interest.

**Multi-trait colocalization problem and colocalization events.** The multi-trait colocalization problem concerns identifying shared causal variants across $L$ traits within a genomic region of interest. The result of a *ColocBoost* analysis at a locus is a set of *Colocalization Events*, where each event $\{s\}$ is summarized by the triplet $\{g(s), T(s), \text{CoS}_\alpha(s)\}$, where:

(i) $g(s)$ denotes a *Genomic Region* or locus underlying the *ColocBoost* analysis;

(ii) $T(s)$ is the *Causal Configuration* specifying which subset of the analyzed traits share the same causal variant;

(iii) $\text{CoS}_\alpha(s)$ is a *Colocalization Confidence Set* representing the smallest set of variants containing the causal variant with probability at least $\alpha$ (default, $95\%$).

The $\text{CoS}_\alpha$ plays the same role in multi-trait analysis that the credible set plays in single-trait fine-mapping: under realistic linkage disequilibrium (LD), the data cannot uniquely identify a single causal variant from its tightly linked proxies, but it can identify the set of variants among which the causal variant must lie. We therefore adopt $\text{CoS}_\alpha$ as the primary unit of colocalization inference, in contrast to existing approaches that report a regional colocalization probability paired with a single nominated variant, such as COLOC's PP.H4 with the top SNP or HyPrColoc's regional posterior probability with a candidate variant.

Consider a total of $L$ traits, *ColocBoost* formulates colocalization as a multi-task regression problem for $L$ traits with phenotype vectors $\boldsymbol{Y}_1 \in \mathbb{R}^{N_1\times 1}, \cdots, \boldsymbol{Y}_L \in \mathbb{R}^{N_L\times 1}$ and corresponding standardized genotype matrices $\mathbf{X}_1 \in \mathbb{R}^{N_1\times P_1}, \cdots, \mathbf{X}_L \in \mathbb{R}^{N_L\times P_L}$, where $N_l$ is the number of independent individuals and $P_l$ is the number of genetic variants for trait $l$. Distinct from multivariate regression based models, here we do not make restrictions of locus having the same variants for different traits in a region of interest. That is, there may exist $P_{l_1} \neq P_{l_2}$ for $l_1 \neq l_2$ (see details about handling paritally overlapping variants across traits in Supplementary Note S.2.5.2). Let $P = |\bigcup_{l=1}^L P_l|$ be the total number of genetic variants for $L$ traits. For each trait $l$, a multiple linear regression model for genetic association can be written as

$$\boldsymbol{Y}_l = \mathbf{X}_l\boldsymbol{\beta}_l + \boldsymbol{\epsilon}_l, \quad \boldsymbol{\epsilon}_l \sim N(\mathbf{0}, \sigma_l^2\mathbf{I}_{N_l}), \tag{S.2.1}$$

where $\sigma_l^2$ is the variance of the phenotype vector, $\mathbf{I}_N$ is an identity matrix, and $\boldsymbol{\beta}_l = (\beta_{l,1}, \cdots, \beta_{l,P})$ is the effect-size vector of $P$ genetic variants on $\boldsymbol{Y}_l$. A variable $j$ is considered *colocalized* if it has non-zero effects ($\beta_{j,l} \neq 0$) for two or more traits.

Based on the loss function for one trait shown in Equation (S.1.4), *ColocBoost* considers a joint loss function for $L$ traits,

$$\mathcal{L}(\boldsymbol{\beta}_1, \cdots, \boldsymbol{\beta}_L) := \sum_{l=1}^{L} \frac{1}{2}||\boldsymbol{Y}_l - \mathbf{X}_l\boldsymbol{\beta}_l||_2^2, \tag{S.2.2}$$

with the gradients with respect to $\boldsymbol{\beta}_l$

$$\nabla\mathcal{L}(\boldsymbol{\beta}_1, \cdots, \boldsymbol{\beta}_L) = \begin{pmatrix} \boldsymbol{X}_1^T(\boldsymbol{Y}_1 - \mathbf{X}_1\boldsymbol{\beta}_1) \\ \vdots \\ \boldsymbol{X}_L^T(\boldsymbol{Y}_L - \mathbf{X}_L\boldsymbol{\beta}_L) \end{pmatrix}_{L\times P} = \begin{pmatrix} \nabla\mathcal{L}(\boldsymbol{\beta}_1) \\ \vdots \\ \nabla\mathcal{L}(\boldsymbol{\beta}_L) \end{pmatrix}_{L\times P}. \tag{S.2.3}$$

*ColocBoost* seeks to minimize $\ell_{(\infty,1)}$ norm of $\nabla\mathcal{L}(\boldsymbol{\beta}_1, \cdots, \boldsymbol{\beta}_L)$ by first aggregating contribution across all traits at variant $j$ through an $\ell_1$-type sum, and then maximizing over $j$ for variant with largest cross-trait gradient through an $\ell_\infty$-type maximum,

$$||\nabla\mathcal{L}(\boldsymbol{\beta}_1, \cdots, \boldsymbol{\beta}_L)||_{(\infty,1)} = \max\left\{\sum_{l=1}^{L} |\boldsymbol{X}_{l,j}^T(\boldsymbol{Y}_l - \mathbf{X}_l\boldsymbol{\beta}_l)|;\ \forall j = 1, 2, \cdots, P\right\}. \tag{S.2.4}$$

In general, for multiple traits $L$, we define the best single-effect variant $j_k^\star$ at boosting round

$k$ for *joint updates* across traits as

$$j_k^\star := \arg\max_j \left( \sum_{l=1}^{L} |\boldsymbol{X}_{l,j}^T \left( \boldsymbol{Y}_l - \mathbf{X}_l \beta_l^k \right) | \right) = \arg\max_j \left( \sum_{l=1}^{L} |(\boldsymbol{X}_{l,j}^T \boldsymbol{r}_l^k| \right). \quad \text{(S.2.5)}$$

and the *trait-specific* best single-effect variant $j_k^l = \arg\max_j \left( |(\boldsymbol{X}_{l,j}^T \boldsymbol{r}_l^k| \right)$, where $\boldsymbol{r}_l^k = \boldsymbol{Y}_l - \mathbf{X}_l \beta_l^k$.

#### S.2.1 *ColocBoost* algorithm for pair-wise colocalization analysis using SEC

*ColocBoost* introduces multi-task weak learners called *Single-Effect Couplers* (SEC), which, at each iteration, models a single causal variant by first evaluating the coupling across traits, and then performing proximity smoothed update at the variant for each set of coupled traits.

We first introduce how *ColocBoost* works for two traits ($L = 2$) via SEC. At boosting round $k$, the best single-effect variant $j_k^\star$ for *joint update* across two traits and the *trait-specific* best single-effect variant $j_k^l$ are given by

$$j_k^\star = \arg\max_j \left( \sum_{l=1}^{2} |(\boldsymbol{X}_{l,j}^T \boldsymbol{r}_l^k| \right) \quad \text{and} \quad j_k^l = \arg\max_j \left( |(\boldsymbol{X}_{l,j}^T \boldsymbol{r}_l^k| \right) \text{ for } l = 1, 2. \quad \text{(S.2.6)}$$

We define the *equivalence* of $j_k^l \sim j_k^\star$ between the trait-specific best update variant $j_k^l$ and joint best update variant $j_k^\star$ if (i) $j_k^l = j_k^\star$ or in strong LD with each other ($|r| > 0.8$), and (ii) have comparable effects, i.e., $\Delta loglik(\cdot; \boldsymbol{Y}_l)$ for trait $l$ at two variants is smaller than $\varepsilon$ ($\varepsilon = 0.1$), where $\Delta loglik(\cdot; \boldsymbol{Y}_l)$ measures the drop in trait $l$'s profile log-likelihood if the effect at a given variant is removed. Therefore, determining which traits to update jointly simply involves checking which traits' best update variant is equivalent to $j_k^*$. To do so, we propose a *dynamic coupling strategy* as follows:

*Logic 1 (SEC)*. If $j_k^1 \sim j_k^\star$ and $j_k^2 \sim j_k^\star$, it indicates that $j_k^\star$ is a putative colocalized variant for two traits; so we *couple* the updates across both traits at variant $j_k^*$ with trait-specific proximity smoothing. That is,

$$(\xi_{jl}^k, \boldsymbol{r}_l^{k+1}, \beta_{jl}^{k+1}) \leftarrow SEC(j_k^\star | \mathbf{X}_l, \boldsymbol{r}_l^k), \quad \forall l = 1, 2, \quad \text{(S.2.7)}$$

where the detailed algorithm of above *proximity smoothed gradient boosting* update via SEC can be found in Supplementary Note S.1.2.

*Logic 2*. If $j_k^1 \nsim j_k^\star$ and $j_k^2 \nsim j_k^\star$, it implies that there are potentially stronger trait-specific effects for both traits to account for in the current boosting round, compared to potential shared signal; so we update each of trait 1 and trait 2 separately at $j_k^1$ and $j_k^2$, respectively. That is,

$$(\xi_{jl}^k, \boldsymbol{r}_l^{k+1}, \beta_{jl}^{k+1}) \leftarrow SEC(j_k^l | \mathbf{X}_l, \boldsymbol{r}_l^k), \quad \forall l = 1, 2. \quad \text{(S.2.8)}$$

*Logic 3 (Delayed SEC)*. If $j_k^1 \sim j_k^\star$ but $j_k^2 \nsim j_k^\star$ (or vice versa), we treat this signal as uncolocalized and compare $\Delta loglik(\cdot; \boldsymbol{Y}_l)$ to decide which trait (either trait 1 or trait 2) to update at *its own best* during the current boosting round, deferring any potentially coupled effects on $j_k^*$ to future rounds.

In *sub-logic 3.1*, if $\Delta loglik(j_k^\star; \boldsymbol{r}_1^k) > \Delta loglik(j_k^2; \boldsymbol{r}_2^k)$ holds, we further evaluate if $\Delta loglik(j_k^\star; \boldsymbol{r}_2^k)$ is substantial (above given threshold - a proportional threshold in terms of the maximal $\Delta loglik(\cdot; \boldsymbol{r}_l^k)$ for $l = 1, 2$; see details in Supplementary Note S.6.2). If so, we update trait 2 at $j_k^2$, that is,

$(\xi_{j2}^k, \boldsymbol{r}_2^{k+1}, \beta_{j2}^{k+1}) \leftarrow SEC(j_k^2|\mathbf{X}_2, \boldsymbol{r}_2^k)$, thus delaying the coupled update at $j_k^\star$. Otherwise, we update trait 1 at $j_k^1 \sim j_k^\star$ using $(\xi_{j1}^k, \boldsymbol{r}_1^{k+1}, \beta_{j1}^{k+1}) \leftarrow SEC(j_k^1|\mathbf{X}_1, \boldsymbol{r}_1^k)$.

In *sub-logic 3.2*, if $\Delta loglik(j_k^\star; \boldsymbol{r}_1^k) \leq \Delta loglik(j_k^2; \boldsymbol{r}_2^k)$, we skip any update at $j_k^\star$, instead update trait 2 at $j_k^2$. That is, $(\xi_{j2}^k, \boldsymbol{r}_2^{k+1}, \beta_{j2}^{k+1}) \leftarrow SEC(j_k^2|\mathbf{X}_2, \boldsymbol{r}_2^k)$.

The delay avoids classifying the joint candidate immediately when one trait still carries a stronger trait-specific signal elsewhere. Fitting that competing signal first allows genuine shared evidence to remain in the residuals and be coupled later, while apparent sharing induced by LD can weaken after the trait-specific effect is removed.

#### S.2.2 *ColocBoost* algorithm: Coupling updates across traits to identify shared causal effects in multi-trait colocalization analysis

For multiple traits $L > 2$, at boosting round $k$, the best single-effect variant $j_k^\star$ for *joint update* across $L$ traits and the *trait-specific* best single-effect variant $j_k^l$ are given by

$$j_k^\star = \arg\max_j \left(\sum_{l=1}^L |(\boldsymbol{X}_{l,j}^T \boldsymbol{r}_l^k|\right) \quad \text{and} \quad j_k^l = \arg\max_j \left(|(\boldsymbol{X}_{l,j}^T \boldsymbol{r}_l^k|\right) \text{ for } l = 1, \cdots, L. \qquad \text{(S.2.9)}$$

We check the equivalence of $j_k^l \sim j_k^\star$ for each trait $l$ as previously described. Let $\mathcal{R}_k^\star$ be a subset of traits with $j_k^l \sim j_k^\star$ and the complementary subset $\mathcal{R}_k^{\neg\star}$ with $j_k^l \nsim j_k^\star$ at boosting round $k$. We extend the *dynamic coupling strategy* with $L = 2$ to $L > 2$, as follows.

<u>*Logic 1 (SEC)*</u>. If $j_k^l \sim j_k^\star$ for all $l = 1, \cdots, L$ ($\mathcal{R}_k^{\neg\star} = \emptyset$), we *couple* all updates across traits at the same variant $j_k^\star$ with trait-specific proximity smoothing. That is,

$$(\xi_{jl}^k, \boldsymbol{r}_l^{k+1}, \beta_{jl}^{k+1}) \leftarrow SEC(j_k^\star|\mathbf{X}_l, \boldsymbol{r}_l^k), \quad \forall l = 1, \cdots, L. \qquad \text{(S.2.10)}$$

<u>*Logic 2*</u>. If $j_k^l \nsim j_k^\star$ for all $l = 1, \cdots, L$ ($\mathcal{R}_k^\star = \emptyset$), we consider the following two sub-logics to determine either trait-specific update or coupled effect update.

In *sub-logic 2.1*, if for any trait $l \in \mathcal{R}_k^{\neg\star}$ there exists at least another trait $l'$ such that $j_k^l \sim j_k^{l'}$, we partition traits into different equivalence groups and update *the trait group* $\mathcal{R}_k^{\star'}$ where maximum $\Delta loglik(j_k^l; \boldsymbol{r}_l^k)$ of $\mathcal{R}_k^{\star'}$ is the highest among all groups. To do so, we re-compute $j_k^{\star'}$ limited to $\mathcal{R}_k^{\star'}$, then implement *Logic 1* on $\mathcal{R}_k^{\star'}$ for the current boosting round. That is,

$$(\xi_{jl}^k, \boldsymbol{r}_l^{k+1}, \beta_{jl}^{k+1}) \leftarrow SEC(j_k^{\star'}|\mathbf{X}_l, \boldsymbol{r}_l^k), \quad \forall l \in \mathcal{R}_k^{\star'}. \qquad \text{(S.2.11)}$$

In *sub-logic 2.2*, if there exists a subset of uncoupled traits ($\mathcal{R}_k^{\neg\star'} \subset \mathcal{R}_k^{\neg\star}$), each having a unique best single-effect variant not equivalent to other traits ($j_k^l \nsim j_k^{l'}$ for any $l' = 1, \cdots, L$), we update each trait $l$ in $\mathcal{R}_k^{\neg\star'}$ at its own best $j_k^l$ and skip updates for other traits. That is,

$$(\xi_{jl}^k, \boldsymbol{r}_l^{k+1}, \beta_{jl}^{k+1}) \leftarrow SEC(j_k^l|\mathbf{X}_l, \boldsymbol{r}_l^k), \quad \forall l \in \mathcal{R}_k^{\neg\star'}. \qquad \text{(S.2.12)}$$

<u>*Logic 3 (Delayed SEC)*</u>. If $j_k^l \sim j_k^\star$ for a subset of traits $\mathcal{R}_k^\star$ and $j_k^l \nsim j_k^\star$ for the complementary subset $\mathcal{R}_k^{\neg\star}$ ($\mathcal{R}_k^\star \neq \emptyset$ and $\mathcal{R}_k^{\neg\star} \neq \emptyset$), it indicates we have both potential colocalization at $j_k^\star$ for $\mathcal{R}_k^\star$ and one or more trait-specific effects elsewhere for $\mathcal{R}_k^{\neg\star}$. To avoid premature coupled updates based only on $\mathcal{R}_k^\star$, we introduce a *delayed SEC* (D-SEC) approach to update $\mathcal{R}_k^{\neg\star}$, deferring updates at $j_k^\star$ to future boosting rounds. This delay accounts for the possibility that some traits in $\mathcal{R}_k^{\neg\star}$ do in fact share causal effect at $j_k^\star$ but currently fail the *equivalence* check because

their best single-effect update is another stronger, trait specific effect at a different variant. To implement D-SEC:

In *sub-logic 3.1*, if $\min_{l\in\mathcal{R}_k^\star}\{\Delta loglik(j_k^\star;\boldsymbol{r}_l^k)\} > \max_{l\in\mathcal{R}_k^{\neg\star}}\{\Delta loglik(j_k^l;\boldsymbol{r}_l^k)\}$, we identify $\mathcal{R}_k^{\neg\star'}$ ($\mathcal{R}_k^{\neg\star'}\subseteq\mathcal{R}_k^{\neg\star}$) whose $\Delta loglik$ at $j_k^\star$ is substantial (above given threshold), and implement *Logic 2* on $\mathcal{R}_k^{\neg\star'}$. If no such additional traits exist, we implement *Logic 1* on $\mathcal{R}_k^\star$ at variant $j_k^\star$.

In *sub-logic 3.2*, if $\min_{l\in\mathcal{R}_k^\star}\{\Delta loglik(j_k^\star;\boldsymbol{r}_l^k)\} < \max_{l\in\mathcal{R}_k^{\neg\star}}\{\Delta loglik(j_k^l;\boldsymbol{r}_l^k)\}$, we skip any updates at $j_k^\star$, instead implement *Logic 2* on $\mathcal{R}_k^{\neg\star}$.

Note that in *Logic 3*, there are other alternative metrics to compare $\Delta loglik(j_k^\star;\boldsymbol{r}_l^k)$ for $l\in\mathcal{R}^\star$ and $\Delta loglik(j_k^l;\boldsymbol{r}_l^k)$ for $l\in\mathcal{R}^{\neg\star}$. For example,

- *P.1 (median).* $\text{median}_{l\in\mathcal{R}_k^\star}\{\Delta loglik(j_k^\star;\boldsymbol{r}_l^k)\} > \text{median}_{l\in\mathcal{R}_k^{\neg\star}}\{\Delta loglik(j_k^l;\boldsymbol{r}_l^k)\}$.
- *P.2 (max-min).* $\max_{l\in\mathcal{R}_k^\star}\{\Delta loglik(j_k^\star;\boldsymbol{r}_l^k)\} > \min_{l\in\mathcal{R}_k^{\neg\star}}\{\Delta loglik(j_k^l;\boldsymbol{r}_l^k)\}$.
- *P.3 (max-max).* $\max_{l\in\mathcal{R}_k^\star}\{\Delta loglik(j_k^\star;\boldsymbol{r}_l^k)\} > \max_{l\in\mathcal{R}_k^{\neg\star}}\{\Delta loglik(j_k^l;\boldsymbol{r}_l^k)\}$.
- *P.4 (min-min).* $\min_{l\in\mathcal{R}_k^\star}\{\Delta loglik(j_k^\star;\boldsymbol{r}_l^k)\} > \min_{l\in\mathcal{R}_k^{\neg\star}}\{\Delta loglik(j_k^l;\boldsymbol{r}_l^k)\}$.

The reason we set *min-max* as the default situation is that we would like to delay the coupled update at $j_k^\star$ to as late as it is reasonable, ensuring all traits are affected by $j_k^\star$ can be coupled to update at later boosting rounds (see details in Supplementary Note S.6.2).

#### S.2.3 Expediting SEC learners in disease-prioritized *ColocBoost*

For disease-prioritized mode of *ColocBoost*, we introduce *expedited SEC* (E-SEC) to prioritize identifying colocalization with a focal trait of interest, e.g., a GWAS. Here, we start with the best single-effect variant for trait of interest $j_k^e = \arg\max_j(|\mathbf{X}_{e,j}^T\boldsymbol{r}_e^k|)$, and construct its equivalence group $\mathcal{R}_k^e$ with respect to $j_k^l = \arg\max_j(|\mathbf{X}_{l,j}^T\boldsymbol{r}_l^k|)$ for each trait $l\neq e$, that is, we evaluate if $j_k^e\sim j_k^l$. If $\mathcal{R}_k^e\neq\emptyset$, we re-compute $j_k^e = \arg\max_j\left(\sum_{l\in\mathcal{R}_k^e}|\mathbf{X}_{l,j}^T\boldsymbol{r}_l^k|\right)$ across traits in $\mathcal{R}_k^e$ as the candidate variant to update in current bosting round, where by design, $j_k^e\sim j_k^*$. We iterate this procedure until $\mathcal{R}_k^e=\emptyset$, then employ the dynamic coupling strategy previously described to guide subsequent boosting rounds. E-SEC effectively couples traits in $\mathcal{R}_k^e$ at variant $j_k^e$ during earlier boosting rounds. This mechanism enhances the identification of colocalizations with the focal trait.

#### S.2.4 Inference on colocalization events from *ColocBoost*

*ColocBoost* characterize a colocalization event $s$ using three attributes $\{CoS_s(\alpha),T(s),g(s)\}$, where $g(s)$ specifies the region where the colocalization event $s$ occurred, the trait configuration $T(s)$ can also be analogously represented as a configuration vector $\boldsymbol{T}^s=(T_1^s,\cdots,T_L^s)$, where $T_l^s\in\{0,1\}$ indicates whether the variant has a putative effect on trait $l$, and $CoS_s(\alpha)$ represents a single-effect colocalization confidence set (CoS) of coverage $\alpha$ (default 0.95). For any given variant, the total number of possible configurations across $L$ traits is $2^L-L-1$. For example, there are $2^3=8$ putative causal configurations for 3 traits of interest, such as $(0,0,0),(1,0,0),(0,1,0),(0,0,1),(1,1,0),(1,0,1),(0,1,1),(1,1,1)$. Here, there are $4=2^3-3-1$ colocalized causal configurations, where at least two traits share the same causal variant

$((1,1,0),(1,0,1),(0,1,1),(1,1,1))$. These attributes describe an event-level object. The confidence set records uncertainty about which variant drives one shared effect, whereas the trait configuration records which traits are assigned to that effect. Both should be kept distinct from VCP, which is a variant-level summary obtained after evidence is propagated across iterations, traits, and retained single-effect components.

##### S.2.4.1 Aggregation of PSGB iterations into colocalization events

*ColocBoost* provides variant-level probability measures that propagate uncertainty from individual boosting iterations through multi-trait aggregation to a final per-variant quantity. The probability hierarchy, $\hat{\omega}_{jl}^{k} \to \xi_{jl}^{s} \to \xi_{j}^{s} \to \text{VCP}_j$, comprises four levels: (1) the per-iteration variant probability $\widehat{\omega}_{jl}^{k}$, arising from the Nesterov-smoothed objective (Proposition S1; Supplementary Note S.3.2); (2) the trait-specific single-effect probability $\xi_{jl}^{s}$, obtained by averaging $\widehat{\omega}_{jl}^{k}$ across iterations within a given trait and single-effect component; (3) the single-effect colocalization probability $\xi_{j}^{s}$, obtained by combining the trait-specific probabilities $\xi_{jl}^{s}$ across the colocalizing traits in that component; and (4) the variant colocalization probability $\text{VCP}_j$, obtained by combining evidence across single-effect components under the same conditional-independence assumption used by SuSiE to calculate PIP[1]. The full derivation of this probability hierarchy is summarized below.

Upon convergence, *ColocBoost* identifies $S$ candidate colocalization events within genomic region $g$. Each event $s$ is summarized by the triplet $\{\text{CoS}_s(\alpha), T(s), g\}$, where $T(s)$ is its trait configuration and $\text{CoS}_s(\alpha)$ is its $\alpha$-level colocalization confidence set.

*Determination of* $T(s)$. The trait configuration is represented by the binary vector $\boldsymbol{T}^s = (T_1^s, \ldots, T_L^s)$, where $T_l^s \in \{0,1\}$ indicates whether trait $l$ participates in the putative shared effect. Excluding the null and $L$ single-trait configurations, there are $2^L - L - 1$ possible colocalization configurations. The configuration is determined by identifying traits that share evidence for genetic effects across SEC updates. For a configuration $\boldsymbol{T}$, let $\mathcal{K}(\boldsymbol{T})$ denote the corresponding boosting rounds. After clustering, each single-effect component $s$ inherits a configuration $\boldsymbol{T}^s$ and a component-specific subset $\mathcal{K}_s \subseteq \mathcal{K}(\boldsymbol{T}^s)$. We write $T(s) = \{l : T_l^s = 1\}$ for its colocalizing traits.

*Determination of* $\text{CoS}_s(\alpha)$. For each configuration and its associated boosting rounds, the confidence set is determined through the following sequential steps.

*(1) Clustering of single-effect components.* For each participating trait $l$ and boosting round $k$, let $\hat{\boldsymbol{\omega}}_l^k = (\hat{\omega}_{1l}^k, \ldots, \hat{\omega}_{Pl}^k)$ denote the proximity-smoothed probability vector over the $P$ candidate variants. Because SEC are weak learners, several rounds can revisit the same causal effect or nearby variants that are proxies in linkage disequilibrium (LD). If the selected variants $j_k^\star$ and $j_{k'}^\star$ from two rounds are close LD proxies, proximity smoothing produces similar profiles $\hat{\boldsymbol{\omega}}_l^k$ and $\hat{\boldsymbol{\omega}}_l^{k'}$ across variants and participating traits. This repeated representation accommodates uncertainty about variant identity in high-LD regions. By contrast, distinct causal effects under the same trait configuration generally produce different probability profiles. We therefore apply modularity-based hierarchical clustering within each configuration (Supplementary Note S.5.1). Each resulting cluster defines a candidate single-effect component $s$, and $\mathcal{K}_s$ contains the boosting rounds assigned to that component. For every $l \in T(s)$, the collection $\mathcal{W}_l^s = \{\hat{\boldsymbol{\omega}}_l^k : k \in \mathcal{K}_s\}$ contains $|\mathcal{K}_s|$ iteration-level probability vectors.

*(2) Defining the single-effect colocalization probability.* Within component $s$, we retain the pre-

viously defined joint profile log-likelihood change,

$$\Delta\mathcal{L}(k+1,k) = \mathcal{L}(\boldsymbol{\beta}_1^{k+1},\dots,\boldsymbol{\beta}_L^{k+1}) - \mathcal{L}(\boldsymbol{\beta}_1^{k},\dots,\boldsymbol{\beta}_L^{k}).$$

We aggregate the iteration-level probabilities for trait $l$ using the magnitude of this change,

$$\xi_{jl}^s = \frac{\sum\limits_{k\in\mathcal{K}_s} |\Delta\mathcal{L}(k+1,k)|\,\hat{\omega}_{jl}^k}{\sum\limits_{k\in\mathcal{K}_s} |\Delta\mathcal{L}(k+1,k)|}. \tag{S.2.13}$$

The denominator is positive for an accepted component. Weighting weak learners by their change in fit follows the gradient boosting ensemble construction[7]. Because each $\hat{\boldsymbol{\omega}}_l^k$ is non-negative and sums to one across variants, $\boldsymbol{\xi}_l^s = (\xi_{1l}^s,\dots,\xi_{Pl}^s)$ is also a probability distribution over the $P$ candidate variants.

We next combine the trait-specific probabilities for $l \in T(s)$ using normalized logarithmic pooling[8],

$$\widetilde{\xi}_j^s = \exp\left\{\frac{\nu}{|T(s)|}\sum_{l\in T(s)} \log \xi_{jl}^s\right\} = \prod_{l\in T(s)} \left(\xi_{jl}^s\right)^{\nu/|T(s)|},$$

$$\xi_j^s = \frac{\widetilde{\xi}_j^s}{\sum\limits_{m=1}^{P} \widetilde{\xi}_m^s}. \tag{S.2.14}$$

Here, $\widetilde{\xi}_j^s$ is the unnormalized logarithmic-pooling score. Only after normalization is $\xi_j^s$ a single-effect colocalization probability distribution over the candidate variants. For every $\nu > 0$, changing $\nu$ changes the concentration of this distribution without changing the variant ranking.

When $\nu = 1$, the unnormalized score is the geometric mean of the trait-specific probabilities. When $\nu = |T(s)|$, it is their full product. Interpreting the latter as accumulation of independent evidence requires an additional independence assumption. We use $\nu = 1.5$ as an empirically calibrated intermediate setting. This choice tempers evidence accumulation when the trait-specific inputs may be partially dependent or redundant, while keeping the pooled score comparable across different numbers of colocalizing traits. After normalization, $\nu$ acts as a concentration or sharpening parameter. It is not an estimate of residual correlation, genetic-effect correlation, or the effective number of independent traits.

For example, consider the following scenarios:

1) *Two-trait colocalization.* If $T(s) = \{1,2\}$, the unnormalized pooled score is

$$\widetilde{\xi}_j^s = \left(\xi_{j1}^s \xi_{j2}^s\right)^{\nu/2},$$

$$\log \widetilde{\xi}_j^s = \frac{\nu}{2}\left(\log \xi_{j1}^s + \log \xi_{j2}^s\right) = \nu\, \text{average}_{l\in\{1,2\}}\left(\log \xi_{jl}^s\right).$$

Thus, $\nu = 1$ gives geometric-mean pooling, $\nu = 1.5$ gives an intermediate sharpened pool, and $\nu = 2 = |T(s)|$ gives the full product.

2) *Ten-trait colocalization.* If $T(s) = \{1, \ldots, 10\}$, the unnormalized pooled score is

$$\widetilde{\xi}_j^s = \prod_{l=1}^{10} \left(\xi_{jl}^s\right)^{\nu/10},$$

$$\log \widetilde{\xi}_j^s = \frac{\nu}{10} \sum_{l=1}^{10} \log \xi_{jl}^s = \nu \, \text{average}_{l \in \{1,\ldots,10\}} \left(\log \xi_{jl}^s\right).$$

Here, $\nu = 1$ again gives geometric-mean pooling, $\nu = 1.5$ gives moderate sharpening, and only $\nu = 10 = |T(s)|$ gives the full product. In particular, $\nu = 2$ is not full-product pooling in the ten-trait setting.

In both examples, averaging the trait-specific log-probabilities before applying $\nu$ prevents the exponent from growing mechanically with the number of colocalizing traits. The single-effect colocalization probability is obtained by normalizing $\widetilde{\xi}_j^s$ across all $P$ variants, as shown in Equation (S.2.14).

Across simulations with 2, 5 and 10 traits, we evaluated $\nu \in \{1, 1.5, 2\}$ (Figure S15; Supplementary Note S.1.4). Power changed modestly and generally decreased as $\nu$ increased, whereas FDR remained below 0.05. The setting $\nu = 1$ was the most sensitive and could produce larger $\alpha$-level CoS. Within the two-trait simulations, FDR increased as $\nu$ approached the full-product value of 2 but remained below 0.05. The default $\nu = 1.5$ balances this empirical sensitivity and concentration behavior.

##### S.2.4.2 Construction of $\alpha$-level colocalized confidence sets and variant colocalization probabilities

*(3) Construction of* $\text{CoS}_s(\alpha)$. For each single-effect component $s$, order the normalized probabilities as $\xi_{(1)}^s \geq \cdots \geq \xi_{(P)}^s$. Let $p_0$ be the first rank at which their cumulative probability reaches $\alpha$. The corresponding $\alpha$-level single-effect colocalization confidence set is

$$p_0 = \min \left\{ p : \sum_{r=1}^{p} \xi_{(r)}^s \geq \alpha \right\},$$

$$\text{CoS}_s(\alpha) = \left\{ j : \xi_j^s \geq \xi_{(p_0)}^s \right\}. \qquad \text{(S.2.15)}$$

The second line includes all variants tied at the cutoff, consistent with the software implementation. We use $\alpha = 0.95$ by default. Each CoS is associated with $T(s)$, represents one putative shared effect, and is designed to contain its causal variant together with high-LD proxies.

A CoS is a set-level inferential object. In simulations, a set-level call succeeds when the retained set contains the causal variant and recovers the relevant trait configuration. High CoS power therefore measures coverage by a set of LD-linked candidates, not identification of one causal variant. Variant-level identification is evaluated separately using VCP and precision–recall analyses.

Following SuSiE, we discard a CoS when its purity is below 0.5 by default[1]. Purity is the minimum absolute correlation among all pairs of variants in the set. This filter removes sets spanning poorly correlated variants, which have limited single-effect interpretation.

We separately filter components with small contributions to the fitted profile log-likelihood because weak learners can capture random fluctuations as putative signals. Let $\Delta\mathcal{L}_l$ denote the overall trait-specific contribution, and let $\Delta\mathcal{L}_l^s$ denote the conditional contribution of component $s$ to trait $l$. By default, a component is retained if at least one trait $l \in T(s)$ satisfies both $\Delta\mathcal{L}_l \geq 0.025$ and $\Delta\mathcal{L}_l^s \geq 0.1\Delta\mathcal{L}_l$. Otherwise, it is discarded as a low-evidence component (Supplementary Note S.5.2). These likelihood filters are distinct from the LD-purity filter. Highly correlated CoS are refined separately in Supplementary Note S.2.4.3. Diagnostics for incomplete LD or mismatched LD and summary statistics are described in Supplementary Note S.2.5.

*Determination of variant colocalization probability (VCP).* Suppose that $S$ candidate components were constructed before filtering. Let $\widetilde{S} \leq S$ be the number retained, and relabel them as $s = 1, \ldots, \widetilde{S}$. For each retained component, let $Z_s \in \{1, \ldots, P\}$ denote its variant identity, with marginal distribution $\text{Pr}_Q(Z_s = j) = \xi_j^s$. We use the factorized working distribution

$$Q(Z_1 = z_1, \ldots, Z_{\widetilde{S}} = z_{\widetilde{S}}) = \prod_{s=1}^{\widetilde{S}} \xi_{z_s}^s.$$

Under this working distribution, the probability that variant $j$ is included in at least one retained component is

$$\text{VCP}_j = 1 - \prod_{s=1}^{\widetilde{S}} \left(1 - \xi_j^s\right). \tag{S.2.16}$$

The product form is exact under the stated factorized working distribution. The factorization itself is a modeling approximation. Fitting each component conditionally on residuals from the other components can reduce redundant representations of one signal, but it does not establish probabilistic independence. This construction parallels the aggregation of single-effect posterior probabilities into PIP in SuSiE[1,9,10]. Multiple causal variants in high LD can violate the approximation and increase false discoveries. Thus, CoS quantifies uncertainty within one shared effect, whereas $\text{VCP}_j$ summarizes support that variant $j$ participates in at least one retained effect.

##### S.2.4.3 Refining colocalization confidence sets

A key challenge arises when different SEC updates produce equivalent causal effects across different subsets of traits, analogous to ensemble methods where individual weak learners capture complementary aspects of the underlying pattern. This fragmentation across multiple SEC can result in detection of multiple incomplete colocalization events, and cannot be naturally coped with using the clustering approach when SEC are assembled. To address this, we further process CoS to merge different $\boldsymbol{T}^s$ sharing the same causal effects, in which process CoS with sub-threshold effects for some SEC might be recovered through recalibration after merger. To do so, we define *between-purity* as the minimum absolute correlation between variant pairs across two confidence sets, and refine CoS for the following scenarios:

- Merging two colocalization confidence sets $\text{CoS}_s$ and $\text{CoS}_{s'}$ with trait sets $T(s)$ and $T(s')$ respectively, requiring (i) at least one shared variant between the sets, and (ii) sufficient correlation, measured by both median between-purity exceeding 0.8 and minimum between-purity exceeding 0.5.

- Merging a trait-specific confidence set $\text{CS}_{s'}$ for trait $l \notin T(s)$ (where $T(s') = \{l\}$) into a colocalization set $\text{CoS}_s$ with trait set $T(s)$, requiring (i) at least one shared variant between the sets, and (ii) strong correlation, minimum between-purity exceeding 0.8.

- Merging two trait-specific confidence sets $\text{CS}_s$ and $\text{CS}_{s'}$ for traits $T(s) = \{l\}$ and $T(s') = \{l'\}$ respectively, requiring (i) at least one shared variant between the sets, and (ii) strong correlation, minimum between-purity exceeding 0.8.

Refined CoS on merged trait set $T(s) \cup T(s')$ will be recalibrated using $W_j^s$ and $W_j^{s'}$, $W_j = 1 - (1 - W_j^s)(1 - W_j^{s'})$, followed by applying Equation S.2.15. By constructing these refined CoS, we enhance the detection of shared causal effect across multiple traits, even if they were not simultaneously updated in one SEC. In our software implementation we added the prefix `merged_` to such refined CoS, to caution the users that the CoS was initially identified by distinct SEC but were deemed similar enough to be considered effectively the same update.

##### S.2.4.4 Normalized probability of colocalization in *ColocBoost*

Similar to configuration probability provided by Bayesian colocalization methods, such as PP.H4 from COLOC to quantify the probability of configuration $\boldsymbol{T}^s = (1, 1)$, we propose a Normalized Probability of Colocalization (NPC) as an empirical evidence in favor of colocalization over a trait-specific configuration. For each trait $l \in T(s)$ corresponding to $CoS_s(\alpha)$, we define the normalized probability of trait $l$ to be included in this configuration as

$$NP_l^s = 1 - \exp(-\lambda_l LRT_l^s), \quad \text{for} \quad l \in T(s), \tag{S.2.17}$$

where $LRT_l^s = -2\log(LR_l^s)$ is a log-likelihood ratio test statistic between the following two models, $M_{0,l}$ and $M_{1,l}^s$. Here, $M_{0,l}$ is the null model where all variants have zero effects $\beta_l = 0$ and $M_{1,l}^s$ is an alternative model that variants in $CoS_s(\alpha)$ have non-zero effects, with $\tilde{\beta}_l \neq 0$ estimated from *ColocBoost*. $\lambda_l$ is a trait-specific rate that adjust the scale of normalization based on a baseline log-likelihood ratio for each trait $l$. For example, $\lambda_l = 1/\Delta\mathcal{L}_{l,noise}$ is the accepted baseline of trait-specific log-likelihood for trait $l$ with a noise $\tilde{\beta}_{l,noise}$ in model $M_{1,l}$.

Let $NP_{max}^s = \max_{l \in T(s)}\{NP_l^s\}$. Then, the probability that only the strongest trait effect is significant among all colocalized traits, $NPUC_s$, can be defined as,

$$NPUC_s = NP_{max}^s \prod_{l \neq max} (1 - NP_l^s). \tag{S.2.18}$$

A higher $NPUC_s$ indicates that only one trait in $T(s)$ has strong $NP$, suggesting the signal is likely trait-specific. Then, Normalized Probability of Colocalization (NPC) is defined by

$$NPC_s = 1 - NPUC_s. \tag{S.2.19}$$

Given sufficient evidence of colocalization for the most likely configuration reported by *ColocBoost* (based on $NPC_s$), the best minimal colocalization configuration can be identified using $NP_l^s$ for trait $l$ within this most likely trait configuration. Our numerical studies suggest that applying lenient filters on these quantities ($NPC_s \geq 0.5$ and $NP_l^s \geq 0.2$) is sufficient to achieve well-controlled FDR while maintaining reasonable detection power.

#### S.2.5 Summary statistics extension and related practical features

In this section we provide implementation details describing how *ColocBoost* addresses common challenges in real-world genetic data integration. First, *ColocBoost* supports both individual-level and summary-statistics-based analyses, with optional use of external LD reference panels when individual-level data are unavailable. Second, it accommodates mixed data types across traits, as in our GWAS–xQTL integration. Third, it handles partial overlapping variant sets without requiring imputation or variant exclusion while retaining variants with partially missing LD data. Finally, it provides an LD-free mode under a single causal variant assumption, enabling analysis when LD is unavailable or poorly matched.

##### S.2.5.1 Mixed use of individual-level data and summary association statistics

Most multi-trait colocalization methods rely solely on summary statistics. Consequently, when individual-level data are available, it is necessary to first convert these data into summary statistics, such as z-scores or estimates of genetic effects for variants. This conversion process can result in a loss of statistical power due to the inherent loss of information such as sample size and MAF if not reported. Most crucially, using an estimated LD matrix from reference panel data or a single in-sample LD across multiple traits may lead to mismatched genetic variants being incorrectly estimated to affect trait specific association through LD, thereby increasing the risk of false positives or failure to capture true signals.

The flexibility of multi-task regression model enables *ColocBoost* to integrate data both at the individual-level or at the summary statistic level—*ColocBoost* can handle scenarios where the individual data is available for some traits (like xQTLs) and the summary data is available for other traits (disease/trait GWAS) (**Figure 1**), eliminating the need for separate analytical approaches when working with mixed data sources. This capability ensures that *ColocBoost* can leverage all available data optimally, preserving the statistical power and enhancing the accuracy of multi-trait colocalization analysis.

As introduced in Supplementary Note S.1.5 and **Algorithm S2**, we provide how SEC can operate on sufficient statistics and two strategies to approximate sufficient statistics from association summary data and external population-level reference data. Let $\mathbf{X}_l^T \boldsymbol{Y}_l$, $\mathbf{X}_l^T \mathbf{X}_l$, and $\boldsymbol{Y}_l^T \boldsymbol{Y}_l$ be the sufficient statistics for trait $l$. For *ColocBoost*, we assume both $\boldsymbol{Y}_l$ and $\mathbf{X}_l$ are standardized so that $\boldsymbol{Y}_l^T \boldsymbol{Y}_l = 1$, the summary statistic $\hat{\boldsymbol{\beta}}_l = \mathbf{X}_l^T \boldsymbol{Y}_l / N_l$, and $\mathbf{X}_l^T \mathbf{X}_l$ is equivalent to the LD matrix of the genotypes. The optimization problem in Equation (S.2.2) based on sufficient statistics can be written as

$$\begin{aligned}\mathcal{L}(\boldsymbol{\beta}_1, \cdots, \boldsymbol{\beta}_L) &= \sum_{l=1}^{L} \frac{1}{2} ||\boldsymbol{Y}_l - \mathbf{X}_l \boldsymbol{\beta}_l||_2^2 \\ &= \sum_{l=1}^{L} \frac{1}{2} (\boldsymbol{Y}_l - \mathbf{X}_l \boldsymbol{\beta}_l)^T (\boldsymbol{Y}_l - \mathbf{X}_l \boldsymbol{\beta}_l) \\ &:= \frac{1}{2} \sum_{l=1}^{L} \left[ \boldsymbol{Y}_l^T \boldsymbol{Y}_l - 2\boldsymbol{\beta}_l^T \mathbf{X}_l^T \boldsymbol{Y}_l + \boldsymbol{\beta}_l^T \mathbf{X}_l^T \mathbf{X}_l \boldsymbol{\beta}_l \right]. \qquad \text{(S.2.20)}\end{aligned}$$

and also the summary-based best single-effect variant $j_k^\star$ at boosting round $k$ for *joint updates* across traits can be written exclusively in terms of the summary statistic-level information

$$
\begin{aligned}
j_k^\star &= \arg\max_j \left( \sum_{l=1}^{L} |\boldsymbol{X}_{l,j}^T \boldsymbol{r}_l^k| \right) \\
&= \arg\max_j \left( \sum_{l=1}^{L} |\boldsymbol{X}_{l,j}^T \boldsymbol{Y}_l - \boldsymbol{X}_{l,j}^T \mathbf{X}_l \boldsymbol{\beta}_l^k| \right) \\
&= \arg\max_j \left( \sum_{l=1}^{L} |N_l \hat{\beta}_{l,j} - \mathbf{LD}_{l,j} \boldsymbol{\beta}_l^k| \right). \qquad \text{(S.2.21)}
\end{aligned}
$$

Therefore, the proximity smoothed gradient boosting algorithm is also a function of the summary statistics introduced in **Algorithm S2**. When individual-level data are available, we can directly calculate the sufficient statistics. When only association summary data are available, we adopt the two strategies to derive sufficient statistics from the summary data as detailed in Supplementary Note S.1.5. More technical details on the derivations of all necessary calculations in *ColocBoost* based on sufficient statistics are provided in Supplementary Note S.4.3 and S.4.4.

##### S.2.5.2 Handling partial overlapping variants across traits in *ColocBoost*

Heterogeneous association datasets from different sources often contain non-overlapping genetic variants due to differences in study design (population/cohort specific allele spectrum, sampling variations, different genotyping platforms, quality control procedures, etc). While genotype imputation methods (IMPUTE2[11] and BEAGLE[12] for individual level data, RAISS[13] and ImpG[14] for summary statistics) can recover missing genotypes and variants, they are not always reliable or feasible. Although discarding non-overlapping regions might be acceptable for pairwise colocalization, it can be severely problematic for multi-trait analysis as the chances of at least one trait missing a variant increase with each additional trait included. *ColocBoost* handles non-overlapping variants within its SEC by ignoring traits at loci where variants are missing, rather than discarding the entire loci from all traits. This approach ensures that only specific loci with missing data are impacted while preserving all available information from other loci. Thus, *ColocBoost* maintains detection power at minimum loss of information from data.

##### S.2.5.3 Accommodating missing or partial-overlapping LD in *ColocBoost*

When some traits completely lack LD information or share only partial variant coverage with other traits, the standard SEC updates will fail. A two-step procedure is adopted as a degenerated dynamic coupling strategy to cope with the situation. Firstly, for each trait, we assume at most one causal variant among those variants lacking LD information for trait $l$, and perform one iteration of *FineBoost* with proximity smoothing via a data-driven simplex $\delta_j^k$ without involving LD, by setting $LD_{j,j_k^\star}$ in Equation S.1.13 to 1. The simplex will be applied to each trait to yield a 95% confidence set from its trait-specific weights. We then evaluate whether pairs of traits potentially share the same causal variant, defined as having overlapping variant weights summing above 0.5. Traits will be temporarily partitioned into separate groups each representing a potential colocalization event. Next, we complete the boosting round by adopting the dynamic coupling strategy previously described: for each group involving more than one trait we apply *Logic 1*, and for groups only involving one trait we process them using *Logic 2.1*. A special case of this procedure is the LD-free operational mode, wherein *ColocBoost* completely ignores LD information and apply the described two-step algorithm across all traits and variants in the analysis.

##### S.2.5.4 *ColocBoost* with sample overlap or residual correlation

Overlapping samples are common in xQTL analyses spanning multiple modalities. Since this can induce dependencies (correlated residuals independent of genetic effects) across traits, in principle, accounting for these trait-trait correlations may improve methods performance. However, Foley et al.[15] showed that treating studies as independent typically yielded controlled FDR and robust power in capturing colocalizing traits while conferring an advantage of reduced computational complexity. To validate this, we considered additional multi-trait simulation studies which incorporated realistic trait-trait correlations estimated from FunGen-xQTL molecular trait datasets (see details in Supplemental Note S.7.2). Despite not modeling these trait correlations by default, *ColocBoost* maintained comparable power and FDR across all simulation scenarios (Figure S3c,d).

The equations below outline an optional diagonal precision-weighted extension. Because only diagonal precision terms enter the displayed objective, this should not be interpreted as a full multivariate residual-covariance likelihood. The extension is not used in the default analyses. Nevertheless, it is straightforward to incorporate overlapping sample adjustments in *ColocBoost*. Let $\mathbf{V} \in \mathbb{R}^{L\times L}$ be the residual covariance matrix of $L$ traits and $\mathbf{V} \neq \mathbf{I}_L$ for the independent studies. Let $\boldsymbol{\Theta} = \mathbf{V}^{-1}$ is the precision matrix of $\mathbf{V}$. We minimize the following optimization problem with the adjustment of residual covariance matrix,

$$\mathcal{L}(\boldsymbol{\beta}_1, \cdots, \boldsymbol{\beta}_L) := \sum_{l=1}^{L} \frac{1}{2}\Theta_{ll}||\boldsymbol{Y}_l - \mathbf{X}_l\boldsymbol{\beta}_l||_2^2, \tag{S.2.22}$$

where $\Theta_{ll}$ is the diagonal elements of $\boldsymbol{\Theta}$ for $l = 1, \cdots, L$. When the phenotypes contain the overlap samples, it is important to account for possible correlations among the measurements of the different phenotypes. Therefore, the best single-effect variant $j_k^\star$ at boosting round $k$ for *joint updates* across traits is given by

$$j_k^\star := \arg\max_j \left( \sum_{l=1}^{L} \Theta_{ll}|(\boldsymbol{X}_{j,l})^T \boldsymbol{r}_l^k| \right). \tag{S.2.23}$$

Then, the proximity smoothed gradient boosting algorithm is also adjusted for this residual covariance. The trait-specific proximity weight and smooth version of objective function can be written as (see detailed derivation in Supplementary Note S.4.2)

$$\xi_j(\boldsymbol{r}^k) := \frac{\dfrac{\Theta_{ll}}{N-1} \cdot \delta_j^k \exp\left(\dfrac{\Theta_{ll}|\boldsymbol{X}_j^T \boldsymbol{r}^k|/(N-1)}{\tau}\right)}{\sum_{j=1}^{P} \delta_j^k \exp\left(\dfrac{\Theta_{ll}|\boldsymbol{X}_j^T \boldsymbol{r}^k|/(N-1)}{\tau}\right)}, \tag{S.2.24}$$

with a close-form, smoothed solution of the sub-gradient optimization problem as follows

$$g^{\tau}_{smooth}(\boldsymbol{r}^k) = \tau \log \left( \sum_{j=1}^{P} \delta_j^k \exp \left( \frac{\Theta_{ll} |\boldsymbol{X}_j^T \boldsymbol{r}^k| / (N-1)}{\tau} \right) \right). \quad \text{(S.2.25)}$$

To estimate the residual covariance matrix **V**, we adapted the approach described in Urbut et al.[16], in which **V** was estimated from $z$-scores obtained from marginal association tests under the null: First, we pool the $z$-scores from all the fine-mapping regions considered, then filter out large (in magnitude) $z$-scores by only considering variants in which the largest absolute $z$-score across traits less than $2$. **V** is then estimated by correlation of these $z$-scores: denoting the number of variants used in this calculation by $J$, and letting $\hat{\boldsymbol{z}}_{0j}$ be the vector of null $z$-scores obtained from the $L$ association tests for variant $j$, we estimated **V** as

$$\hat{\mathbf{V}} = \frac{1}{J} \sum_{j=1}^{J} \hat{\boldsymbol{z}}_{0j} \hat{\boldsymbol{z}}_{0j}^T. \quad \text{(S.2.26)}$$

### S.3 Justifications and Theoretical properties of *FineBoost* and *ColocBoost*

This section establishes properties of the single-effect conditional inclusion probability $\hat{\boldsymbol{\omega}}^k$ used in the proximity-smoothed update. The formal results are stated for the single-trait case (*FineBoost*, $L = 1$) and, where specified, for a fixed positive reference measure. Applying the update trait by trait within a Single-Effect Coupler does not extend the convergence result to the complete multi-trait configuration search. Computational derivations, summary-statistics calculations, and parameter assessments are presented in the sections that follow.

#### S.3.1 Connection to the Correlation Minimization framework.

The starting point is the Correlation Minimization (CM) problem introduced by Freund et al.[2],

$$\text{CM} : \quad f^\star = \min_{\boldsymbol{r} \in \mathcal{P}_{\text{res}}} f(\boldsymbol{r}), \quad f(\boldsymbol{r}) = \|\mathbf{X}^T \boldsymbol{r}/(N-1)\|_\infty, \quad \text{(S.3.1)}$$

where $\mathcal{P}_{\text{res}} = \{\boldsymbol{r} \in \mathbb{R}^N : \boldsymbol{r} = \boldsymbol{Y} - \mathbf{X}\boldsymbol{\beta} \text{ for some } \boldsymbol{\beta}\}$ is the affine space of residuals. As shown by Freund et al.[2] (Proposition 3.2), the $\text{FS}_\varepsilon$ algorithm is an instance of subgradient descent on the CM problem, with a subgradient at $\boldsymbol{r}^k$ given by $\boldsymbol{g}^k = \text{sgn}(\boldsymbol{X}_{j_k^\star}^T \boldsymbol{r}^k)\, \boldsymbol{X}_{j_k^\star}/(N-1)$. Since $f(\boldsymbol{r})$ is non-smooth, the $\text{FS}_\varepsilon$ algorithm achieves only sublinear convergence at rate $O(1/\sqrt{k})$ (Theorem 3.1 in Freund et al.[2]).

We extend the standard CM by writing the objective equivalently as

$$f(\boldsymbol{r}) = \max_{\boldsymbol{\omega} \in \Delta_P} \sum_{j=1}^{P} \omega_j |\boldsymbol{X}_j^T \boldsymbol{r}/(N-1)|. \quad \text{(S.3.2)}$$

This proximity-smoothed objective is obtained by adding an entropic regularizer $-\tau D_{\text{KL}}(\boldsymbol{\omega} \| \boldsymbol{\delta})$ to this variational representation, following from the *Nesterov smoothing*[17] of the $\ell_\infty$-norm objective $f$, with the KL divergence serving as the prox-function on $\Delta_P$ and $\boldsymbol{\delta}$ as the prox-center. The resulting smoothed function satisfies the standard approximation bounds (Nesterov et al.[17],

Theorem 1),

$$f(\boldsymbol{r}) \;\le\; g^{\tau}_{\text{smooth}}(\boldsymbol{r}) \;\le\; f(\boldsymbol{r}) + \tau\, D_{\text{KL,max}}, \tag{S.3.3}$$

where $D_{\text{KL,max}} = \max_{\boldsymbol{\omega}\in\Delta_P} D_{\text{KL}}(\boldsymbol{\omega}\|\boldsymbol{\delta}) \le \log P$ (the upper bound attained when $\boldsymbol{\delta}$ is uniform). Thus $g^{\tau}_{\text{smooth}}$ uniformly approximates $f$ with accuracy controlled by $\tau \log P$. The cost of smoothing is small at our default $\tau = 0.01$.

#### S.3.2 Properties of $\hat{\omega}^k$

**Proposition S1** (Variational characterization and robustness of $\hat{\boldsymbol{\omega}}^k$). *Let $\boldsymbol{\delta} \in \Delta_P$ satisfy $\delta_j > 0$ for all $j$, and let $\tau > 0$. Define $v_j = |\boldsymbol{X}_j^T \boldsymbol{r}^k|/(N-1)$ for each variant $j$. The maximization problem*

$$\max_{\boldsymbol{\omega}\in\Delta_P} \left\{ \sum_{j=1}^{P} \omega_j\, v_j - \tau\, D_{\text{KL}}(\boldsymbol{\omega}\|\boldsymbol{\delta}) \right\}$$

*has a unique solution*

$$\hat{\omega}^k_j = \frac{\delta_j \exp(v_j/\tau)}{\sum_{m=1}^{P} \delta_m \exp(v_m/\tau)}, \tag{S.3.4}$$

*which satisfies the following properties.*

*I. Distributional properties.*

*(a) $\hat{\boldsymbol{\omega}}^k$ is a proper probability distribution on $\{1,\ldots,P\}$, with $\hat{\omega}^k_j > 0$ for all $j$ and $\sum_j \hat{\omega}^k_j = 1$.*

*(b) As $\tau \to 0^+$, $\hat{\boldsymbol{\omega}}^k \to \boldsymbol{f}e_{j^\star_k}$ (the point mass on the variant with maximal $v_j$, assuming uniqueness), recovering the standard FS$_\varepsilon$ subgradient[2].*

*(c) As $\tau \to \infty$, $\hat{\boldsymbol{\omega}}^k \to \boldsymbol{\delta}$.*

*(d) For a set $S \subseteq \{1,\ldots,P\}$ of variants in perfect LD (i.e., $\boldsymbol{X}_i = \pm\boldsymbol{X}_j$ for all $i,j \in S$, so that $v_i = v_j$ for all $i,j \in S$) with common reference weight ($\delta_i = \delta_j$ for $i,j \in S$), the solution assigns equal probability $\hat{\omega}^k_i = \hat{\omega}^k_j$ for all $i,j \in S$.*

*II. Robustness to $\tau$.*

*(e) (Ranking invariance.) The pairwise ratio satisfies*

$$\frac{\hat{\omega}^k_j}{\hat{\omega}^k_m} = \frac{\delta^k_j}{\delta^k_m} \exp\!\left(\frac{v_j - v_m}{\tau}\right). \tag{S.3.5}$$

*Hence $\hat{\omega}^k_j \ge \hat{\omega}^k_m$ if and only if $v_j - v_m \ge \tau \log(\delta^k_m/\delta^k_j)$. For uniform $\boldsymbol{\delta}^k$ within the LD neighborhood, the ordering is determined entirely by the data evidence $v_j$ and is invariant to $\tau$. More generally, the ranking can change only when $\tau$ crosses a critical value $\tau^\star_{jm} = (v_m - v_j)/\log(\delta^k_j/\delta^k_m)$ for some pair $(j,m)$, so the ordering is piecewise constant in $\tau$.*

*(f) (Bounded sensitivity.) For any two temperatures $\tau, \tau' > 0$, the maximum change in any pairwise log-ratio is bounded by*

$$\max_{j,m} \left| \log \frac{\hat{\omega}_j(\tau)}{\hat{\omega}_m(\tau)} - \log \frac{\hat{\omega}_j(\tau')}{\hat{\omega}_m(\tau')} \right| \le (v_{\max} - v_{\min}) \left| \frac{1}{\tau} - \frac{1}{\tau'} \right|, \tag{S.3.6}$$

*where $v_{\max} = \max_j v_j$ and $v_{\min} = \min_j v_j$. For a multiplicative perturbation $\tau' = (1+\epsilon)\tau$ with $|\epsilon|$ small, the right-hand side reduces to $O(\epsilon\,(v_{\max} - v_{\min})/\tau)$, so the total variation distance $\|\hat{\boldsymbol{\omega}}(\tau) - \hat{\boldsymbol{\omega}}(\tau')\|_{\text{TV}}$ is continuous in $\tau$ and vanishes as $\tau' \to \tau$.*

*(g) (Stability of credible sets.) Define the level-$\alpha$ credible set $\mathcal{C}_\alpha(\tau)$ as the smallest subset $S \subseteq \{1, \ldots, P\}$, ordered by decreasing $\hat{\omega}_j(\tau)$, such that $\sum_{j \in S} \hat{\omega}_j(\tau) \geq \alpha$. From the ranking invariance in (e), the greedy construction selects variants in the same order for all $\tau$ within a stability interval. When the lead variant $j_k^\star$ has substantially larger evidence $v_{j_k^\star}$ than competing variants—the typical scenario in well-powered GWAS loci—the credible set $\mathcal{C}_\alpha(\tau)$ is invariant to $\tau$ over a wide range.*

*Proof.* The objective is strictly concave in $\boldsymbol{\omega}$ on the relative interior of $\Delta_P$ (the KL divergence is strictly convex). Forming the Lagrangian with multiplier $\mu$ for the constraint $\sum_j \omega_j = 1$, the first-order condition for $\omega_j > 0$ gives $v_j - \tau \log(\omega_j/\delta_j) - \tau = \tau\mu$. Solving for $\omega_j$ yields $\omega_j = \delta_j \exp(v_j/\tau) \cdot \exp(-1-\mu)$, and enforcing $\sum_j \omega_j = 1$ produces Eqn. (S.3.4).

Property (a) is immediate from Eqn. (S.3.4). Properties (b) and (c) follow from the standard behavior of the softmax function as the temperature varies. Property (d) follows because $v_i = v_j$ and $\delta_i = \delta_j$ imply identical numerators, hence equal weights.

For property (e), dividing Eqn. (S.3.4) for indices $j$ and $m$ yields Eqn. (S.3.5) directly. The threshold condition and piecewise constancy follow from the monotonicity of the exponential function.

For property (f), note that $\log(\hat{\omega}_j(\tau)/\hat{\omega}_m(\tau)) = \log(\delta_j/\delta_m) + (v_j - v_m)/\tau$. The $\boldsymbol{\delta}$-dependent term cancels in the difference, leaving $(v_j - v_m)(1/\tau - 1/\tau')$, which is maximized when $|v_j - v_m| = v_{\text{max}} - v_{\text{min}}$. The continuity of the total variation distance follows because the map $\tau \mapsto \hat{\boldsymbol{\omega}}(\tau)$ is continuous (as a composition of continuous functions) and the total variation is a continuous metric on probability distributions.

For property (g), the credible set $\mathcal{C}_\alpha(\tau)$ is constructed by a greedy algorithm that adds variants in decreasing order of $\hat{\omega}_j(\tau)$. By (e), this ordering is constant for $\tau$ in each interval between consecutive critical values $\tau_{jm}^\star$. Within such an interval, the same variants are added in the same order; the only quantity that changes is when the cumulative sum reaches $\alpha$, which can only shrink or grow the set by boundary variants. When the evidence gap $v_{j_k^\star} - v_m$ is large for $m \neq j_k^\star$, the critical values $\tau_{jm}^\star$ are pushed to extreme values, widening the stability interval. □

#### S.3.3 Convergence guarantee of proximity smoothing gradient boosting algorithm

Freund et al.[2] established convergence properties for the corresponding forward-selection problem. Here we provide analogous convergence guarantees for the single-trait PGSB under the assumptions given below.

**Property 1**. Using Proposition 3.1 from Freund et al[2], we can write

$$\min_{k \in \{0,1,2,\cdots,K\}} g^{\tau}_{smooth}(\boldsymbol{r}^k) \leq g^{\tau}_{smooth}(\boldsymbol{r}^\star) + \frac{||\boldsymbol{r}^0 - \boldsymbol{r}^\star||_2^2}{2\eta_k(K+1)} + \frac{\eta_k G^2}{2} \tag{S.3.7}$$

where $\eta_k > 0$ is a dynamic learning rate of the gradient descent algorithm. $G$ is an upper bound on the gradient magnitudes such that $||\nabla g^{\tau}_{smooth}(\boldsymbol{r}^k)||_2$ is less than or equal to $G$ for each $k$.

*Proof.*

$$||\nabla g^{\tau}_{smooth}(\boldsymbol{r}^k)||_2^2 = ||\sum_j \xi_j(\boldsymbol{r}^k)\text{sgn}(\boldsymbol{X}_j^T \boldsymbol{r}^k)\boldsymbol{X}_j||_2^2 \leq \sum_j \xi_j^2(\boldsymbol{r}^k) + \sum_{j \neq j'} \xi_j(\boldsymbol{r}^k)\xi_{j'}(\boldsymbol{r}^k)|\rho_{jj'}| \tag{S.3.8}$$

where $\rho_{jj'}$ is the correlation between the columns $\boldsymbol{X}_j$ and $\boldsymbol{X}_{j'}$, which is LD between two variants $j$ and $j'$. Note that we considered the standardized genotype matrix, where the columns are $\ell_2$ normalized meaning $\boldsymbol{X}_j^T \boldsymbol{X}_j = 1$ for any $j$.

Note that the right hand side of Equation (S.3.8) is a convex function of the simplex vector $\left(\xi_1(\boldsymbol{r}^k), \xi_2(\boldsymbol{r}^k), \cdots, \xi_P(\boldsymbol{r}^k)\right)$ and it can be deduced that the maximum value of this quantity on this simplex will be attained at the corners of the simplex, where only one of the indices is $1$ and the rest $0$. This implies that

$$||\nabla g^{\tau}_{smooth}(\boldsymbol{r}^k)||_2^2 \leq 1 = G. \tag{S.3.9}$$

Also, the iterates $\boldsymbol{r}^k$ will eventually converge to the residuals for the least squares regression, $\boldsymbol{r}^\star = \hat{\boldsymbol{r}}_{LS} = \boldsymbol{Y} - \mathbf{X}\hat{\boldsymbol{\beta}}_{LS}$ and we can write

$$||\boldsymbol{r}^0 - \boldsymbol{r}^\star||_2 = ||\boldsymbol{Y} - (\boldsymbol{Y} - \mathbf{X}\hat{\boldsymbol{\beta}}_{LS})||_2 = ||\mathbf{X}\hat{\boldsymbol{\beta}}_{LS}||_2, \tag{S.3.10}$$

and also $\boldsymbol{X}_j^T \boldsymbol{r}^\star = \boldsymbol{X}_j^T \hat{\boldsymbol{r}}_{LS} = 0$ for each $j$. So, from Equation (S.1.14), we get

$$g^{\tau}_{smooth}(\boldsymbol{r}^\star) = 0. \tag{S.3.11}$$

Then using derivations in Equations (S.3.9), (S.3.10) and (S.3.11), we can write Equation (S.3.7) as

$$\min_{k \in \{0,1,2,\cdots,K\}} g^{\tau}_{smooth}(\boldsymbol{r}^k) \leq \frac{||\mathbf{X}\hat{\boldsymbol{\beta}}_{LS}||_2^2}{2\epsilon(K+1)} + \frac{\epsilon}{2}. \tag{S.3.12}$$

Additionally, it has been shown previously in Freund et al[2] that

$$\sup_{\boldsymbol{r} \in \mathbb{R}^N} |g(\boldsymbol{r}) - g^{\tau}_{smooth}(\boldsymbol{r})| \leq \tau \log(P) \tag{S.3.13}$$

implying

$$\min_{k \in \{0,1,2,\cdots,K\}} g(\boldsymbol{r}^k) \leq \frac{||\mathbf{X}\hat{\boldsymbol{\beta}}_{LS}||_2^2}{2\epsilon(K+1)} + \frac{\epsilon}{2} + \tau \log(P), \tag{S.3.14}$$

where $\tau > 0$ can be any positive quantity. Then, we can write

$$\min_{k \in \{0,1,2,\cdots,K\}} g(\boldsymbol{r}^k) = \min_{k \in \{0,1,2,\cdots,K\}} ||\nabla \mathcal{L}_N(\boldsymbol{\beta}^k)||_\infty \leq \frac{||\mathbf{X}\hat{\boldsymbol{\beta}}_{LS}||_2^2}{2N\epsilon(K+1)} + \frac{\epsilon}{2N}, \tag{S.3.15}$$

where $\mathcal{L}_N(\cdot)$ is the least squares loss with $N$ samples. This derivation replicates Equation 39 of A.3.2 in Freund et al[2]. Therefore, we can replicate all the results in Theorem 3.1 of Freund et al[2]. □

For analytical clarity, we state the result for a fixed reference measure $\boldsymbol{\delta}$; in practice, *ColocBoost* uses an adaptive $\boldsymbol{\delta}^k$ that is updated at each iteration, as discussed in Remark 2 below.

**Proposition S2** (Convergence of PGSB). *Consider the PGSB algorithm with fixed reference measure $\boldsymbol{\delta}$, constant learning rate $\eta > 0$, and initialization $\boldsymbol{r}^0 = \boldsymbol{f}Y$. Assume the columns of $\boldsymbol{X}$*

*have unit $\ell_2$ norm. Then for any $k \geq 0$, there exists an index $i \in \{0, \ldots, k\}$ such that*

$$\|\boldsymbol{X}^T \boldsymbol{r}^i/(N-1)\|_\infty \;\leq\; \frac{\|\boldsymbol{X}\hat{\boldsymbol{\beta}}^{\mathsf{LS}}\|_2^2}{2\eta(K+1)(N-1)^2} + \frac{\eta}{2(N-1)^2} + \tau D_{\mathsf{KL,max}},$$

*where $\hat{\boldsymbol{\beta}}^{\mathsf{LS}}$ is a least squares solution and $D_{\mathsf{KL,max}} \leq \log P$.*

*Proof.* Since the gradient of $g^\tau_{\mathsf{smooth}}$ is a convex combination of vectors $\boldsymbol{X}_j/(N-1)$ (weighted by $\hat{\omega}^k_j \geq 0$ summing to 1), the triangle inequality gives $\|\nabla g^\tau_{\mathsf{smooth}}\|_2 \leq (N-1)^{-1}$. Setting $G = (N-1)^{-1}$ and applying the standard convergence bound for projected gradient descent (Property 1 above; Freund et al.[2], Proposition 3.1; Nesterov et al.[17]) with step-size $\eta$ on the smoothed objective over $\mathcal{P}_{\mathsf{res}}$,

$$\min_{i\in\{0,\ldots,k\}} g^\tau_{\mathsf{smooth}}(\boldsymbol{r}^i) \;\leq\; g^\tau_{\mathsf{smooth}}(\boldsymbol{r}^\star_\tau) + \frac{\|\boldsymbol{r}^0 - \boldsymbol{r}^\star_\tau\|_2^2}{2(k+1)\eta} + \frac{\eta G^2}{2},$$

where $\boldsymbol{r}^\star_\tau$ minimizes $g^\tau_{\mathsf{smooth}}$ over $\mathcal{P}_{\mathsf{res}}$. Using the Nesterov approximation bounds (S.3.3), $f(\boldsymbol{r}^i) \leq g^\tau_{\mathsf{smooth}}(\boldsymbol{r}^i)$ and $g^\tau_{\mathsf{smooth}}(\boldsymbol{r}^\star_\tau) \leq f(\boldsymbol{r}^{\mathsf{LS}}) + \tau D_{\mathsf{KL,max}} = \tau D_{\mathsf{KL,max}}$, since $f(\boldsymbol{r}^{\mathsf{LS}}) = 0$. Noting $\|\boldsymbol{r}^0 - \boldsymbol{r}^\star_\tau\|_2 \leq \|\boldsymbol{X}\hat{\boldsymbol{\beta}}^{\mathsf{LS}}\|_2$ and $G = (N-1)^{-1}$ completes the bound. □

**Remark 1.** *Comparing with Theorem 3.1(iv) of Freund et al.[2] for FS$_\varepsilon$, the bound above has the same structural form with an additional $\tau D_{\mathsf{KL,max}}$ term reflecting the cost of smoothing. This additive penalty is paid for distributing the update across LD-proximal variants rather than selecting a single one. Setting $\tau = 0$ eliminates this term and recovers the original CM convergence guarantee.*

**Remark 2.** *In practice, ColocBoost uses an iteration-dependent reference measure $\boldsymbol{\delta}^k$ that adapts to the current "seed" variant $j^\star_k$ identified from the residual $\boldsymbol{r}^k$. This adaptive scheme is motivated by the sequential nature of the algorithm that we leverage to avoid pre-specifying the LD neighborhood of all potential signals simultaneously, which is not feasible when the number and locations of causal signals are unknown a priori for fixed $\boldsymbol{\delta}$. This is directly analogous to the iterative structure of SuSiE[1], where each single-effect component is fitted conditionally on the residuals from all other components.*

#### S.3.4 Structural analogy with single-effect posterior inclusion probability in SuSiE.

We now formalize the relationship between the single-effect conditional inclusion probability $\hat{\boldsymbol{\omega}}^k$ in *FineBoost* and the single-effect posterior $\boldsymbol{\alpha}_l$ in SuSiE[1], where the variational posterior probability that the $l$-th causal effect is at variant $j$ takes the form

$$\alpha_{l,j} \propto \pi_j \cdot \mathsf{BF}_j, \quad \text{where } \log \mathsf{BF}_j = \frac{z_j^2}{2} \cdot \frac{\sigma_0^2\|\boldsymbol{X}_j\|_2^2}{\sigma^2 + \sigma_0^2\|\boldsymbol{X}_j\|_2^2} + C,$$

with $\pi_j$ denoting the prior inclusion probability, $\mathsf{BF}_j$ the Bayes factor under a normal prior $\beta \sim N(0, \sigma_0^2)$, and $z_j = \boldsymbol{X}_j^T \boldsymbol{r}/\sqrt{N-1}$ the marginal z-score. For standardized genotypes this simplifies to $\log \mathsf{BF}_j \approx z_j^2/(2c)$ for a constant $c = 1 + \sigma^2/(\sigma_0^2 N)$.

**Proposition S3.** *The single-effect probabilities in FineBoost and SuSiE share a common soft-max structure. FineBoost (from Proposition S1):*

$$\hat{\omega}_j \propto \delta_j \cdot \exp\Big(\frac{|z_j|}{\tau'}\Big), \quad \tau' = \tau\sqrt{N-1}.$$

*SuSiE[1]:*

$$\alpha_j \propto \pi_j \cdot \exp\Big(\frac{z_j^2}{2c}\Big).$$

*Both have the form* (reference measure) $\times$ exp(data evidence)*, normalized to sum to* $1$ *over variants.*

The data evidence terms are structurally analogous between the two frameworks. In SuSiE, the quadratic form $z_j^2/2$ arises from the Gaussian likelihood and Gaussian prior on effect sizes, yielding the standard normal-theory Bayes factor[18]. In *FineBoost*, the linear form $|z_j|$ arises from the $\ell_\infty$-norm CM objective and its Nesterov smoothing: the log-sum-exp function is the smooth approximation of the maximum, and the corresponding Gibbs weights naturally involve $\exp(|z_j|/\tau')$ rather than $\exp(z_j^2)$. From a Bayesian perspective, $|z_j|$ can also be interpreted as arising from a Laplace (double-exponential) prior on the effect size $\beta$.

The reference measures differ between SuSiE and *FineBoost* because as a Bayesian method, $\pi_j$ in SuSiE is a prior inclusion probability typically uniform or estimated via empirical Bayes from the full set of data[19]. In *FineBoost*, $\boldsymbol{\delta}^k$ serves as the prox-center of the KL divergence in the Nesterov smoothing formulation and is data-driven to guide the allocation toward variants in the LD neighborhood of $j_k^\star$.

The analogy extends from single iterations to single-effect components in *FineBoost*, where within each component, $\hat{\boldsymbol{\omega}}^k$ are aggregated into a trait-specific single-effect probability $\xi_{jl}^s$ by weighting its contribution proportionally to its profile log-likelihood gain, following the standard gradient boosting ensemble principle[7], in which each weak learner's influence on the aggregate is determined by the loss reduction it achieved. Since a convex combination of probability distributions on $\{1, \ldots, P\}$ is itself a probability distribution, $W_{l,j}^s$ inherits the properties established for $\hat{\boldsymbol{\omega}}^k$ in Proposition S1.

#### S.3.5 Clarification of the probability hierarchy in *ColocBoost*: from per-iteration weights to variant-level inference

*ColocBoost* provides variant-level probability measures that propagate uncertainty from individual boosting iterations through multi-trait aggregation to a final per-variant quantity. This section clarifies the layered structure of probability hierarchy, tracing how per-iteration variant probability are successively aggregated across iterations, traits, and single-effect components to yield the variant-level measures (VPA and VCP) used for downstream inference.

1. *Per-iteration trait-specific variant probability* $\hat{\omega}_{l,j}^k$: the probability assigned to variant $j$ for trait $l$ at boosting iteration $k$, arising from the Nesterov-smoothed objective (Proposition S1; Supplementary Note S.3.2).

2. *Per-single-effect, trait-specific probability* $\xi_{jl}^s$ (trait-specific single-effect probability): a weighted average of $\hat{\omega}_{jl}^k$ across iterations $k$ belonging to single-effect component $s$, with

weights proportional to profile log-likelihood gains. Specifically, for each colocalization event $s$ with trait configuration $T(s)$, we re-normalized $\hat{\omega}_{l,j}^k$ for $l \in T(s)$ by their corresponding profile log-likelihood gains $\Delta\mathcal{L}(k+1,k) = \mathcal{L}(\boldsymbol{\beta}_1^{k+1}, \cdots, \boldsymbol{\beta}_L^{k+1}) - \mathcal{L}(\boldsymbol{\beta}_1^k, \cdots, \boldsymbol{\beta}_L^k)$. This yields a trait-specific single-effect probability for each variant $j$ and trait $l \in T(s)$,

$$\xi_{jl}^s = \frac{\sum_{k \in \mathcal{K}_s} \hat{\omega}_{l,j}^k |\Delta\mathcal{L}(k+1,k)|}{\sum_{k \in \mathcal{K}_s} |\Delta\mathcal{L}(k+1,k)|}.$$

This aggregation follows the standard gradient boosting ensemble principle[7], in which each weak learner's influence on the aggregate is determined by the loss reduction it achieved. Since a convex combination of probability distributions on $\{1, \ldots, P\}$ is itself a probability distribution, $\xi_{jl}^s$ inherits the properties established for $\hat{\omega}^k$ in Proposition S1.

3. *Per-single-effect, cross-trait probability* $\xi_j^s$ (single-effect colocalization probability): the geometric mean of $\xi_{jl}^s$ across colocalizing traits, aggregating independent sources of evidence via logarithmic pooling. For multi-trait colocalization, trait-specific probabilities $\xi_{jl}^s$ are combined across colocalizing traits via a geometric mean, $\xi_j^s = \prod_{l \in T(s)} (\xi_{jl}^s)^{\nu/|\boldsymbol{T}^s=1|}$ which aggregates all traits with $T_l^s = 1$ into a single-effect colocalization inclusion probability for variant j supporting $\boldsymbol{T}^s$. Equivalently, $\log \xi_j^s = \frac{1}{|\boldsymbol{T}^s=1|} \sum_l \log \xi_{jl}^s$, which averages log-probabilities across traits. This logarithmic pooling is the natural aggregation when combining conditionally independent sources of evidence on the same event[8]. We calibrate the single-effect colocalization inclusion probability $\xi_j^s$ through extensive simulation studies (Figure S15; Supplementary Note S1.4), evaluating the coverage properties and ensuring that the probabilities accurately reflect uncertainty in variant selection across different numbers of colocalizing traits. We set $\nu = 1.5$ by default to ensure consistent scale across different numbers of colocalizing traits.

4. *Variant-level probability* (VPA for single-trait, VCP for multi-trait): $1 - \prod_s (1 - \xi_j^s)$, combining evidence across components under the same conditional independence assumption used by SuSiE for PIP. Specially, the VCP aggregation across single-effect components, VCP$_j = 1 - \prod_s (1 - \xi_j^s)$, assumes conditional independence of the component-level probabilities $\xi_j^s$ given the data. This is the same independence structure employed by SuSiE to compute PIP from single-effect posteriors, where PIP$_j = 1 - \prod_{l=1}^L (1 - \alpha_{l,j})$ . In both frameworks, the independence assumption arises because each component is fitted *conditionally* on residuals from all other components.

### S.4 Computational derivations of *FineBoost* and *ColocBoost*

In this section, we will show important computational details as well as some theoretical properties of *FineBoost* and *ColocBoost*, including the derivation of the objective function and weights for the proximity smoothing gradient boosting algorithm for single trait (S.4.1), the convergence guarantee of sub-gradient with the smoothing constraint (S.3.3), derivation of the objective function and weights for independent traits (S.4.2), derivation of sufficient statistics from association summary statistics data (S.4.3), derivation of profile log-likelihood using sufficient statistics (S.4.4), and statistical reliability of the model parameters (S.4.5).

#### S.4.1 Derivation of the objectives and weights based on KL divergence for the proximity smoothing gradient boosting algorithm

For a given simplex $\delta_j$ for $j$-th genetic variant, the close-form, smoothed solution of the sub-gradient optimization objective function is given by

$$g^{\tau}_{smooth}(\boldsymbol{r}) = \tau \log \left( \sum_{j=1}^{P} \delta_j \exp \left( \frac{|\boldsymbol{X}_j^T \boldsymbol{r}|/(N-1)}{\tau} \right) \right). \tag{S.1.14}$$

*Proof.* Recall that the objective function with smoothing constraint is given by Equation (S.1.12),

$$g_{smooth}(\boldsymbol{r}) = \max_{\boldsymbol{\omega}} \sum_j \omega_j |\boldsymbol{X}_j^T \boldsymbol{r}|/(N-1) - \tau\rho(\boldsymbol{\omega}), \quad \sum_{j=1}^{P} \omega_j = 1, \ \omega_j \geq 0 \tag{S.1.12}$$

where we use $\rho(\boldsymbol{\omega})$ to captures the KL-divergence of a probability vector $\boldsymbol{\omega}$ with respect to a simplex $\boldsymbol{\delta}$,

$$\rho(\boldsymbol{\omega}) = \sum_{j=1}^{P} \omega_j \log(\omega_j) - \sum_{j=1}^{P} \omega_j \log(\delta_j) := KL(\boldsymbol{\omega}||\boldsymbol{\delta})$$

where $\delta_j$ is a probability vector that assigns weights to each variant $j$ (see detailed formulation of data-driven local association simplex in each boosting round in Supplementary Note S.1.2). Therefore, we consider the following constraint optimization problem to optimize the Equation (S.1.14). Let the loss function be

$$f(\boldsymbol{\omega}, \lambda) = \sum_j \omega_j |\boldsymbol{X}_j^T \boldsymbol{r}|/(N-1) - \tau \left( \sum_{j=1}^{P} \omega_j \log(\omega_j) - \sum_{j=1}^{P} \omega_j \log(\delta_j) \right) - \lambda \left( \sum_{j=1}^{P} \omega_j - 1 \right) \tag{S.4.1}$$

where $\lambda > 0$ is the Lagrange multiplier. Then, we take the derivative of $f(\boldsymbol{\omega}, \lambda)$ with respect to $\omega_j$ for $j = 1, \cdots, P$ and $\lambda$.

$$\begin{aligned} \frac{\partial f(\boldsymbol{\omega}, \lambda)}{\partial \omega_j} &= |\boldsymbol{X}_j^T \boldsymbol{r}|/(N-1) - \tau \left( \log(\omega_j) + 1 - \log(\delta_j) \right) - \lambda \\ &= |\boldsymbol{X}_j^T \boldsymbol{r}|/(N-1) - \lambda - \tau - \tau \log(\omega_j/\delta_j) \end{aligned} \tag{S.4.2}$$

$$\frac{\partial f(\boldsymbol{\omega}, \lambda)}{\partial \lambda} = \sum_{j=1}^{P} \omega_j - 1 = 0 \tag{S.4.3}$$

From Equation (S.4.2), we solve for $\omega_j$, which is

$$\begin{aligned} \log\left(\frac{\hat{\omega}_j}{\delta_j}\right) &= \frac{|\boldsymbol{X}_j^T \boldsymbol{r}|/(N-1) - \lambda - \tau}{\tau} \\ \Leftrightarrow \hat{\omega}_j &= \delta_j \exp\left( \frac{|\boldsymbol{X}_j^T \boldsymbol{r}|/(N-1) - \lambda - \tau}{\tau} \right) \end{aligned} \tag{S.4.4}$$

Then, we plug Equation (S.4.4) into Equation (S.4.2) to solve for $\lambda$.

$$\begin{aligned}
&\sum_{j=1}^{P} \delta_j \,\text{exp}\left(\frac{|\boldsymbol{X}_j^T \boldsymbol{r}|/(N-1) - \tau - \lambda}{\tau}\right) - 1 = 0 \\
\Leftrightarrow &\sum_{j=1}^{P} \delta_j \,\text{exp}\left(\frac{|\boldsymbol{X}_j^T \boldsymbol{r}|/(N-1)}{\tau}\right) = \text{exp}\left(1 + \frac{\lambda}{\tau}\right) \\
\Leftrightarrow &\hat{\lambda} = \tau \,\text{log}\left(\sum_{j=1}^{P} \delta_j \,\text{exp}\left(\frac{|\boldsymbol{X}_j^T \boldsymbol{r}|/(N-1)}{\tau}\right)\right) - \tau \qquad \text{(S.4.5)}
\end{aligned}$$

Rewrite Equation (S.4.4) as $|\boldsymbol{X}_j^T \boldsymbol{r}|/(N-1) = \tau\,\text{log}(\hat{\omega}_j/\delta_j) + \tau + \lambda$ and then plug Equation (S.4.4) and Equation (S.4.5) into Equation (S.4.1). Recall that we also have $\sum_{j=1}^{P} \hat{\omega}_j = 1$.

$$\begin{aligned}
f(\hat{\boldsymbol{\omega}}, \hat{\lambda}) &= \sum_{j=1}^{P} \hat{\omega}_j(\tau\,\text{log}(\hat{\omega}_j/\delta_j) + \tau + \hat{\lambda}) - \tau \sum_{j=1}^{P} \hat{\omega}_j \,\text{log}(\hat{\omega}_j/\delta_j) - \hat{\lambda}\left(\sum_{j=1}^{P} \hat{\omega}_j - 1\right) \\
&= \tau + \hat{\lambda} \\
&= \tau\,\text{log}\left(\sum_{j=1}^{P} \delta_j \,\text{exp}\left(\frac{|\boldsymbol{X}_j^T \boldsymbol{r}|/(N-1)}{\tau}\right)\right) \\
&= g^{\tau}_{smooth}(\boldsymbol{r}) \qquad \text{(S.4.6)}
\end{aligned}$$

□

Therefore, we take the derivative of $g^{\tau}_{smooth}(\boldsymbol{r})$.

$$\begin{aligned}
\frac{\partial g^{\tau}_{smooth}(\boldsymbol{r})}{\partial \boldsymbol{r}} =& \tau \times \frac{1}{\sum_{j=1}^{P} \delta_j \,\text{exp}\left(\frac{|\boldsymbol{X}_j^T \boldsymbol{r}|/(N-1)}{\tau}\right)} \times \sum_{j=1}^{P} \delta_j \left[\frac{\partial}{\partial \boldsymbol{r}} \text{exp}\left(\frac{|\boldsymbol{X}_j^T \boldsymbol{r}|/(N-1)}{\tau}\right)\right] \\
=& \tau \times \sum_{j=1}^{P} \left[\frac{\delta_j \,\text{exp}\left(\frac{|\boldsymbol{X}_j^T \boldsymbol{r}|/(N-1)}{\tau}\right)}{\sum_{j=1}^{P} \delta_j \,\text{exp}\left(\frac{|\boldsymbol{X}_j^T \boldsymbol{r}|/(N-1)}{\tau}\right)}\right] \times \frac{1/(N-1)}{\tau} \times \text{sgn}(\boldsymbol{X}_j^T \boldsymbol{r})\boldsymbol{X}_j \\
=& \sum_{j=1}^{P} \left[\frac{\frac{1}{N-1} \cdot \delta_j \,\text{exp}\left(\frac{|\boldsymbol{X}_j^T \boldsymbol{r}|/(N-1)}{\tau}\right)}{\sum_{l=1}^{P} \delta_l \,\text{exp}\left(\frac{|\boldsymbol{X}_l^T \boldsymbol{r}|/(N-1)}{\tau}\right)}\right] \times \text{sgn}(\boldsymbol{X}_j^T \boldsymbol{r})\boldsymbol{X}_j \qquad \text{(S.4.7)}
\end{aligned}$$

Therefore,

$$\nabla g^{\tau}_{smooth}(\boldsymbol{r}) = \sum_{j=1}^{P} \hat{\omega}^{k}_{j} \text{sgn}(\boldsymbol{X}_j^T \boldsymbol{r}) \boldsymbol{X}_j \tag{S.4.8}$$

where

$$\hat{\omega}^{k}_{j} := \frac{\frac{1}{N-1} \cdot \delta_j \exp\left(\frac{|\boldsymbol{X}_j^T \boldsymbol{r}|/(N-1)}{\tau}\right)}{\sum_{l=1}^{P} \delta_l \exp\left(\frac{|\boldsymbol{X}_l^T \boldsymbol{r}|/(N-1)}{\tau}\right)}. \tag{S.4.9}$$

#### S.4.2 Derivation of the objectives and weights based on KL divergence for independent traits

For a given simplex $\delta_j$ for $j$-th genetic variant and the precision matrix of the residual covariance, $\boldsymbol{\Theta} = \mathbf{V}^{-1}$, the close-form, smoothed solution of the sub-gradient optimization objective function for independent $L$ traits is given by

$$g^{\tau}_{smooth}(\boldsymbol{r}) = \tau \log \left( \sum_{j=1}^{P} \delta_j \exp\left( \frac{\Theta_{ll} |\boldsymbol{X}_j^T \boldsymbol{r}|/(N-1)}{\tau} \right) \right). \tag{S.2.25}$$

*Proof.* Recall that the optimization problem with the adjustment of residual covariance matrix is given by Equation (S.2.22),

$$\mathcal{L}(\boldsymbol{\beta}_1, \cdots, \boldsymbol{\beta}_R) := \sum_{l=1}^{L} \frac{1}{2} \Theta_{ll} ||\boldsymbol{Y}_l - \mathbf{X}_l \boldsymbol{\beta}_r||_2^2. \tag{S.2.22}$$

The objective function with smoothing constraint can be written as

$$g_{smooth}(\boldsymbol{r}) = \max_{\boldsymbol{\omega}} \sum_{j} \omega_j \Theta_{ll} |\boldsymbol{X}_j^T \boldsymbol{r}|/(N-1) - \tau \rho(\boldsymbol{\omega}), \quad \sum_{j=1}^{P} \omega_j = 1,\ \omega_j \geq 0 \tag{S.4.10}$$

with same $\rho(\boldsymbol{\omega})$ introduced in Supplementary Note S.4.1). Therefore, we consider the following constraint optimization problem to optimize the Equation (S.4.10) and the loss function is

$$f(\boldsymbol{\omega}, \lambda) = \sum_{j} \omega_j \Theta_{ll} |\boldsymbol{X}_j^T \boldsymbol{r}|/(N-1) - \tau \left( \sum_{j=1}^{P} \omega_j \log(\omega_j) - \sum_{j=1}^{P} \omega_j \log(\delta_j) \right) - \lambda \left( \sum_{j=1}^{P} \omega_j - 1 \right) \tag{S.4.11}$$

where $\lambda > 0$ is the Lagrange multiplier. Similar to Supplementary Note S.4.1, we take the derivative of $f(\boldsymbol{\omega}, \lambda)$ with respect to $\omega_j$ for $j = 1, \cdots, P$ and $\lambda$.

$$\frac{\partial f(\boldsymbol{\omega}, \lambda)}{\partial \omega_j} = \Theta_{ll} |\boldsymbol{X}_j^T \boldsymbol{r}|/(N-1) - \lambda - \tau - \tau \operatorname{log}(\omega_j/\delta_j) \tag{S.4.12}$$

$$\frac{\partial f(\boldsymbol{\omega}, \lambda)}{\partial \lambda} = \sum_{j=1}^{P} \omega_j - 1 = 0 \tag{S.4.13}$$

From Equation (S.4.12), we solve for $\omega_j$, which is

$$\begin{aligned} \operatorname{log}\left(\frac{\hat{\omega}_j}{\delta_j}\right) &= \frac{\Theta_{ll} |\boldsymbol{X}_j^T \boldsymbol{r}|/(N-1) - \lambda - \tau}{\tau} \\ \Leftrightarrow \hat{\omega}_j &= \delta_j \operatorname{exp}\left(\frac{\Theta_{ll} |\boldsymbol{X}_j^T \boldsymbol{r}|/(N-1) - \lambda - \tau}{\tau}\right) \end{aligned} \tag{S.4.14}$$

Then, we plug Equation (S.4.14) into Equation (S.4.12) to solve for $\lambda$.

$$\begin{aligned} &\sum_{j=1}^{P} \delta_j \operatorname{exp}\left(\frac{\Theta_{ll} |\boldsymbol{X}_j^T \boldsymbol{r}|/(N-1) - \tau - \lambda}{\tau}\right) - 1 = 0 \\ \Leftrightarrow \hat{\lambda} &= \tau \operatorname{log}\left(\sum_{j=1}^{P} \delta_j \operatorname{exp}\left(\frac{\Theta_{ll} |\boldsymbol{X}_j^T \boldsymbol{r}|/(N-1)}{\tau}\right)\right) - \tau \end{aligned} \tag{S.4.15}$$

Rewrite Equation (S.4.14) as $|\boldsymbol{X}_j^T \boldsymbol{r}|/(N-1) = \tau \operatorname{log}(\hat{\omega}_j/\delta_j) + \tau + \lambda$ and then plug Equation (S.4.14) and Equation (S.4.15) into Equation (S.4.11). Recall that we also have $\sum_{j=1}^{P} \hat{\omega}_j = 1$.

$$f(\hat{\boldsymbol{\omega}}, \hat{\lambda}) = \tau \operatorname{log}\left(\sum_{j=1}^{P} \delta_j \operatorname{exp}\left(\frac{\Theta_{ll} |\boldsymbol{X}_j^T \boldsymbol{r}|/(N-1)}{\tau}\right)\right) = g_{smooth}^{\tau}(\boldsymbol{r}) \tag{S.4.16}$$

□

Take the derivative of $g_{smooth}^{\tau}(\boldsymbol{r})$,

$$\frac{\partial g_{smooth}^{\tau}(\boldsymbol{r})}{\partial \boldsymbol{r}} = \sum_{j=1}^{P} \left[\frac{\frac{1}{N-1} \cdot \delta_j \operatorname{exp}\left(\frac{\Theta_{ll} |\boldsymbol{X}_j^T \boldsymbol{r}|/(N-1)}{\tau}\right)}{\sum_{l=1}^{P} \delta_l \operatorname{exp}\left(\frac{\Theta_{ll} |\boldsymbol{X}_l^T \boldsymbol{r}|/(N-1)}{\tau}\right)}\right] \times \operatorname{sgn}(\boldsymbol{X}_j^T \boldsymbol{r}) \boldsymbol{X}_j \tag{S.4.17}$$

we obtain

$$\nabla g_{smooth}^{\tau}(\boldsymbol{r}) = \sum_{j=1}^{P} \xi_j(\boldsymbol{r}) \operatorname{sgn}(\boldsymbol{X}_j^T \boldsymbol{r}) \boldsymbol{X}_j \tag{S.4.18}$$

where

$$\xi_j(\boldsymbol{r}) := \frac{\frac{1}{N-1} \cdot \delta_j \exp\left(\frac{\Theta_{ll}|\boldsymbol{X}_j^T \boldsymbol{r}|/(N-1)}{\tau}\right)}{\sum_{l=1}^{P} \delta_l \exp\left(\frac{\Theta_{ll}|\boldsymbol{X}_l^T \boldsymbol{r}|/(N-1)}{\tau}\right)}. \tag{S.4.19}$$

#### S.4.3 Derivation of sufficient statistics from association summary statistics data

Without loss of generality, we assume genotype of genetic variant $j$, $\boldsymbol{X}_j$, and phenotype, $\boldsymbol{Y}$, are mean-centered. That is, $\sum_{i=1}^{N} Y_i/N = \sum_{i=1}^{N} X_{ij}/N = 0$. Then, we consider the following simple linear regression between $\boldsymbol{Y}$ and $\boldsymbol{X}_j$ for $j = 1, \cdots, P$. This calculation identifies the reported association quantities and LD information needed by the summary-data implementation. It changes the input representation but not the model fitted by the individual-level implementation.

$$\boldsymbol{Y} = \boldsymbol{X}_j \beta_j + \boldsymbol{\epsilon}_{N\times 1} \text{ with } \boldsymbol{\epsilon} \sim N(\boldsymbol{0}, \sigma^2 \mathbf{I}_N). \tag{S.4.20}$$

To estimate $\hat{\beta}_j$ and $\hat{se}(\hat{\beta}_j)$, the loss function of the simple linear regression is given by $l_j := \sum_{i=1}^{N}(Y_i - X_{ij}\beta_j)^2/N$. We then have

$$\begin{aligned} \frac{\partial l_j}{\partial \beta_j} &= \frac{1}{N}\sum_{i=1}^{N}(Y_i - X_{ij})X_{ij} \\ &= \frac{1}{N}\boldsymbol{X}_j^T \boldsymbol{Y} - \frac{1}{N}\boldsymbol{X}_j^T \boldsymbol{X}_j \beta_j = 0. \end{aligned} \tag{S.4.21}$$

The estimates $\hat{\beta}_j$ and $\hat{se}(\hat{\beta}_j)$ between genetic variant $j$ and phenotype are given by

$$\hat{\beta}_j = \boldsymbol{X}_j^T \boldsymbol{Y} / \boldsymbol{X}_j^T \boldsymbol{X}_j, \tag{S.4.22}$$

$$\hat{se}(\hat{\beta}_j) = \hat{s}/\sqrt{\boldsymbol{X}_j^T \boldsymbol{X}_j} \quad \text{with} \quad \hat{s} = \sqrt{||\boldsymbol{Y} - \hat{\boldsymbol{Y}}||_2^2/(N-2)}. \tag{S.4.23}$$

We can estimate the numerator of $\hat{s}^2$ as

$$\begin{aligned} ||\boldsymbol{Y} - \hat{\boldsymbol{Y}}||_2^2 &= (\boldsymbol{Y} - \boldsymbol{X}_j\hat{\beta}_j)^T(\boldsymbol{Y} - \boldsymbol{X}_j\hat{\beta}_j) \\ &= \boldsymbol{Y}^T\boldsymbol{Y} - \hat{\beta}_j^2 \boldsymbol{X}_j^T \boldsymbol{X}_j \\ &= (N-1)\sigma^2 - (N-1)\hat{\beta}_j^2\sigma_j^2, \end{aligned} \tag{S.4.24}$$

where $\sigma_j^2 = \boldsymbol{X}_j^T \boldsymbol{X}_j/(N-1)$ represent the variance of genotype $\boldsymbol{X}_j$. Therefore, z-score for testing the association effect can be written as

$$\begin{aligned} & z_j = \hat{\beta}_j/\hat{se}(\hat{\beta}_j) = \hat{\beta}_j \times \frac{\sigma_j\sqrt{N-2}}{\sqrt{\sigma^2 - \hat{\beta}_j^2\sigma_j^2}} \\ \Leftrightarrow \quad & \hat{\beta}_j = \frac{z_j}{\sqrt{N-2+z_j^2}} \cdot \frac{\sigma}{\sigma_j}. \end{aligned} \tag{S.4.25}$$

Based on Equation (S.4.22), we can calculate $\boldsymbol{X}_j^T\boldsymbol{Y}$ as

$$\begin{aligned}\boldsymbol{X}_j^T\boldsymbol{Y} &= \boldsymbol{X}_j^T\boldsymbol{X}_j\hat{\beta}_j \\ &= (N-1)\sigma_j^2 \times \frac{z_j}{\sqrt{N-2+z_j^2}} \cdot \frac{\sigma}{\sigma_j} \\ &= \frac{(N-1)z_j}{\sqrt{N-2+z_j^2}} \cdot \sigma_j \cdot \sigma. \end{aligned} \tag{S.4.26}$$

Let $\text{MAF}_j$ denote the minor allele frequency of variant $j$, then, we can estimate $\sigma_j^2 \approx 2 \times \text{MAF}_j \times (1 - \text{MAF}_j)$ if $\text{MAF}_j$ provided in the summary data; otherwise, we can assume the summary data is obtained from scaled genotype such that $\sigma_j^2 = 1$.

#### S.4.4 Derivation of relative change of marginal profile log-likelihood

In this subsection, we introduce the derivations of change in marginal profile log-likelihood with respect to phenotype and residual using sufficient statistics.

##### S.4.4.1 Relative change in profile log-likelihood with respect to phenotype for a trait

The relative change of marginal profile log-likelihood for $\boldsymbol{Y}$ at variant $j$ is used to determine the equivalent between two different variants $j \sim j'$. We define $\Delta loglik(\cdot;\boldsymbol{Y})$ is the relative change of marginal profile log-likelihood with respect to phenotype $\boldsymbol{Y}$, which can be written as

$$\Delta loglik(j;\boldsymbol{Y}) = \frac{|loglik(\boldsymbol{Y}) - loglik(\mathbf{X}_j,\boldsymbol{Y})|}{|loglik(\boldsymbol{Y})|}. \tag{S.4.27}$$

It is also easy to calculate using only the sufficient statistics. Given the ordinary least square (OLS) estimator of effect between $\mathbf{X}_j$ and $\boldsymbol{Y}$ as $\hat{\boldsymbol{\beta}}_j = (\mathbf{X}_j^T\mathbf{X}_j)^{-1}\mathbf{X}_j^T\boldsymbol{Y} = \dfrac{\mathbf{X}_j^T\boldsymbol{Y}}{N-1}$ for $j = 1, \cdots, P$ and $\sigma^2 = \dfrac{\boldsymbol{Y}^T\boldsymbol{Y}}{N-1}$, we can have

$$\begin{aligned} loglik(\mathbf{X}_j,\boldsymbol{Y}) &= -\frac{N}{2}\log(2\pi\sigma^2) - \frac{1}{2\sigma^2}(\boldsymbol{Y} - \mathbf{X}_j\hat{\boldsymbol{\beta}}_j)^T(\boldsymbol{Y} - \mathbf{X}_j\hat{\boldsymbol{\beta}}_j) \\ &= -\frac{N}{2}\log\left(\frac{2\pi\boldsymbol{Y}^T\boldsymbol{Y}}{N-1}\right) - \frac{N-1}{2\boldsymbol{Y}^T\boldsymbol{Y}}\left[\boldsymbol{Y}^T\boldsymbol{Y} - \frac{(\mathbf{X}_j^T\boldsymbol{Y})^T(\mathbf{X}_j^T\boldsymbol{Y})}{N-1}\right] \\ &= -\frac{N}{2}\log\left(\frac{2\pi\boldsymbol{Y}^T\boldsymbol{Y}}{N-1}\right) - \frac{N-1}{2} + \frac{(\mathbf{X}_j^T\boldsymbol{Y})^T(\mathbf{X}_j^T\boldsymbol{Y})}{2\boldsymbol{Y}^T\boldsymbol{Y}} \end{aligned} \tag{S.4.28}$$

$$loglik(\boldsymbol{Y}) = -\frac{N}{2}\log(2\pi\sigma^2) - \frac{1}{2\sigma^2}\boldsymbol{Y}^T\boldsymbol{Y} = -\frac{N}{2}\log\left(\frac{2\pi\boldsymbol{Y}^T\boldsymbol{Y}}{N-1}\right) - \frac{N-1}{2} \tag{S.4.29}$$

Therefore, the relative change of marginal profile log-likelihood is given by

$$\begin{aligned}\Delta loglik(j;\boldsymbol{Y}) &= \frac{|loglik(\boldsymbol{Y}) - loglik(\mathbf{X}_j,\boldsymbol{Y})|}{|loglik(\boldsymbol{Y})|}\\ &= \frac{(\mathbf{X}_j^T\boldsymbol{Y})^T(\mathbf{X}_j^T\boldsymbol{Y})}{2\boldsymbol{Y}^T\boldsymbol{Y}} \Bigg/ \left|\frac{N}{2}\log\left(\frac{2\pi\boldsymbol{Y}^T\boldsymbol{Y}}{N-1}\right) + \frac{N-1}{2}\right|\\ &\approx \frac{\left(\mathbf{X}_j^T\boldsymbol{Y}/(N-1)\right)^T\left(\mathbf{X}_j^T\boldsymbol{Y}/(N-1)\right)}{2\boldsymbol{Y}^T\boldsymbol{Y}/(N-1)} \Bigg/ \left|\frac{1}{2}\log\left(\frac{2\pi\boldsymbol{Y}^T\boldsymbol{Y}}{N-1}\right) + \frac{1}{2}\right|. \qquad \text{(S.4.30)}\end{aligned}$$

##### S.4.4.2 Relative change with respect to residual for a trait

In *Logic 3* of the dynamic coupling strategy, to determine which subset of traits need to updated with the highest $\Delta loglik(\cdot;\boldsymbol{r}^k)$ at boosting round $k$, we also need to compute the relative change of margin log-likelihood for $\boldsymbol{r}^k$ at $j$, $\Delta loglik(j;\boldsymbol{r}^k) = |loglik(\boldsymbol{r}^k) - loglik(\mathbf{X}_j,\boldsymbol{r}^k)|/|loglik(\boldsymbol{r}^k)|$.

Similarly, given that $\hat{\boldsymbol{\beta}}_j^k = (\mathbf{X}_j^T\mathbf{X}_j)^{-1}\mathbf{X}_j^T\boldsymbol{r}^k = \dfrac{\mathbf{X}_j^T\boldsymbol{r}^k}{N-1}$ and $\sigma_{res}^2 = \dfrac{(\boldsymbol{r}^k)^T\boldsymbol{r}^k}{N-1}$, we can have

$$\begin{aligned}loglik(\mathbf{X}_j,\boldsymbol{r}^k) &= -\frac{N}{2}\log(2\pi\sigma_{res}^2) - \frac{1}{2\sigma_{res}^2}(\boldsymbol{r}^k - \mathbf{X}_j\hat{\boldsymbol{\beta}}_j^k)^T(\boldsymbol{r}^k - \mathbf{X}_j\hat{\boldsymbol{\beta}}_j^k)\\ &= -\frac{N}{2}\log\left(\frac{2\pi(\boldsymbol{r}^k)^T\boldsymbol{r}^k}{N-1}\right) - \frac{N-1}{2(\boldsymbol{r}^k)^T\boldsymbol{r}^k}\left[(\boldsymbol{r}^k)^T\boldsymbol{r}^k - \frac{(\mathbf{X}_j^T\boldsymbol{r}^k)^T(\mathbf{X}_j^T\boldsymbol{r}^k)}{N-1}\right]\\ &= -\frac{N}{2}\log\left(\frac{2\pi((\boldsymbol{r}^k)^T\boldsymbol{r}^k}{N-1}\right) - \frac{N-1}{2} + \frac{(\mathbf{X}_j^T\boldsymbol{r}^k)^T(\mathbf{X}_j^T\boldsymbol{r}^k)}{2(\boldsymbol{r}^k)^T\boldsymbol{r}^k}, \qquad \text{(S.4.31)}\\ loglik(\boldsymbol{r}^k) &= -\frac{N}{2}\log(2\pi\sigma_{res}^2) - \frac{1}{2\sigma_{res}^2}(\boldsymbol{r}^k)^T\boldsymbol{r}^k = -\frac{N}{2}\log\left(\frac{2\pi(\boldsymbol{r}^k)^T\boldsymbol{r}^k}{N-1}\right) - \frac{N-1}{2}. \qquad \text{(S.4.32)}\end{aligned}$$

Here, we can estimate $(\boldsymbol{r}^k)^T\boldsymbol{r}^k$ using $(\boldsymbol{Y} - \mathbf{X}\hat{\boldsymbol{\beta}}_j^k)^T(\boldsymbol{Y} - \mathbf{X}\hat{\boldsymbol{\beta}}_j^k) = \boldsymbol{Y}^T\boldsymbol{Y} - 2(\hat{\boldsymbol{\beta}}_j^k)^T\mathbf{X}^T\boldsymbol{Y} + (\hat{\boldsymbol{\beta}}_j^k)^T\mathbf{X}^T\mathbf{X}\hat{\boldsymbol{\beta}}_j^k$. Therefore, the relative change of marginal log-likelihood for $\boldsymbol{r}^k$ at $j$ is given by

$$\begin{aligned}\Delta loglik(j;\boldsymbol{r}^k) &= \frac{|loglik(\boldsymbol{r}^k) - loglik(\mathbf{X}_j,\boldsymbol{r}^k)|}{|loglik(\boldsymbol{r}^k)|}\\ &= \frac{(\mathbf{X}_j^T\boldsymbol{r}^k)^T(\mathbf{X}_j^T\boldsymbol{r}^k)}{2(\boldsymbol{r}^k)^T\boldsymbol{r}^k} \Bigg/ \left|\frac{N}{2}\log\left(\frac{2\pi(\boldsymbol{r}^k)^T\boldsymbol{r}^k}{N-1}\right) + \frac{N-1}{2}\right|\\ &\approx \frac{\left(\mathbf{X}_j^T\boldsymbol{r}^k/(N-1)\right)^T\left(\mathbf{X}_j^T\boldsymbol{r}^k/(N-1)\right)}{2\left((\boldsymbol{r}^k)^T\boldsymbol{r}^k/(N-1)\right)} \Bigg/ \left|\frac{1}{2}\log\left(\frac{2\pi(\boldsymbol{r}^k)^T\boldsymbol{r}^k}{N-1}\right) + \frac{1}{2}\right|. \qquad \text{(S.4.33)}\end{aligned}$$

Note that $0 \le (\boldsymbol{r}^k)^T\boldsymbol{r}^k \le \boldsymbol{Y}^T\boldsymbol{Y}$ theoretically. However, if LD information from a suitable reference panel from the same study population is missing for the summary statistics, then the approximation of $\mathbf{X}^T\mathbf{X}$ maybe inaccurate or ill-conditioned. in such cases, it is even possible to observe $(\boldsymbol{r}^k)^T\boldsymbol{r}^k < 0$ or $(\boldsymbol{r}^k)^T\boldsymbol{r}^k > \boldsymbol{Y}^T\boldsymbol{Y}$, which is theoretically invalid and indicative of instability in the estimation process. Therefore, we stop updating the specific trait if this estimation is invalid. Notice that we also leverage this property as a diagnostic tool in the `colocboost` package to detect potential LD panel mismatches and provide a user warning.

##### S.4.4.3 Log-likelihood ratio test statistic of a CoS for a trait

To calculate the normalized probability of a trait according to CoS $s$ (ignoring index of trait in the corresponding trait configuration), we introduce a log-likelihood ratio test statistic, $LRT^s = -2\log(LR^s)$, between $M_0$, the null model where all variants have zero effects $\boldsymbol{\beta} = 0$ and $M_1^s$, an alternative model that variants in $CoS_s$ have non-zero effects, with $\tilde{\boldsymbol{\beta}}(cos) \neq 0$ estimated from *ColocBoost*. Here, $LRT^s = 2 \times [\mathcal{L}(\boldsymbol{\beta} = 0) - \mathcal{L}(\tilde{\boldsymbol{\beta}}(cos)]$ with

$$\begin{aligned}
\mathcal{L}(\tilde{\boldsymbol{\beta}}(cos)) &= -\frac{N}{2}\log(2\pi\sigma^2) - \frac{1}{2\sigma^2}(\boldsymbol{Y} - \mathbf{X}\tilde{\boldsymbol{\beta}}(cos))^T(\boldsymbol{Y} - \mathbf{X}\tilde{\boldsymbol{\beta}}(cos)) \\
&= -\frac{N}{2}\log(2\pi\sigma^2) - \frac{1}{2\sigma^2}\left(\boldsymbol{Y}^T\boldsymbol{Y} - 2\tilde{\boldsymbol{\beta}}(cos)^T\mathbf{X}^T\boldsymbol{Y} + \tilde{\boldsymbol{\beta}}(cos)^T\mathbf{X}^T\mathbf{X}\tilde{\boldsymbol{\beta}}(cos)\right), \\
\mathcal{L}(\boldsymbol{\beta} = 0)) &= -\frac{N}{2}\log(2\pi\sigma^2) - \frac{1}{2\sigma^2}\boldsymbol{Y}^T\boldsymbol{Y}, \qquad \text{(S.4.34)}
\end{aligned}$$

where $\mathcal{L}(\boldsymbol{\beta})$ indicates the loss function introduced in Equation S.1.4 and $\sigma^2 = \dfrac{\boldsymbol{Y}^T\boldsymbol{Y}}{N-1}$.

#### S.4.5 Statistical reliability of the model parameters

Proximity smoothing gradient boosting algorithm in *FineBoost* relies on several important model parameters, including the dynamic learning rate $\eta_k$ and the smoothing parameter $\tau$ that controls uncertainty in variable selection. In this subsection, we discuss the statistical reliability of these parameters.

##### S.4.5.1 Dynamic learning rate $\eta_k$

In *FineBoost*, we employed a dynamic learning rate $\eta_k = \eta_0/(1 + kd)$, where $\eta_0$ (default as 0.5) represents the initial learning rate and $d$ (default as 1) controls the decay rate of $\eta_k$ over iterations. We use the negative log-likelihood as the loss function introduced in Equation S.1.4, $\mathcal{L}(\boldsymbol{\beta}) := \frac{1}{2}||\boldsymbol{Y} - \mathbf{X}\boldsymbol{\beta}||_2^2$. The gradient of $\mathcal{L}(\boldsymbol{\beta})$ is Lipschitz continuous, a well-known property of quadratic loss functions in regression settings, as discussed in ref[20]. The choice of $\eta_k$ significantly impacts convergency and the design of $\eta_k = \eta_0/(1 + kd)$ initially satisfies the Robbins-Monro conditions for convergence[21],

$$\sum_{k=0}^{\infty} \eta_k = \infty \quad \text{and} \quad \sum_{k=0}^{\infty} \eta_k^2 < \infty, \qquad \text{(S.4.35)}$$

where $\sum \eta_k \to \infty$ (harmonic series) and $\sum \eta_k^2 < \infty$ (p-series with $p = 2$) ensuring convergence to a local minimum in convex optimization.

##### S.4.5.2 Temperature parameter $\tau$

To account for LD-induced uncertainty in variable selection, $\tau$ in Equation S.1.12 plays a critical role in controlling the degree of smoothing over the objective function $g$,

$$g_{smooth}(\boldsymbol{r}^k) = \max_{\boldsymbol{\omega}^k} \sum_j \omega_j^k |\boldsymbol{X}_j^T \boldsymbol{r}^k|/(N-1) - \tau \cdot \rho(\boldsymbol{\omega}^k). \qquad \text{(S.1.12)}$$

As $\tau \to 0$, the objective function of *FineBoost*, $g_{smooth}(\boldsymbol{r}^k)$, reduces to the unsmoothed objective of standard gradient boosting, $g(\boldsymbol{r}^k)$. It greedily focuses on the variant with the strongest association, assigning all weight to $j_k^\star$. Conversely, as $\tau \to \infty$, the weights $\boldsymbol{\omega}^k$ converge to the data-driven association simplex $\boldsymbol{\delta}^k$. This results in a highly smoothed solution where the update is distributed across all variants solely according to $\boldsymbol{\delta}^k$, potentially diluting the focus on the strongest signals and risking underfitting by overemphasizing LD proxies. We recommends a default $\tau = 0.01$, a relatively small value that indicates a preference for minimal smoothing. A theoretical trade-off emerges: increasing $\tau$ enhances robustness to LD by distributing weights across correlated variants, reducing redundancy in variable selection, but it may obscure weaker, independent signals, leading to a loss of sensitivity for detecting subtle effects.

### S.5 Details of inferences on colocalization events

#### S.5.1 Modularity-based hierarchical clustering approach used in defining confidence sets

In *FineBoost*, let $\mathcal{W} = \{\xi_j^k\}$ be a collection of weight vectors, where $\xi_j^k$ is the weight of variant $j$ updated at boosting round $k$ for $j = 1, \cdots, P$ and $k = 1, \cdots, K$. Let $\boldsymbol{\Sigma}$ be an $K \times K$ correlation matrix of weights in $\mathcal{W}$. A modularity-based hierarchical clustering approach we previously developed[6] can be applied to assemble these vectors, which involves the following three steps.
**Step 1**. We first divide $K$ iterations into $s = 1, \cdots, K$ clusters using the hierarchical clustering method based on the correlation matrix $\boldsymbol{\Sigma}$. Let $\delta\left(\boldsymbol{\xi}_{k_1}^{(s)}, \boldsymbol{\xi}_{k_2}^{(s)}\right)$ be the Kronecker function with

$$\delta\left(\boldsymbol{\xi}_{k_1}^{(s)}, \boldsymbol{\xi}_{k_2}^{(s)}\right) = \begin{cases} 1, & \text{if } \boldsymbol{\xi}_{k_1} \text{ and } \boldsymbol{\xi}_{k_2} \text{ are in the same cluster} \\ 0, & \text{otherwise} \end{cases}. \tag{S.5.1}$$

**Step 2**. We then calculate the modularity of the network with $s$ clusters. Denote the modularity of the network, $\boldsymbol{\Sigma}$, as

$$Q_s = \frac{1}{2D} \sum_{k_1, k_2 = 1}^{K} \left( \Sigma_{k_1, k_2} - \frac{d_{k_1} d_{k_2}}{2D} \right) \delta\left(\boldsymbol{\xi}_{k_1}^{(s)}, \boldsymbol{\xi}_{k_2}^{(s)}\right), \tag{S.5.2}$$

where $d_{k_1} = \sum_{k_2=1}^{K} \Sigma_{k_1,k_2}$ and $d_{k_2} = \sum_{k_1=1}^{K} \Sigma_{k_1,k_2}$ are the degrees of $\boldsymbol{\xi}_{k_1}$ and $\boldsymbol{\xi}_{k_2}$, respectively. $D = \sum_{k_1=1}^{K} d_{k_1} = \sum_{k_2=1}^{K} d_{k_2}$ is the total degree of the correlation matrix $\boldsymbol{\Sigma}$.
**Step 3**. Note that the modularity is designed to measure the strength of division of a network into different clusters[22]. The larger modularity indicates that the fraction of edges within the cluster is larger than that of edges between the clusters. Therefore, the optimal number of clusters can be obtained by maximizing the modularity, that is,

$$S = \arg\max\{Q_1, Q_2, \cdots, Q_K\}. \tag{S.5.3}$$

In case that $Q_1 = 0$ by the definition of the modularity of the network from Equation (S.5.2), we always obtain the number of clusters $S \geq 2$. However, if there is only one confidence set included, that is, we determine to update the same $j_k^\star$. Obtaining more than one cluster is not accurate. So, we also include a *minimum cluster correlation* metric to ensure that we can obtain only one confidence set if all boosting rounds are highly correlated (default to 0.8).

#### S.5.2 Criteria to remove potentially spurious signals

Weak learners SEL and SEC in *FineBoost* and *ColocBoost* may capture noise as putative signals, potentially introducing false positives to our findings. To identify and filter spurious signals, we discard the $\alpha$-level (colocalized) confidence sets exhibit low marginal contribution to the overall profile log-likelihood of traits conditional on other (colocalized) confidence sets to ensure that the result reflect true underlying patterns and associations, rather than artifacts of random fluctuations or inherent noise in the data. The procedure has been described in Supplementary Note S.1.3 and Supplementary Note S.2.4. Details are given as follows.

Let $\tilde{\boldsymbol{\beta}}_l$ be coefficients for trait $l$ estimated from *ColocBoost*. The change in overall trait-specific profile log-likelihood for trait $l$ is defined by, $\Delta\mathcal{L}_l = \mathcal{L}(\boldsymbol{\beta}_l = 0) - \mathcal{L}(\tilde{\boldsymbol{\beta}}_l)$,

$$\begin{aligned}
\mathcal{L}(\tilde{\boldsymbol{\beta}}_l) &= -\frac{N_l}{2}\log(2\pi\sigma_l^2) - \frac{1}{2\sigma_l^2}(\boldsymbol{Y}_l - \mathbf{X}_l\tilde{\boldsymbol{\beta}}_l)^T(\boldsymbol{Y}_l - \mathbf{X}_l\tilde{\boldsymbol{\beta}}_l) \\
&= -\frac{N_l}{2}\log(2\pi\sigma_l^2) - \frac{1}{2\sigma_l^2}\left(\boldsymbol{Y}_l^T\boldsymbol{Y}_l - 2\tilde{\boldsymbol{\beta}}_l^T\mathbf{X}_l^T\boldsymbol{Y}_l + \tilde{\boldsymbol{\beta}}_l^T\mathbf{X}_l^T\mathbf{X}_l\tilde{\boldsymbol{\beta}}_l\right), \\
\mathcal{L}(\boldsymbol{\beta}_l = 0) &= -\frac{N_l}{2}\log(2\pi\sigma_l^2) - \frac{1}{2\sigma_l^2}\boldsymbol{Y}_l^T\boldsymbol{Y}_l,
\end{aligned} \tag{S.5.4}$$

where $\mathcal{L}(\boldsymbol{\beta}_l)$ indicates the loss function introduced in Equation S.1.4 and $\sigma_l^2 = \dfrac{\boldsymbol{Y}_l^T\boldsymbol{Y}_l}{N_l - 1}$. Similarly, let $\tilde{\boldsymbol{\beta}}_l^{\neg s} := \tilde{\boldsymbol{\beta}}_l(cos_s|cos_{s'}, s' \neq s))$ be coefficients for trait $l$ estimated from *ColocBoost*, where only variants in $j \in CoS_s$ have zero effects for trait $l$ and other variants have $\tilde{\beta}_{jl}$ for $j \notin CoS_s$. The change in CoS-trait-specific log-likelihood is given by $\Delta\mathcal{L}_l^s = \mathcal{L}(\tilde{\boldsymbol{\beta}}_l) - \mathcal{L}(\tilde{\boldsymbol{\beta}}_l^{\neg s})$, where

$$\mathcal{L}(\tilde{\boldsymbol{\beta}}_l^{\neg s}) = -\frac{N_l}{2}\log(2\pi\sigma_l^2) - \frac{1}{2\sigma_l^2}\left(\boldsymbol{Y}_l^T\boldsymbol{Y}_l - 2(\tilde{\boldsymbol{\beta}}_l^{\neg s})^T\mathbf{X}_l^T\boldsymbol{Y}_l + (\tilde{\boldsymbol{\beta}}_l^{\neg s})^T\mathbf{X}_l^T\mathbf{X}_l\tilde{\boldsymbol{\beta}}_l^{\neg s}\right). \tag{S.5.5}$$

We fine-tuned these thresholds using extensive simulations to balance sensitivity and specificity under various simulation designs. We observed an overall robust performance of *ColocBoost* to these choices with slightly better performance under the default choice, as detailed in Supplementary Note S.6.2. We consider $\Delta\mathcal{L}_l < 0.025$ and $\Delta\mathcal{L}_l^s < 0.1\Delta\mathcal{L}_l$ as default cutoff for *ColocBoost*, and a slightly less conservative yet still well calibrated cutoff $\Delta\mathcal{L}_l < 0.015$ for *FineBoost*.

### S.6 Details on objective functions, tuning parameters, and implementation

In this section, we discuss the design of the objective functions through *FineBoost* with multiple causal effects in different SEL (S.6.1) (*ColocBoost* trivially follow from the same arguments), details in fine-tunning the default model parameters for *ColocBoost* and *FineBoost* (S.6.2), and implementation considerations (S.6.3).

#### S.6.1 Objective function of *FineBoost*

We first discuss the difference of the objective functions between the standard gradient boosting algorithm (Equation (S.1.7)) and the proximity smoothing gradient boosting algorithm (Equation (S.1.14)). Given the details shown in Supplementary Note S.1.2, we recall that the loss function and minimization problem are as follow,

$$\begin{aligned} \mathcal{L}(\boldsymbol{\beta}) &:= \frac{1}{2}||\boldsymbol{Y} - \mathbf{X}\boldsymbol{\beta}||_2^2, \\ \boldsymbol{\beta}^\star &:= \arg\min_{\boldsymbol{\beta}} \left[\max\left\{|\boldsymbol{X}_j^T\boldsymbol{r}|; \forall j = 1, 2, \cdots, P\right\}\right]. \end{aligned} \tag{S.6.1}$$

Therefore, the objective function od the standard gradient boosting algorithm with respect to the residuals $\boldsymbol{r}$ is given in Equation (S.1.7), $g(\boldsymbol{r}) := \max\left\{|\mathbf{X}_j^T\boldsymbol{r}|; \forall j = 1, 2, \cdots, P\right\}$, and $j_k^\star := \arg\max_j |\boldsymbol{X}_j^T\boldsymbol{r}^k|$ at boosting round $k$. Therefore, the objective function given the selected $j_k^\star$ is denoted as

$$g(\boldsymbol{r}^k) = |\mathbf{X}_{j_k^\star}^T\boldsymbol{r}^k|. \tag{S.6.2}$$

Given the positive learning rate $\eta_k > 0$, the gradient descent updates might have the decent objective function, that is, $g(\boldsymbol{r}^{k+1}) < g(\boldsymbol{r}^k)$ for all boosting rounds[2].

In *FineBoost*, we consider a proximity smoothing gradient boosting algorithm, where we bring in a smoothing constraint based on $j_k^\star$ and a data-driven association simplex at boosting round $k$. Therefore, the objective function is introduced in Equation (S.1.14), as follows

$$g_{smooth}^\tau(\boldsymbol{r}^k) = \tau \log\left(\sum_{j=1}^P \delta_j^k \exp\left(\frac{|\boldsymbol{X}_j^T\boldsymbol{r}^k|/(N-1)}{\tau}\right)\right). \tag{S.6.3}$$

Here, $g_{smooth}^\tau(\boldsymbol{r}^k)$ is a function of the optimal weight $\boldsymbol{w}^k := \boldsymbol{w}(j_k^\star)$, which denotes as $g_{smooth}^\tau(\boldsymbol{r}^k) = g_{smooth}^\tau(\boldsymbol{r}^k|\boldsymbol{w}^k)$ (details in Supplementary Note S.4.1). Therefore, across different SEL at different boosting rounds, it is possible that

$$g_{smooth}^\tau(\boldsymbol{r}^{k+1}|\boldsymbol{w}^{k+1}) \geq g_{smooth}^\tau(\boldsymbol{r}^k|\boldsymbol{w}^k) \quad \text{for} \quad j_k^\star \neq j_{k+1}^\star. \tag{S.6.4}$$

This discrepancy may arise because the optimal weights $\boldsymbol{\omega}^k$ and $\boldsymbol{\omega}^{k+1}$ are influenced by the local LD structure and the strength of associations in the region, which can affect the evaluation of the objective function. Notably, if $j_k^\star = j_{k+1}^\star$, the following inequality should hold:

$$g_{smooth}^\tau(\boldsymbol{r}^{k+1}|\boldsymbol{w}^{k+1}) < g_{smooth}^\tau(\boldsymbol{r}^k|\boldsymbol{w}^k). \tag{S.6.5}$$

To illustrate the theoretical behavior described above, we present a simulated example with three true causal variants, demonstrating the progression of the smoothed objective function and the corresponding profile log-likelihood of *FineBoost* across boosting rounds until converged. The left panel shows the trajectory of the objective function values, while the right panel displays the corresponding profile log-likelihood, both indexed by boosting round.

We observe that the profile log-likelihood decreases consistently across iterations, indicating that the residual norm satisfies $\|\boldsymbol{r}^{k+1}\| < \|\boldsymbol{r}^k\|$ as expected. However, the smoothed

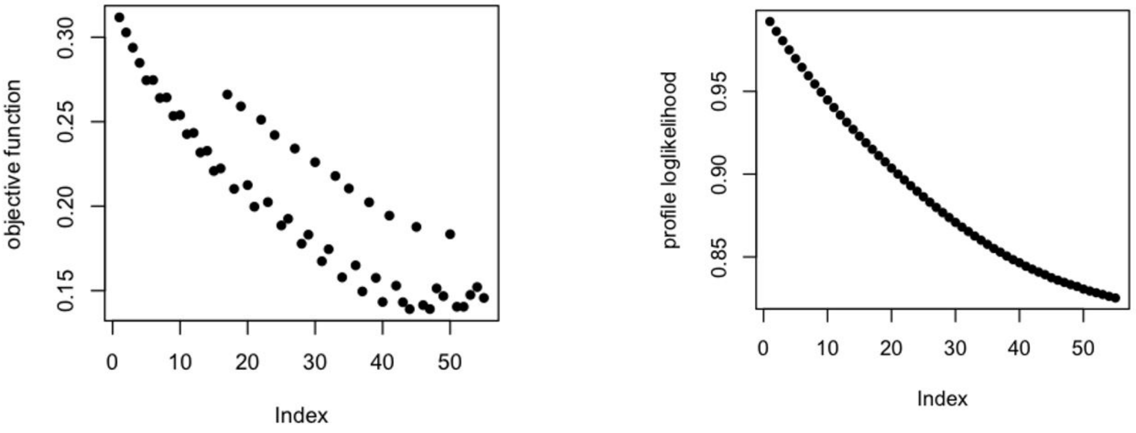

objective function decreases if and only if $j_k^\star = j_{k+1}^\star$. The objective function exhibits a consistent decrease along the boosting round for each of the three causal signals. This behavior reflects alignment between boosting updates and true signals. However, in general, the optimized smoothing weight $\omega^k$ can influence the value of the smoothed objective differently for each selected $j_k^\star$, particularly in regions with complex LD structures.

It is important to highlight a key difference between the standard gradient boosting algorithm and the proximity smoothing gradient boosting algorithm used in *FineBoost*. Specifically, the standard objective function serves as an upper bound on the smoothed objective function across all $j$. That is, $\max_j \left\{\omega_j |\boldsymbol{X}_j^T \boldsymbol{r}|\right\} \leq |\boldsymbol{X}_{j_k^\star}^T \boldsymbol{r}|$, which reflects the additional penalization introduced by proximity-aware smoothing.

#### S.6.2 Choice of model parameter and robustness assessment

Several key parameters influence performance and convergence of the proximity smoothing gradient boosting algorithm used in *FineBoost* and *ColocBoost*, including the learning rate $\eta_k$, smoothing parameter $\tau$, type of data-driven association simplex $\boldsymbol{\delta}_k$, regularizer for simplex $\lambda$, equivalence threshold across best updates, sensitivity of parameters related to D-SEC, weight fudge factor $\nu$, robustness against weak spurious signals, and the sensitivity of correlation between confidence sets. We perform parameter fine-tuning through the extensive simulation studies introduced in Supplementary Note S.7.2.

##### S.6.2.1 Proximity smoothing simplex $\delta^k$

Proximity smoothing in *ColocBoost* uses a local association simplex $\boldsymbol{\delta}^k$ that incorporates both LD and marginal association signals (Equation S.1.13). The default choice implemented in *ColocBoost* (code-named `LD_z2z`) ensures that the update remains local around $j_k^*$ ,

$$\delta_j^k \propto \exp\left(\sqrt{|LD_{j,j_k^\star}|} \times h(Z_{\boldsymbol{X}_j,\boldsymbol{r}^k})\right) \quad \text{where} \quad h(Z_{\boldsymbol{X}_j,\boldsymbol{r}^k}) = (1-\lambda)|Z_{\boldsymbol{X}_j,\boldsymbol{r}^k}| + \frac{\lambda}{2} Z^2_{\boldsymbol{X}_j,\boldsymbol{r}^k}. \quad \text{(S.6.6)}$$

This resemble Bayes factor-based fine-mapping methods like SuSiE[23], which computes posterior inclusion probabilities as $\text{PIP}_j \propto \text{BF}_j = \sqrt{\frac{V+W}{V}} \exp\left(\frac{Z_j^2 W}{2(V+W)}\right)$, where $V$ is variance of estimated effects (1 for z-scores) and $W$ is the prior effect size variance. The $Z^2_{\boldsymbol{X}_j,\boldsymbol{r}^k}$ term mirrors the BF formulation, resulting in normalized weights akin to PIPs. We conducted sensitivity analysis of $\lambda \in [0, 1]$, and observed that increasing $\lambda$ (heavier weights on $Z^2$) enhances power and reduces the size of CoS, but slightly raises FDR (Figure S4b). However, FDR still remains well-controlled across all scenarios. We adopt $\lambda = 0.5$ by default to optimize balance between power and FDR. We observed an overall robust performance of *ColocBoost* to these choices, with slightly better performance under the default choice.

We also considered three alternative choices of $\boldsymbol{\delta}^k$ :

1. Data-driven simplex $\delta_j^k \propto \exp\left(h(Z_{\boldsymbol{X}_j,\boldsymbol{r}^k})\right)$ without using LD (code-named `only_z2z`), designed for cases where LD is unavailable—the *LD-free* mode of *ColocBoost*.

2. Standard simplex $\delta_j^k = 1/P$ (code-named `entropy`), which distributes signal uniformly across all variants for benchmarking purposes.

3. Constrained local association simplex, same as default but bounding $h(Z_{\boldsymbol{X}_j,\boldsymbol{r}^k}) \in [-5, 5]$ (code-named `Range_Z`).

Figure S4a demonstrates that the default (`LD_z2z`) outperforms others, highlighting the advantages of incorporating both association signals and local LD structure for causal variant detection. Nonetheless, the data-driven (`only_z2z`) simplex performed well in single-causal variant scenarios, confirming its utility when LD references are absent.

##### S.6.2.2 Temperature parameter $\tau$

The temperature parameter $\tau$ controls the degree of smoothing over $g$, with smaller values concentrating the update closer around $j_k^\star$ (default $\tau = 0.01$).

$$\xi_j(\boldsymbol{r}^k) := \frac{\dfrac{1}{N-1} \cdot \delta_j^k \exp\left(\dfrac{|\boldsymbol{X}_j^T \boldsymbol{r}^k|/(N-1)}{\tau}\right)}{\displaystyle\sum_{j=1}^{P} \delta_j^k \exp\left(\dfrac{|\boldsymbol{X}_j^T \boldsymbol{r}^k|/(N-1)}{\tau}\right)}. \tag{S.6.7}$$

$$\lim_{\tau \to \infty} \xi_j(\boldsymbol{r}^k) = \delta_j^k/(N-1) \tag{S.6.8}$$

The calibration analyses compare the resulting iteration-level probabilities with simulated causal variants and with SuSiE single-effect probabilities. Separate sensitivity analyses vary the temperature and the association–LD mixing choice so that concentration and reference-measure effects are not conflated.

##### S.6.2.3 Calibration of equivalence across best updates

In *dynamic coupling strategy* of *ColocBoost*, we defined the *equivalence* of $j_k^l \sim j_k^\star$ between the trait-specific best update variant $j_k^l$ and joint best update variant $j_k^\star$,if $j_k^l = j_k^\star$ or in strong LD with each other ($|r| > 0.8$). Similarly, we also establish equivalence between any pair of the trait-specific best update variants $j_k^l$ and $j_k^{l'}$ for $l \neq l'$ using the same criteria of identity or strong LD. The choice of appropriate threshold (code-named `jk_equiv_corr`) of LD for determining this equivalence is crucial, therefore, we established this threshold of LD between best update variants based on the empirical evaluations in our simulation studies (Figure S4c). We established a lower bound of 0.7 for this threshold (LD = $0.7^2 \approx 0.5$) and, also note that with a threshold of 1, each trait will be updated independently without any coupling strategy. As shown in our simulation comparison, a threshold of 0.8 provides an optimal balance of statistical power and false discovery rate control by varing the number of traits. Importantly, all three thresholds maintain FDR well below the conventional 0.05 level (shown by the red dashed line). The stability of performance across different numbers of traits further supports 0.8 as the default threshold for determining best update equivalence in the *dynamic coupling strategy*.

##### S.6.2.4 Sensitivity of parameters related to D-SEC

The *delayed* SEC (D-SEC) was designed to remove or substantially weaken other uncoupled signals by postponing coupled updates at potential colocalized signal to later boosting rounds while prioritizing trait-specific updates in the initial rounds. We recap the previously described the conditions to trigger D-SEC as follows (complete details and notations are given in Supplementary Note S.2.2)

- We identify $\mathcal{R}_k^{\neg\star'}$ ($\mathcal{R}_k^{\neg\star'} \subseteq \mathcal{R}_k^{\neg\star}$) whose $\Delta loglik$ at $j_k^\star$ is substantial (above given threshold), and used a proportional threshold ($p_\Delta$, code-named `coloc_thresh`) in terms of the maximal $\Delta loglik(\cdot; \boldsymbol{r}_l^k)$ for $l = 1, \cdots, L$. Specifically, we considered a threshold of $p_\Delta \times \max_l\{\Delta loglik(\cdot; \boldsymbol{r}_l^k)\}$.
- There are five choices of metrics to compare $\Delta loglik(j_k^\star; \boldsymbol{r}_l^k)$ for $l \in \mathcal{R}^\star$ and $\Delta loglik(j_k^l; \boldsymbol{r}_l^k)$ for $l \in \mathcal{R}^{\neg\star}$ (code-named `func_compare`: *min-max* (default), *median*, *max-min*, *max-max*, *min-min*).

Based on our comprehensive simulation evaluations (Figure S4d-e), we observed remarkable stability in performance across different parameter settings for the D-SEC. We varied $p_\Delta$ from 0.05 to 0.2, maintaining consistent statistical power and well-controlled FDR across different numbers of traits and different causal variant. Similarly, our investigations into alternative comparison metrics demonstrated robust performance across all tested scenarios. The choices of the default setting ($p_\Delta = 0.1$ and *min-max*) align with our design goal to delay the coupled update at $j_k^\star$ to as late as possible in the boosting process, thus ensuring all traits effected by $j_k^\star$ can be coupled for update at later boosting rounds. The stability of performance across different parameter settings further validates the robustness of ColocBoost and its effectiveness in handling complex multi-trait genetic architectures.

##### S.6.2.5 Weight fudge factor $\nu$

Recall that we use the *integrative* weight to define the variant-level probability for variant $j$ supporting $\boldsymbol{T}^s$, $W_j^s := \prod_{l \in \boldsymbol{T}^s} (W_{jl}^s)^{\nu/|\boldsymbol{T}^s = 1|}$, where $\boldsymbol{T}^s$ is a multi-trait configuration for CoS $s$. This integrative weight aggregates all traits with $T_l^s = 1$ into variant-level probability for variant $j$ supporting $\boldsymbol{T}^s$. When $\nu = 1$ this represents a geometric mean and when $\nu = |\boldsymbol{T}^s = 1|$ this is simply the product of all weights assuming complete independence of genetic effects. We set

$\nu = 1.5$ by default to account for partial correlation between genetic traits while ensuring the integrated weight scales consistently regardless of how many traits are colocalizing.

We conducted extensive simulation studies to calibrate the weight fudge factor, comparing performance at values of 1, 1.5, and 2 (Figure S4f). Our empirical results across different values of $\nu$ demonstrated that while there is a modest decrease in statistical power as $\nu$ increases from 1 to 2, this effect is consistent across different numbers of traits. Importantly, $\nu = 1$ maximizes sensitivity by capturing more putative colocalization signals, it potentially increases size of $\alpha$-level CoS with more variants. At $\nu = 2$, we observed increased yet still controlled false discoveries. The default value of $\nu = 1.5$ represents a balance to account for partial correlation between genetic effects across traits while maintaining statistical rigor in identifying true colocalization signals.

##### S.6.2.6 Robustness against weak spurious signals

Weak learners SEL and SEC in *FineBoost* and *ColocBoost* may capture noise as putative signals, potentially introducing false positives to our findings. To identify and filter spurious signals, we discard the $\alpha$-level (colocalized) confidence set $s$ exhibiting low $\Delta\mathcal{L}_l$ and $\Delta\mathcal{L}_l^s$. The procedure has been described in Supplementary Note S.1.3 and Supplementary Note S.2.4, and the detailed calculation of CoS-trait evidence $\Delta\mathcal{L}_l$ and trait-specific evidence $\Delta\mathcal{L}_l^s$ is introduced in Supplementary Note S.5.2.

Based on our simulation studies results (Figure S4g-h), we varied $\Delta\mathcal{L}_l$ from 0.01 to 0.03 (code-named `check_null_max`) and tested different proportional thresholds for $\Delta\mathcal{L}_l^s$ (code-named `check_null`) . Specifically, we considered three proportional values of 0.1, 0.15, and 0.2, that is, $0.1\Delta\mathcal{L}_l$, $0.15\Delta\mathcal{L}_l$, $0.2\Delta\mathcal{L}_l$. The results indicate consistent performance across these parameters, effectively controlling the FDR at the level of 0.05. Notably, the 0.1 threshold achieved the optimal balance between statistical power and FDR control across varying numbers of traits and causal variants. Consequently, we selected 0.1 as the default proportional threshold for the CoS-trait evidence $\Delta\mathcal{L}_l^s$ and 0.025 as the default trait-specific evidence $\Delta\mathcal{L}_l$. We also evaluated the robustness of $\Delta\mathcal{L}_l$ in a single-trait fine-mapping analysis using *FineBoost*, we suggested a slightly less conservative yet still well calibrated cutoff $\Delta\mathcal{L}_l < 0.015$ for *FineBoost*.

##### S.6.2.7 Correlation between colocalization confidence sets (the *between-purity)*

As we discussed in Supplementary Note S.2.4.3, a key challenge arises when different SEC updates produce equivalent causal effects across different subsets of traits—analogous to ensemble methods where individual weak learners capture complementary aspects of the underlying pattern. We introduced *between-purity* as the minimum absolute correlation between variant pairs across two confidence sets, to refine the colocalization confidence sets by additionally considering between-purity in the inference of colocalization events. In our extensive simulation studies, we evaluated different thresholds of between-purity ranging from 0.8 to 0.9, as shown in Figure S4i. Based on this analysis, we selected 0.8 as the default between-purity threshold for refining CoS, as it maintains robust performance across different trait configurations while ensuring high confidence in the detected colocalizations. However, compared to many other tuning parameters in *ColocBoost*, this parameter exhibits greater sensitivity which may change what we define as different colocalization events. We strongly advise users against reducing these thresholds below 0.8 without solid justification, as doing so may force *ColocBoost* to merge multiple potentially separate colocalization events into one larger set and thus undermine the sensitivity to identify separate colocalization loci.

#### S.6.3 Implementation tricks to optimize computational efficiency

Despite its capacity in the scale of genetic variables and traits to integrate, *ColocBoost* is designed for high computational efficiency for large-scale molecular QTL and GWAS applications, enabling rapid inference for datasets with tens to hundreds of traits within minutes (Table 1). This efficiency is not only a result of the model and algorithm by nature, but also achieved through a combination of strategic optimizations tailored to handle both individual-level and summary-level data while minimizing computational overhead. Below, we outline several key strategies employed:

1. To support both individual-level and summary statistics, rather than converting all data to sufficient statistics, *ColocBoost* processes individual-level data directly when available, yielding greater computational efficiency for molecular QTL studies with typically small samples (a few hundred to thousand) compared to GWAS, and reduced memory requirements by eliminating the need for complete LD matrix which can be large for genetic regions with tens of thousands of variants. Our implementation differentially processes data types—applying **Algorithm S1** with SEC for individual-level data and **Algorithm S2** with $\text{SEC}_{\text{ss}}$ for sufficient statistics estimated from summary data, balancing speed and memory usage while maintaining precision in integrative QTL with or without GWAS summary statistics.

2. *ColocBoost* requires only local LD computation for the best update in each boosting round rather than full LD matrices for all traits. A significant bottleneck in large-scale genomic analysis is the multiplication of LD matrix—either computing from individual-level genotype data or loading pre-computed LD reference panels—which can be computationally expensive. *ColocBoost* circumvents this by focusing solely on a local LD vector, limited to LD between the best update variant and all other variants within each SEC. Once the LD vector is obtained for the best update in a given SEC, *ColocBoost* reuses this vector for subsequent updates, avoiding redundant calculations.

3. When analyzing multiple phenotypes with shared genotypes or multiple summary statistics with a common LD matrix, *ColocBoost* implement memory-efficient indexing strategies. Instead of duplicating genotype matrices $\mathbf{X}_l$ to match phenotype dimensions $Y_{l'}$, *ColocBoost* implement a dictionary-based indexing system ($l \to l'$) to map between genotype/LD data and phenotype/summary statistics.

### S.7 Comparison with other competing colocalization methods

Multi-trait colocalization integrating multi-omics data is a challenging problem. Among methods developed for multi-trait colocalization and multi-omics data integration, only a handful are reliable enough to be used in practice. In this section we decribed details of simulation studies comparing *ColocBoost* with these selected methods.

#### S.7.1 Unified variant-level benchmarking for other competing colocalization methods

Because many existing colocalization methods report only a single top-ranked variant, we observed inflated FDR when assessing colocalization events at variant level, particularly under multiple causal variant scenarios, due to failure to account for LD-driven uncertainty in variant selection (Figure S2e). In contrast, our proposed colocalization confidence set (CoS) framework offers a more informative and robust summary (Figure 2c). However, since CoS was initially designed for *ColocBoost* and not naturally available from competing methods, we developed a unified set-level metric by extending each method to construct 95% credible sets analogous to our 95% CoS. These were built using reported variant-level probabilities of each method and filtered with the same LD-based purity threshold (<0.5) to ensure fair and interpretable comparisons, as detailed below.

- **COLOC (V5)**[24,25]: for colocalization configuration where PP.H4 is greater than other PPs (PP.H0, PP.H1, PP.H2, PP.H3), we use `SNP.PP.H4` as variant-level scores to define 95% colocalization credible set similar to how we defined 95% CoS. Unlike many analyses that apply a high PP.H4 cutoff (e.g., 0.7–0.8), we do not impose one here for methods comparison purposes, as the COLOC paper does not rigorously assess or recommend a particular threshold, and our simulations show that omitting it preserves well-controlled FDR while maintaining power comparable to *ColocBoost*. Nonetheless, users requiring more stringent evidence of calling a colocalization event can adopt a higher PP.H4 cutoff in practice.
- **HyPrColoc**[15]: for identified colocalizations with posterior probability higher than uncolocalized configurations, we used `snp.scores` as the variant-level scores to define 95% colocalization credible set. As with COLOC, we omit an explicit configuration probability cutoff here for fair comparison.
- **MOLOC**[26]: for colocalization configuration where PPA.ab is greater than other PPAs (`PPA.a`, `PPA.b`, `PPA.a.b`, and zero), we used `SNP.PP.ab` as the variant-level scores to define 95% colocalization credible set.

#### S.7.2 Detailed simulation settings

We compared *ColocBoost* against three established colocalization methods, COLOC (V5)[24,25], HyPrColoc[15], and MOLOC[26], including realistic simulation analyses, simulation designs adopted in competing methods, and simulation with correlated traits, encompassing multiple scenarios varying in the number of causal variants, variant-trait causal configurations, and effect-size heterogeneity (Figure S2a).

##### S.7.2.1 Simulation settings for multi-trait colocalization methods

We considered genotype data from $N = 1,162$ ROSMAP individuals with available genotypes, focusing on 1,287 unique gene regions ($\pm$1.5Mb from gene TSS) randomly drawn from topologically associating domains (TAD) out of 1,381 human brain TADs genome-wide. Each region contains variants filtered by missing rate less than 0.1 and minor allele frequency (MAF) greater than 0.05, resulting in between 2,069 to 40,000 variants per region (average=9,216). Let $\mathbf{X}$ be the standardized genotype matrix in each region and $\boldsymbol{\beta}_l$ be the normalized effect size for variants on trait $l$, where $P_c$ approximately independent variants were sampled to have causal (non-zero) effects and all the remaining were set to non-causal (zero effect). We simulate the normalized effect sizes such that a fixed percentage of phenotypic variance explained (PVE) by each variant as follows

$$\phi = \frac{\mathbb{V}ar(\boldsymbol{X}_{j_1}\beta_{l,j_1})}{\mathbb{V}ar(\boldsymbol{Y}_l)} \quad \text{for } j_1, \ldots, j_{P_c}.$$

Then, we generated simulated traits based on $Y_l = \mathbf{X}\boldsymbol{\beta}_l + \boldsymbol{\epsilon}$ with $\boldsymbol{\epsilon} \sim \mathcal{N}(0, \sigma_y^2\mathbf{I})$, where $\sigma_y^2$ is chosen such that a modest $\phi = 0.05$ was used for the xQTL-only colocalization setting, and a lower $\phi = 0.02$ for the disease-prioritized setting. While this $\phi$ may appear relatively large for some GWAS traits, real GWAS typically benefit from much larger sample sizes to detect weaker effects with comparable power. Our choice of $\phi$ for simulation purposes is merely a practical trade-off given limited availability of large-scale genotype data.

For simulations under the null, we generated simulated trait based on $\boldsymbol{Y} \sim \mathcal{N}(0, \sigma_y^2)$, where we set $\boldsymbol{\beta}_l = 0$ and varied $\sigma_y^2$ values $\sigma \in \{0.1, 0.5, 1\}$. To simulate dependency between traits due to non-genetic factors such as sample overlap, we model residuals as $\boldsymbol{\epsilon} \sim \mathcal{N}(0, \sigma_y^2\boldsymbol{\Sigma})$, where $\boldsymbol{\Sigma}$ was estimated from the FunGen-xQTL atlas used in our real data analysis (see Supplementary Note S.2.5.4 for details). In our simulation studies, we considered the following simulation designs:

***(1) Primary numerical study***

Our primary numerical study considered three broad scenarios based on different numbers of traits simulated: (i) For simulations with only two traits, we assumed both traits shared the same $P_c$ causal variants, where $P_c$ varied from 1 to 3. (ii) For simulations with 5, 10, and 20 traits, we employed a more complex simulation design to mimic the number of causal variants and variant-trait configurations in real-world data analyses. Specifically, we varied the number of causal variants (1 to 5), and randomly assigned each variant to a subset of traits to determine the colocalization patterns at each gene locus based on an empirical probability distribution derived from the observed genome-wide overlap of fine-mapping credible sets in the FunGen-xQTL atlas (Table S10). For example, when simulating 3 causal variants per gene across 10 traits, we randomly selected 2-10 colocalized traits for each causal variant with probabilities 38.0%, 28.5%, 16.3%, 7.9%, 5.1%, 2.4%, 1.2%, 0.4%, 0.2%. (iii) For simulations with 50 traits, we also varied the number of causal variants (1 to 5), and randomly selected 10–25 colocalized traits per variant. Notably, as the number of traits increases, colocalization patterns tend to become sparser. In some cases, distinct subsets of traits may share different causal variants (e.g., traits {1,2,3} share one variant, while traits {4,5,6} share another).

In (ii), to realistically model colocalization patterns observed in empirical datasets, we utilized genome-wide fine-mapping results derived from 62 FunGen-xQTL contexts. The number of non-overlapping fine-mapped loci within each genomic region served as a proxy for causal variant count. For loci exhibiting overlap across two or more traits (indicating potential colocalization based on fine-mapping evidence), we quantified the number of colocalized traits. This approach yielded a joint probability distribution of colocalized trait counts, conditioned on the total number of traits and causal variants, without dependence on any specific colocalization methodology. During simulation procedures, for each genomic region and causal variant, we stochastically sampled the number of colocalizing traits from this empirically derived distribution.

***(2) Fully colocalized design***

To compare with HyPrColoc with large number of traits, we also adopted a fully colocalized scenario from the simulation design in the HyPrColoc, where all traits share the same causal variants. In our simulation, we extended this framework to compare *ColocBoost* and HyPrColoc under conditions where multiple causal variants simultaneously influence all traits. Our assessment spanned settings with varying phenotypic complexity (5, 10, and 20 traits), with each configuration incorporating up to five true causal variants. This approach allowed us to evaluate method performance under increasingly complex genetic architectures while maintaining complete pleiotropy across all traits.

***(3) Clustered and randomized colocalization design***

We also evaluated simulation designs inspired by HyPrColoc to benchmark performance under more complex trait-variant relationships. We simulated 10 traits under four distinct configurations: (i) `3+3+2+2`, with traits partitioned into four clusters, each sharing a distinct causal variant; (ii) `5+5`, comprising two clusters of five traits, each cluster sharing one causal variant; (iii) `5+5+1rand`, featuring two main clusters plus one additional random variant colocalizing with a third subset of traits; and (iv) `5+5+1rand+1rand`, incorporating two additional random variants each influencing separate trait subsets. These designs enabled comprehensive assessment under both structured and partially unstructured colocalization patterns.

***(4) Residual-correlation simulations under an AR(1) model***

We repeated the causal configurations from the independent-sample baseline under an AR(1) residual model. Specifically, for traits $i, j \in \{1, \ldots, L\}$ with residuals $\varepsilon_i$ and $\varepsilon_j$, we defined the residual correlation as Cor$(\varepsilon_i, \varepsilon_j) = \rho^{|i-j|}$, where $\rho \in \{0.3, 0.5, 0.7, 0.9\}$. Thus, adjacent traits had residual correlation $\rho$, and correlation decayed geometrically with the distance between trait indices. The analysis covered 2, 5, and 10 traits and the same range of causal-variant settings used in the baseline. The corresponding $\rho = 0$ simulations served as the independent-sample reference. We compared power, FDR, median CoS size, and mean set purity. Power, CoS size, and set purity were largely stable, whereas FDR increased under some configurations at $\rho \geq 0.7$. Estimated residual correlations in the xQTL analyses remained below 0.2.

***(5) Prior-sensitivity analysis for the COLOC comparison***

In COLOC, $p_1$ and $p_2$ denote the prior probabilities that a randomly selected variant is associated with trait 1 or trait 2, respectively. The shared-association prior $p_{12}$ denotes the probability that the variant is associated with both traits. We assessed sensitivity to $p_{12}$ using the same two-trait simulations as the primary benchmark, in which the traits shared one, two, or three causal variants. We fixed $p_1 = p_2 = 10^{-4}$ and reran COLOC with $p_{12} \in \{10^{-4}, 10^{-5}, 10^{-6}, 10^{-7}\}$, together with its default $p_{12} = 5 \times 10^{-6}$. All other COLOC inputs and event-calling rules were held fixed across settings. For each setting, we selected the colocalized configuration when PP.H4 exceeded PP.H0, PP.H1, PP.H2, and PP.H3, without imposing an additional PP.H4 cutoff. We then used `SNP.PP.H4` to construct a 95% credible set and evaluated power and FDR using the same set-level definitions as in the primary benchmark. COLOC power decreased from 0.95 at $p_{12} = 10^{-4}$ to 0.40 at $p_{12} = 10^{-7}$, corresponding to an absolute decrease of 0.55 and a relative decrease of approximately 58%. In contrast, ColocBoost does not require a corresponding user-specified colocalization prior.

##### S.7.2.2 Performance metrics for benchmarking multi-trait colocalization methods

We used three principal metrics to compare *ColocBoost* with other methods:

(i) Statistical power: proportion of correctly identified CoS-trait pairs over the total number of true variant-trait pairs in simulated causal configurations.

(ii) False discovery rate (FDR): proportion of incorrectly detected CoS over all detected CoS.

(iii) Variant-level precision-recall curve (AUPRC): precision-recall curves generated by varying the threshold of variant-level colocalization probability from 0 to 1.

Statistical power and FDR are evaluated at the level of a colocalization configuration, for both the 95% CoS and the set of traits involved in this configuration. Specifically, let $\mathcal{T}_{j,true}$ be the set of true colocalized traits for the true causal variant $j$ for $j \in \mathcal{M}_{causal}$ where $\mathcal{M}_{causal}$ is the set of true causal variants. Let $CoS_s$ denote the estimated 95% CoS for colocalization event $s$ and $\hat{\mathcal{T}}_s$ be the set of colocalizing traits associated with this event. For each true causal variant $j$, power is calculated as

$$p_j = \begin{cases} \text{max}\{|\mathcal{T}_{j,true} \cap \hat{\mathcal{T}}_s|\}; & \text{if } \exists s \text{ s.t. } j \in CoS_s \text{ and } \hat{\mathcal{T}}_s \subset \mathcal{T}_{j,true}, \\ 0; & \text{otherwise,} \end{cases}$$

$$\text{Power} = \frac{\sum_{j \in \mathcal{M}_{causal}} p_j}{\sum_{j \in \mathcal{M}_{causal}} |\mathcal{T}_{j,true}|},$$

where the notation $|\mathcal{T}|$ denotes the number of elements in the set $\mathcal{T}$. This definition provides a stringent measure of detection accuracy: ensures that the detected colocalized traits must be a subset of the true set for that variant, and among such cases, power is proportional to the number of correctly identified traits.

To compute FDR, for each detected $CoS_s$ and colocalized traits $\hat{\mathcal{T}}_s$, we considered two stringent conditions to classify false discoveries: (i) $CoS_s$ does not include the true causal variant and (ii) $CoS_s$ includes a causal variant $j$, but also includes incorrect traits ($\hat{\mathcal{T}}_s \not\subset \mathcal{T}_{j,true}$). We assigned $F_s = 1$ if either of these two conditions was met and $F_s = 0$ otherwise. The FDR is calculated as

$$FDR = \frac{\sum_{s=1}^{S} F_s}{S}$$

We further evaluate variant-level detection accuracy using precision-recall curves (PRC), a standard practice adopted in a similar context (fine-mapping) when variant-level scores are determined by both signal strength and its LD proximity and is difficult to separate apart these two factors. In brief, we applied a range of thresholds [0,1] to each variant's colocalization score to compute precision and recall rates. For *ColocBoost*, we used VCP as the colocalization score; see Supplementary Note S.7.1 for details of colocalization score of other methods.

##### S.7.2.3 Comparison with OPERA using "target trait" benchmarks

To benchmark *ColocBoost* with the recently proposed multi-omics colocalization method OPERA[27] in simulation studies, given that OPERA focuses on colocalization with a "target trait" (GWAS) and requiring *genome-wide* GWAS summary statistics for estimating its hyperparameters, we computed summary statistics for 500 of the 1,287 independent, non-overlapping gene regions previously described, to serve as 500 replicates for power and FDR calculations. In our simulation studies, we considered the following simulation designs:

***(1) Primary numerical study***

In our primary simulation comparison, we simulated at the region level for 2, 5, and 10 traits. In each locus, the first trait served as the *target* phenotype with per-variant PVE, $\phi$, ranging from 0.02 to 0.05, while the remaining traits each had $\phi = 0.05$. The number of causal variants and colocalizing traits per variant were randomly assigned based on distribution drawn from genome-wide overlaps of fine-mapped credible sets previously described.

***(2) Simulation design with specific trait configurations in OPERA***

In the secondary analysis, we adopted a probability distribution framework from the simulation design used in the OPERA to model realistic colocalization patterns between target trait and others. In this design, we ensured that the target trait always contains exactly one causal variant while implementing a structured probability configuration for other traits. For the 4-trait simulation (3 + 1 target), we assigned higher probability (0.78) to scenarios with no colocalization, with decreasing probabilities for configurations with increasing numbers of colocalized traits: 0.05 for single-trait colocalization, 0.02 for two-trait colocalization, and 0.01 for complete colocalization. We extended this pattern to the 6-trait simulation (5 + 1 target), where non-colocalized configurations received 0.655 probability, followed by systematically decreasing probabilities for scenarios with one (0.02), two (0.01), three (0.01), four (0.008), and five (0.005) colocalized traits. This design enabled fair comparison between methods by replicating the exact probabilistic framework underlying OPERA's original benchmarks while maintaining consistent evaluation criteria.

##### S.7.2.4 Region-level performance metrics for OPERA comparison

Unlike *ColocBoost* and other methods which identifies variant-level (VCP) and locus-level (CoS) colocalizations, OPERA (i) does not perform xQTL-only colocalization because it requires a target trait (e.g. GWAS), and (ii) only provides gene-level test for whether a colocalized signal exists within an entire gene, without pinpointing the specific variant(s). To enable a fair comparison between OPERA and *ColocBoost*, we consider the GWAS-prioritized mode of *ColocBoost*, and defined power and FDR at gene-level evaluated on the target trait.

Specifically, we defined power as the proportion of correctly identified gene regions colocalized with the target trait over the total true colocalized gene regions. FDR is defined as the proportion of incorrectly identified gene regions colocalized with the target trait over all identified regions. In practice, OPERA may report multiple overlapping subsets ambiguous as to which true colocalization they correspond to due to a lack of variant level information. For instance, for cases with two distinct colocalizations {1,2} and {1,2,3} where trait 1 is the target trait, OPERA often reports three subsets {1,2,3}, {1,2} and {1,3}, the last of which is ambiguous as to whether it is a false positive driven by a incorrect causal variant, or merely subset of {1,2,3} with the correct underlying causal variant. We adopted a lenient evaluation strategy for OPERA: for both the ground truth and reported colocalization, we merged any subsets that are identical or strictly nested into their largest compatible set (here, {1,2,3}) to enable a consistent gene-level assessment. To compare on unified gene-level basis, we likewise apply the same merging strategy to *ColocBoost* CoS, despite it often identifies multiple distinct and correct colocalization events per gene without merging. This procedure ensures a fair comparison for OPERA at gene-level despite underestimating the power of *ColocBoost* at event-level and overlooking finer colocalization patterns it can capture.

### S.8 MaxVCP-based variant annotation and heritability analysis

In this study, we highlight a new idea to evaluate whether variants implicated by multi-trait colocalization inform disease risk, creating an annotation for colocalized xQTLs. To do so, we introduce a new variant annotation based on the *maximum variant colocalization probability* (MaxVCP) from *ColocBoost* as an annotation. In xQTL studies when *ColocBoost* is applied separately for each region of interest (i.e., gene), a single variant may receive multiple VCP corresponding to multiple *cis*-genes. We therefore assigns each variant its highest VCP across all genes—analogous to the MaxCPP annotation in fine-mapping[9]. Specifically, for variants in 95% CoS obtained by *ColocBoost*, let $\mathcal{G}_j$ denote the set of genes whose 95% CoS contains variant $j$, we then define

$$MaxVCP_j = \max_{g \in \mathcal{G}_j} \{VCP_j^g\}, \tag{S.8.1}$$

where $VCP_j^g$ is the variant colocalization probability score for variant $j$ and gene $g \in \mathcal{G}_j$. Similar to Finucane et al. 2018[28], we can use the stratified LD score regression (S-LDSC[29]) to estimate per-SNP heritability, enrichment, and standardized effect size of the colocalizing annotation. In S-LDSC analysis, we model the disease trait of interest, $\boldsymbol{Y}_{disease}$, as depending linearly on genotype, $\boldsymbol{Y}_{disease} = \mathbf{X}\boldsymbol{\beta} + \boldsymbol{\epsilon}$. Because each variant is standardized, S-LDSC assumes the variance of effect size, $\mathbb{V}ar(\beta_j)$, is a linear additive contribution to annotations. We also can call $\mathbb{V}ar(\beta_j)$ the per-variant heritability of variant $j$. Denote $h_g^2 = \sum_j \mathbb{V}ar(\beta_j)$ as the estimated variant heritability across all annotations, then $h_g^2/M$ represents the proportion of heritability explained by per-variant within all annotations, where $M$ is the total number of variants .

Given $C$ functional annotations from the baseline-LD model (v2.2), including binary annotation, conserved, and regulatory annotations (e.g., promoter, enhancer, histone marks, TF binding sites) and continuous-valued LD-related annotations. Let $a_{j,MaxVCP}$ and $a_{j,c}$ be the annotation value of variant $j$ based on MaxVCP from *ColocBoost* and functional annotation $c$ from baseline LD model for $c = 1, \cdots, C$, respectively. The per-variant heritability of variant $j$ is given by

$$\mathbb{V}ar(\beta_j) = a_{j,MaxVCP} \cdot \tau_{MaxVCP} + \sum_c a_{j,c} \cdot \tau_c, \tag{S.8.2}$$

where $\tau_{MaxVCP}$ and $\tau_c$ quantify heritability explained by per-variant within colocalized annotation and annotation $c$, respectively. We can estimate $\tau_{MaxVCP}$ under a linear model between association test statistics and LD score as follows,

$$\mathbb{E}\left[\chi_j^2\right] = N\left(\ell(j, MaxVCP) \cdot \tau_{MaxVCP} + \sum_c \ell(j,c)\tau_c\right) + 1, \tag{S.8.3}$$

where $\ell(j,c)$ is LD score of the variant $j$ to annotation $c$, defined as $\ell(j,c) = \sum_k a_{k,c} r_{jk}^2$ with $r_{jk}$ is correlation between variants $j$ and $k$. Similar definition of $\ell(j, MaxVCP) = \sum_k a_{k,MaxVCP} r_{jk}^2$.

Enrichment is defined as the relative contribution of heritability explained by colocalized annotation (MaxVCP),

$$\text{Enrichment} = \frac{\tau_{MaxVCP}}{h_g^2/M}, \tag{S.8.4}$$

where $\tau_{MaxVCP} = h^2_{g,MaxVCP}/\sum_j a_{j,MaxVCP}$ quantifies heritability explained by per-variant within colocalized annotation, correcting for functional annotations in the baseline LD model (v2.2). Enrichment$> 1$ represents MaxVCP-based annotation enriched to disease heritability. Standardized effect size $\tau^{\star}_{MaxVCP}$ is defined as the proportionate change in per-variant heritability associated with a one standard deviation increase in the value of the colocalized annotation, conditional on other annotations included in the model. $\tau^{\star}_{MaxVCP} > 0$ if MaxVCP-based annotation increases per-variant heritability, accounting for all other annotations included[30]. Both enrichment and $\tau^{\star}_{MaxVCP}$ are estimated via a genomic block-jackknife procedure with 200 blocks that also provides standard errors[29].

We considered two versions of $\tau^{\star}_{MaxVCP}$: a marginal version conditional on baseline-LD annotations only, and a joint version where the conditioning set includes other colocalization-derived annotations in addition to baseline-LD. In the joint analysis, we use a metric, joint $\tau^*$, to quantify the conditional informativeness of a heritability model[31], generalizing the joint $\tau^*$ metrix to more than two annotations. In detail, given a joint model defined by $C$ annotations (conditional on a published set of annotations such as the baseline-LD model), we use

$$\text{Joint } \tau^* = \sqrt{\sum_{c=1}^{C} (\tau_c^*)^2 + \sum_{c \neq l} r_{cl} \tau_c^* \tau_l^*}, \tag{S.8.5}$$

where $r_{cl}$ is the pairwise correlation of the annotations $c$ and $l$, and $r_{cl}\tau_c^*\tau_l^*$ is expected to be positive since two positively correlated annotations typically have the same direction of effect[31]. We calculated standard errors for joint $\tau^*$ using a genomic block-jackknife with 200 blocks.

### S.9 Differences between the two Alzheimer's disease (AD) GWAS studies investigated

Although Bellenguez et. al.[32] and Wightman et. al.[33] AD GWAS have partial cohort overlap, each meta-analysis also draws from distinct datasets. Specifically, Wightman includes cohorts such as HUNT and BioVU, whereas Bellenguez incorporates EADB-TOPMed, among others. For UK Biobank data where proxy AD/dementia cases were defined, these two studies also differ from each other. In Wightman et. al. , the phenotype was derived by counting the number of affected parents (ranging from 0–2) and weighting unaffected parents by their age relative to 100. Bellenguez et. al. on the other hand, classified anyone reporting at least one parent or sibling with dementia (at baseline or follow-up) as a proxy case, leading to 46,828 proxy cases in total. These distinctions in cohort composition, proxy definitions, and sample sizes highlight that a replication study in a downsampling-based fashion between the two GWAS is not straightforward, potentially underestimating the true replication rate.

### S.10 Split-sample reproducibility analyses

We evaluated robustness to reduced sample size in two real-data settings without simulating causal configurations. In each setting, we generated one random two-way partition, denoted $d_1$ and $d_2$, and, for the DLPFC analysis, one random three-way partition, denoted $d_1$, $d_2$, and $d_3$. Each individual was assigned to exactly one nonoverlapping subset within a partition. The complete dataset was denoted $d$. Split-specific association statistics were generated separately for each subset, while the phenotype definition, variant-quality-control criteria, covariate specification, genomic regions, and non-sample-specific method settings were retained from the corresponding complete-data analysis. Thus, "nonoverlapping" refers to the individuals on the partitioned side; the paired dataset on the other side was shared across the subset analyses.

The first setting used ROSMAP DLPFC bulk-eQTL data from $n = 784$ donors. The two-way partition contained 392 donors in each subset, and the three-way partition contained one subset of 262 donors and two subsets of 261 donors. We paired the complete DLPFC dataset and every DLPFC subset with the same primary AD GWAS from Bellenguez et al.[32]. The AD GWAS was not partitioned. We applied *ColocBoost* and COLOC to the complete-data pairing and to each subset pairing. We also applied SuSiE to the complete DLPFC eQTL dataset and separately to every DLPFC subset. The 95% credible sets (CS) from complete-data SuSiE fine-mapping provided a fixed single-trait reference. For the two-way partition, we evaluated CS recovery for *ColocBoost*, COLOC, and split-specific SuSiE fine-mapping. For the three-way partition, the CS-recovery comparison included *ColocBoost* and split-specific SuSiE fine-mapping; COLOC was retained in the locus-overlap analysis but was not included in this three-way CS-recovery comparison.

The second setting used directly measured low-density lipoprotein (LDL) cholesterol in $n = 344{,}613$ White British participants from UK Biobank. We randomly divided these participants into two nonoverlapping subsets of $n = 172{,}306$ and $n = 172{,}307$ and generated the GWAS association statistics separately in each subset. The complete and split-specific LDL GWAS inputs were each paired with the same GTEx v8 liver eQTL data from $n = 208$ donors; the GTEx data were not partitioned. We applied *ColocBoost* and COLOC to the complete-data pairing and to both subset pairings, using the same analyzed loci and method settings throughout.

We used four complementary reproducibility summaries. First, full-data locus recovery was the proportion of loci called in $d$ that were also called in at least one subset, with the complete-data call set from the same method as the denominator. Second, for the DLPFC analysis, full-data CS recovery was the proportion of complete-data SuSiE 95% CS represented by an overlapping split-derived colocalization or fine-mapping set in the same gene region. We evaluated this quantity across all complete-data CS and separately across single-CS gene regions, defined as genes for which complete-data SuSiE returned exactly one CS. Third, directional cross-subset concordance was calculated in both directions: the number of loci shared by $d_1$ and $d_2$ was divided in turn by the total number called in each subset. We also retained the absolute number of loci shared by the two subsets because the directional proportions depend on the total number of calls made by each method. Fourth, for each *ColocBoost* CoS, purity was defined as the minimum absolute pairwise LD correlation among variants in the set, using the LD matrix supplied to the corresponding analysis; singleton CoS were assigned purity 1. We summarized the mean and range of this call-level quantity within each DLPFC subset.

These evaluations therefore quantify robustness and reproducibility under sample partitioning rather than formal statistical power or FDR. Because each subset contains fewer individuals than the complete dataset, lower recovery in a subset is expected and is not interpreted as evidence that a complete-data locus is false.
